# Genome-wide association study meta-analysis of suicide attempt identifies twelve genome-wide significant loci and implicates genetic risks for specific health factors

**DOI:** 10.1101/2022.07.03.22277199

**Authors:** Anna R Docherty, Niamh Mullins, Allison E Ashley-Koch, Xuejun Qin, Jonathan R I Coleman, Andrey Shabalin, JooEun Kang, Balasz Murnyak, Frank Wendt, Mark Adams, Adrian I Campos, Emily DiBlasi, Janice M Fullerton, Henry R Kranzler, Amanda Bakian, Eric T Monson, Miguel E Rentería, Consuelo Walss-Bass, Ole A Andreassen, Cynthia M Bulik, Howard J Edenberg, Ronald C Kessler, J John Mann, John I Nurnberger, Giorgio Pistis, Fabian Streit, Robert J Ursano, Renato Polimonti, Michelle Dennis, Melanie Garrett, Lauren Hair, Philip Harvey, Elizabeth R Hauser, Michael A Hauser, Jennifer Huffman, Daniel Jacobson, Jennifer H Lindquist, Ravi Madduri, Benjamin McMahon, David W Oslin, Jodie Trafton, Swapnil Awasthi, Andrew W Bergen, Wade H Berrettini, Martin Bohus, Harry Brandt, Xiao Chang, Hsi-Chung Chen, Wei J Chen, Erik D Christensen, Steven Crawford, Scott Crow, Philibert Duriez, Alexis C Edwards, Fernando Fernández-Aranda, Manfred M Fichter, Hanga Galfalvy, Steven Gallinger, Michael Gandal, Philip Gorwood, Yiran Guo, Jonathan D Hafferty, Hakon Hakonarson, Katherine A Halmi, Akitoyo Hishimoto, Sonia Jain, Stéphane Jamain, Susana Jiménez-Murcia, Craig Johnson, Allan S Kaplan, Walter H Kaye, Pamela K Keel, James L Kennedy, Minsoo Kim, Kelly L Klump, Daniel F Levey, Dong Li, Shih-Cheng Liao, Klaus Lieb, Lisa Lilenfeld, Adriana Lori, Pierre J Magistretti, Christian R Marshall, James E Mitchell, Richard M Myers, Satoshi Okazaki, Ikuo Otsuka, Dalila Pinto, Abigail Powers, Nicolas Ramoz, Stephan Ripke, Stefan Roepke, Vsevolod Rozanov, Stephen W Scherer, Christian Schmahl, Marcus Sokolowski, Anna Starnawska, Michael Strober, Mei-Hsin Su, Laura M Thornton, Janet Treasure, Erin B Ware, Hunna J Watson, Stephanie H Witt, D Blake Woodside, Zeynep Yilmaz, Lea Zillich, Rolf Adolfsson, Ingrid Agartz, Tracy M Air, Martin Alda, Lars Alfredsson, Adebayo Anjorin, Vivek Appadurai, María Soler Artigas, Sandra Van der Auwera, M Helena Azevedo, Nicholas Bass, Claiton HD Bau, Bernhard T Baune, Frank Bellivier, Klaus Berger, Joanna M Biernacka, Tim B Bigdeli, Elisabeth B Binder, Michael Boehnke, Marco P Boks, Rosa Bosch, David L Braff, Richard Bryant, Monika Budde, Enda M Byrne, Wiepke Cahn, Miguel Casas, Enrique Castelao, Jorge A Cervilla, Boris Chaumette, Sven Cichon, Aiden Corvin, Nicholas Craddock, David Craig, Franziska Degenhardt, Srdjan Djurovic, Ayman H Fanous, Jerome C Foo, Andreas J Forstner, Mark Frye, Justine M Gatt, Pablo V Gejman, Ina Giegling, Hans J Grabe, Melissa J Green, Eugenio H Grevet, Maria Grigoroiu-Serbanescu, Blanca Gutierrez, Jose Guzman-Parra, Steven P Hamilton, Marian L Hamshere, Annette M Hartmann, Joanna Hauser, Stefanie Heilmann-Heimbach, Per Hoffmann, Marcus Ising, Ian Jones, Lisa A Jones, Lina Jonsson, René S Kahn, John R Kelsoe, Kenneth S Kendler, Stefan Kloiber, Karestan C Koenen, Manolis Kogevinas, Bettina Konte, Marie-Odile Krebs, Mikael Landén, Jacob Lawrence, Marion Leboyer, Phil H Lee, Douglas F Levinson, Calwing Liao, Jolanta Lissowska, Susanne Lucae, Fermin Mayoral, Susan L McElroy, Patrick McGrath, Peter McGuffin, Andrew McQuillin, Divya Mehta, Ingrid Melle, Yuri Milaneschi, Philip B Mitchell, Esther Molina, Gunnar Morken, Preben Bo Mortensen, Bertram Müller-Myhsok, Caroline Nievergelt, Vishwajit Nimgaonkar, Markus M Nöthen, Michael C O’Donovan, Roel A Ophoff, Michael J Owen, Carlos Pato, Michele T Pato, Brenda WJH Penninx, Jonathan Pimm, James B Potash, Robert A Power, Martin Preisig, Digby Quested, Josep Antoni Ramos-Quiroga, Andreas Reif, Marta Ribasés, Vanesa Richarte, Marcella Rietschel, Margarita Rivera, Andrea Roberts, Gloria Roberts, Guy A Rouleau, Diego L Rovaris, Dan Rujescu, Cristina Sánchez-Mora, Alan R Sanders, Peter R Schofield, Thomas G Schulze, Laura J Scott, Alessandro Serretti, Jianxin Shi, Stanley I Shyn, Lea Sirignano, Pamela Sklar, Olav B Smeland, Jordan W Smoller, Edmund J S Sonuga-Barke, Gianfranco Spalletta, John S Strauss, Beata Świątkowska, Maciej Trzaskowski, Ming T Tsuang, Gustavo Turecki, Laura Vilar-Ribó, John B Vincent, Henry Völzke, James TR Walters, Cynthia Shannon Weickert, Thomas W Weickert, Myrna M Weissman, Leanne M Williams, Naomi R Wray, Clement C Zai, Esben Agerbo, Anders D Børglum, Gerome Breen, Ditte Demontis, Annette Erlangsen, Tõnu Esko, Joel Gelernter, Stephen J Glatt, David M Hougaard, Hai-Gwo Hwu, Po-Hsiu Kuo, Cathryn M Lewis, Qingqin S Li, Chih-Min Liu, Nicholas G Martin, Andrew M McIntosh, Sarah E Medland, Ole Mors, Merete Nordentoft, Catherine M Olsen, David Porteous, Daniel J Smith, Eli A Stahl, Murray B Stein, Danuta Wasserman, Thomas Werge, David C Whiteman, Virginia Willour, the VA Million Veteran Program (MVP), the MVP Suicide Exemplar Workgroup, Suicide Working Group of the Psychiatric Genomics Consortium, Major Depressive Disorder Working Group of the Psychiatric Genomics Consortium, Bipolar Disorder Working Group of the Psychiatric Genomics Consortium, Schizophrenia Working Group of the Psychiatric Genomics Consortium, Eating Disorder Working Group of the Psychiatric Genomics Consortium, German Borderline Genomics Consortium, Hilary Coon, Jean C Beckham, Nathan A Kimbrel, Douglas M Ruderfer

**Author notes:** Corresponding author **Address for reprint:** Corresponding author, Anna R. Docherty, Department of Psychiatry, University of Utah School of Medicine, 501 Chipeta Way, Salt Lake City, Utah 84110, USA. These authors have contributed equally (Anna R Docherty, Niamh Mullins, Allison E Ashley-Koch). Senior authors (Jean C Beckham, Nathan A Kimbrel, Douglas M Ruderfer). **Previous presentation:** World Congress of Psychiatric Genetics, 2022, Florence, Italy. **Disclosures:** Dr. Qingqin Li is employed by Janssen Pharmaceuticals. Dr. Eli Stahl is a full-time employee of the Regeneron Genetics Center, a subsidiary of Regeneron Pharmaceuticals Inc. All other authors have no financial relationships with commercial interests.

## Abstract

**Objective:** Suicidal behavior is heritable and a major cause of death worldwide. Two large-scale genome-wide association studies (GWAS) recently discovered and cross-validated genome-wide significant (GWS) loci for suicide attempt (SA). The current study leveraged the genetic cohorts from both studies to conduct the largest GWAS meta-analysis of SA to date. Multi-ancestry and admixture-specific meta-analyses were conducted within groups of significant African, East Asian, and European ancestry admixtures.

**Methods:** This study was comprised of 22 cohorts, including 43,871 SA cases and 915,025 ancestry-matched controls. Analytical methods across multi-ancestry and individual ancestry admixtures included inverse variance-weighted fixed effects meta-analyses, followed by gene, gene-set, tissue-set, and drug-target enrichment, as well as summary-data-based Mendelian Randomization with brain eQTL data, phenome-wide genetic correlation, and genetic causal proportion analyses.

**Results:** Multi-ancestry and European ancestry admixture GWAS meta-analyses identified 12 risk loci at p<5×10^−8^. These loci were mostly intergenic and implicated *DRD2, SLC6A9, FURIN, NLGN1, SOX5, PDE4B*, and *CACNG2*. The multi-ancestry SNP-based heritability estimate of SA was 5.7% on the liability scale (SE=0.003, p = 5.7×10^−80^). Significant brain tissue gene expression and drug set enrichment was observed. There was shared genetic variation of SA with ADHD, smoking, and risk tolerance after conditioning SA on both major depressive disorder and post-traumatic stress disorder. Genetic causal proportion analyses implicated shared genetic risk for specific health factors.

**Conclusions:** This multi-ancestry analysis of suicide attempt identified several loci contributing to risk, and establishes significant shared genetic covariation with clinical phenotypes. These findings provide insight into genetic factors associated with suicide attempt across major ancestry admixtures, in veteran and civilian populations, and in attempt versus death.

## Introduction

Suicide was the fourth leading cause of death among 15–29-year-olds in 2019, accounting for more than 700,000 deaths worldwide (1). Suicide attempts (SA; defined as self-injurious behaviors with an intent to die) are even more common (2-4). Suicide attempts are strongly associated with psychiatric conditions, poor quality of life, traumatic experiences, and social and economic burden (1), and are the single strongest predictor of future suicide death (5).

Heritability estimates for suicidal thoughts and behaviors from twin and family studies range from 30-55% (6), and recent large-scale genome-wide association studies (GWAS) have yielded promising and replicable results. The International Suicide Genetics Consortium (ISGC; total *N*=549,743; 29,782 cases) identified two loci reaching genome-wide significance for suicide attempt in individuals of primarily European ancestry admixtures, on chromosomes 6 (index SNP rs71557378, *p =* 1.97×10^−8^) and 7 (index SNP rs62474683, *p =* 1.91×10^−10^) (7). The intergenic locus on chromosome 7 remained significant after conditioning on psychiatric disorders, and was independently replicated (*p =* 3.27×10^−3^) (8) within the Million Veteran Program (MVP) cohort (9). The MVP cohort GWAS of SA (total *N*=409,153; 14,089 cases) resulted in two genome-wide significant multi-ancestry loci, on chromosomes 20 (index SNP rs56817213, *p =* 3.64×10^−9^) and 1 (index SNP rs72730526, *p =* 3.69×10^−8^) (8). A top signal identified at the *Dopamine Receptor D2* locus (*p =* 1.77×10^−7^) also showed moderate association in the ISGC GWAS (*p =* 7.97×10^−4^) (7).

These studies established the complexity of the common variant genetic architecture of suicide attempt and demonstrated the critical role of sample size for discovering novel, replicable risk loci for suicide phenotypes through GWAS (10). Together, these GWAS suggested that larger studies will identify additional genomic risk loci and refine genetic risk metrics.

The objective of the current study was to conduct a meta-analysis of the ISGC and MVP studies (total *N*=958,896; 43,871 suicide attempt and suicide death cases). Moreover, there is considerable need to increase the diversity and generalizability of GWAS data (11). Combining all ISGC and MVP cohorts allowed for the largest GWAS meta-analyses of European, African, and East Asian ancestry admixtures to date. We also tested for gene set enrichment and functional follow-up specific to these primary ancestral admixtures.

## Methods GWAS Cohorts and Phenotype Ascertainment

### The International Suicide Genetics Consortium (ISGC) Cohort

The ISGC analyses included 29,782 suicide attempt (SA) and/or suicide death (SD) cases and 519,961 controls from 18 cohorts (15 SA, 2 SD, and 1 both), 12 of which were ascertained clinically for the purpose of studying psychiatric disorders. Details about the specific cohorts have been described previously (7) and cohort references and ascertainment methods are summarized in Supplementary Table S1. Twelve SA cohorts ascertained information on SA via in-person structured psychiatric interviews conducted by trained clinicians/researchers, two SA cohorts used self-report, and two SA cohorts used ICD codes or hospital records. All interviews and self-report items asked explicitly about SA rather than self-harm (which would also include non-suicidal self-injury). ICD codes were coupled with information from emergency room settings, reason for contact information, and attempt methods that were mined from physician notes, in order to maximize evidence that suicidal intent was present. For the cohorts using interviews or self-report to ascertain SA information, the SA was non-fatal. An additional two cohorts explicitly ascertained cases of suicide death (SD). The majority of SD cases were ascertained from the Utah Office of the Medical Examiner (Utah *n* = 4,692). In these cases, suicide cause-of-death determination results from a detailed investigation of the scene of the death and circumstances of death, determination of medical conditions by full autopsy, review of medical and other public records concerning the case, interviews with survivors, and standard toxicology workups (12). Suicide determination is traditionally conservative due to its impact on surviving relatives. In the 746 suicide deaths from Kobe, Japan, autopsies on suicides were performed and cause of death was determined through discussion with the Medical Examiner’s Office and the Division of Legal Medicine in the Kobe University Graduate School of Medicine. The Columbia University cohort of both SA and SD included 317 suicide deaths that were determined by psychological autopsy and the coroner or medical examiner. A psychological autopsy is a method of determining the psychological factors that may have contributed to a death, considering additional information from family members, friends, acquaintances, medical records and other relevant documents to better characterize a death of uncertain cause, including suspected suicides.

### The Million Veteran Program (MVP) Cohort

MVP recruitment and study procedures have been described previously (8) and included veterans providing a blood sample, consenting to genetic analyses and the linking of one’s genetic information to the VA’s electronic health records (EHR), and completing two optional surveys (9, 13). SA was defined as an act of deliberate self-harm with the intent to cause death that occurred at any point over the lifetime. Briefly, cases were defined as veterans with a documented history of SA in the EHR (N=14,089) and controls were defined as veterans with no documented history of suicidal thoughts or behaviors in the EHR (N=395,064). VA EHR sources were utilized to create a SA phenotype using: (a) diagnostic codes for intentional self-harm; (b) suicidal behavior reports from the VA’s Suicide Prevention Applications Network (SPAN) database; and (c) mental health survey responses from the VA’s Mental Health Assistant database indicating a history of attempting suicide. Veterans who had a history of suicidal ideation but no SA were excluded from analysis. For all ISGC and MVP cohorts, it remains undetermined which individuals with SA may have later died by suicide. Details of sample sizes by genetic ancestry admixture for the ISGC and MVP cohorts are presented in Table 1.

**Table 1.** Summary of GWAS cohorts and primary ancestry admixtures

| **Cohort** | **Attempt/Death** | **Ascertainment** | **Cases** | **Controls** |
| --- | --- | --- | --- | --- |
| **EUR** |  |  |  |  |
| Army STARRS | Attempt | Military | 670 | 10,637 |
| Australian Genetics of Depression Study | Attempt | Psychiatric | 2,792 | 20,193 |
| Columbia University | Attempt & Death | Psychiatric | 577 | 1,233 |
| Genetic Investigation of Suicide and SA (GISS) | Attempt | Psychiatric | 660 | 660 |
| German Borderline Genomics Consortium | Attempt | Psychiatric | 481 | 1,653 |
| iPSYCH | Attempt | Population | 7,003 | 52,227 |
| Janssen | Attempt | Psychiatric | 255 | 1,684 |
| Million Veteran Program | Attempt | Military | 9,196 | 287,370 |
| Psychiatric Genomics Consortium BIP | Attempt | Psychiatric | 3,214 | 17,642 |
| Psychiatric Genomics Consortium ED | Attempt | Psychiatric | 170 | 5,070 |
| Psychiatric Genomics Consortium MDD | Attempt | Psychiatric | 1,528 | 16,626 |
| Psychiatric Genomics Consortium SCZ | Attempt | Psychiatric | 1,640 | 7,112 |
| UK Biobank | Attempt | Population | 2,433 | 334,766 |
| University of Utah | Death | Population | 4,692 | 20,702 |
| Yale-Penn | Attempt | Psychiatric | 475 | 1,817 |
| **Total** |  |  | **35,786** | **779,392** |
| **EAS** |  |  |  |  |
| CONVERGE Consortium | Attempt | Psychiatric | 1,148 | 6,515 |
| Kobe University | Death | Population | 746 | 14,049 |
| Million Veteran Program | Attempt | Military | 115 | 4,082 |
| **Total** |  |  | **2,009** | **24,646** |
| **AA** |  |  |  |  |
| Grady Trauma Project | Attempt | General medical | 669 | 4,473 |
| Million Veteran Program | Attempt | Military | 3,507 | 74,306 |
| Yale-Penn | Attempt | Psychiatric | 629 | 2,902 |
| **Total** |  |  | **4,805** | **81,681** |
| **LAT** |  |  |  |  |
| Million Veteran Program | Attempt | Military | 1,271 | 29,306 |
| **Multi-ancestry Total** |  |  | **43,871** | **915,025** |

Note: EUR = European, EAS = East Asian, AFR = African, LAT = Hispanic/Latino, BIP = bipolar disorder, ED = eating disorders, MDD = major depressive disorder, SCZ = schizophrenia

## Genotyping, quality control, and imputation

Details of genotyping, quality control (QC), and imputation for the ISGC and MVP data sets have been described previously (7, 8). In the ISGC analyses, genotyping was performed locally by each of the research teams using comparable procedures^8^ (details per cohort are available in the Table S1). Standard parameters were used to retain individuals and SNPs after quality control for missingness, relatedness, and Hardy-Weinberg equilibrium. Genetic ancestry was defined by the contributing cohorts, and we include all ascertainment, QC, and analysis details of the ISGC and MVP cohorts in Supplemental Table 1. Imputation was performed using the largest available ancestrally matched reference panels, either from 1000 Genomes or the Haplotype Reference Consortium. We confirmed the comparability of imputation across the cohorts by comparing the final set of SNPs in the meta-analysis, including the number of cohorts in which they were present, and the INFO scores across cohorts and within ancestral admixture groups. Sample overlap and/or cryptic relatedness across cohorts was assessed and corrected for using the meta-analytic tools described below. Eight of the cohorts had high control:case ratios (using an arbitrary cut-off of >15:1). In these cases, the LD Score regression (14) (LDSC) attenuation ratio statistics were examined for evidence of population stratification or uncontrolled type 1 error in the cohort. For any evidence of inflation, the intercept was used to adjust the SE of the summary statistics.

## GWAS meta-analysis of suicide attempt

For both the ISGC and MVP cohorts, the initial GWAS analysis was conducted within genetic ancestral admixture groups. For the ISGC meta-analysis, GWAS were conducted within study and genetic ancestry admixture group, covarying for at least 10 principal components of genetic ancestry, genomic relatedness matrices or factors capturing site of recruitment or genotyping batch, as required (7). For the MVP cohort, ancestry was assigned for four mutually exclusive ancestral groups utilizing a previously defined approach harmonizing genetic ancestry admixture and self-identified ancestry grouping (HARE) (15). Subsequent MVP GWAS analyses were performed within ancestral admixture group using PLINK2 (16), covarying for genetic ancestry principal components, age, and sex.

A multi-ancestry meta-analysis of SA GWAS summary statistics was conducted using an inverse variance-weighted fixed effects model (standard error) in METAL (17), assuming shared risk effects across ancestry admixtures. SNPs with a mean weighted minor allele frequency of <1%, mean weighted imputation INFO score <0.6 or SNPs present in <80% of the total effective sample size were removed to ensure adequate statistical power at every variant included.

Ancestry admixture-specific GWAS meta-analyses were conducted with cohorts of significant European (EUR), African (AFR), and East Asian (EAS) ancestry admixtures using the same procedures. Only one primary ancestral admixture, Hispanic/Latino (LAT), was limited to a single cohort and thus could not be meta-analyzed. Inflation of test statistics due to polygenicity or cryptic relatedness were assessed using the LDSC attenuation ratio ((LDSC intercept - 1)/(mean of association chi-square statistics - 1)). Resulting genome-wide significant (GWS) loci were defined as those with *p*<5×10^−8^ with LD *r*^2^>0.1, within a 3,000 kb window, based on the structure of the Haplotype Reference Consortium (HRC) EUR reference panel for the multi-ancestry meta-analysis, or the HRC ancestry-appropriate reference panel otherwise. GWS loci for SA were examined for heterogeneity across cohorts via the *I*^*2*^ inconsistency metric and forest plots.

## Estimation of heritability and genetic association with other disorders

LDSC (14) and cov-LDSC (18) methods were used to estimate the phenotypic variance in SA explained by common SNPs (SNP-based heritability, *h*^2^_SNP_) from the GWAS meta-analysis summary statistics. LD scores from 1000 Genomes (EUR and EAS) were used to derive *h*^2^_SNP_ for the multi-ancestry GWAS meta-analysis and meta-analyses of European and East Asian ancestry admixtures. To obtain acceptable attenuation ratios for Hispanic/Latino and African ancestry admixture *h*^2^_SNP_ estimates, we used covariate-adjusted AMR LD scores from Pan UK BioBank (Pan UKBB, https://pan.ukbb.broadinstitute.org) and AA LD scores from gnomAD v2.1.1 (19). *h*^2^_SNP_ was calculated on the liability scale assuming a lifetime prevalence of SA in the general population of 2% (middle of the range reported worldwide) (20). The default script of LDSC was used to exclude SNPs with MAF<1% and INFO<0.9 and also to restrict variants to the list of approximately 1.2 million HAPMAP SNPs that are typically well-imputed across datasets. *h*^2^ estimates remained stable across >2% and >5% MAF thresholds. The genetic correlation attributable to genome-wide SNPs (*r*_G_) was estimated between the ancestral admixture groups using the Popcorn package (21), and with a range of psychiatric disorders using LDSC and the largest available discovery GWAS meta-analysis summary statistics (22-33). The latter analyses were confined to European ancestry admixture for consistency with the discovery summary data. Tests were Bonferroni-corrected, adjusting for up to 18 phenotypes hypothesized to be associated with SA based on previous epidemiological association and/or previous evidence of genetic association in LD Hub (34). Previous LD Hub analyses in ISGC were pre-categorized manually into risk factor groups relevant to SA (5, 35, 36): autoimmune disease, neurologic disease, heart disease, hypertension, diabetes, kidney disease, cancer, alcohol use, smoking, pain, psychiatric, sleep, life stressors, socioeconomic, and education/cognition. *r*_G_ of SA in ISGC and MVP in this study were calculated using LDSC, and references for the discovery GWAS are listed in Table S2. Differences in *r*_G_ across other phenotypes using EUR GWAS meta-analyses were tested as a deviation from 0, using the block jackknife method implemented in LDSC (37). To examine phenome-wide partial genetic causality, the Complex-Traits Genetics Virtual Lab (CTG-VL) (38) was used to conduct FDR-corrected Genetic Causal Proportion (GCP) analyses on the EUR summary data.

## Conditioning suicide attempt on major depressive disorder and PTSD

The results of the EUR GWAS SA meta-analysis were conditioned on genetic risks for major depressive disorder (MDD) (27) and post-traumatic stress disorder (PTSD) (32) in secondary analyses, to examine genetic associations both shared with and unique to suicide risk. Results were conditioned because MDD and PTSD are both highly co-morbid with SA, and because PTSD is particularly prevalent within military veteran populations (i.e., MVP). Conditioning was conducted using mtCOJO (multi-trait-based COnditional & JOint analysis using GWAS summary data) (39), implemented in GCTA software (40). mtCOJO estimates the effect size of a SNP on an outcome trait (e.g., SA) conditioned on exposure trait(s) (e.g., MDD). GWS SNPs for the exposure are used as instruments to estimate the effect of the exposure on the outcome, and this effect is used to perform genome-wide conditioning, yielding conditioned effect sizes and *p*-values for the outcome trait. The EUR-only SA GWAS summary statistics were used as the outcome trait, because mtCOJO requires GWAS summary statistics for the exposure trait, which were derived from EUR ancestry discovery GWAS. To select independent SNPs as instruments, we selected those more than 1 megabase (Mb) apart or with an LD r^2^<0.05 based on the 1000 Genomes Project Phase 3 EUR reference panel (41). mtCOJO is robust to sample overlap between the GWAS of the exposure and outcome. In this analysis, statistical power to detect genetic associations at individual SNPs was reduced relative to the unconditioned analysis by the additional model parameters, but the genetic correlations using the conditioned summary statistics provide valuable insights into the relevant risk factors for SA over and above those related to MDD and PTSD.

## Gene, gene pathway, and tissue enrichment analyses

Enrichment analyses of the GWAS results were performed to probe genes, biological pathways, and tissues implicated in SA, using the multi-ancestry and ancestry admixture-specific GWAS results. *P*-values quantifying the degree of association of genes and gene sets with SA were calculated using MAGMA v1.08 (42), implemented in FUMA v1.3.7 (43). Input SNPs were mapped to 18,627 protein-coding genes. Genome-wide significance was defined at *p* = 0.05/18,627 = 2.68×10^−6^. Curated gene sets that included at least 10 genes from MSigDB V7.0 were tested for association with SA. Competitive gene-set tests were conducted to correct for gene size, variant density and LD within and between genes. Tissue-set enrichment analyses were also performed using MAGMA implemented in FUMA, to test for enrichment of association signal in genes expressed in 54 tissue types from GTEx V8 (44) (Bonferroni-corrected *p*-value threshold = 9.26×10^−4^).

## Drug target enrichment analyses

Additional gene-set enrichment analyses of both the multi-ancestry and EUR GWAS meta-analysis results were performed, restricted to genes targeted by drugs, in order to investigate putative relationships of suicide attempt with specific drug types. These analyses do not identify causal relationships, but may implicate genes relevant to pharmacotherapy. This approach has been described previously (45). Gene-level and gene-set analyses were performed in MAGMA v1.08. Gene boundaries were defined using build 37 reference data from the NCBI, available on the MAGMA website (https://ctg.cncr.nl/software/magma), extended 35kb upstream and 10kb downstream to increase the likelihood of including regulatory regions outside of the transcribed region. Gene-level association statistics were defined as the aggregate of the mean and the lowest variant-level *p*-value within the gene boundary, converted to a *Z*-value. Gene sets were defined comprising the targets of each drug in the Drug-Gene Interaction database DGIdb v.2 (46) and in the Psychoactive Drug Screening Ki Database(47), both downloaded in June 2016 (45). Analyses were performed using competitive gene-set analyses in MAGMA.

Results from the drug-set analysis were then grouped according to the Anatomical Therapeutic Chemical class of the drug (45). Only drug classes containing at least 10 valid drug gene sets within them were analyzed, and drug-class analysis was performed using enrichment curves. All drug gene sets were ranked by their association in the drug-set analysis, and then for a given drug class, an enrichment curve was drawn scoring a “hit” if the drug gene set was within the class, or a “miss” if it was outside of the class. The area under the curve was calculated, and a *p*-value for this calculated as the Wilcoxon Mann-Whitney test comparing drug gene sets within the class to drug gene sets outside of the class (45). A Bonferroni-corrected significance threshold of *p* < 5.79×10^−5^ and *p* < 4.35 ×10^−4^ were used for the drug-set and the drug-class analysis, respectively, accounting for 863 drug-sets and 115 drug classes.

## Summary data-based Mendelian randomization

Summary data-based Mendelian randomization (SMR) (v1.03) (48, 49) was applied to detect GWAS signals that co-localize with expression quantitative trait loci (eQTLs), in order to investigate putative causal relationships between SNPs and SA via gene expression. SMR was performed using eQTL summary statistics from the MetaBrain consortium (50), a cortex-derived eQTL dataset consisting of 2,970 EUR-cortex samples. The analysis was conducted using the EUR-only GWAS meta-analysis results, for consistency with the eQTL data. Brain eQTL data from comparable sample sizes in other ancestral groups is not currently available. SMR analysis was limited to transcripts with at least one significant *cis*-eQTL (*p* < 5×10^−8^) in the dataset (of 8,753 in MetaBrain). The Bonferroni-corrected significance threshold for the SMR analysis was *p* < 5.71×10^−6^ and the significance threshold for the HEIDI test (HEterogeneity In Dependent Instruments) (51) was *p* ≥ 0.01. A non-significant HEIDI test suggests a direct causal role, rather than a pleiotropic effect, of the SA-associated SNPs on gene expression.

## Polygenic risk scoring

Polygenic risk scores (PRS) for SA were tested for association with SA versus controls in six target cohorts: PGC MDD, BIP and SCZ (all European ancestry admixtures), CONVERGE (East Asian ancestry admixtures), and Yale-Penn and Grady Trauma Project cohorts (both primarily African ancestry admixtures, located in the United States). The SA GWAS meta-analysis was repeated, excluding each cohort in turn, to create independent discovery datasets. PRS were generated using PRS-CS (51), which uses a Bayesian regression framework to place continuous shrinkage priors on the effect sizes of SNPs in the PRS, adaptive to the strength of their association signal in the discovery GWAS and the LD structure from an external reference panel. The 1000 Genomes EUR, EAS or AFR reference panels (41) were used to estimate LD between SNPs, as appropriate for each target cohort. PLINK 1.9 (16) was used to weight SNPs by their effect sizes calculated using PRS-CS and sum all SNPs into PRS for each individual in the target cohorts. PRS were tested for association with case versus control status in the target cohort using a logistic regression model including covariates as per the GWAS. The amount of phenotypic variance explained by the PRS (*R*^2^) was calculated on the liability scale, assuming a lifetime prevalence of SA in the general population of 2% (20). The Bonferroni-corrected significance threshold adjusting for six tests was P<0.008.

## Results

### Significant shared genetic architecture of SA between civilian (ISGC) and military populations (MVP)

The multi-ancestry GWAS included 43,871 cases and 915,025 controls from 22 cohorts (Table 1). Cases were of predominantly European ancestry admixtures (EUR, 81%), with 11% of cases with significant African ancestry admixtures located in the U.S. (AFR), 5% with East Asian ancestry admixtures (EAS), and 3% with Hispanic/Latino ancestry admixtures located in the U.S. (LAT). Case definition was lifetime SA, with ∼13% of all cases having died by suicide. Additional information on study characteristics and ascertainment methods is presented in Supplementary Table S1.

Cohorts across ISGC and MVP differed with respect to ascertainment, with ISGC being largely civilian and MVP being military (Table 1a). However, examination of the genetic correlation of EUR GWAS meta-analyses for ISGC and MVP (*r*_G_ = 0.81, SE = 0.091, *p* = 2.85×10^−19^) indicated consistency of common-variant genetic architecture across these meta-analyses. Results from both fixed and meta-regression models were comparable in the multi-ancestry and EUR GWAS meta-analyses (all GWS effect size correlations >.99) indicating that ancestry and cohort ascertainment were unlikely to confound observed genetic effects (Table 1b).

### GWAS meta-analysis of SA across and within ancestries identified 12 GWS loci

The multi-ancestry GWAS meta-analysis identified eight genome-wide significant (GWS) loci (P<5×10^−8^) (Figure 1). The *h*^2^_SNP_ of SA was significant at 5.7% (SE=0.003, *p* = 5.70×10^−80^) on the liability scale assuming an SA population prevalence of 2%. The cov-LDSC intercept was 1.04 (SE=0.01, *p* = 1.59×10^−5^) and the attenuation ratio was 0.13 (SE=0.03), indicating that the majority of inflation of GWAS test statistics is likely due to polygenicity (Supplementary Figure S1).

**Figure 1:**
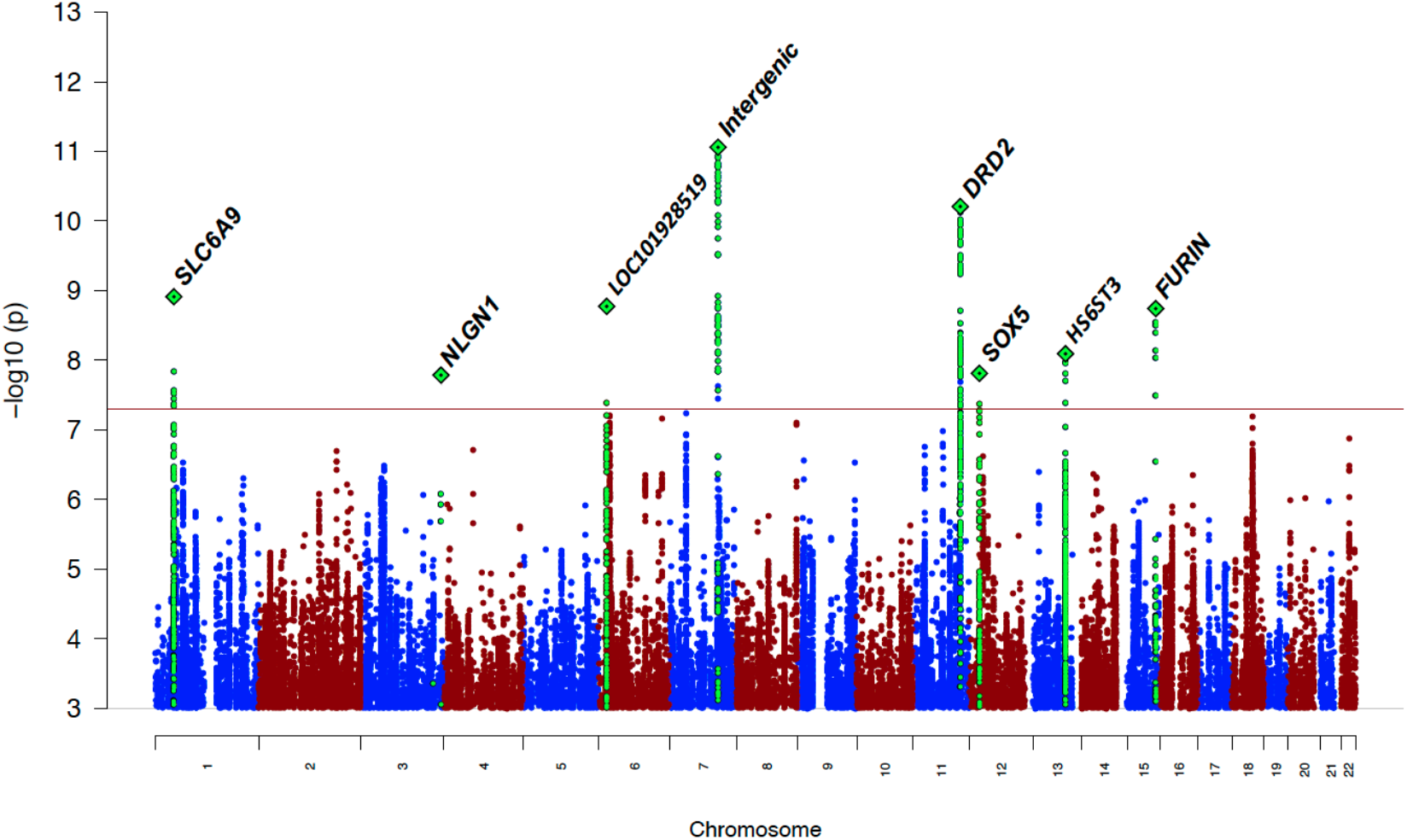
Manhattan plot of multi-ancestry GWAS meta-analysis of suicide attempt. The *x-*axis shows genomic position and the *y-*axis shows statistical significance as –log_10_(P value). The horizontal line shows the genome-wide significance threshold (P<5.0×10^−8^). Labels represent the nearest gene to the index SNP. Regional plots of the eight genome-wide significant loci across ancestry admixtures and the four genome-wide significant loci in EUR are presented in Supplementary Figures S3-S14.

The locus most strongly associated with SA was in an intergenic region on chromosome 7 (index SNP rs62474683, odds ratio (OR) A allele = 1.05 [1.04-1.07], *p* = 8.72×10^−12^, frequency in cases = 0.57, frequency in controls = 0.56, Forest plot Figure S2). At other GWS loci, index SNPs were intronic in the *SLC6A9, DRD2, HS6ST3* and *FURIN* genes (Table 2; additional summary data of all GWS loci are provided in Table S1b). On chromosome 3, a GWS SNP localized to the 5’ untranslated region of the *NLGN1* gene, though the index SNP lacked neighboring SNPs in LD. There was no evidence of heterogeneity of effects across cohorts for any GWS locus according to *I*^2^ heterogeneity indices (Table S1b). Forest plots for GWS loci are included in Figures S2-S9.

**Table 2:**
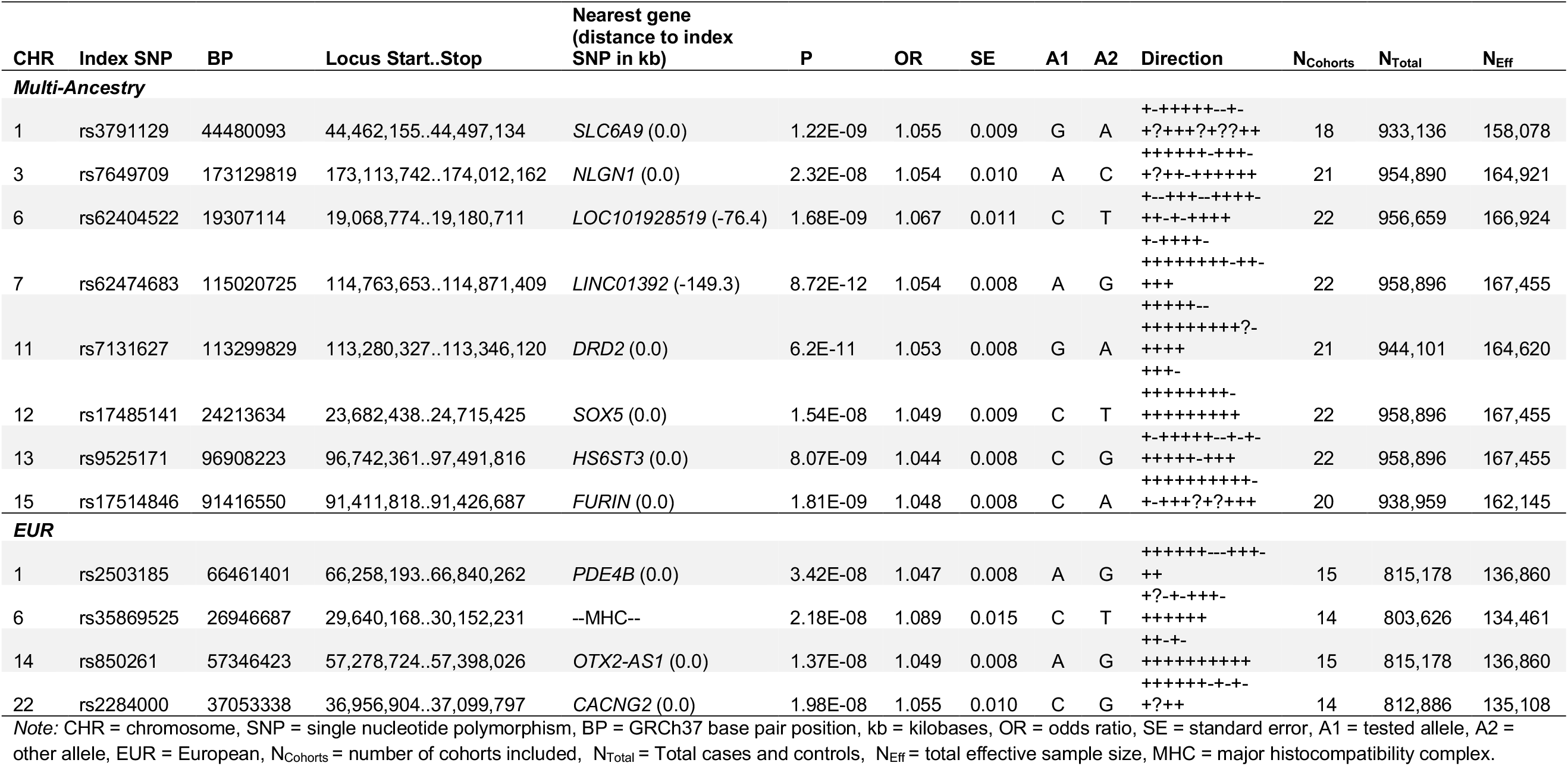
Results from meta-analyses of suicide attempt showing the index SNP from each genome-wide significant locus.

The EUR GWAS meta-analysis *h*^2^_SNP_ was estimated at 7.0% (SE=0.4%) and identified four additional GWS loci (Table 2, Figure S10, Forest plots Figures S11-14), composed of mostly intergenic index SNPs. The nearest genes were *PDE4B, OTX2-AS1, CACNG2*, and one locus was in the major histocompatibility complex (MHC). GWAS meta-analyses in AFR (*h*^2^_SNP_ = 9.8%, SE = 1.8%) and EAS (*h*^2^_SNP_ = 9.8%, SE = 4.5%) produced no GWS loci. The LAT SA *h*^2^_SNP_ (from the MVP GWAS) was estimated at 10.0% (SE = 6.5%). Regional plots of the 12 GWS risk loci across all meta-analyses are presented in Supplementary Figures S15-S26.

Mapped genes from the top loci in multi-ancestry and ancestry admixture-specific meta-analyses are presented in Supplementary Tables S3-S6.

### Genetic correlations of SA across ancestry GWAS

The genetic correlations of SA across each of the ancestral groupings were attenuated, with estimated *r*_G_s between 0.064 (SE = 0.574) (EAS with LAT) and 0.997 (SE = .537) (EUR with LAT) Popcorn *r*_G_ results can be found in Table S7. Individual cohort GWAS were variably powered to estimate genetic correlation estimates with the other cohorts. LDSC estimates across all individual GWAS are presented in Table S8, though cov-LDSC *h*^2^_SNP_ and Popcorn *r*_G_s in Table S7 are the preferred sources for statistics involving ancestry admixtures.

### SA GWS loci are enriched for brain-expressed genes and overlap with previous genetic associations to known risk factors

Significant signal enrichment was observed in genes expressed in pituitary gland and brain tissues, based on the multi-ancestry GWAS (Table S9). Significant gene expression in brain was also observed in the EUR analysis (Table S10). Tissue-set enrichment analyses and corresponding GTEx gene expression heatmaps for all of the multi- and ancestry admixture-specific GWAS are provided in Tables S9-S12 and Figures S31-S34.

Several GWS genes were identified in MAGMA analyses of the multi-ancestry and EUR meta-analyses (Table S13; enrichment of SA signal with genes and gene sets across all meta-analyses are presented in Supplementary Tables S13-S14). MAGMA gene-based tests of the GWAS meta-analyses, with GWS results, are presented in Manhattan plots and QQ-plots in Figures S27-S30. EAS and AA *p*-value thresholds for inclusion of GWAS variants in follow-up analysis were relaxed to *p* < 1×10^−5^ and 1×10^−6^, respectively, in order to explore gene-based tests of top ancestry-specific GWAS variants. Top genes implicated in the EAS analysis included *C11orf87, MYO1C*, and *FAXC*, and top genes implicated in the AFR analysis included *CNTNAP2, IGF2R, MAN1B1*, and *SLC22A1*. Neither set of genes was significantly associated with any pathway or tissue enrichment.

Gene-set analyses from the multi-ancestry and EUR GWAS identified 519 significant gene sets (31 and 488, respectively), spanning multiple domains, including epigenetics, gene regulation and transcription, cellular response to stress, DNA repair, and immunologic signatures (Table S14). The 31 multi-ancestry gene sets included schizophrenia and autism, containing protein-coding genes such as *FURIN, FES*, and *DRD2*, mapped from GWS loci. Most of the 488 EUR gene sets were due to overlap with a small group of 35 histone-coding genes.

Significant proportions of overlapping genes in GWAS Catalog (52) gene sets were observed for both multi-ancestry and EUR meta-analyses (Figures S35-S36). The 12 GWS loci from the multi-ancestry and EUR GWAS meta-analyses were tagged in several GWAS including cognition, smoking, insomnia, and risky behavior. Six of the 12 risk loci had *p*-values < 0.005 for the “*Suicide or Other Intentional Self-Harm*” analysis in FinnGen. A comprehensive list of results of SNP associations from the GWAS Catalog is presented in Table S15. Examination of the pheWAS results (*p* < 0.005) across UK Biobank, FinnGen and the GWAS Catalog resulted in the identification of several psychiatric, weight/BMI- and immune-related traits (Table S16).

Two loci implicated specific genes, *FES* and *TIAF1*, that were significantly associated with SA in SMR analyses and passed the HEIDI test. SMR results suggested that SA risk may be mediated by an increased expression of *FES* (previously implicated in cross-ancestry schizophrenia(53)) and decreased expression of *TIAF1* in cortex (Table S17).

### Significant overlap of SA GWS loci and targets of antipsychotics and antidepressants

Drug target enrichment results suggested that SA risk is most associated with the targets of antipsychotic and antidepressant drug classes. In the multi-ancestry gene-set analysis of the targets of drug classes defined by their Anatomical Therapeutic Chemical (ATC) classes (45), there was significant enrichment in the targets of four drug classes: *Antipsychotics, Psychoanaleptics*, which includes individually significant *Antidepressants* and its subclass *Other Antidepressants* (Table S18). The class of *Other Antidepressants* includes those not classified as selective serotonin reuptake inhibitors, monoamine oxidase inhibitors or monoamine reuptake inhibitors.

In the EUR ancestry admixture GWAS analysis, there was significant enrichment in the targets of just three drug classes, including *Antipsychotics*, the broad class of *Psycholeptics* (drugs with a calming effect on behavior), and the class *Cytotoxic Antibiotics and Related Substances* (Table S19). Only one drug, the insecticide cyfluthrin, was significantly enriched when grouping genes targeted by individual drugs (from the Drug-Gene Interaction Database DGIdb v.2 and the Psychoactive Drug Screening Ki Database) and this was only observed in the EUR GWAS results (see Tables S20 and S21 for multi-ancestry and EUR results).

### Significant genetic correlation of SA with known non-psychiatric risk factors minimally affected after conditioning on MDD and PTSD

The out-of-sample polygenic risk analyses based on the new ISGC+MVP discovery GWAS meta-analysis statistics resulted in higher *R*^2^ estimates than were observed in ISGC1, particularly for AA (maximum variance explained *R*^2^ = 0.66%, *p* = 0.01, and a maximum increase of 146%) and EAS (*R*^2^ = 0.34%, *p* = 8.1×10^−6^, a 36% increase. EUR maximum variance explained = 1.11%, *p* = 6.2×10^−22^, a 24% increase from ISGC1 (Table S22). Figure 2 presents a forest plot of the genetic correlations of the EUR GWAS meta-analyses of suicide attempt with several physical and mental health phenotypes, as well as one control phenotype (body mass index, BMI). Significant shared genetic covariation of EUR SA with smoking (*r*_G_ = 0.46, SE = 0.03, *p =* 8.06×10^−63^), ADHD (*r*_*G*_ *= 0*.55, SE = 0.04, *p* = 2.98×10^−41^), risk tolerance (*r*_*G*_ *= 0*.32, SE = 0.02, *p* = 1.34×10^−59^), and chronic pain (*r*_*G*_ *=* .45, SE = 0.03, *p* = 9.50×10^−50^) were observed both before and after conditioning on MDD and PTSD. Significant positive genetic correlations of neuroticism, schizophrenia, bipolar disorder, and self-harm ideation with SA (*r*_*G*_ *= 0*.45 SE = 0.03, *p* = 1.0×10^−52^ ; *r*_*G*_ *= 0*.43, SE = 0.03, *p* = 1.32×10^−55^; *r*_*G*_ *=* 0.48, SE = 0.04, *p* = 1.81×10^−37^; *r*_*G*_ *=* 0.83, SE = 0.06, *p* = 1.94×10^−51^) did not remain significant after conditioning on both MDD & PTSD.

**Figure 2:**
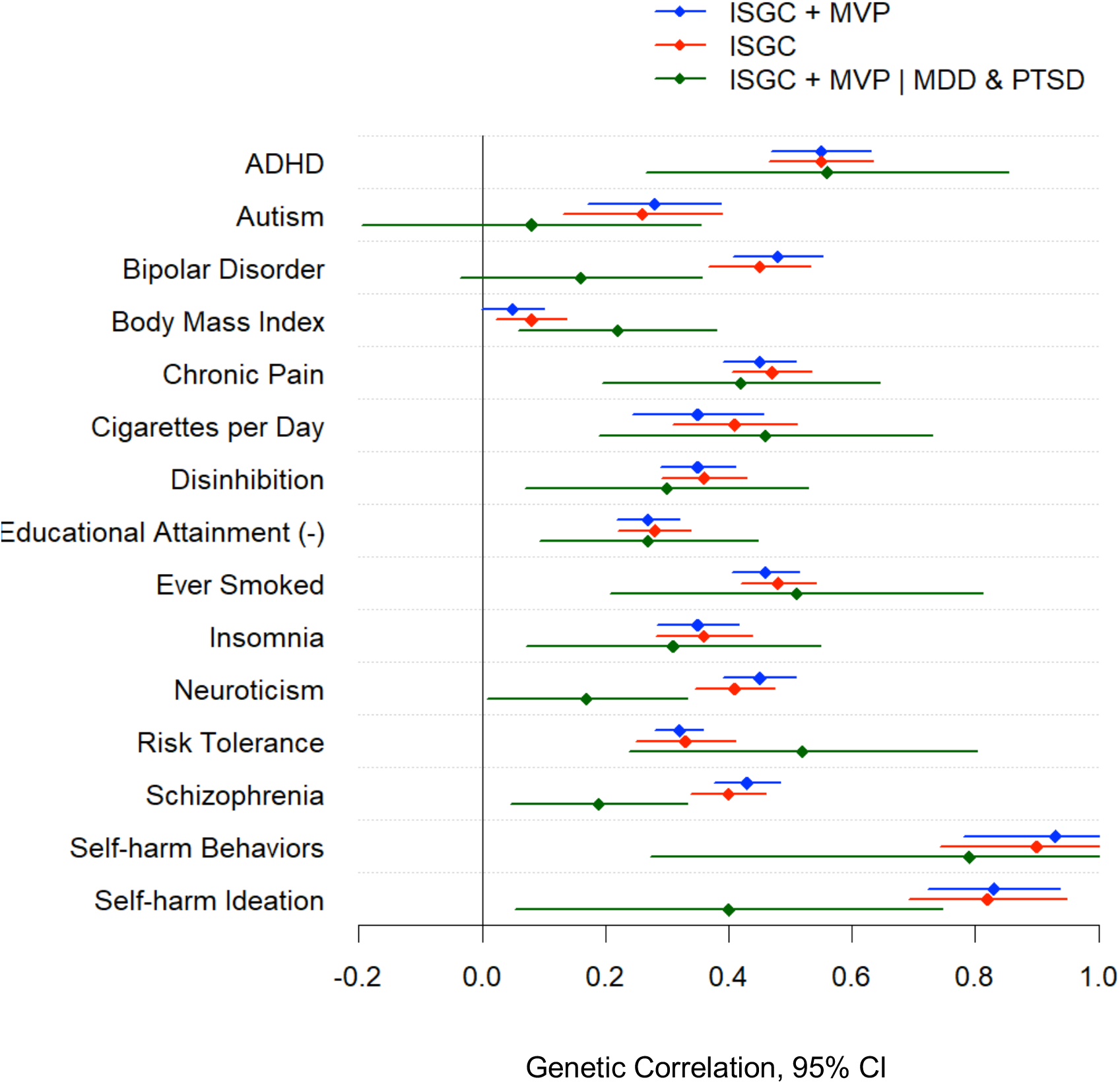
Forest plot of genetic correlations of the multi-ancestry GWAS meta-analyses of suicide attempt with physical and mental health phenotypes. The *x-*axis presents genetic correlation values with 95% confidence intervals (CI), and the *y-*axis presents the discovery GWAS for multiple phenotypes. ISGC = International Suicide Genetics Consortium meta-analysis; ISGC + MVP = the primary meta-analysis including GWAS from both ISGC and Million Veterans Program sets of cohorts; ISGC + MVP | MDD & PTSD = the combined GWAS meta-analysis of both cohorts conditioning on major depressive disorder and post-traumatic stress disorder.

For completeness of comparison across cohorts and phenotypic subgroups (SA versus SD), genetic correlation estimates for phenotypes are presented in Table S23 using the European ancestry admixture GWAS summary statistics from 1) ISGC + MVP, 2) ISGC only, 3) MVP only, 4) ISGC without suicide death, 5) ISGC suicide death only (the Utah Suicide Study, current N = 4,692 EUR suicide deaths and 20,702 controls), and 6) conditioning on MDD and PTSD for MVP, ISGC, and MVP + ISGC. LDSC jackknife tests of differences between these genetic correlation estimates are presented in Table S24, and more exhaustive comparison of phenome-wide *r*_G_ and genetic causal proportion analyses, with the European admixture GWAS meta-analysis, are provided in Table S25. Genetic causal proportion analyses implicated several non-psychiatric genetic risks in EUR SA, including particulate air matter pollution exposure (pm 2.5), smoking exposures, and pulmonary health factors. Risk factors with significant partial genetic causality estimates are presented in Table S25.

## Discussion

This study presents the largest GWAS meta-analysis of SA to date, incorporating multiple ancestries and expanding the set of GWS loci from four to 12. Discovery of three of the novel GWS loci, and improved out-of-sample PRS prediction across ancestry, was only possible with the aggregation of all ancestral cohorts. For the first time, we show that implicated genes are highly expressed in brain tissue, enriched in pathways related to gene regulation and transcription, cellular response to stress, DNA repair, and immunologic signatures, and are shared with epidemiological risk factors. Genetic correlation and causal proportion analyses implicate a number of non-psychiatric genetic risks in SA, including pulmonary health factors. We also provide important evidence that a significant proportion of the common variant genetic architecture of SA is shared across large civilian and veteran populations with disparate demographics.

One advantage of combining the ISGC with MVP was the opportunity to examine genetic effects across heterogeneous cohorts. For example, the sample composition and ascertainment across the ISGC is predominantly civilian and international, with a large proportion of females (7). A number of the ISGC samples from the Psychiatric Genomics Consortium cohorts (Table 1) are collected from individuals with major psychiatric disorders, representing a more clinical population. In contrast, the MVP cohorts are predominantly male (8), and all are military veterans ascertained through the U.S. Department of Veterans Affairs (VA) healthcare system. The consistency of SA common variant genetic architecture across EUR MVP and ISGC cohorts indicates that power may be further enhanced by combining future cohorts with differing ascertainments.

As expected, the increase in sample size, and resulting increase in power, led to the identification of several new GWS loci and improved out-of-sample PRS prediction, across ancestries, relative to the previous ISGC-only analyses. The loci identified in this study implicate genes expressed in brain. Genes associated with SA in this study are highly enriched among psychiatric phenotypes and overall health and wellness risk factors for SA. Brain is the predominant tissue enriched for associated genes, and there is also significant enrichment in pituitary gland, consistent with previous association of SA with hypothalamic-pituitary-adrenal system dysregulation (54). In addition, the enrichment of pathways related to epigenetics and gene regulation and transcription suggest that epigenetic modifications, such as DNA methylation, may play a role in modulating the effect of SA-associated genetic variants. However, epigenetic pathways were only enriched in GWAS of European ancestry admixture, pointing to the potential importance and varied impact of epigenetic mechanisms in diverse biological systems that may contribute to SA risk. Pathways enriched in the multi-ancestry GWAS were absent of histone-coding genes, and contained protein-coding genes mapped from GWS loci such as *FURIN, FES*, and *DRD2*. These multi-ancestry pathway results, while harder to interpret, may be more generalizable to the global population.

Drug target enrichment results suggest that SA risk is associated with the targets of antipsychotic and antidepressant drug classes. One explanation may be that psychiatric symptoms associated with SA risk are also associated with these drug targets, though the direction of any association of drugs with risk cannot be assumed and is not directly tested here. The SMR analysis of EUR results implicated *FES* and *TIAF1* in SA. *FES* has been previously implicated in cross-ancestry schizophrenia (53).

Genetic correlations of SA with ADHD, smoking, pain, and risk tolerance remained significant after conditioning SA on both MDD and PTSD, while schizophrenia, bipolar disorder, and neuroticism did not. This suggests a potential role for health factors in SA risk that are both shared with and distinct from psychiatric disorders, as proposed in Mann & Rizk’s stress diathesis model (55) of suicidal behavior based on clinical and biological studies. The suicide diathesis includes altered decision-making that may be more pronounced in the context of ADHD and smoking, and may be aggravated by sleep problems. Pain is associated with the stress domain of suicidal behavior, and is also associated with increased access to prescription opioids. Overall, this study leverages genetic data to examine important risk phenotypes that may or may not be present in medical records.

Some limitations of this study should be considered. First, a meta-analysis of such a large number of diverse cohorts, with different assessments of SA, could reduce statistical power by increasing heterogeneity. Our analyses remain still more conservative with the inclusion of age and sex covariates in three of the ISGC cohorts and MVP. However, GWAS of the primary datasets typically produced significant—and high—genetic correlation estimates. GWS loci produced similar effect sizes across cohorts and across fixed and meta-regression models (correlations of EUR and multi-ancestry GWS effect sizes across models exceeded 0.99). Indeed, the apparent consistency of genetic architecture across EUR ISGC and MVP cohorts is important given marked demographic and ascertainment differences.

This study also provides GWAS meta-analyses specific to African and East Asian ancestry admixtures. The lack of GWS loci specific to these SA meta-analyses underscores a strong need for greater ancestral diversity and representation in suicide genetics research. With high variability of sample sizes of individual ISGC and MVP ancestral cohorts (case *n*s ranging from 115 to 9,196) some GWAS yielded *h*^2^ and *r*_G_ estimates, while others did not. Variability in *r*_G_ indicates that increasing the examination of non-European ancestries in the future will significantly increase the generalizability of the genetic risk signals identified from studies of suicide phenotypes and the portability of polygenic scores. Importantly, broader ancestral representation, particularly from population-dense areas such as India, Western Asia, and the Global South, will be critical for improving the rigor and generalizability of GWAS results in future research.

Implicated genes and established genetic relationships with ADHD, smoking, and risk tolerance help to inform our understanding of biological contributions to risk of SA. From a clinical standpoint, impulsivity, smoking status, and risk-taking behaviors are intuitive co-morbid indicators of suicide risk. Genetic causal proportion analyses implicate these and other health factors—pulmonary and cardiovascular—in risk for SA. And our preliminary comparison of genetic correlations across SA vs. SD GWAS cohorts appears to implicate risk tolerance in the severity of the suicide phenotype. Further study, comparing SD and SA with subjects with suicidal ideation, will allow for a comparison of those who think about suicide and those who act. Importantly, genetic risk for SA, calculated in new independent cohorts using these GWAS summary data, will contribute to a deeper understanding of the clinical implications of genetic risk for suicide. The future addition of multiple ancestral cohorts is likely to yield continued discovery and increased opportunity for clinical translation.

## Data Availability

All data produced in the present study are available upon reasonable request to the authors.

https://www.ncbi.nlm.nih.gov/pmc/articles/PMC6675659/

## Acknowledgements

This work was supported by the National Institute of Mental Health (R01MH123619, R01MH123489, R01MH099134, K01MH109765, R01MH116269, R01MH121455). This research is also based on data from the U.S. Department of Veterans Affairs (VA) Million Veteran Program (MVP) program and was supported by Award #I01CX001729 from the Clinical Science Research and Development Service of the Veterans Health Administration Office of Research and Development. J.C. Beckham was also supported by a Senior Research Career Scientist Award (#lK6BX003777) from CSR&D. Research reported in this publication was supported by NIGMS of the National Institutes of Health under award number T32GM007347 (J.K.). This work was also supported by the Brain & Behavior Research Foundation/NARSAD Young Investigator Awards No. 29551 to N. Mullins, No. 28686 to A. Shabalin, and No. 28132 to E. DiBlasi. Work was supported by the Huntsman Mental Health Institute, the American Foundation for Suicide Prevention (A. Docherty, A. Shabalin, E. DiBlasi, A. Bakian, H. Coon), and the Clark Tanner Research Foundation. This work was also supported by research funding from Janssen Research & Development, LLC to University of Utah. Data from the Utah cohort is available through the Utah Population Database, which is partially supported by the Huntsman Cancer Institute and award number P30CA42014 from the National Cancer Institute. Several statistical analyses were carried out on the Mount Sinai high performance computing cluster (http://hpc.mssm.edu), which is supported by the Office of Research Infrastructure of the National Institutes of Health (Grant Nos. S10OD018522 and S10OD026880). This work was also conducted in part using the resources of the Advanced Computing Center for Research and Education at Vanderbilt University, Nashville, TN and the Huntsman Mental Health Institute at the University of Utah School of Medicine, Salt Lake City, UT. This publication does not represent the views of the Department of Veteran Affairs or the United States Government. We also thank and acknowledge MVP, the MVP Suicide Exemplar Workgroup, and the ISGC for their contributions to this manuscript. A complete listing of contributors from the MVP, MVP Suicide Exemplar Workgroup, and the ISGC is provided in the Supplementary Materials.

## References

1. Organization TWH: Suicide.

2. Organization TWH: Preventing suicide: A global imperative.

3. (SAMHSA) SAaMHSA.

4. Control CfD.

5. Franklin JC, Ribeiro JD, Fox KR, Bentley KH, Kleiman EM, Huang X, Musacchio KM, Jaroszewski AC, Chang BP, Nock MK. Risk factors for suicidal thoughts and behaviors: A meta-analysis of 50 years of research. Psychol Bull. 2017;143:187–232.

6. Voracek M, Loibl LM. Genetics of suicide: a systematic review of twin studies. Wien Klin Wochenschr. 2007;119:463–475.

7. Mullins N, Kang J, Campos AI, Coleman JRI, Edwards AC, Galfalvy H, Levey DF, Lori A, Shabalin A, Starnawska A, Su MH, Watson HJ, Adams M, Awasthi S, Gandal M, Hafferty JD, Hishimoto A, Kim M, Okazaki S, Otsuka I, Ripke S, Ware EB, Bergen AW, Berrettini WH, Bohus M, Brandt H, Chang X, Chen WJ, Chen HC, Crawford S, Crow S, DiBlasi E, Duriez P, Fernandez-Aranda F, Fichter MM, Gallinger S, Glatt SJ, Gorwood P, Guo Y, Hakonarson H, Halmi KA, Hwu HG, Jain S, Jamain S, Jimenez-Murcia S, Johnson C, Kaplan AS, Kaye WH, Keel PK, Kennedy JL, Klump KL, Li D, Liao SC, Lieb K, Lilenfeld L, Liu CM, Magistretti PJ, Marshall CR, Mitchell JE, Monson ET, Myers RM, Pinto D, Powers A, Ramoz N, Roepke S, Rozanov V, Scherer SW, Schmahl C, Sokolowski M, Strober M, Thornton LM, Treasure J, Tsuang MT, Witt SH, Woodside DB, Yilmaz Z, Zillich L, Adolfsson R, Agartz I, Air TM, Alda M, Alfredsson L, Andreassen OA, Anjorin A, Appadurai V, Soler Artigas M, Van der Auwera S, Azevedo MH, Bass N, Bau CHD, Baune BT, Bellivier F, Berger K, Biernacka JM, Bigdeli TB, Binder EB, Boehnke M, Boks MP, Bosch R, Braff DL, Bryant R, Budde M, Byrne EM, Cahn W, Casas M, Castelao E, Cervilla JA, Chaumette B, Cichon S, Corvin A, Craddock N, Craig D, Degenhardt F, Djurovic S, Edenberg HJ, Fanous AH, Foo JC, Forstner AJ, Frye M, Fullerton JM, Gatt JM, Gejman PV, Giegling I, Grabe HJ, Green MJ, Grevet EH, Grigoroiu-Serbanescu M, Gutierrez B, Guzman-Parra J, Hamilton SP, Hamshere ML, Hartmann A, Hauser J, Heilmann-Heimbach S, Hoffmann P, Ising M, Jones I, Jones LA, Jonsson L, Kahn RS, Kelsoe JR, Kendler KS, Kloiber S, Koenen KC, Kogevinas M, Konte B, Krebs MO, Landen M, Lawrence J, Leboyer M, Lee PH, Levinson DF, Liao C, Lissowska J, Lucae S, Mayoral F, McElroy SL, McGrath P, McGuffin P, McQuillin A, Medland SE, Mehta D, Melle I, Milaneschi Y, Mitchell PB, Molina E, Morken G, Mortensen PB, Muller-Myhsok B, Nievergelt C, Nimgaonkar V, Nothen MM, O’Donovan MC, Ophoff RA, Owen MJ, Pato C, Pato MT, Penninx B, Pimm J, Pistis G, Potash JB, Power RA, Preisig M, Quested D, Ramos-Quiroga JA, Reif A, Ribases M, Richarte V, Rietschel M, Rivera M, Roberts A, Roberts G, Rouleau GA, Rovaris DL, Rujescu D, Sanchez-Mora C, Sanders AR, Schofield PR, Schulze TG, Scott LJ, Serretti A, Shi J, Shyn SI, Sirignano L, Sklar P, Smeland OB, Smoller JW, Sonuga-Barke EJS, Spalletta G, Strauss JS, Swiatkowska B, Trzaskowski M, Turecki G, Vilar-Ribo L, Vincent JB, Volzke H, Walters JTR, Shannon Weickert C, Weickert TW, Weissman MM, Williams LM, Wray NR, Zai CC, Ashley-Koch AE, Beckham JC, Hauser ER, Hauser MA, Kimbrel NA, Lindquist JH, McMahon B, Oslin DW, Qin X, Major Depressive Disorder Working Group of the Psychiatric Genomics C, Bipolar Disorder Working Group of the Psychiatric Genomics C, Eating Disorders Working Group of the Psychiatric Genomics C, German Borderline Genomics C, Workgroup MVPSE, Program VAMV, Agerbo E, Borglum AD, Breen G, Erlangsen A, Esko T, Gelernter J, Hougaard DM, Kessler RC, Kranzler HR, Li QS, Martin NG, McIntosh AM, Mors O, Nordentoft M, Olsen CM, Porteous D, Ursano RJ, Wasserman D, Werge T, Whiteman DC, Bulik CM, Coon H, Demontis D, Docherty AR, Kuo PH, Lewis CM, Mann JJ, Renteria ME, Smith DJ, Stahl EA, Stein MB, Streit F, Willour V, Ruderfer DM. Dissecting the Shared Genetic Architecture of Suicide Attempt, Psychiatric Disorders, and Known Risk Factors. Biol Psychiatry. 2022;91:313–327.

8. Kimbrel NA, Ashley-Koch AE, Qin XJ, Lindquist JH, Garrett ME, Dennis MF, Hair LP, Huffman JE, Jacobson DA, Madduri RK, Trafton JA, Coon H, Docherty AR, Kang J, Mullins N, Ruderfer DM, Program VAMV, Workgroup MVPSE, International Suicide Genetics C, Harvey PD, McMahon BH, Oslin DW, Hauser ER, Hauser MA, Beckham JC. A genome-wide association study of suicide attempts in the million veterans program identifies evidence of pan-ancestry and ancestry-specific risk loci. Mol Psychiatry. 2022;27:2264–2272.

9. Gaziano JM, Concato J, Brophy M, Fiore L, Pyarajan S, Breeling J, Whitbourne S, Deen J, Shannon C, Humphries D, Guarino P, Aslan M, Anderson D, LaFleur R, Hammond T, Schaa K, Moser J, Huang G, Muralidhar S, Przygodzki R, O’Leary TJ. Million Veteran Program: A mega-biobank to study genetic influences on health and disease. J Clin Epidemiol. 2016;70:214–223.

10. Sullivan PF, Agrawal A, Bulik CM, Andreassen OA, Borglum AD, Breen G, Cichon S, Edenberg HJ, Faraone SV, Gelernter J, Mathews CA, Nievergelt CM, Smoller JW, O’Donovan MC, Psychiatric Genomics C. Psychiatric Genomics: An Update and an Agenda. Am J Psychiatry. 2018;175:15–27.

11. Martin AR, Gignoux CR, Walters RK, Wojcik GL, Neale BM, Gravel S, Daly MJ, Bustamante CD, Kenny EE. Human Demographic History Impacts Genetic Risk Prediction across Diverse Populations. Am J Hum Genet. 2017;100:635–649.

12. Docherty AR, Shabalin AA, DiBlasi E, Monson E, Mullins N, Adkins DE, Bacanu SA, Bakian AV, Crowell S, Chen D, Darlington TM, Callor WB, Christensen ED, Gray D, Keeshin B, Klein M, Anderson JS, Jerominski L, Hayward C, Porteous DJ, McIntosh A, Li Q, Coon H. Genome-Wide Association Study of Suicide Death and Polygenic Prediction of Clinical Antecedents. Am J Psychiatry. 2020;177:917–927.

13. Hoffmire C, Stephens B, Morley S, Thompson C, Kemp J, Bossarte RM. VA Suicide Prevention Applications Network: A National Health Care System-Based Suicide Event Tracking System. Public Health Rep. 2016;131:816–821.

14. Bulik-Sullivan BK, Loh PR, Finucane HK, Ripke S, Yang J, Schizophrenia Working Group of the Psychiatric Genomics C, Patterson N, Daly MJ, Price AL, Neale BM. LD Score regression distinguishes confounding from polygenicity in genome-wide association studies. Nat Genet. 2015;47:291–295.

15. Fang H, Hui Q, Lynch J, Honerlaw J, Assimes TL, Huang J, Vujkovic M, Damrauer SM, Pyarajan S, Gaziano JM, DuVall SL, O’Donnell CJ, Cho K, Chang KM, Wilson PWF, Tsao PS, Program VAMV, Sun YV, Tang H. Harmonizing Genetic Ancestry and Self-identified Race/Ethnicity in Genome-wide Association Studies. Am J Hum Genet. 2019;105:763–772.

16. Chang CC, Chow CC, Tellier LC, Vattikuti S, Purcell SM, Lee JJ. Second-generation PLINK: rising to the challenge of larger and richer datasets. Gigascience. 2015;4:7.

17. Willer CJ, Li Y, Abecasis GR. METAL: fast and efficient meta-analysis of genomewide association scans. Bioinformatics. 2010;26:2190–2191.

18. Luo Y, Li X, Wang X, Gazal S, Mercader JM, Me Research T, Consortium STD, Neale BM, Florez JC, Auton A, Price AL, Finucane HK, Raychaudhuri S. Estimating heritability and its enrichment in tissue-specific gene sets in admixed populations. Hum Mol Genet. 2021;30:1521–1534.

19. Karczewski KJ, Francioli LC, Tiao G, Cummings BB, Alfoldi J, Wang Q, Collins RL, Laricchia KM, Ganna A, Birnbaum DP, Gauthier LD, Brand H, Solomonson M, Watts NA, Rhodes D, Singer-Berk M, England EM, Seaby EG, Kosmicki JA, Walters RK, Tashman K, Farjoun Y, Banks E, Poterba T, Wang A, Seed C, Whiffin N, Chong JX, Samocha KE, Pierce-Hoffman E, Zappala Z, O’Donnell-Luria AH, Minikel EV, Weisburd B, Lek M, Ware JS, Vittal C, Armean IM, Bergelson L, Cibulskis K, Connolly KM, Covarrubias M, Donnelly S, Ferriera S, Gabriel S, Gentry J, Gupta N, Jeandet T, Kaplan D, Llanwarne C, Munshi R, Novod S, Petrillo N, Roazen D, Ruano-Rubio V, Saltzman A, Schleicher M, Soto J, Tibbetts K, Tolonen C, Wade G, Talkowski ME, Genome Aggregation Database C, Neale BM, Daly MJ, MacArthur DG. The mutational constraint spectrum quantified from variation in 141,456 humans. Nature. 2020;581:434–443.

20. Nock MK, Borges G, Bromet EJ, Alonso J, Angermeyer M, Beautrais A, Bruffaerts R, Chiu WT, de Girolamo G, Gluzman S, de Graaf R, Gureje O, Haro JM, Huang Y, Karam E, Kessler RC, Lepine JP, Levinson D, Medina-Mora ME, Ono Y, Posada-Villa J, Williams D. Cross-national prevalence and risk factors for suicidal ideation, plans and attempts. Br J Psychiatry. 2008;192:98–105.

21. Brown BC, Asian Genetic Epidemiology Network Type 2 Diabetes C, Ye CJ, Price AL, Zaitlen N. Transethnic Genetic-Correlation Estimates from Summary Statistics. Am J Hum Genet. 2016;99:76–88.

22. Campos AI, Verweij KJH, Statham DJ, Madden PAF, Maciejewski DF, Davis KAS, John A, Hotopf M, Heath AC, Martin NG, Renteria ME. Genetic aetiology of self-harm ideation and behaviour. Sci Rep. 2020;10:9713.

23. Demontis D, Walters RK, Martin J, Mattheisen M, Als TD, Agerbo E, Baldursson G, Belliveau R, Bybjerg-Grauholm J, Baekvad-Hansen M, Cerrato F, Chambert K, Churchhouse C, Dumont A, Eriksson N, Gandal M, Goldstein JI, Grasby KL, Grove J, Gudmundsson OO, Hansen CS, Hauberg ME, Hollegaard MV, Howrigan DP, Huang H, Maller JB, Martin AR, Martin NG, Moran J, Pallesen J, Palmer DS, Pedersen CB, Pedersen MG, Poterba T, Poulsen JB, Ripke S, Robinson EB, Satterstrom FK, Stefansson H, Stevens C, Turley P, Walters GB, Won H, Wright MJ, Consortium AWGotPG, Early L, Genetic Epidemiology C, andMe Research T, Andreassen OA, Asherson P, Burton CL, Boomsma DI, Cormand B, Dalsgaard S, Franke B, Gelernter J, Geschwind D, Hakonarson H, Haavik J, Kranzler HR, Kuntsi J, Langley K, Lesch KP, Middeldorp C, Reif A, Rohde LA, Roussos P, Schachar R, Sklar P, Sonuga-Barke EJS, Sullivan PF, Thapar A, Tung JY, Waldman ID, Medland SE, Stefansson K, Nordentoft M, Hougaard DM, Werge T, Mors O, Mortensen PB, Daly MJ, Faraone SV, Borglum AD, Neale BM. Discovery of the first genome-wide significant risk loci for attention deficit/hyperactivity disorder. Nat Genet. 2019;51:63–75.

24. Grove J, Ripke S, Als TD, Mattheisen M, Walters RK, Won H, Pallesen J, Agerbo E, Andreassen OA, Anney R, Awashti S, Belliveau R, Bettella F, Buxbaum JD, Bybjerg-Grauholm J, Baekvad-Hansen M, Cerrato F, Chambert K, Christensen JH, Churchhouse C, Dellenvall K, Demontis D, De Rubeis S, Devlin B, Djurovic S, Dumont AL, Goldstein JI, Hansen CS, Hauberg ME, Hollegaard MV, Hope S, Howrigan DP, Huang H, Hultman CM, Klei L, Maller J, Martin J, Martin AR, Moran JL, Nyegaard M, Naerland T, Palmer DS, Palotie A, Pedersen CB, Pedersen MG, dPoterba T, Poulsen JB, Pourcain BS, Qvist P, Rehnstrom K, Reichenberg A, Reichert J, Robinson EB, Roeder K, Roussos P, Saemundsen E, Sandin S, Satterstrom FK, Davey Smith G, Stefansson H, Steinberg S, Stevens CR, Sullivan PF, Turley P, Walters GB, Xu X, Autism Spectrum Disorder Working Group of the Psychiatric Genomics C, Bupgen, Major Depressive Disorder Working Group of the Psychiatric Genomics C, andMe Research T, Stefansson K, Geschwind DH, Nordentoft M, Hougaard DM, Werge T, Mors O, Mortensen PB, Neale BM, Daly MJ, Borglum AD. Identification of common genetic risk variants for autism spectrum disorder. Nat Genet. 2019;51:431–444.

25. Karlsson Linner R, Biroli P, Kong E, Meddens SFW, Wedow R, Fontana MA, Lebreton M, Tino SP, Abdellaoui A, Hammerschlag AR, Nivard MG, Okbay A, Rietveld CA, Timshel PN, Trzaskowski M, Vlaming R, Zund CL, Bao Y, Buzdugan L, Caplin AH, Chen CY, Eibich P, Fontanillas P, Gonzalez JR, Joshi PK, Karhunen V, Kleinman A, Levin RZ, Lill CM, Meddens GA, Muntane G, Sanchez-Roige S, Rooij FJV, Taskesen E, Wu Y, Zhang F, and Me Research T, e QC, International Cannabis C, Social Science Genetic Association C, Auton A, Boardman JD, Clark DW, Conlin A, Dolan CC, Fischbacher U, Groenen PJF, Harris KM, Hasler G, Hofman A, Ikram MA, Jain S, Karlsson R, Kessler RC, Kooyman M, MacKillop J, Mannikko M, Morcillo-Suarez C, McQueen MB, Schmidt KM, Smart MC, Sutter M, Thurik AR, Uitterlinden AG, White J, Wit H, Yang J, Bertram L, Boomsma DI, Esko T, Fehr E, Hinds DA, Johannesson M, Kumari M, Laibson D, Magnusson PKE, Meyer MN, Navarro A, Palmer AA, Pers TH, Posthuma D, Schunk D, Stein MB, Svento R, Tiemeier H, Timmers P, Turley P, Ursano RJ, Wagner GG, Wilson JF, Gratten J, Lee JJ, Cesarini D, Benjamin DJ, Koellinger PD, Beauchamp JP. Genome-wide association analyses of risk tolerance and risky behaviors in over 1 million individuals identify hundreds of loci and shared genetic influences. Nat Genet. 2019;51:245–257.

26. Nagel M, Jansen PR, Stringer S, Watanabe K, de Leeuw CA, Bryois J, Savage JE, Hammerschlag AR, Skene NG, Munoz-Manchado AB, andMe Research T, White T, Tiemeier H, Linnarsson S, Hjerling-Leffler J, Polderman TJC, Sullivan PF, van der Sluis S, Posthuma D. Meta-analysis of genome-wide association studies for neuroticism in 449,484 individuals identifies novel genetic loci and pathways. Nat Genet. 2018;50:920–927.

27. Nievergelt CM, Maihofer AX, Klengel T, Atkinson EG, Chen CY, Choi KW, Coleman JRI, Dalvie S, Duncan LE, Gelernter J, Levey DF, Logue MW, Polimanti R, Provost AC, Ratanatharathorn A, Stein MB, Torres K, Aiello AE, Almli LM, Amstadter AB, Andersen SB, Andreassen OA, Arbisi PA, Ashley-Koch AE, Austin SB, Avdibegovic E, Babic D, Baekvad-Hansen M, Baker DG, Beckham JC, Bierut LJ, Bisson JI, Boks MP, Bolger EA, Borglum AD, Bradley B, Brashear M, Breen G, Bryant RA, Bustamante AC, Bybjerg-Grauholm J, Calabrese JR, Caldas-de-Almeida JM, Dale AM, Daly MJ, Daskalakis NP, Deckert J, Delahanty DL, Dennis MF, Disner SG, Domschke K, Dzubur-Kulenovic A, Erbes CR, Evans A, Farrer LA, Feeny NC, Flory JD, Forbes D, Franz CE, Galea S, Garrett ME, Gelaye B, Geuze E, Gillespie C, Uka AG, Gordon SD, Guffanti G, Hammamieh R, Harnal S, Hauser MA, Heath AC, Hemmings SMJ, Hougaard DM, Jakovljevic M, Jett M, Johnson EO, Jones I, Jovanovic T, Qin XJ, Junglen AG, Karstoft KI, Kaufman ML, Kessler RC, Khan A, Kimbrel NA, King AP, Koen N, Kranzler HR, Kremen WS, Lawford BR, Lebois LAM, Lewis CE, Linnstaedt SD, Lori A, Lugonja B, Luykx JJ, Lyons MJ, Maples-Keller J, Marmar C, Martin AR, Martin NG, Maurer D, Mavissakalian MR, McFarlane A, McGlinchey RE, McLaughlin KA, McLean SA, McLeay S, Mehta D, Milberg WP, Miller MW, Morey RA, Morris CP, Mors O, Mortensen PB, Neale BM, Nelson EC, Nordentoft M, Norman SB, O’Donnell M, Orcutt HK, Panizzon MS, Peters ES, Peterson AL, Peverill M, Pietrzak RH, Polusny MA, Rice JP, Ripke S, Risbrough VB, Roberts AL, Rothbaum AO, Rothbaum BO, Roy-Byrne P, Ruggiero K, Rung A, Rutten BPF, Saccone NL, Sanchez SE, Schijven D, Seedat S, Seligowski AV, Seng JS, Sheerin CM, Silove D, Smith AK, Smoller JW, Sponheim SR, Stein DJ, Stevens JS, Sumner JA, Teicher MH, Thompson WK, Trapido E, Uddin M, Ursano RJ, van den Heuvel LL, Van Hooff M, Vermetten E, Vinkers CH, Voisey J, Wang Y, Wang Z, Werge T, Williams MA, Williamson DE, Winternitz S, Wolf C, Wolf EJ, Wolff JD, Yehuda R, Young RM, Young KA, Zhao H, Zoellner LA, Liberzon I, Ressler KJ, Haas M, Koenen KC. International meta-analysis of PTSD genome-wide association studies identifies sex- and ancestry-specific genetic risk loci. Nat Commun. 2019;10:4558.

28. Stahl EA, Breen G, Forstner AJ, McQuillin A, Ripke S, Trubetskoy V, Mattheisen M, Wang Y, Coleman JRI, Gaspar HA, de Leeuw CA, Steinberg S, Pavlides JMW, Trzaskowski M, Byrne EM, Pers TH, Holmans PA, Richards AL, Abbott L, Agerbo E, Akil H, Albani D, Alliey-Rodriguez N, Als TD, Anjorin A, Antilla V, Awasthi S, Badner JA, Baekvad-Hansen M, Barchas JD, Bass N, Bauer M, Belliveau R, Bergen SE, Pedersen CB, Boen E, Boks MP, Boocock J, Budde M, Bunney W, Burmeister M, Bybjerg-Grauholm J, Byerley W, Casas M, Cerrato F, Cervantes P, Chambert K, Charney AW, Chen D, Churchhouse C, Clarke TK, Coryell W, Craig DW, Cruceanu C, Curtis D, Czerski PM, Dale AM, de Jong S, Degenhardt F, Del-Favero J, DePaulo JR, Djurovic S, Dobbyn AL, Dumont A, Elvsashagen T, Escott-Price V, Fan CC, Fischer SB, Flickinger M, Foroud TM, Forty L, Frank J, Fraser C, Freimer NB, Frisen L, Gade K, Gage D, Garnham J, Giambartolomei C, Pedersen MG, Goldstein J, Gordon SD, Gordon-Smith K, Green EK, Green MJ, Greenwood TA, Grove J, Guan W, Guzman-Parra J, Hamshere ML, Hautzinger M, Heilbronner U, Herms S, Hipolito M, Hoffmann P, Holland D, Huckins L, Jamain S, Johnson JS, Jureus A, Kandaswamy R, Karlsson R, Kennedy JL, Kittel-Schneider S, Knowles JA, Kogevinas M, Koller AC, Kupka R, Lavebratt C, Lawrence J, Lawson WB, Leber M, Lee PH, Levy SE, Li JZ, Liu C, Lucae S, Maaser A, MacIntyre DJ, Mahon PB, Maier W, Martinsson L, McCarroll S, McGuffin P, McInnis MG, McKay JD, Medeiros H, Medland SE, Meng F, Milani L, Montgomery GW, Morris DW, Muhleisen TW, Mullins N, Nguyen H, Nievergelt CM, Adolfsson AN, Nwulia EA, O’Donovan C, Loohuis LMO, Ori APS, Oruc L, Osby U, Perlis RH, Perry A, Pfennig A, Potash JB, Purcell SM, Regeer EJ, Reif A, Reinbold CS, Rice JP, Rivas F, Rivera M, Roussos P, Ruderfer DM, Ryu E, Sanchez-Mora C, Schatzberg AF, Scheftner WA, Schork NJ, Shannon Weickert C, Shehktman T, Shilling PD, Sigurdsson E, Slaney C, Smeland OB, Sobell JL, Soholm Hansen C, Spijker AT, St Clair D, Steffens M, Strauss JS, Streit F, Strohmaier J, Szelinger S, Thompson RC, Thorgeirsson TE, Treutlein J, Vedder H, Wang W, Watson SJ, Weickert TW, Witt SH, Xi S, Xu W, Young AH, Zandi P, Zhang P, Zollner S, e QC, Consortium B, Adolfsson R, Agartz I, Alda M, Backlund L, Baune BT, Bellivier F, Berrettini WH, Biernacka JM, Blackwood DHR, Boehnke M, Borglum AD, Corvin A, Craddock N, Daly MJ, Dannlowski U, Esko T, Etain B, Frye M, Fullerton JM, Gershon ES, Gill M, Goes F, Grigoroiu-Serbanescu M, Hauser J, Hougaard DM, Hultman CM, Jones I, Jones LA, Kahn RS, Kirov G, Landen M, Leboyer M, Lewis CM, Li QS, Lissowska J, Martin NG, Mayoral F, McElroy SL, McIntosh AM, McMahon FJ, Melle I, Metspalu A, Mitchell PB, Morken G, Mors O, Mortensen PB, Muller-Myhsok B, Myers RM, Neale BM, Nimgaonkar V, Nordentoft M, Nothen MM, O’Donovan MC, Oedegaard KJ, Owen MJ, Paciga SA, Pato C, Pato MT, Posthuma D, Ramos-Quiroga JA, Ribases M, Rietschel M, Rouleau GA, Schalling M, Schofield PR, Schulze TG, Serretti A, Smoller JW, Stefansson H, Stefansson K, Stordal E, Sullivan PF, Turecki G, Vaaler AE, Vieta E, Vincent JB, Werge T, Nurnberger JI, Wray NR, Di Florio A, Edenberg HJ, Cichon S, Ophoff RA, Scott LJ, Andreassen OA, Kelsoe J, Sklar P, Bipolar Disorder Working Group of the Psychiatric Genomics C. Genome-wide association study identifies 30 loci associated with bipolar disorder. Nat Genet. 2019;51:793–803.

29. Yengo L, Sidorenko J, Kemper KE, Zheng Z, Wood AR, Weedon MN, Frayling TM, Hirschhorn J, Yang J, Visscher PM, Consortium G. Meta-analysis of genome-wide association studies for height and body mass index in approximately 700000 individuals of European ancestry. Hum Mol Genet. 2018;27:3641–3649.

30. Johnston KJA, Adams MJ, Nicholl BI, Ward J, Strawbridge RJ, Ferguson A, McIntosh AM, Bailey MES, Smith DJ. Genome-wide association study of multisite chronic pain in UK Biobank. PLoS Genet. 2019;15:e1008164.

31. Liu M, Jiang Y, Wedow R, Li Y, Brazel DM, Chen F, Datta G, Davila-Velderrain J, McGuire D, Tian C, Zhan X, andMe Research T, Psychiatry HA-I, Choquet H, Docherty AR, Faul JD, Foerster JR, Fritsche LG, Gabrielsen ME, Gordon SD, Haessler J, Hottenga JJ, Huang H, Jang SK, Jansen PR, Ling Y, Magi R, Matoba N, McMahon G, Mulas A, Orru V, Palviainen T, Pandit A, Reginsson GW, Skogholt AH, Smith JA, Taylor AE, Turman C, Willemsen G, Young H, Young KA, Zajac GJM, Zhao W, Zhou W, Bjornsdottir G, Boardman JD, Boehnke M, Boomsma DI, Chen C, Cucca F, Davies GE, Eaton CB, Ehringer MA, Esko T, Fiorillo E, Gillespie NA, Gudbjartsson DF, Haller T, Harris KM, Heath AC, Hewitt JK, Hickie IB, Hokanson JE, Hopfer CJ, Hunter DJ, Iacono WG, Johnson EO, Kamatani Y, Kardia SLR, Keller MC, Kellis M, Kooperberg C, Kraft P, Krauter KS, Laakso M, Lind PA, Loukola A, Lutz SM, Madden PAF, Martin NG, McGue M, McQueen MB, Medland SE, Metspalu A, Mohlke KL, Nielsen JB, Okada Y, Peters U, Polderman TJC, Posthuma D, Reiner AP, Rice JP, Rimm E, Rose RJ, Runarsdottir V, Stallings MC, Stancakova A, Stefansson H, Thai KK, Tindle HA, Tyrfingsson T, Wall TL, Weir DR, Weisner C, Whitfield JB, Winsvold BS, Yin J, Zuccolo L, Bierut LJ, Hveem K, Lee JJ, Munafo MR, Saccone NL, Willer CJ, Cornelis MC, David SP, Hinds DA, Jorgenson E, Kaprio J, Stitzel JA, Stefansson K, Thorgeirsson TE, Abecasis G, Liu DJ, Vrieze S. Association studies of up to 1.2 million individuals yield new insights into the genetic etiology of tobacco and alcohol use. Nat Genet. 2019;51:237–244.

32. Howard DM, Adams MJ, Clarke TK, Hafferty JD, Gibson J, Shirali M, Coleman JRI, Hagenaars SP, Ward J, Wigmore EM, Alloza C, Shen X, Barbu MC, Xu EY, Whalley HC, Marioni RE, Porteous DJ, Davies G, Deary IJ, Hemani G, Berger K, Teismann H, Rawal R, Arolt V, Baune BT, Dannlowski U, Domschke K, Tian C, Hinds DA, and Me Research T, Major Depressive Disorder Working Group of the Psychiatric Genomics C, Trzaskowski M, Byrne EM, Ripke S, Smith DJ, Sullivan PF, Wray NR, Breen G, Lewis CM, McIntosh AM. Genome-wide meta-analysis of depression identifies 102 independent variants and highlights the importance of the prefrontal brain regions. Nat Neurosci. 2019;22:343–352.

33. Jansen PR, Watanabe K, Stringer S, Skene N, Bryois J, Hammerschlag AR, de Leeuw CA, Benjamins JS, Munoz-Manchado AB, Nagel M, Savage JE, Tiemeier H, White T, andMe Research T, Tung JY, Hinds DA, Vacic V, Wang X, Sullivan PF, van der Sluis S, Polderman TJC, Smit AB, Hjerling-Leffler J, Van Someren EJW, Posthuma D. Genome-wide analysis of insomnia in 1,331,010 individuals identifies new risk loci and functional pathways. Nat Genet. 2019;51:394–403.

34. Zheng J, Erzurumluoglu AM, Elsworth BL, Kemp JP, Howe L, Haycock PC, Hemani G, Tansey K, Laurin C, Early G, Lifecourse Epidemiology Eczema C, Pourcain BS, Warrington NM, Finucane HK, Price AL, Bulik-Sullivan BK, Anttila V, Paternoster L, Gaunt TR, Evans DM, Neale BM. LD Hub: a centralized database and web interface to perform LD score regression that maximizes the potential of summary level GWAS data for SNP heritability and genetic correlation analysis. Bioinformatics. 2017;33:272–279.

35. Ahmedani BK, Peterson EL, Hu Y, Rossom RC, Lynch F, Lu CY, Waitzfelder BE, Owen-Smith AA, Hubley S, Prabhakar D, Williams LK, Zeld N, Mutter E, Beck A, Tolsma D, Simon GE. Major Physical Health Conditions and Risk of Suicide. Am J Prev Med. 2017;53:308–315.

36. Fazel S, Runeson B. Suicide. N Engl J Med. 2020;382:266–274.

37. Hubel C, Gaspar HA, Coleman JRI, Finucane H, Purves KL, Hanscombe KB, Prokopenko I, investigators M, Graff M, Ngwa JS, Workalemahu T, Eating Disorders Working Group of the Psychiatric Genomics C, Major Depressive Disorder Working Group of the Psychiatric Genomics C, Schizophrenia Working Group of the Psychiatric Genomics C, Tourette Syndrome/Obsessive-Compulsive Disorder Working Group of the Psychiatric Genomics C, O’Reilly PF, Bulik CM, Breen G. Genomics of body fat percentage may contribute to sex bias in anorexia nervosa. Am J Med Genet B Neuropsychiatr Genet. 2019;180:428–438.

38. Haworth S, Kho PF, Holgerson PL, Hwang LD, Timpson NJ, Renteria ME, Johansson I, Cuellar-Partida G. Assessment and visualization of phenome-wide causal relationships using genetic data: an application to dental caries and periodontitis. Eur J Hum Genet. 2021;29:300–308.

39. Zhu Z, Zheng Z, Zhang F, Wu Y, Trzaskowski M, Maier R, Robinson MR, McGrath JJ, Visscher PM, Wray NR, Yang J. Causal associations between risk factors and common diseases inferred from GWAS summary data. Nat Commun. 2018;9:224.

40. Yang J, Lee SH, Goddard ME, Visscher PM. GCTA: a tool for genome-wide complex trait analysis. Am J Hum Genet. 2011;88:76–82.

41. Genomes Project C, Auton A, Brooks LD, Durbin RM, Garrison EP, Kang HM, Korbel JO, Marchini JL, McCarthy S, McVean GA, Abecasis GR. A global reference for human genetic variation. Nature. 2015;526:68–74.

42. de Leeuw CA, Mooij JM, Heskes T, Posthuma D. MAGMA: generalized gene-set analysis of GWAS data. PLoS Comput Biol. 2015;11:e1004219.

43. Watanabe K, Taskesen E, van Bochoven A, Posthuma D. Functional mapping and annotation of genetic associations with FUMA. Nat Commun. 2017;8:1826.

44. Consortium GT, Laboratory DA, Coordinating Center -Analysis Working G, Statistical Methods groups-Analysis Working G, Enhancing Gg, Fund NIHC, Nih/Nci, Nih/Nhgri, Nih/Nimh, Nih/Nida, Biospecimen Collection Source Site N, Biospecimen Collection Source Site R, Biospecimen Core Resource V, Brain Bank Repository-University of Miami Brain Endowment B, Leidos Biomedical-Project M, Study E, Genome Browser Data I, Visualization EBI, Genome Browser Data I, Visualization-Ucsc Genomics Institute UoCSC, Lead a, Laboratory DA, Coordinating C, management NIHp, Biospecimen c, Pathology, e QTLmwg, Battle A, Brown CD, Engelhardt BE, Montgomery SB. Genetic effects on gene expression across human tissues. Nature. 2017;550:204–213.

45. Gaspar HA, Breen G. Drug enrichment and discovery from schizophrenia genome-wide association results: an analysis and visualisation approach. Sci Rep. 2017;7:12460.

46. Wagner AH, Coffman AC, Ainscough BJ, Spies NC, Skidmore ZL, Campbell KM, Krysiak K, Pan D, McMichael JF, Eldred JM, Walker JR, Wilson RK, Mardis ER, Griffith M, Griffith OL. DGIdb 2.0: mining clinically relevant drug-gene interactions. Nucleic Acids Res. 2016;44:D1036–1044.

47. Roth BL, Lopez E, Patel S, Kroeze WK. The Multiplicity of Serotonin Receptors: Uselessly Diverse Molecules or an Embarrassment of Riches? The Neuroscientist. 2000;6:252–262.

48. Wu Y, Zeng J, Zhang F, Zhu Z, Qi T, Zheng Z, Lloyd-Jones LR, Marioni RE, Martin NG, Montgomery GW, Deary IJ, Wray NR, Visscher PM, McRae AF, Yang J. Integrative analysis of omics summary data reveals putative mechanisms underlying complex traits. Nat Commun. 2018;9:918.

49. Zhu Z, Zhang F, Hu H, Bakshi A, Robinson MR, Powell JE, Montgomery GW, Goddard ME, Wray NR, Visscher PM, Yang J. Integration of summary data from GWAS and eQTL studies predicts complex trait gene targets. Nat Genet. 2016;48:481–487.

50. de Klein N, Tsai EA, Vochteloo M, Baird D, Huang Y, Chen CY, van Dam S, Oelen R, Deelen P, Bakker OB, El Garwany O, Ouyang Z, Marshall EE, Zavodszky MI, van Rheenen W, Bakker MK, Veldink J, Gaunt TR, Runz H, Franke L, Westra HJ. Brain expression quantitative trait locus and network analyses reveal downstream effects and putative drivers for brain-related diseases. Nat Genet. 2023.

51. Ge T, Chen CY, Ni Y, Feng YA, Smoller JW. Polygenic prediction via Bayesian regression and continuous shrinkage priors. Nat Commun. 2019;10:1776.

52. Sollis E, Mosaku A, Abid A, Buniello A, Cerezo M, Gil L, Groza T, Gunes O, Hall P, Hayhurst J, Ibrahim A, Ji Y, John S, Lewis E, MacArthur JAL, McMahon A, Osumi-Sutherland D, Panoutsopoulou K, Pendlington Z, Ramachandran S, Stefancsik R, Stewart J, Whetzel P, Wilson R, Hindorff L, Cunningham F, Lambert SA, Inouye M, Parkinson H, Harris LW. The NHGRI-EBI GWAS Catalog: knowledgebase and deposition resource. Nucleic Acids Res. 2023;51:D977–D985.

53. Ikeda M, Takahashi A, Kamatani Y, Momozawa Y, Saito T, Kondo K, Shimasaki A, Kawase K, Sakusabe T, Iwayama Y, Toyota T, Wakuda T, Kikuchi M, Kanahara N, Yamamori H, Yasuda Y, Watanabe Y, Hoya S, Aleksic B, Kushima I, Arai H, Takaki M, Hattori K, Kunugi H, Okahisa Y, Ohnuma T, Ozaki N, Someya T, Hashimoto R, Yoshikawa T, Kubo M, Iwata N. Genome-Wide Association Study Detected Novel Susceptibility Genes for Schizophrenia and Shared Trans-Populations/Diseases Genetic Effect. Schizophr Bull. 2019;45:824–834.

54. Pfennig A, Kunzel HE, Kern N, Ising M, Majer M, Fuchs B, Ernst G, Holsboer F, Binder EB. Hypothalamus-pituitary-adrenal system regulation and suicidal behavior in depression. Biol Psychiatry. 2005;57:336–342.

55. Mann JJ, Rizk MM. A Brain-Centric Model of Suicidal Behavior. Am J Psychiatry. 2020;177:902–916.

