## Supplementary Figures for "Genome-wide association study meta-analysis of suicide attempt identifies twelve genome-wide significant loci and implicates genetic risks for specific health factors": Supplementary_Figures_Docherty_ISGC_MVP_SA_GWAS_01012023.docx

S1. Multi-Ancestry GWAS QQ Plot of Suicide Attempt

S2. Multi-Ancestry GWAS Meta-analysis Forest Plot of rs62474683 on Chromosome 7

S3. Multi-Ancestry GWAS Meta-analysis Forest Plot of rs7131627 on Chromosome 11

S4. Multi-Ancestry GWAS Meta-analysis Forest Plot of rs3791129 on Chromosome 1

S5. Multi-Ancestry GWAS Meta-analysis Forest Plot of rs62404522 on Chromosome 6

S6. Multi-Ancestry GWAS Meta-analysis Forest Plot of rs17514846 on Chromosome 15

S7. Multi-Ancestry GWAS Meta-analysis Forest Plot of rs9525171 on Chromosome 13

S8. Multi-Ancestry GWAS Meta-analysis Forest Plot of rs17485141 on Chromosome 12

S9. Multi-Ancestry GWAS Meta-analysis Forest Plot of rs7649709 on Chromosome 3

S10. EUR GWAS Manhattan Plot of Suicide Attempt

S11. Multi-Ancestry GWAS Meta-analysis Forest Plot of rs850261 on Chromosome 14

S12. Multi-Ancestry GWAS Meta-analysis Forest Plot of rs2284000 on Chromosome 22

S13. Multi-Ancestry GWAS Meta-analysis Forest Plot of rs35869525 on Chromosome 6

S14. Multi-Ancestry GWAS Meta-analysis Forest Plot of rs2503185 on Chromosome 1

S15. Multi-Ancestry Regional Plot of Genome-Wide Significant Loci on Chromosome 7

S16. Multi-Ancestry Regional Plot of Genome-Wide Significant Loci on Chromosome 11

S17. Multi-Ancestry Regional Plot of Genome-Wide Significant Loci on Chromosome 1

S18. Multi-Ancestry Regional Plot of Genome-Wide Significant Loci on Chromosome 6

S19. Multi-Ancestry Regional Plot of Genome-Wide Significant Loci on Chromosome 15

S20. Multi-Ancestry Regional Plot of Genome-Wide Significant Loci on Chromosome 13

S21. Multi-Ancestry Regional Plot of Genome-Wide Significant Loci on Chromosome 12

S22. Multi-Ancestry Regional Plot of Genome-Wide Significant Loci on Chromosome 3

S23. EUR Regional Plot of Genome-Wide Significant Loci on Chromosome 14

S24. EUR Regional Plot of Genome-Wide Significant Loci on Chromosome 22

S25. EUR Regional Plot of Genome-Wide Significant Loci on Chromosome 6

S26. EUR Regional Plot of Genome-Wide Significant Loci on Chromosome 1

S27. Multi-Ancestry Gene-Based Manhattan Plot of Suicide Attempt

S28. Multi-Ancestry Gene-Based QQ-Plot of Suicide Attempt

S29. EUR Gene-Based Manhattan Plot of Suicide Attempt

S30. EUR Gene-Based QQ-Plot of Suicide Attempt

S31. Multi-Ancestry MAGMA Tissue Expression Analysis

S32. EUR MAGMA Tissue Expression Analysis

S33: Multi-Ancestry GTex Gene Expression Heatmap

S34: EUR GTex Gene Expression Heatmap

S35: Multi-Ancestry Enrichment of Genes in Gene Sets from GWAS Catalog

S36: EUR Enrichment of Genes in Gene Sets from GWAS Catalog

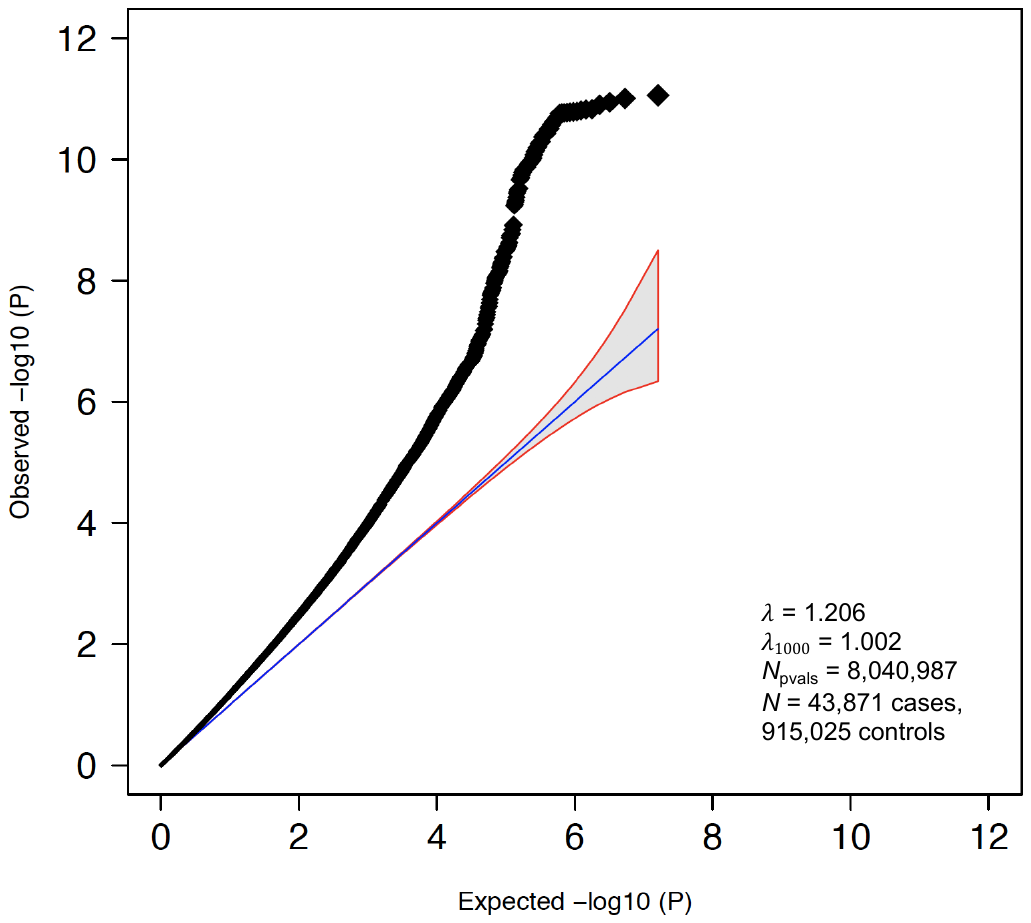

**Figure S1: Multi-Ancestry GWAS QQ Plot of Suicide Attempt.** The y-axis reflects observed p-values. The x-axis is the number of significant p-values expected under *H_0_*. $\lambda$ = lambda, or the genomic inflation factor. $\lambda$_1000_ = the inflation factor for an equivalent study of 1000 cases and 1000 controls.

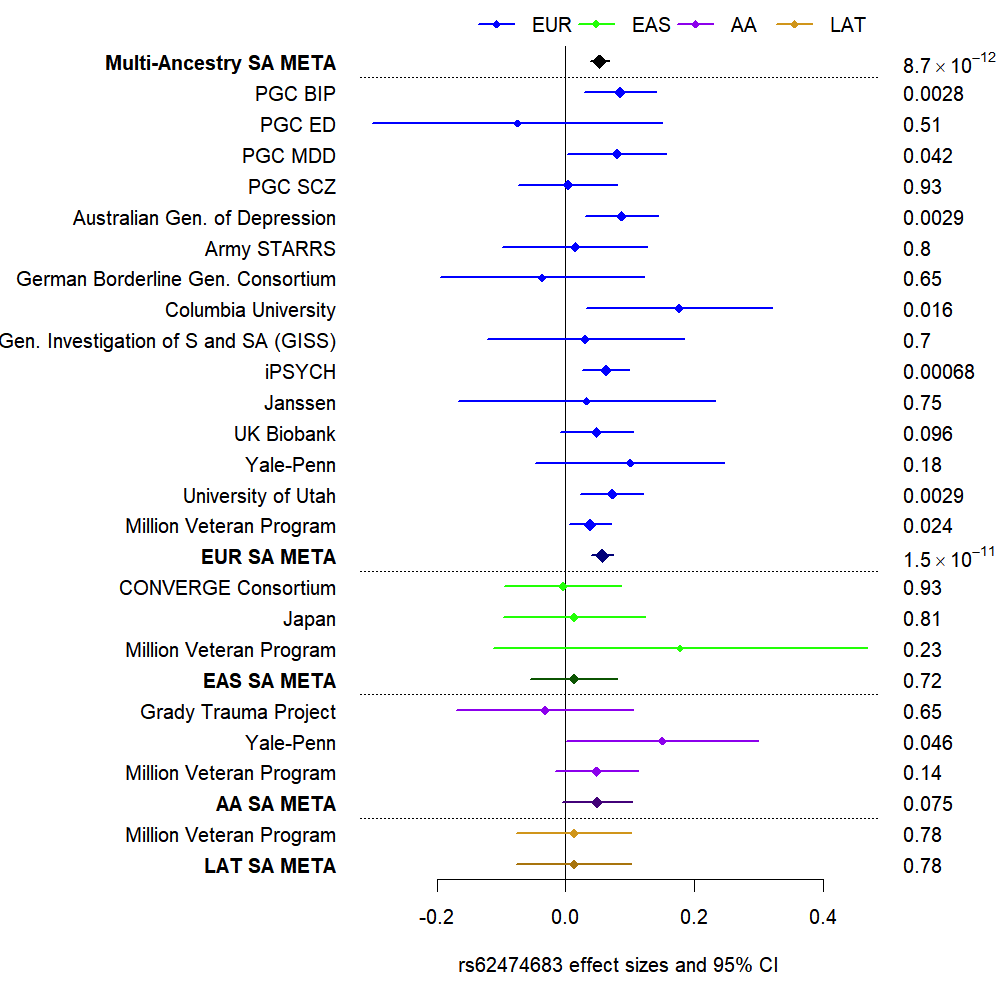

**Figure S2: Multi-Ancestry GWAS Meta-analysis Forest Plot of rs62474683 on Chromosome 7.**

Effect sizes are presented on the x-axis as the log of the odds ratio, ln(OR), of the effect allele on suicide attempt. CI = confidence interval. Cohorts and respective admixture-specific groupings are depicted on the y-axis. The p-values for each test are presented to the right of each cohort ln(OR). EUR = European admixture, EAS = East Asian admixture, AFR = African admixture, LAT = Hispanic/Latino admixture, LAT = Hispanic/Latino admixture, SA = suicide attempt, META = meta-analysis, PGC = Psychiatric Genomics Consortium, MDD = major depressive disorder, BIP = Bipolar disorder, SCZ = schizophrenia, ED = eating disorder.

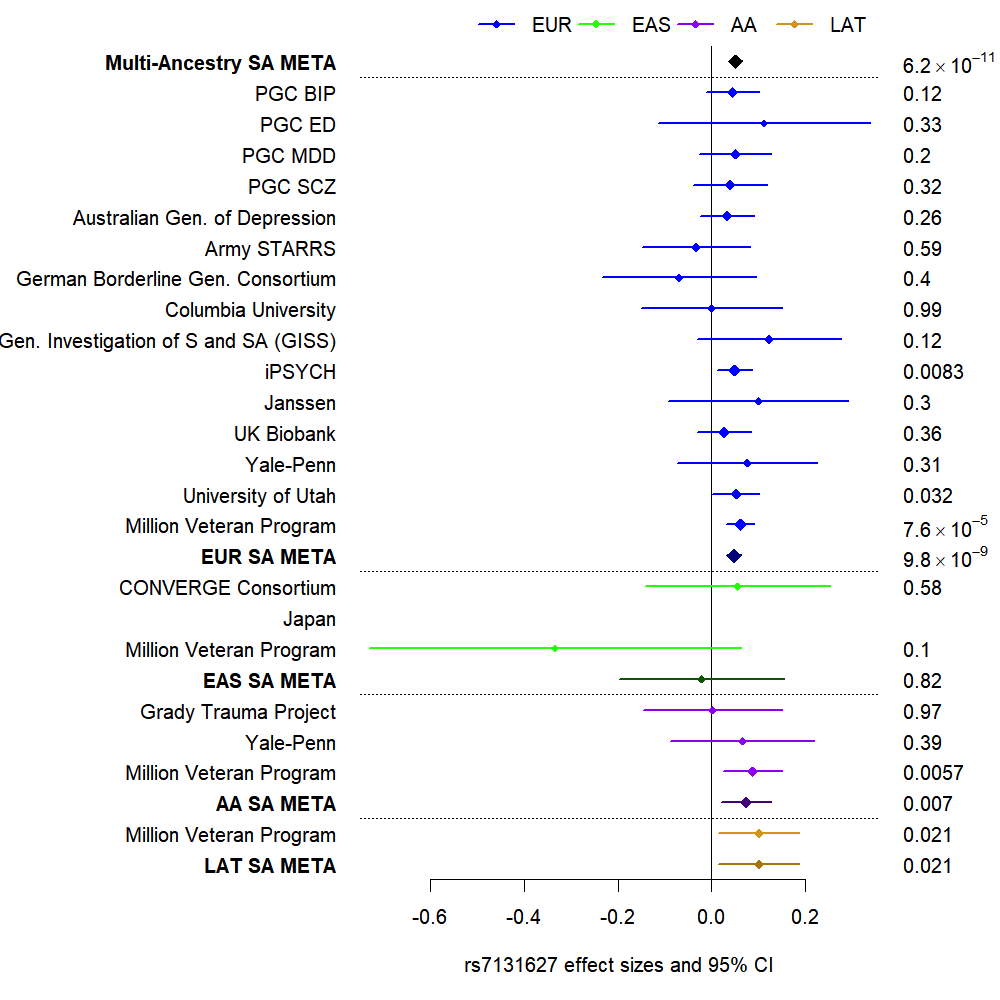

**Figure S3: Multi-Ancestry GWAS Meta-analysis Forest Plot of rs7131627 on Chromosome 11.**

Effect sizes are presented on the x-axis as the log of the odds ratio, ln(OR), of the effect allele on suicide attempt. CI = confidence interval. Cohorts and respective admixture-specific groupings are depicted on the y-axis. The p-values for each test are presented to the right of each cohort ln(OR). EUR = European admixture, EAS = East Asian admixture, AFR = African admixture, LAT = Hispanic/Latino admixture, LAT = Hispanic/Latino admixture, SA = suicide attempt, META = meta-analysis, PGC = Psychiatric Genomics Consortium, MDD = major depressive disorder, BIP = Bipolar disorder, SCZ = schizophrenia, ED = eating disorder.

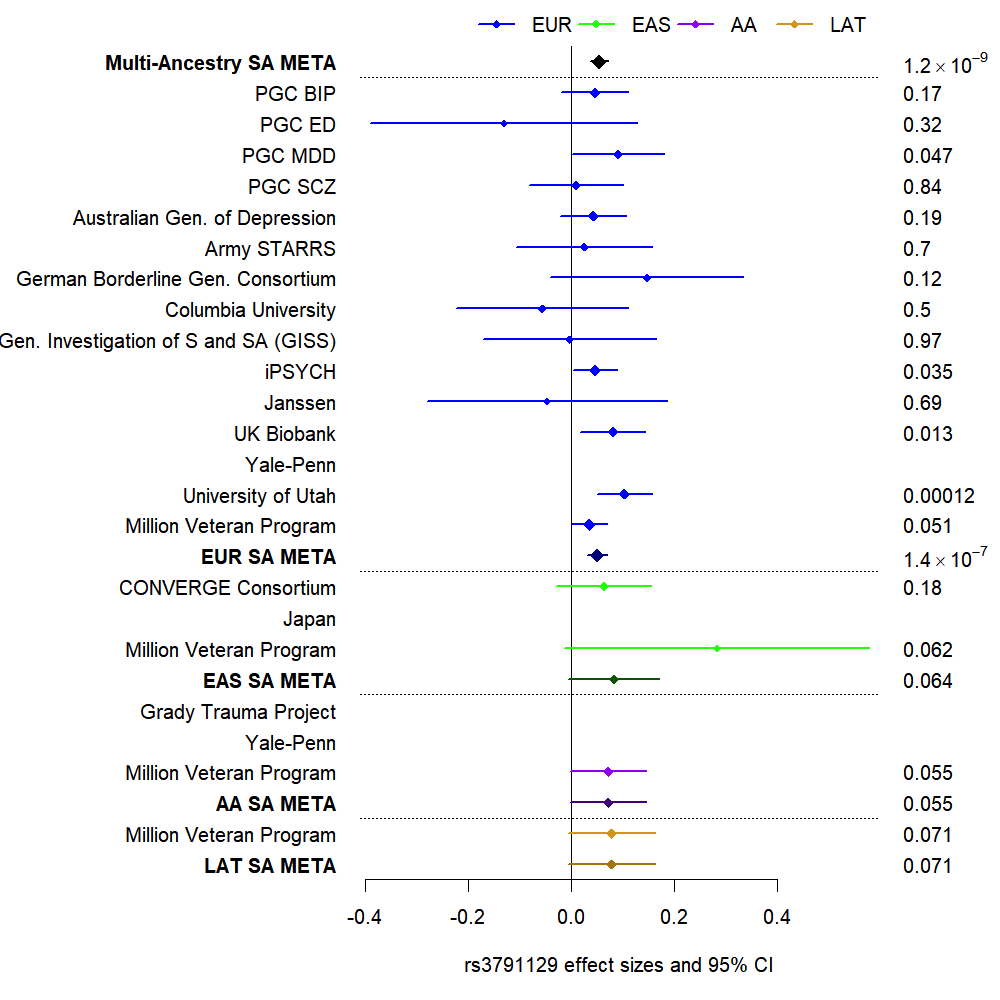

**Figure S4: Multi-Ancestry GWAS Meta-analysis Forest Plot of rs3791129 on Chromosome 1.**

Effect sizes are presented on the x-axis as the log of the odds ratio, ln(OR), of the effect allele on suicide attempt. CI = confidence interval. Cohorts and respective admixture-specific groupings are depicted on the y-axis. The p-values for each test are presented to the right of each cohort ln(OR). EUR = European admixture, EAS = East Asian admixture, AFR = African admixture, LAT = Hispanic/Latino admixture, LAT = Hispanic/Latino admixture, SA = suicide attempt, META = meta-analysis, PGC = Psychiatric Genomics Consortium, MDD = major depressive disorder, BIP = Bipolar disorder, SCZ = schizophrenia, ED = eating disorder.

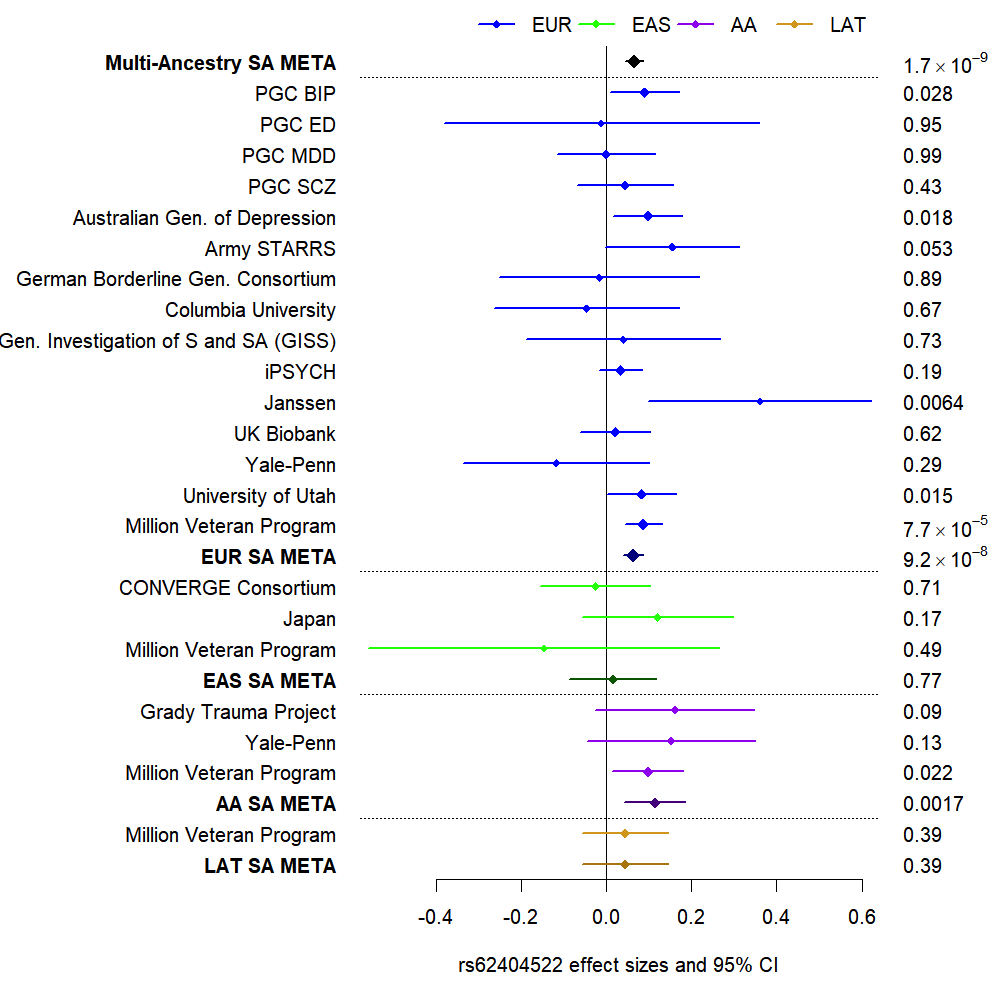

**Figure S5: Multi-Ancestry GWAS Meta-analysis Forest Plot of rs62404522 on Chromosome 6.**

Effect sizes are presented on the x-axis as the log of the odds ratio, ln(OR), of the effect allele on suicide attempt. CI = confidence interval. Cohorts and respective admixture-specific groupings are depicted on the y-axis. The p-values for each test are presented to the right of each cohort ln(OR). EUR = European admixture, EAS = East Asian admixture, AFR = African admixture, LAT = Hispanic/Latino admixture, SA = suicide attempt, META = meta-analysis, PGC = Psychiatric Genomics Consortium, MDD = major depressive disorder, BIP = Bipolar disorder, SCZ = schizophrenia, ED = eating disorder.

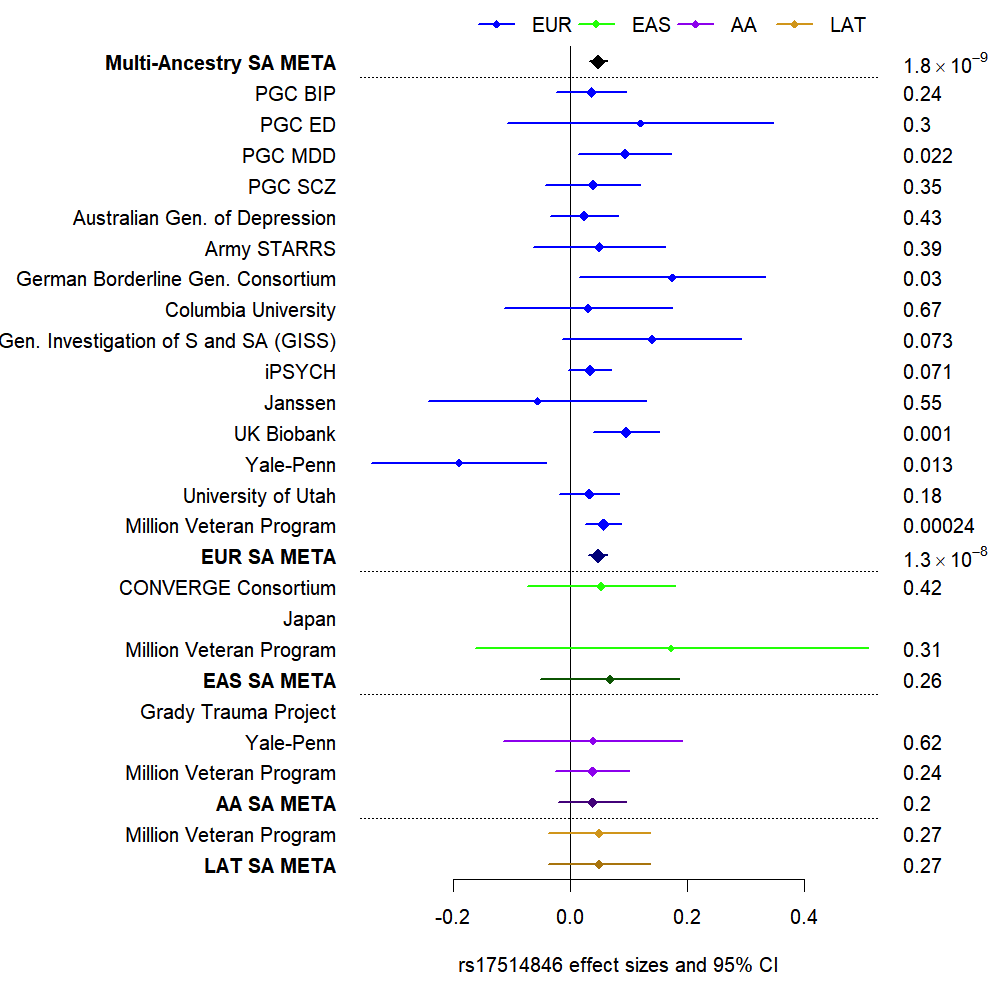

**Figure S6: Multi-Ancestry GWAS Meta-analysis Forest Plot of rs17514846 on Chromosome 15.**

Effect sizes are presented on the x-axis as the log of the odds ratio, ln(OR), of the effect allele on suicide attempt. CI = confidence interval. Cohorts and respective admixture-specific groupings are depicted on the y-axis. The p-values for each test are presented to the right of each cohort ln(OR). EUR = European admixture, EAS = East Asian admixture, AFR = African admixture, LAT = Hispanic/Latino admixture, SA = suicide attempt, META = meta-analysis, PGC = Psychiatric Genomics Consortium, MDD = major depressive disorder, BIP = Bipolar disorder, SCZ = schizophrenia, ED = eating disorder.

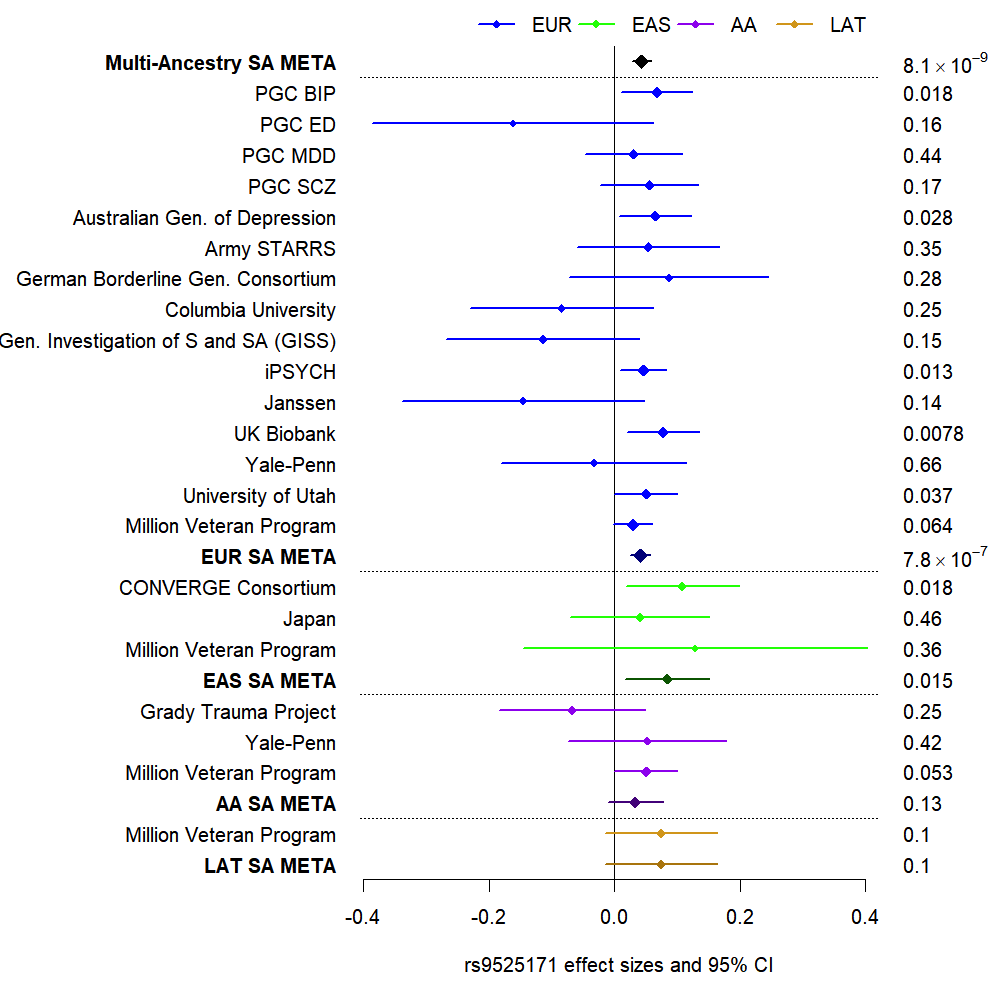

**Figure S7: Multi-Ancestry GWAS Meta-analysis Forest Plot of rs9525171 on Chromosome 13.**

Effect sizes are presented on the x-axis as the log of the odds ratio, ln(OR), of the effect allele on suicide attempt. CI = confidence interval. Cohorts and respective admixture-specific groupings are depicted on the y-axis. The p-values for each test are presented to the right of each cohort ln(OR). EUR = European admixture, EAS = East Asian admixture, AFR = African admixture, LAT = Hispanic/Latino admixture, SA = suicide attempt, META = meta-analysis, PGC = Psychiatric Genomics Consortium, MDD = major depressive disorder, BIP = Bipolar disorder, SCZ = schizophrenia, ED = eating disorder.

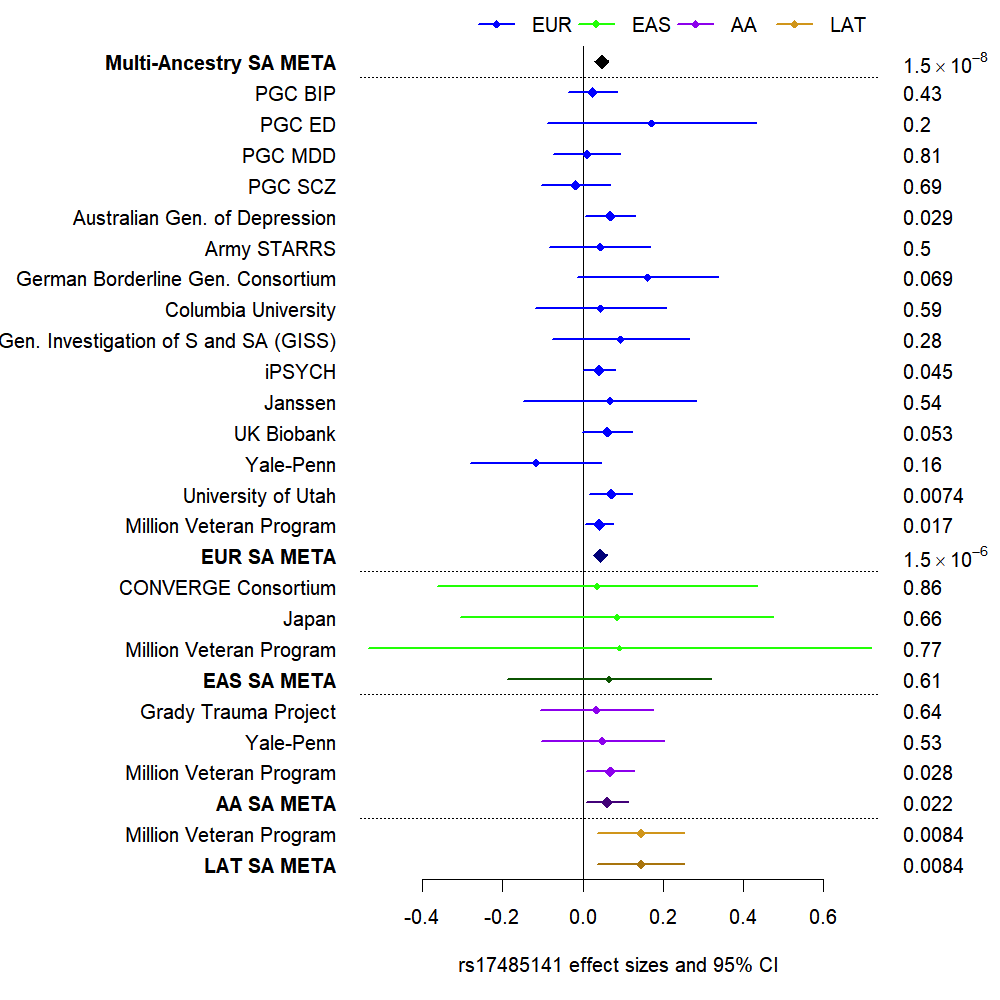

**Figure S8: Multi-Ancestry GWAS Meta-analysis Forest Plot of rs17485141 on Chromosome 12.**

Effect sizes are presented on the x-axis as the log of the odds ratio, ln(OR), of the effect allele on suicide attempt. CI = confidence interval. Cohorts and respective admixture-specific groupings are depicted on the y-axis. The p-values for each test are presented to the right of each cohort ln(OR). EUR = European admixture, EAS = East Asian admixture, AFR = African admixture, LAT = Hispanic/Latino admixture, SA = suicide attempt, META = meta-analysis, PGC = Psychiatric Genomics Consortium, MDD = major depressive disorder, BIP = Bipolar disorder, SCZ = schizophrenia, ED = eating disorder.

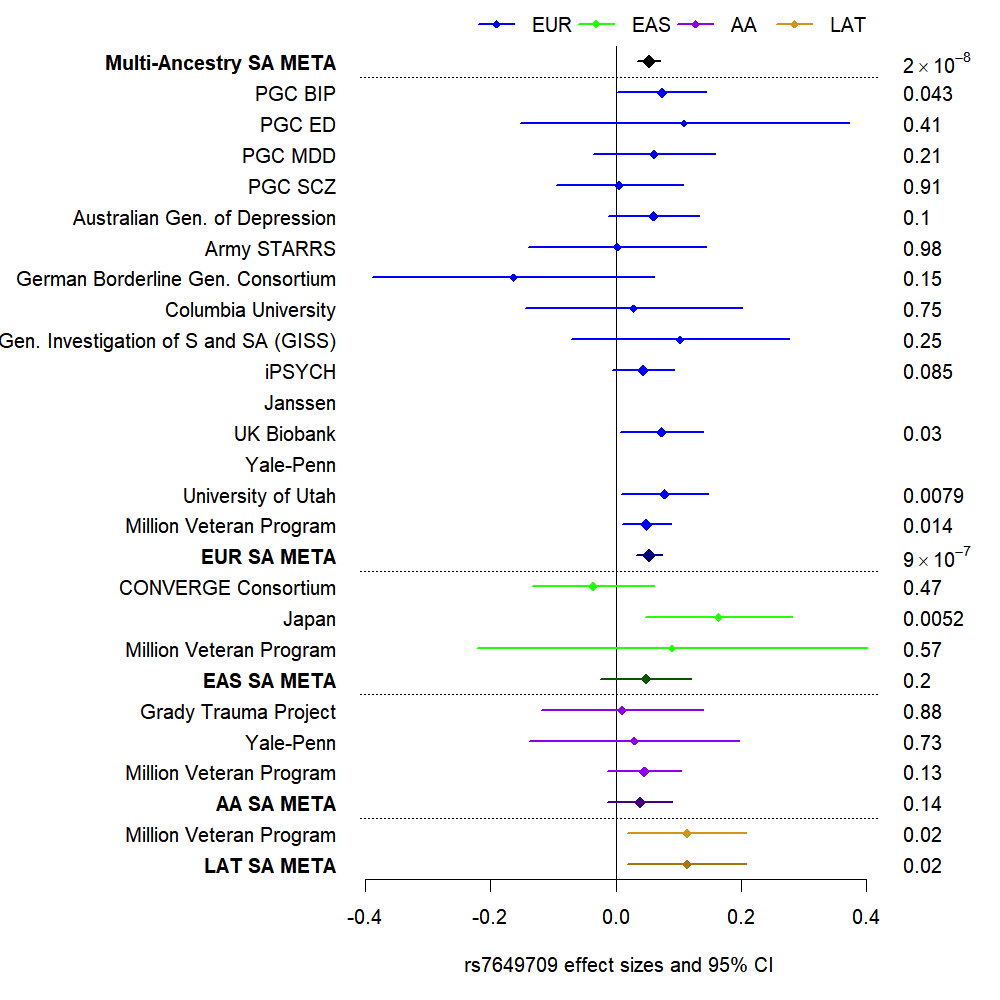

**Figure S9: Multi-Ancestry GWAS Meta-analysis Forest Plot of rs7649709 on Chromosome 3.**

Effect sizes are presented on the x-axis as the log of the odds ratio, ln(OR), of the effect allele on suicide attempt. CI = confidence interval. Cohorts and respective admixture-specific groupings are depicted on the y-axis. The p-values for each test are presented to the right of each cohort ln(OR). EUR = European admixture, EAS = East Asian admixture, AFR = African admixture, LAT = Hispanic/Latino admixture, SA = suicide attempt, META = meta-analysis, PGC = Psychiatric Genomics Consortium, MDD = major depressive disorder, BIP = Bipolar disorder, SCZ = schizophrenia, ED = eating disorder.

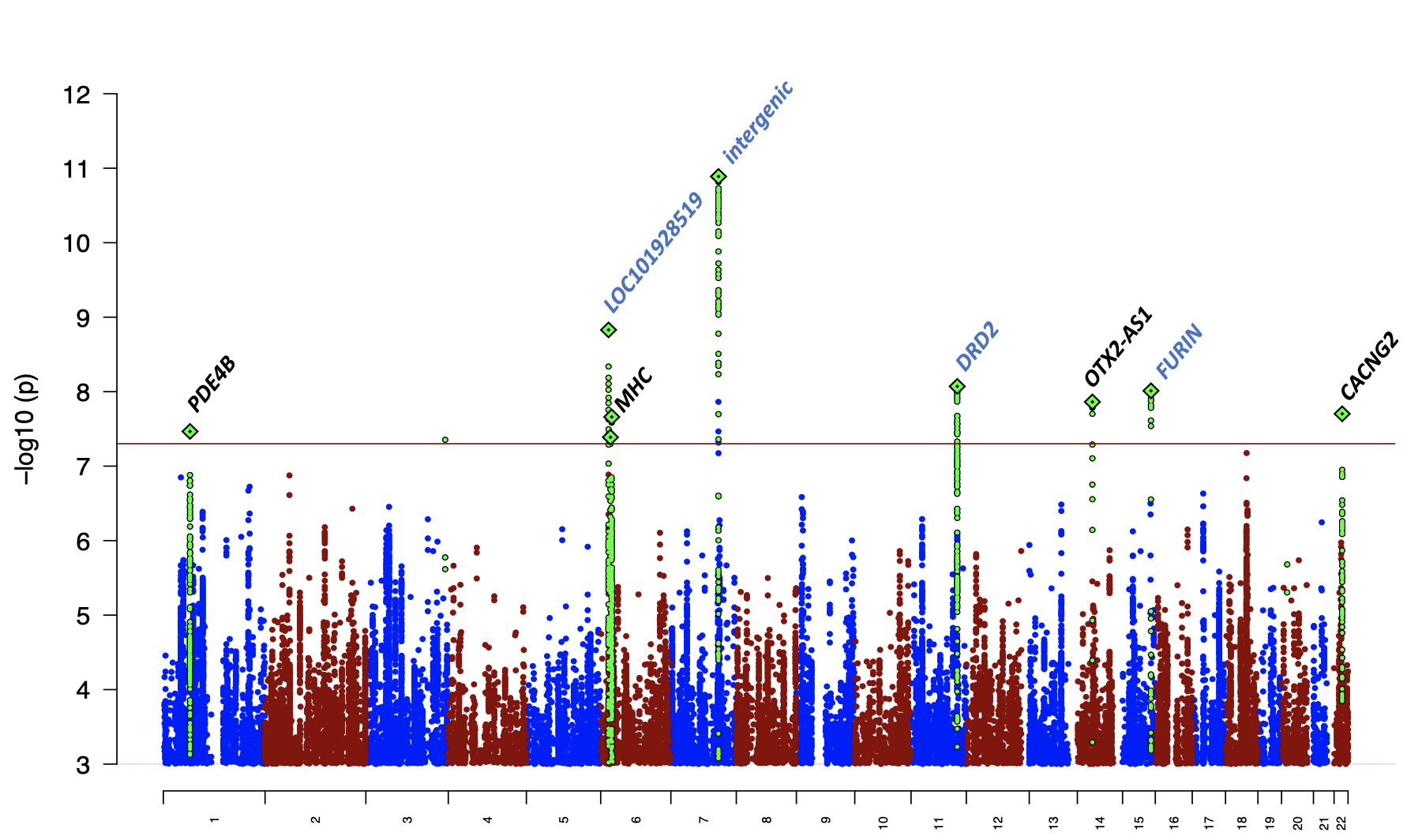

**Figure S10: EUR GWAS Meta-Analysis Manhattan Plot of Suicide Attempt.** The *x-*axis shows genomic position and the *y-*axis shows statistical significance as –log_10_(P value). The horizontal line shows the genome-wide significance threshold (P<5.0x10^-8^). Genes reflect the nearest gene, and regions in blue reflect loci also GWS in multi-ancestry meta-analyses.

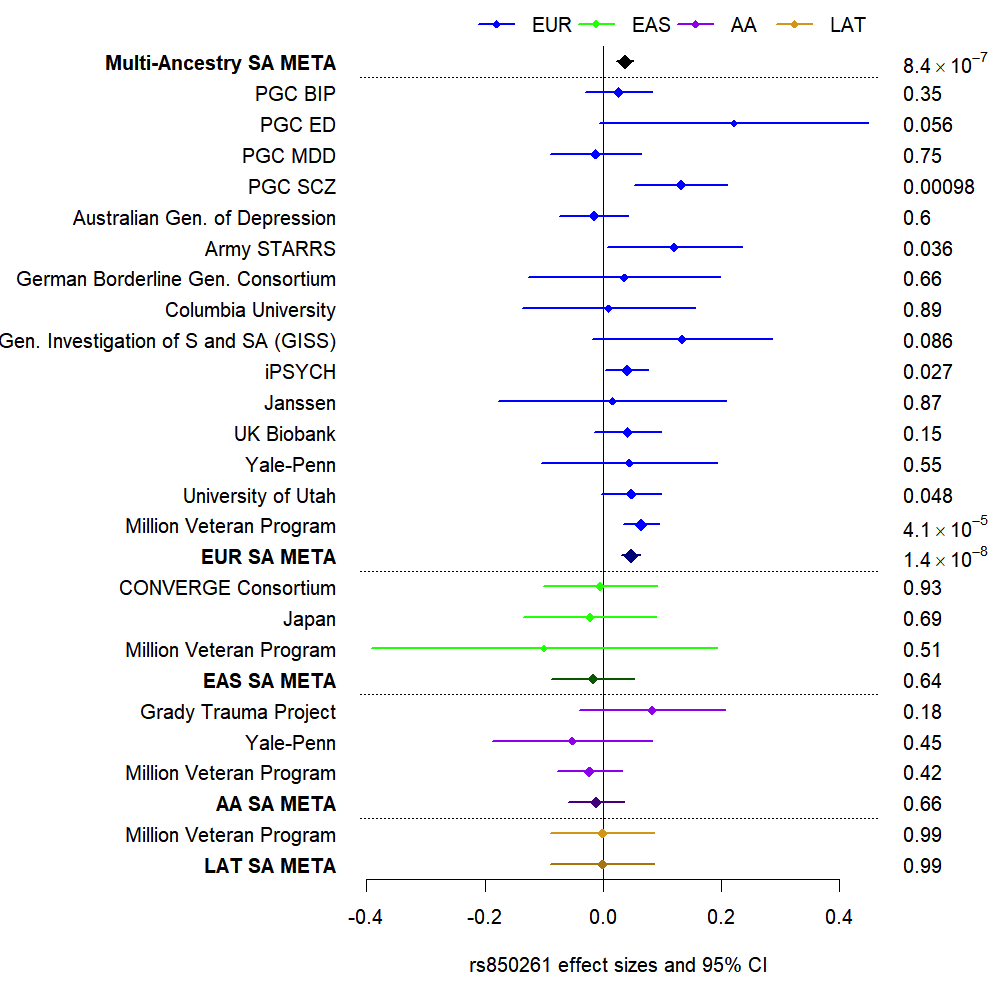

**Figure S11: Multi-Ancestry GWAS Meta-analysis Forest Plot of rs850261 on Chromosome 14.**

Effect sizes are presented on the x-axis as the log of the odds ratio, ln(OR), of the effect allele on suicide attempt. CI = confidence interval. Cohorts and respective admixture-specific groupings are depicted on the y-axis. The p-values for each test are presented to the right of each cohort ln(OR). EUR = European admixture, EAS = East Asian admixture, AFR = African admixture, LAT = Hispanic/Latino admixture, SA = suicide attempt, META = meta-analysis, PGC = Psychiatric Genomics Consortium, MDD = major depressive disorder, BIP = Bipolar disorder, SCZ = schizophrenia, ED = eating disorder.

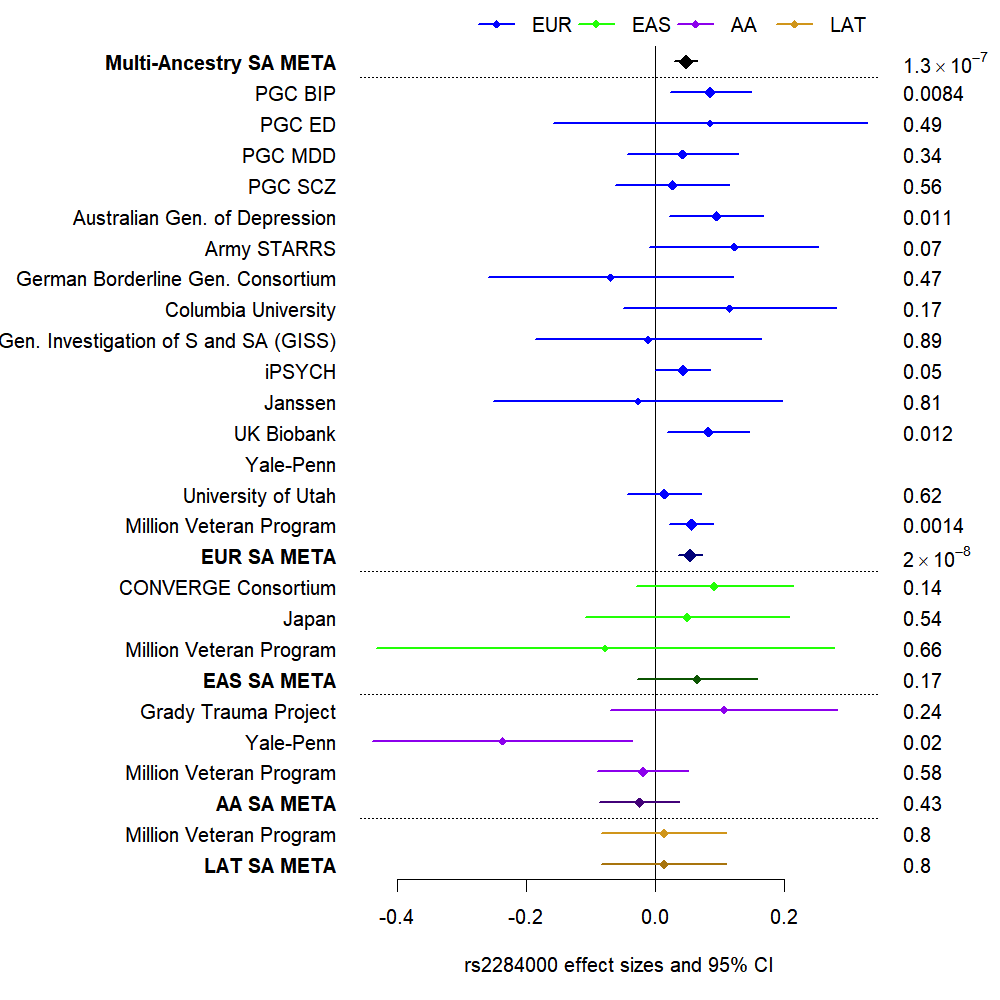

**Figure S12: Multi-Ancestry GWAS Meta-analysis Forest Plot of rs2284000 on Chromosome 22**

Effect sizes are presented on the x-axis as the log of the odds ratio, ln(OR), of the effect allele on suicide attempt. CI = confidence interval. Cohorts and respective admixture-specific groupings are depicted on the y-axis. The p-values for each test are presented to the right of each cohort ln(OR). EUR = European admixture, EAS = East Asian admixture, AFR = African admixture, LAT = Hispanic/Latino admixture, SA = suicide attempt, META = meta-analysis, PGC = Psychiatric Genomics Consortium, MDD = major depressive disorder, BIP = Bipolar disorder, SCZ = schizophrenia, ED = eating disorder.

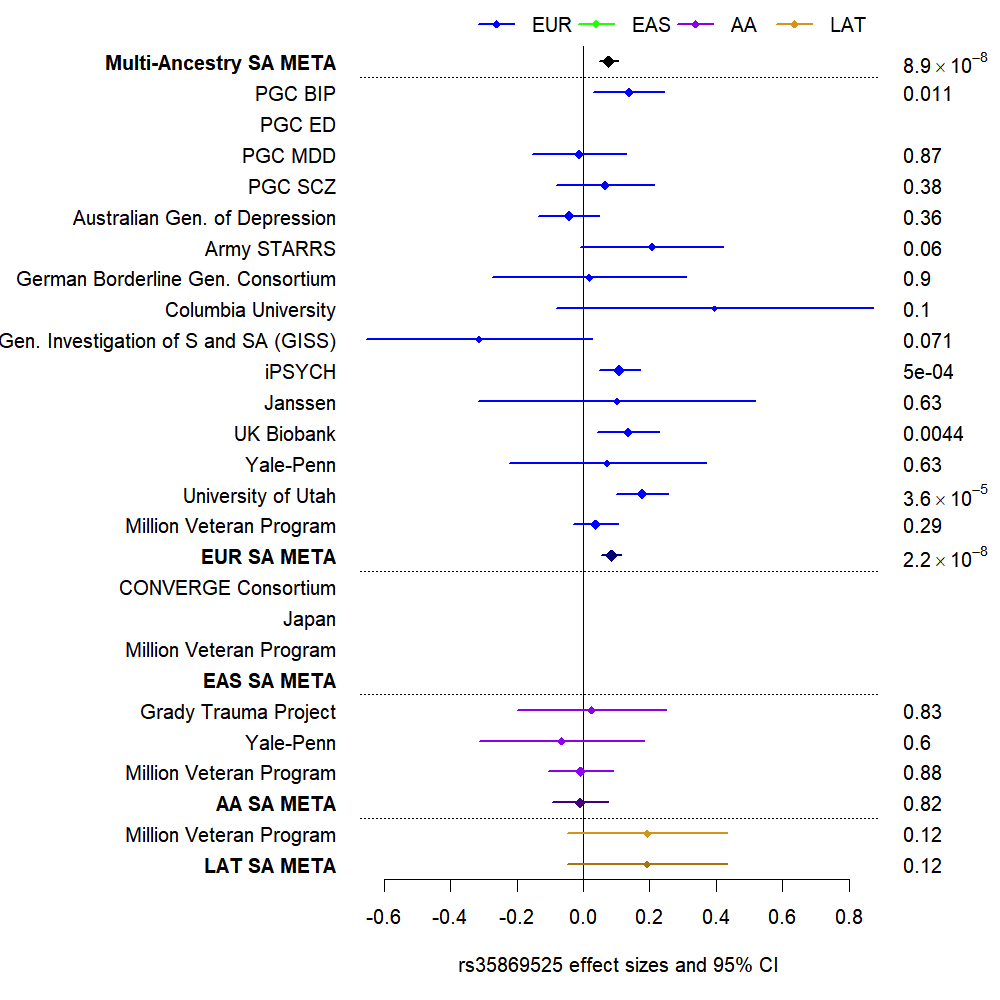

**Figure S13: Multi-Ancestry GWAS Meta-analysis Forest Plot of rs35869525 on Chromosome 6.**

Effect sizes are ln(OR), the log of the odds ratio of the effect allele on suicide attempt, CI = confidence interval

CHR- chromosome,BP - base pair position based on hg19, EUR - European admixture, EAS - East Asian admixture, AFR - admixed African American admixture, PGC - Psychiatric Genomics Consortium, MDD - major depressive disorder, BIP- Bipolar disorder, SCZ - schizophrenia, ED - eating disorder, ln(OR) - log of the odds ratio of the effect allele on suicide attempt, CI - confidence interval

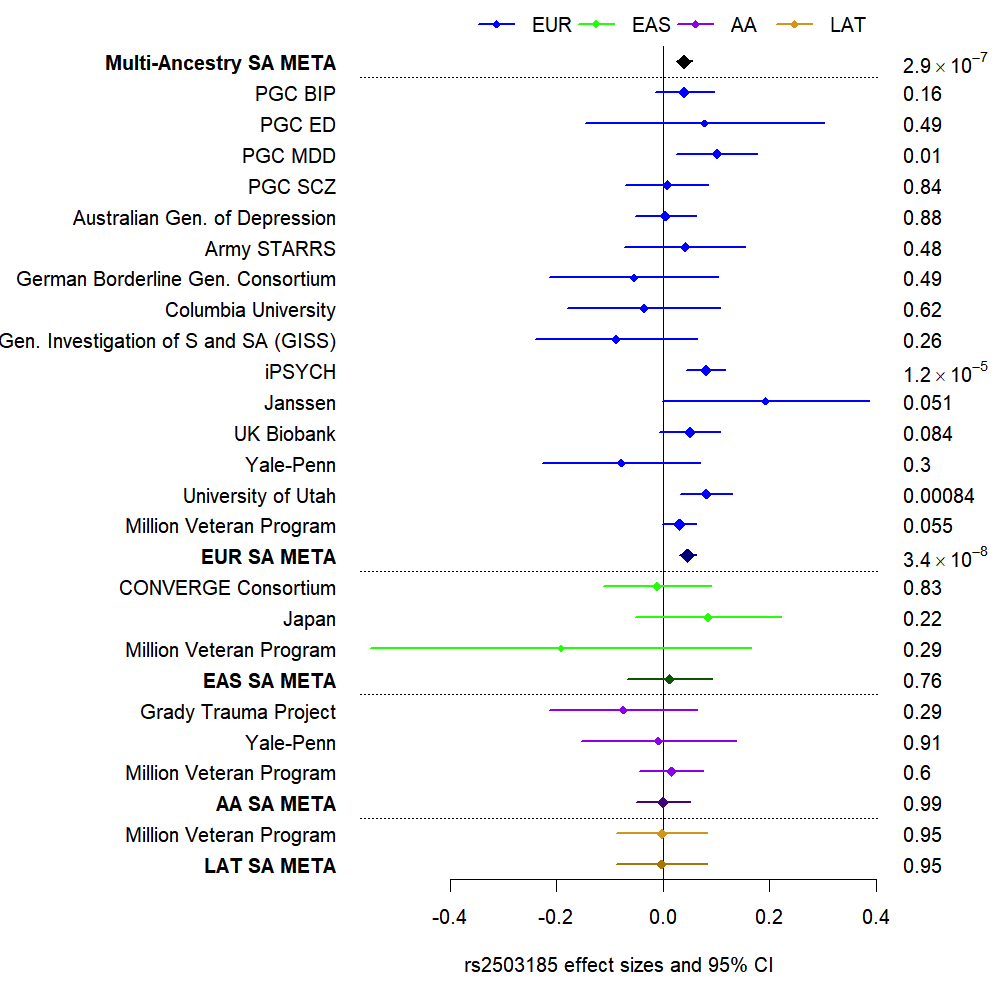

**Figure S14: Multi-Ancestry GWAS Meta-analysis Forest Plot of rs2503185 on Chromosome 1.**

Effect sizes are presented on the x-axis as the log of the odds ratio, ln(OR), of the effect allele on suicide attempt. CI = confidence interval. Cohorts and respective admixture-specific groupings are depicted on the y-axis. The p-values for each test are presented to the right of each cohort ln(OR). EUR = European admixture, EAS = East Asian admixture, AFR = African admixture, LAT = Hispanic/Latino admixture, SA = suicide attempt, META = meta-analysis, PGC = Psychiatric Genomics Consortium, MDD = major depressive disorder, BIP = Bipolar disorder, SCZ = schizophrenia, ED = eating disorder.

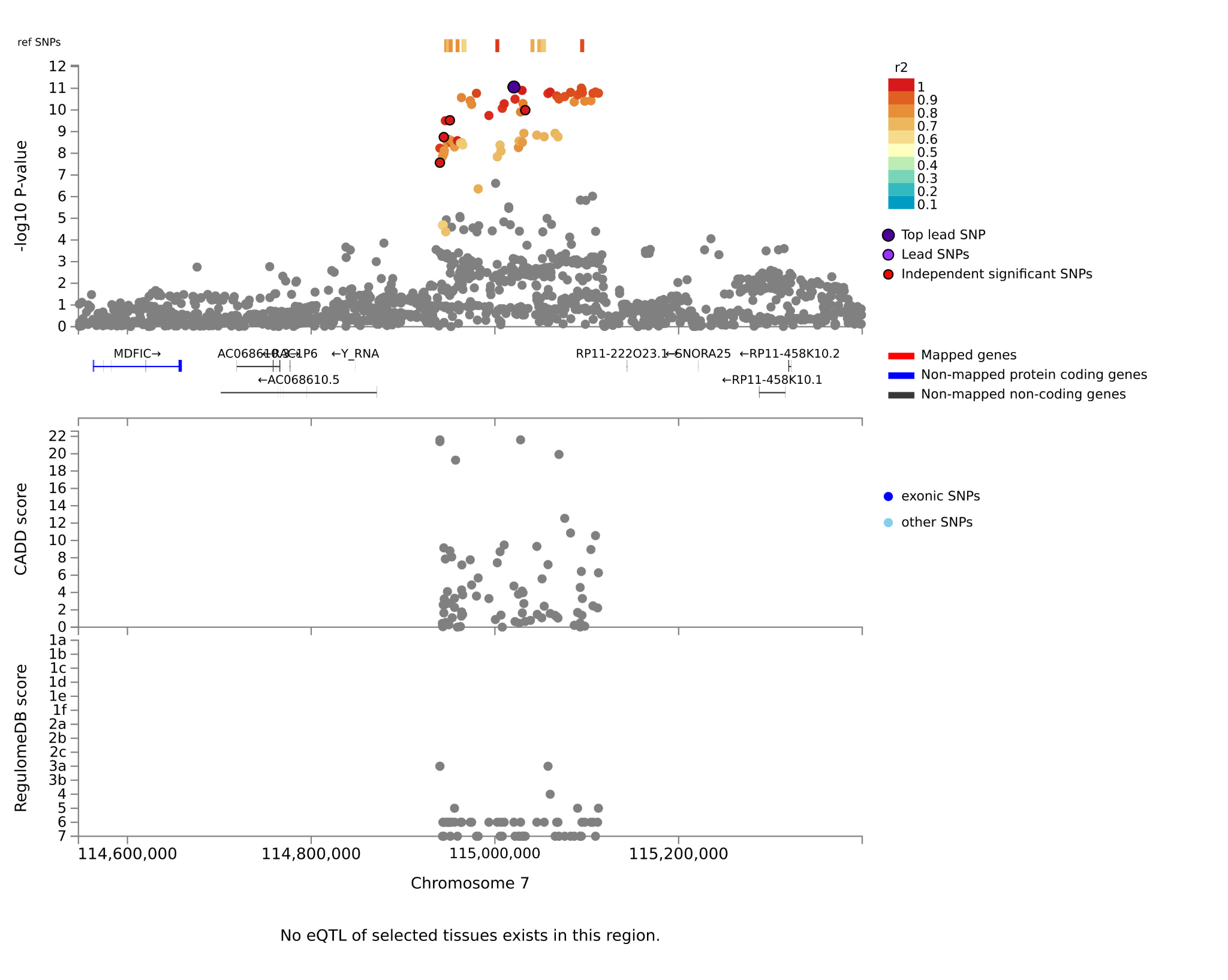

**Figure S15. Multi-Ancestry Regional Plot of Genome-Wide Significant Loci on Chromosome 7.** Plots are zoomed in to highlight the genomic region (x-axis) that contains the index SNP and SNPs in linkage disequilibrium with the index SNP. -log10 *p*-values are presented on the y-axis. Purple circles reflect the variant with the smallest *p*-value in the locus.

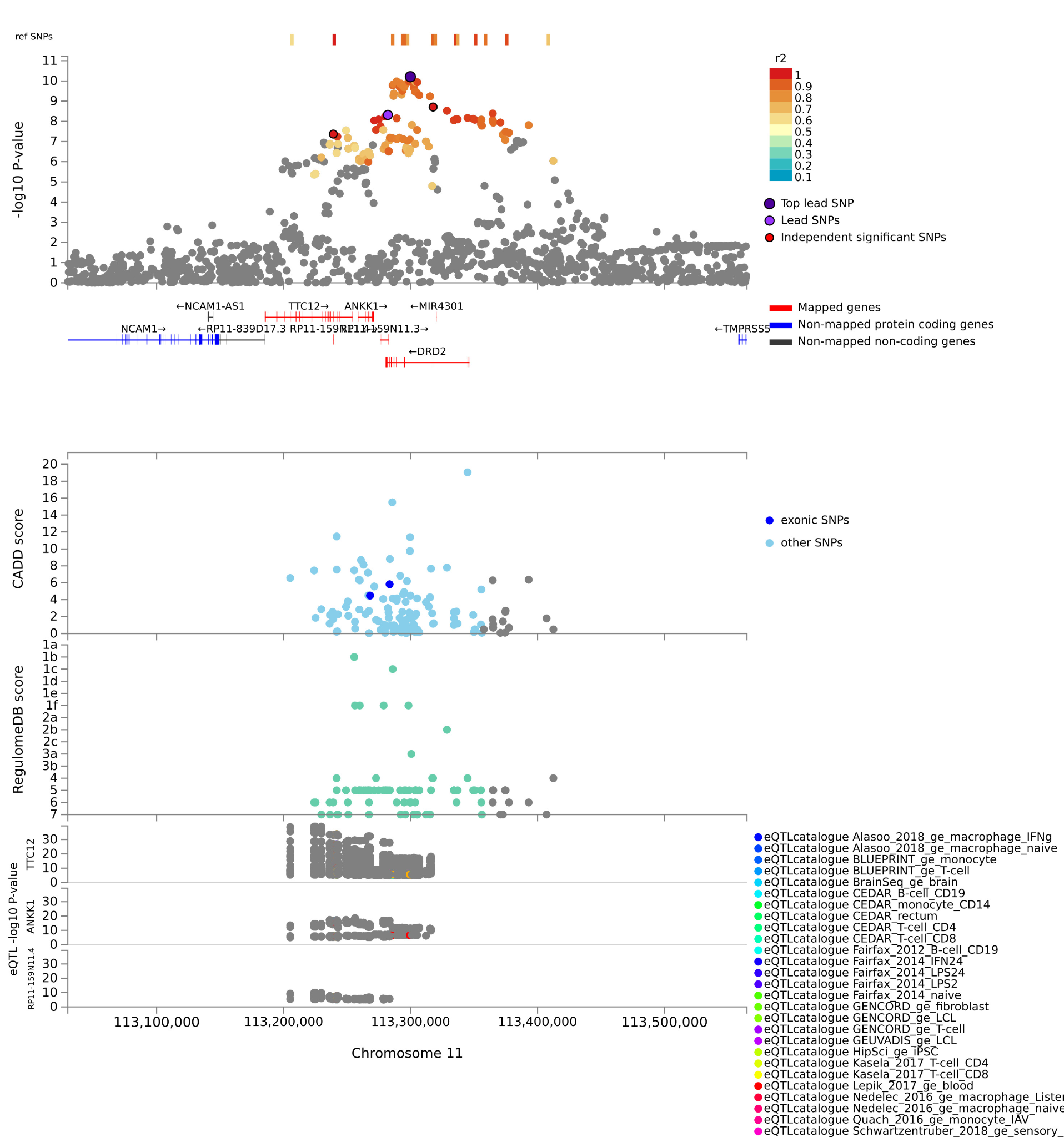

**Figure S16. Multi-Ancestry Regional Plot of Genome-Wide Significant Loci on Chromosome 11.** Plots are zoomed in to highlight the genomic region (x-axis) that contains the index SNP and SNPs in linkage disequilibrium with the index SNP. -log10 *p*-values are presented on the y-axis. Purple circles reflect the variant with the smallest *p*-value in the locus.

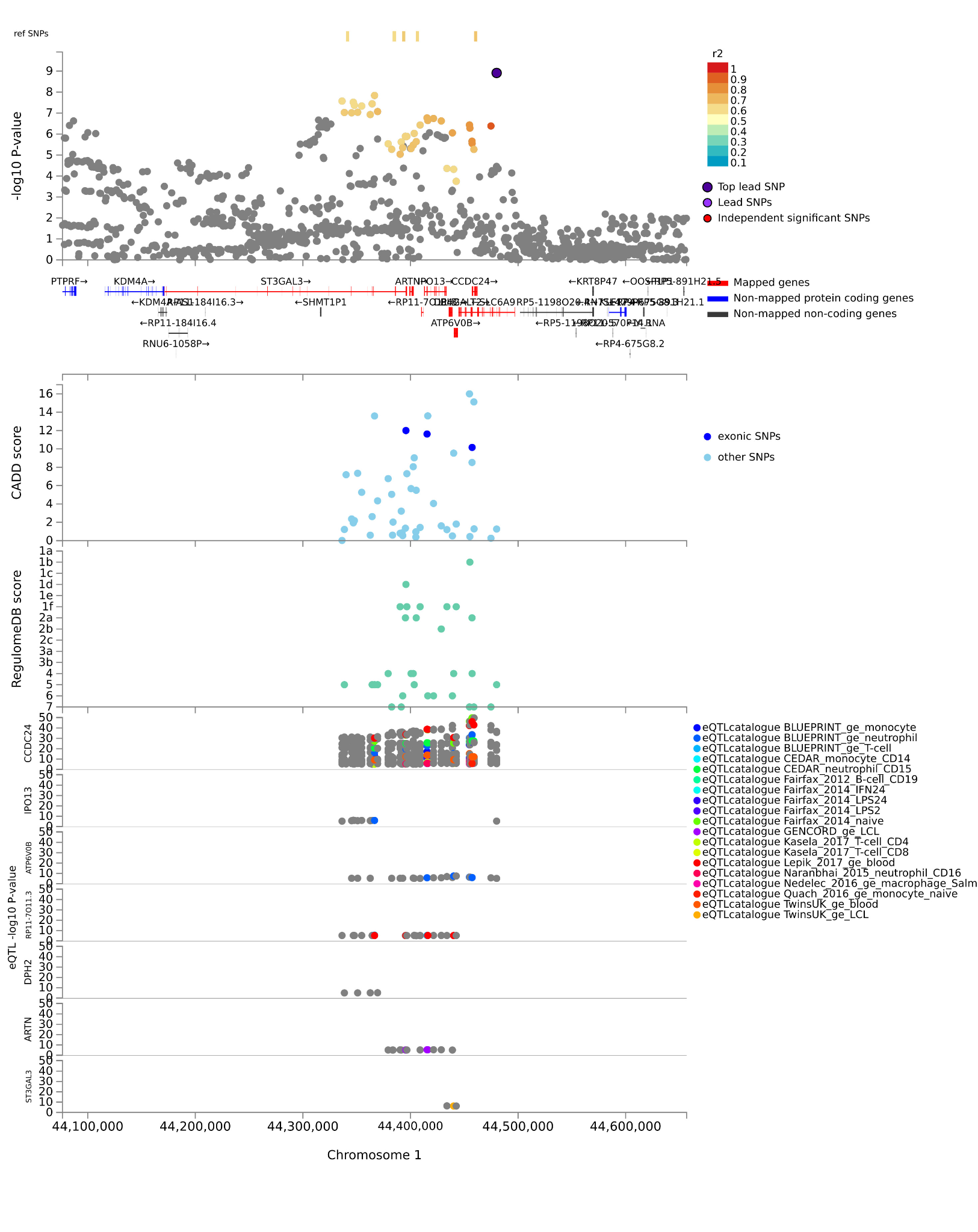

**Figure S17. Multi-Ancestry Regional Plot of Genome-Wide Significant Loci on Chromosome 1.** Plots are zoomed in to highlight the genomic region (x-axis) that contains the index SNP and SNPs in linkage disequilibrium with the index SNP. -log10 *p*-values are presented on the y-axis. Purple circles reflect the variant with the smallest *p*-value in the locus.

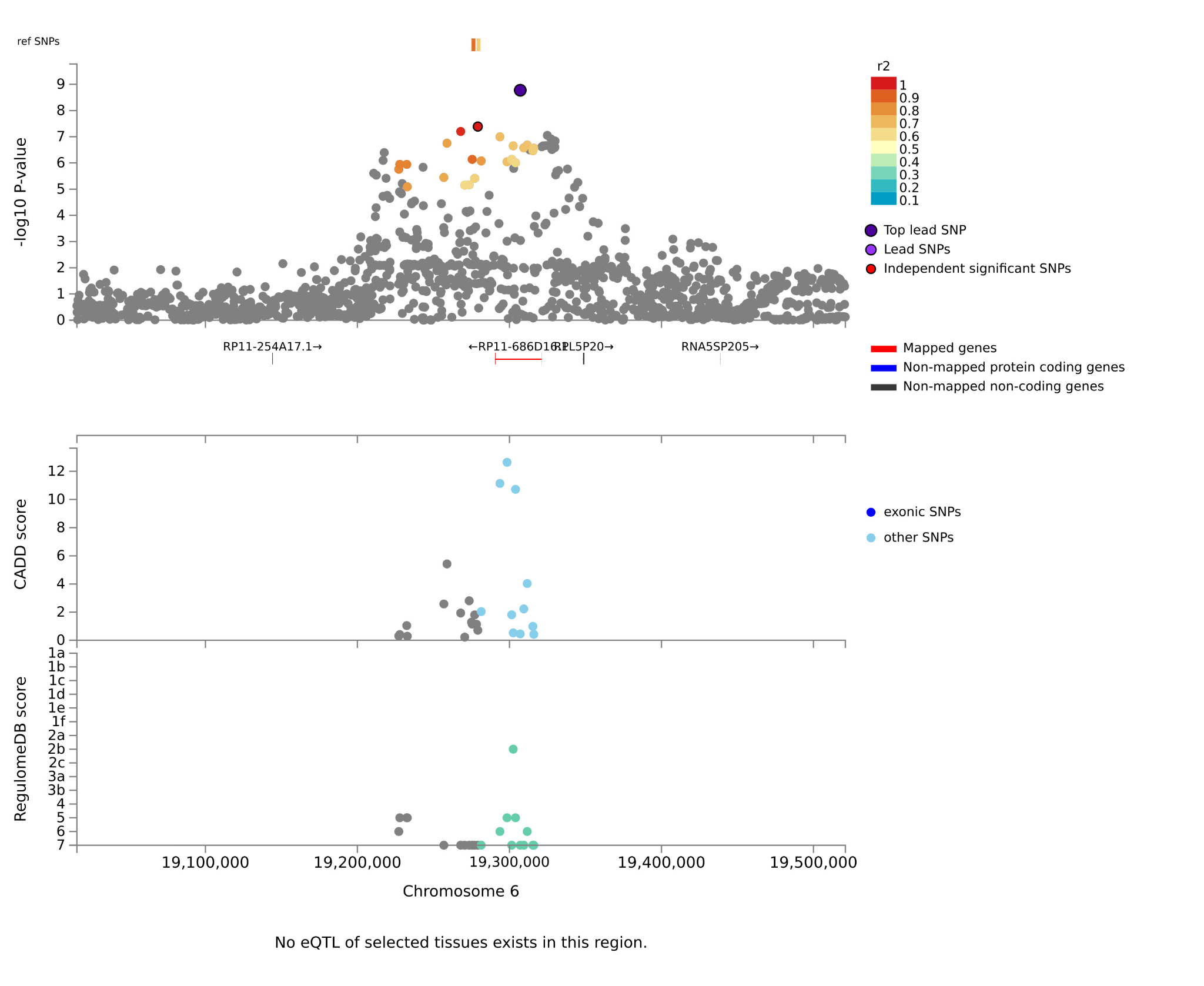

**Figure S18. Multi-Ancestry Regional Plot of Genome-Wide Significant Loci on Chromosome 6.** Plots are zoomed in to highlight the genomic region (x-axis) that contains the index SNP and SNPs in linkage disequilibrium with the index SNP. -log10 *p*-values are presented on the y-axis. Purple circles reflect the variant with the smallest *p*-value in the locus.

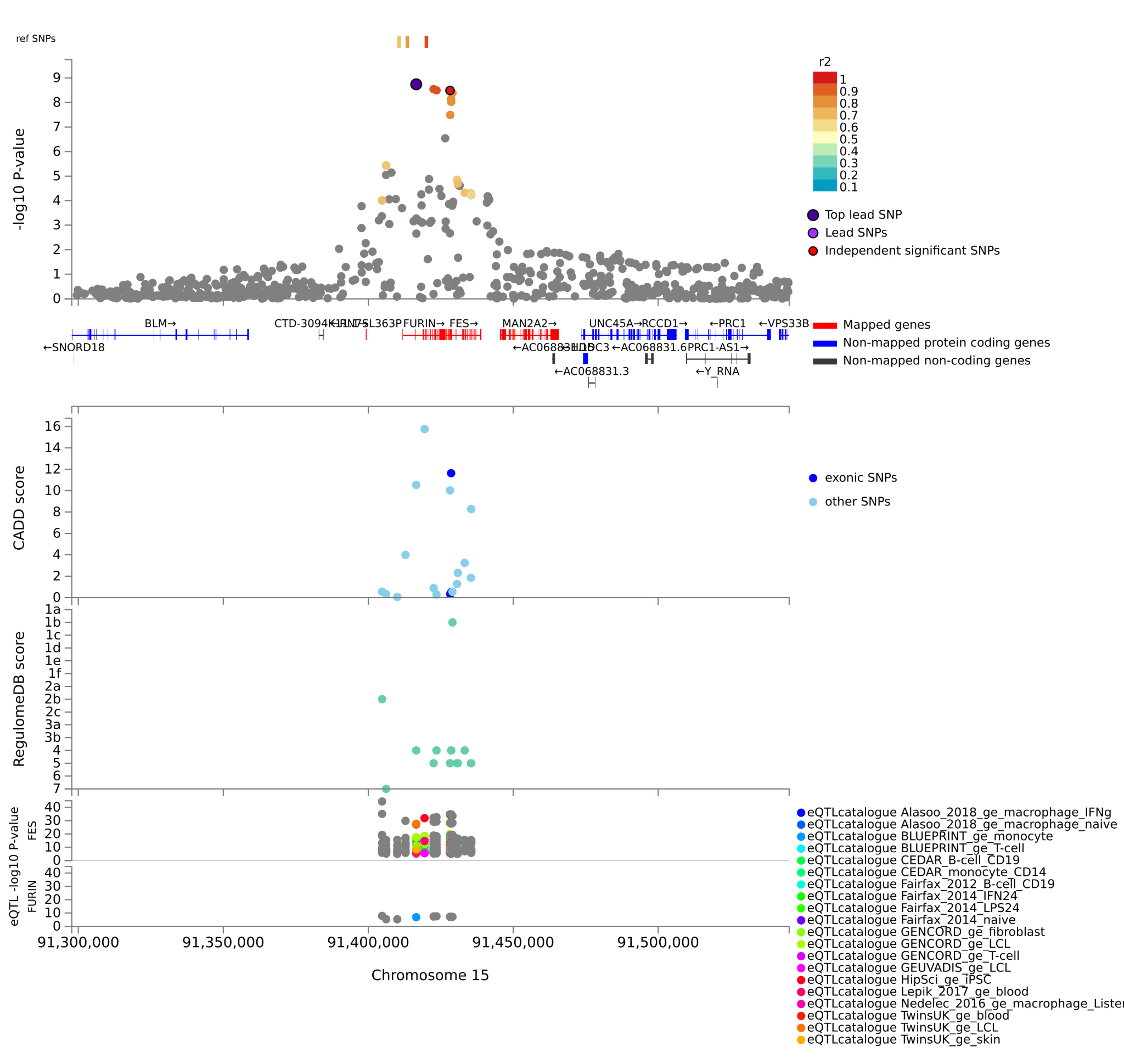

**Figure S19. Multi-Ancestry Regional Plot of Genome-Wide Significant Loci on Chromosome 15.** Plots are zoomed in to highlight the genomic region (x-axis) that contains the index SNP and SNPs in linkage disequilibrium with the index SNP. -log10 *p*-values are presented on the y-axis. Purple circles reflect the variant with the smallest *p*-value in the locus.

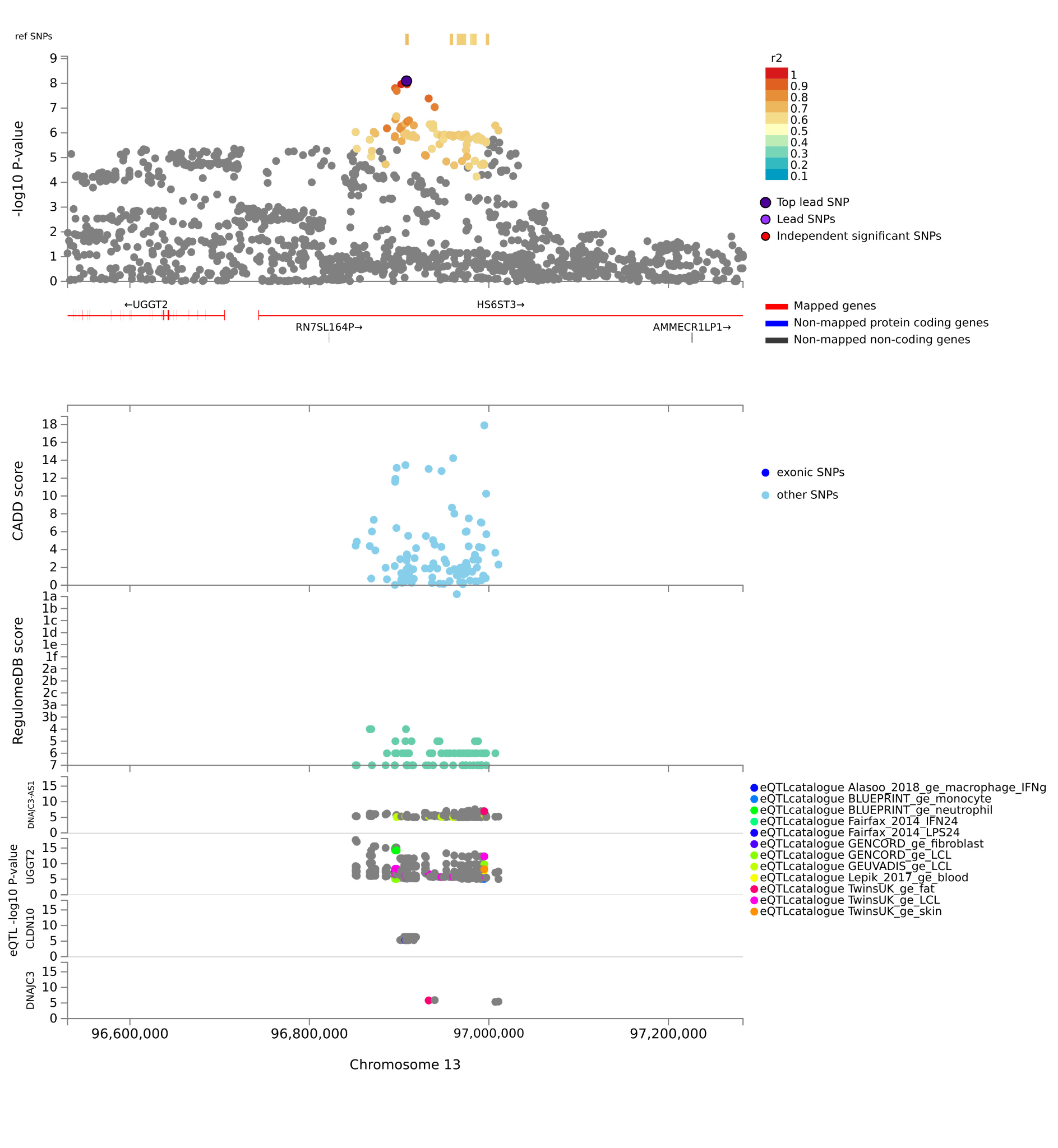

**Figure S20. Multi-Ancestry Regional Plot of Genome-Wide Significant Loci on Chromosome 13.** Plots are zoomed in to highlight the genomic region (x-axis) that contains the index SNP and SNPs in linkage disequilibrium with the index SNP. -log10 *p*-values are presented on the y-axis. Purple circles reflect the variant with the smallest *p*-value in the locus.

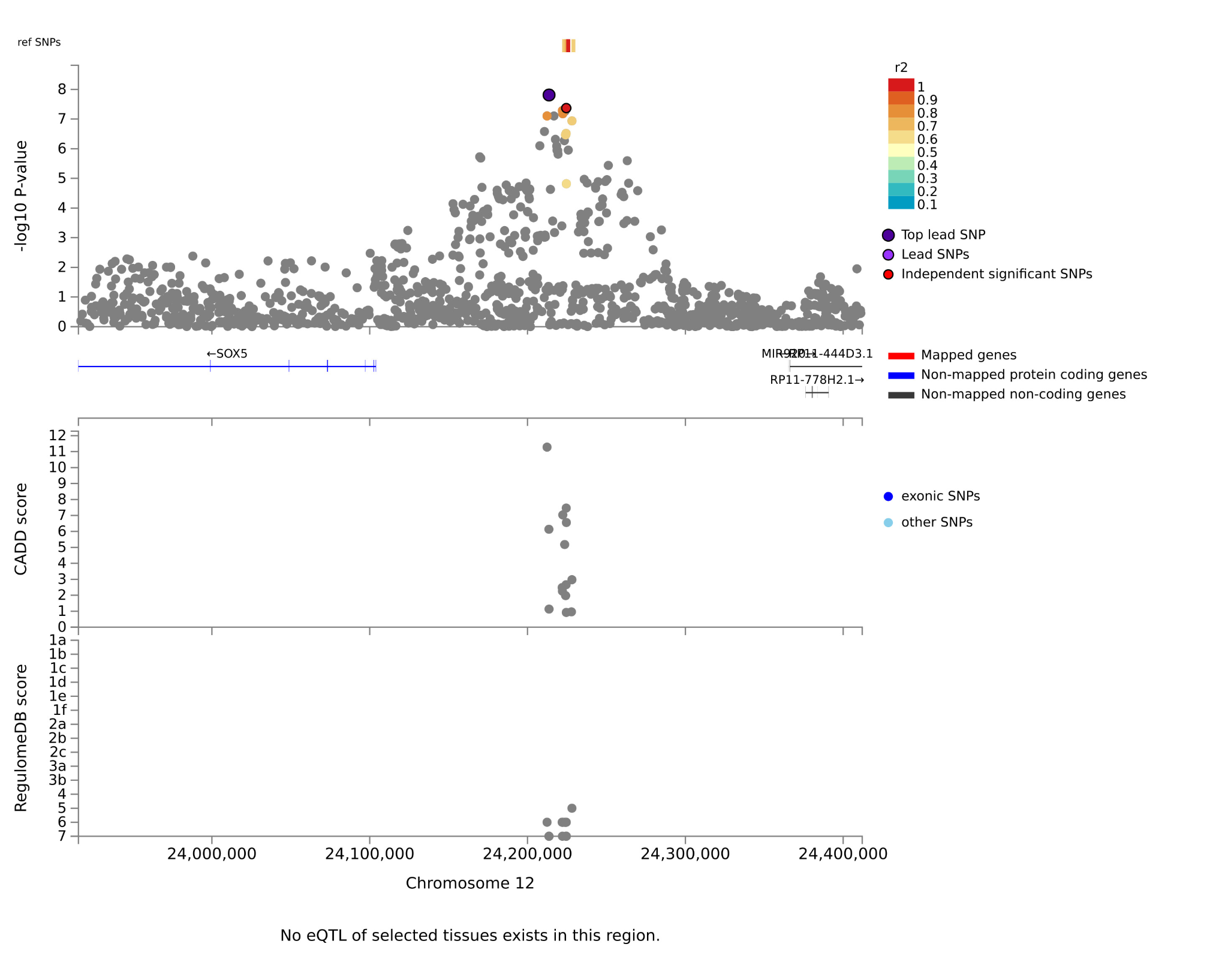

**Figure S21. Multi-Ancestry Regional Plot of Genome-Wide Significant Loci on Chromosome 12.** Plots are zoomed in to highlight the genomic region (x-axis) that contains the index SNP and SNPs in linkage disequilibrium with the index SNP. -log10 *p*-values are presented on the y-axis. Purple circles reflect the variant with the smallest *p*-value in the locus.

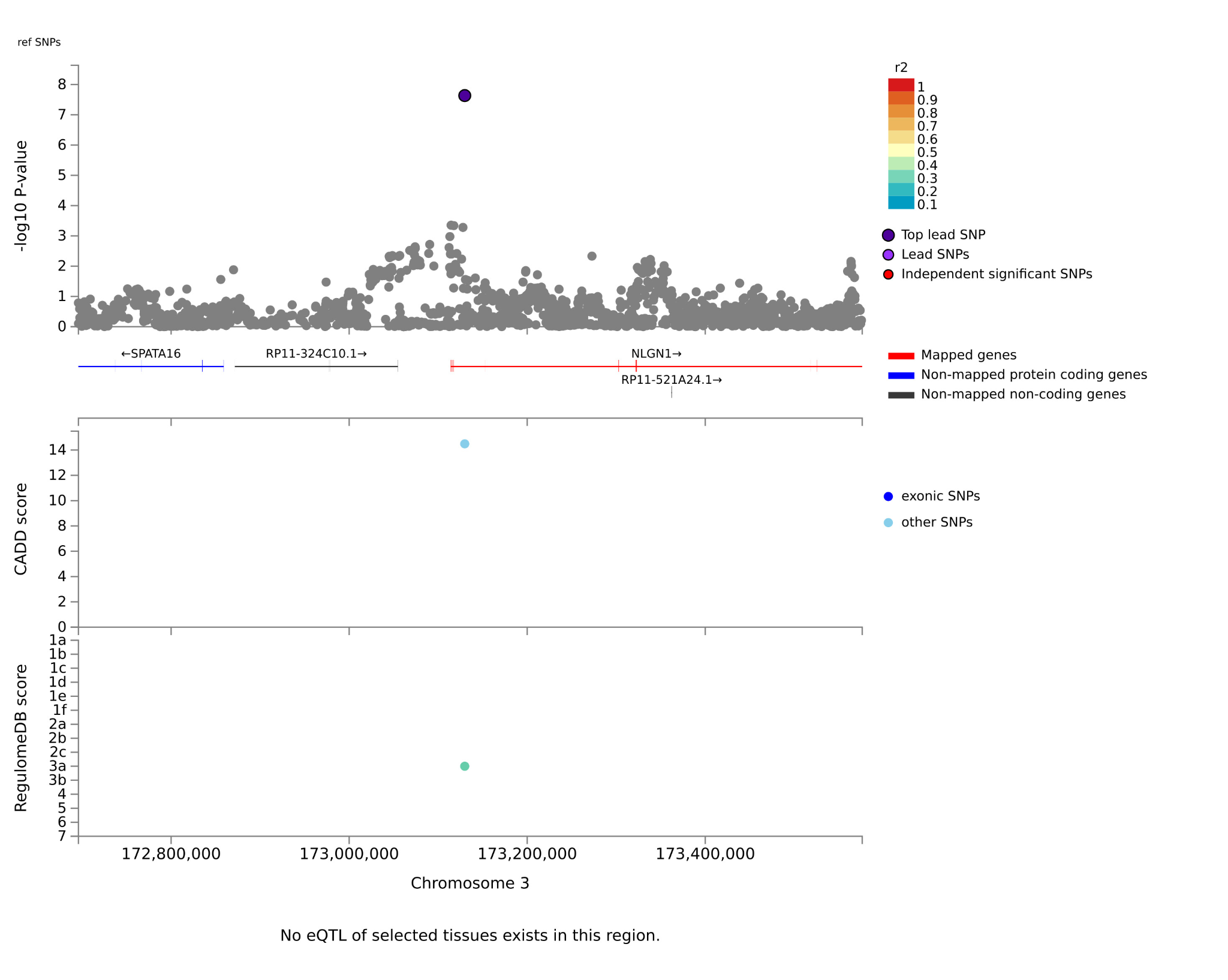

**Figure S22. Multi-Ancestry Regional Plot of Genome-Wide Significant Loci on Chromosome 3.** Plots are zoomed in to highlight the genomic region (x-axis) that contains the index SNP and SNPs in linkage disequilibrium with the index SNP. -log10 *p*-values are presented on the y-axis. Purple circles reflect the variant with the smallest *p*-value in the locus.

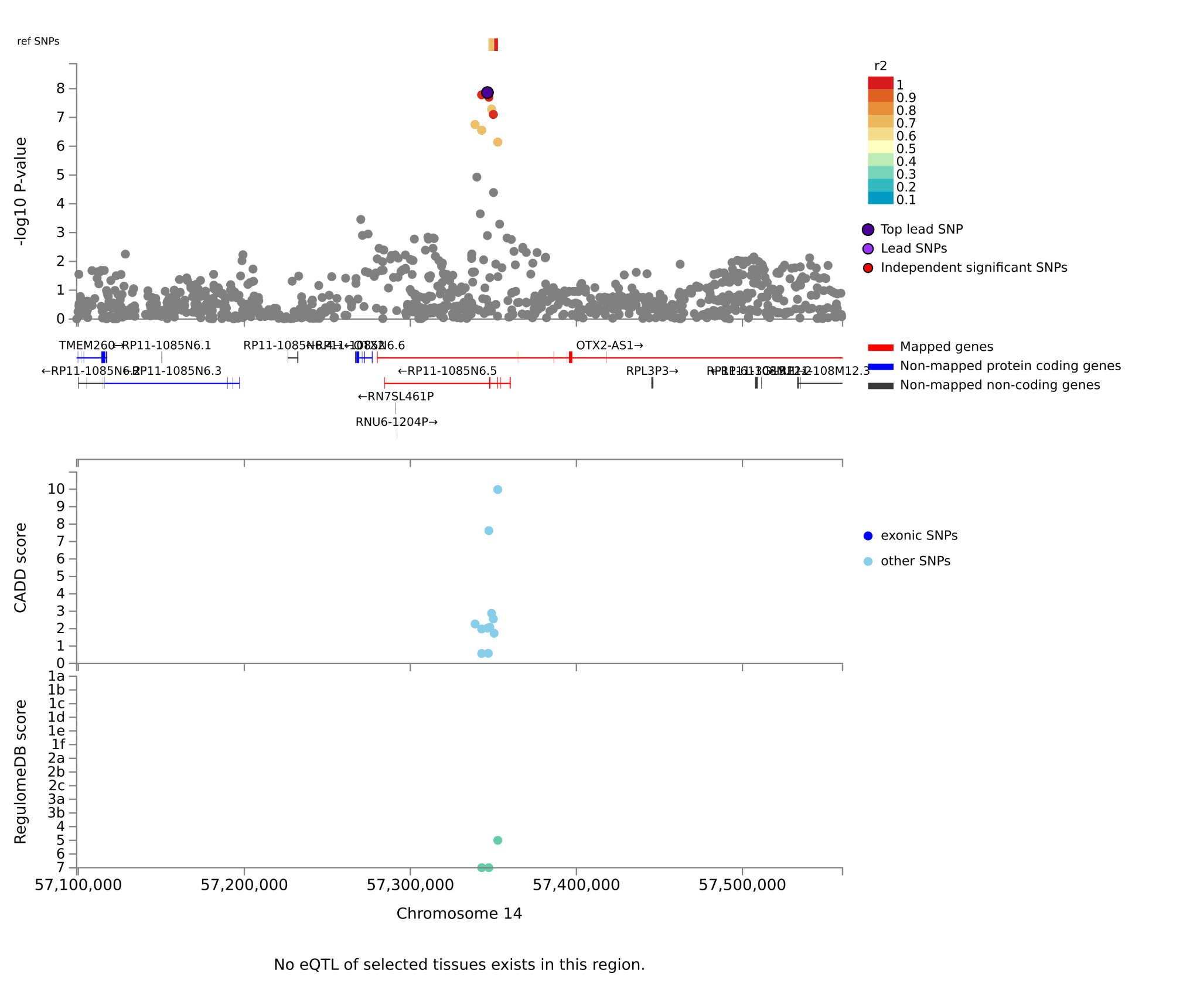

**Figure S23. EUR Regional Plot of Genome-Wide Significant Loci on Chromosome 14.** Plots are zoomed in to highlight the genomic region (x-axis) that contains the index SNP and SNPs in linkage disequilibrium with the index SNP. -log10 *p*-values are presented on the y-axis. Purple circles reflect the variant with the smallest *p*-value in the locus.

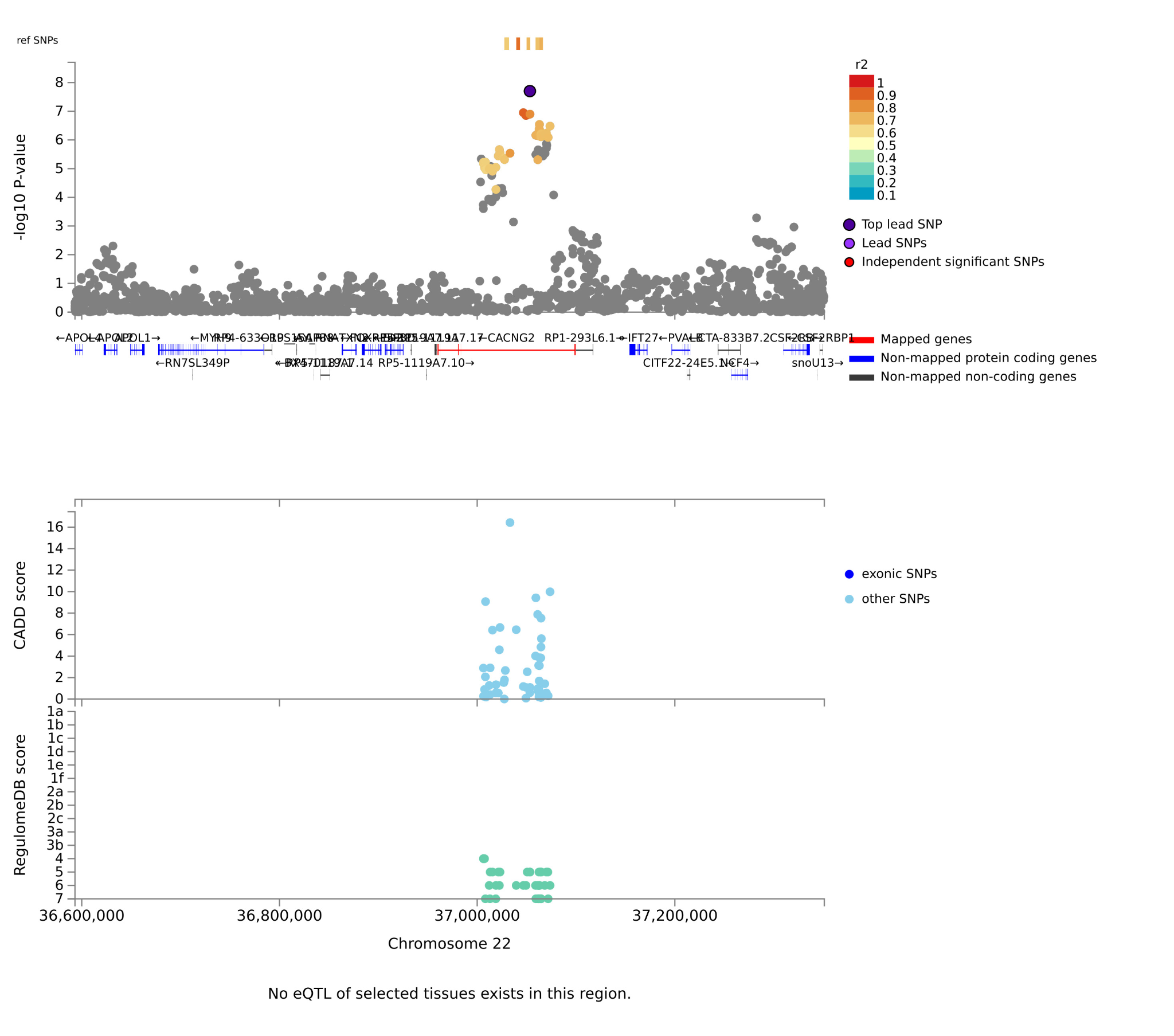

**Figure S24. EUR Regional Plot of Genome-Wide Significant Loci on Chromosome 22.** Plots are zoomed in to highlight the genomic region (x-axis) that contains the index SNP and SNPs in linkage disequilibrium with the index SNP. -log10 *p*-values are presented on the y-axis. Purple circles reflect the variant with the smallest *p*-value in the locus.

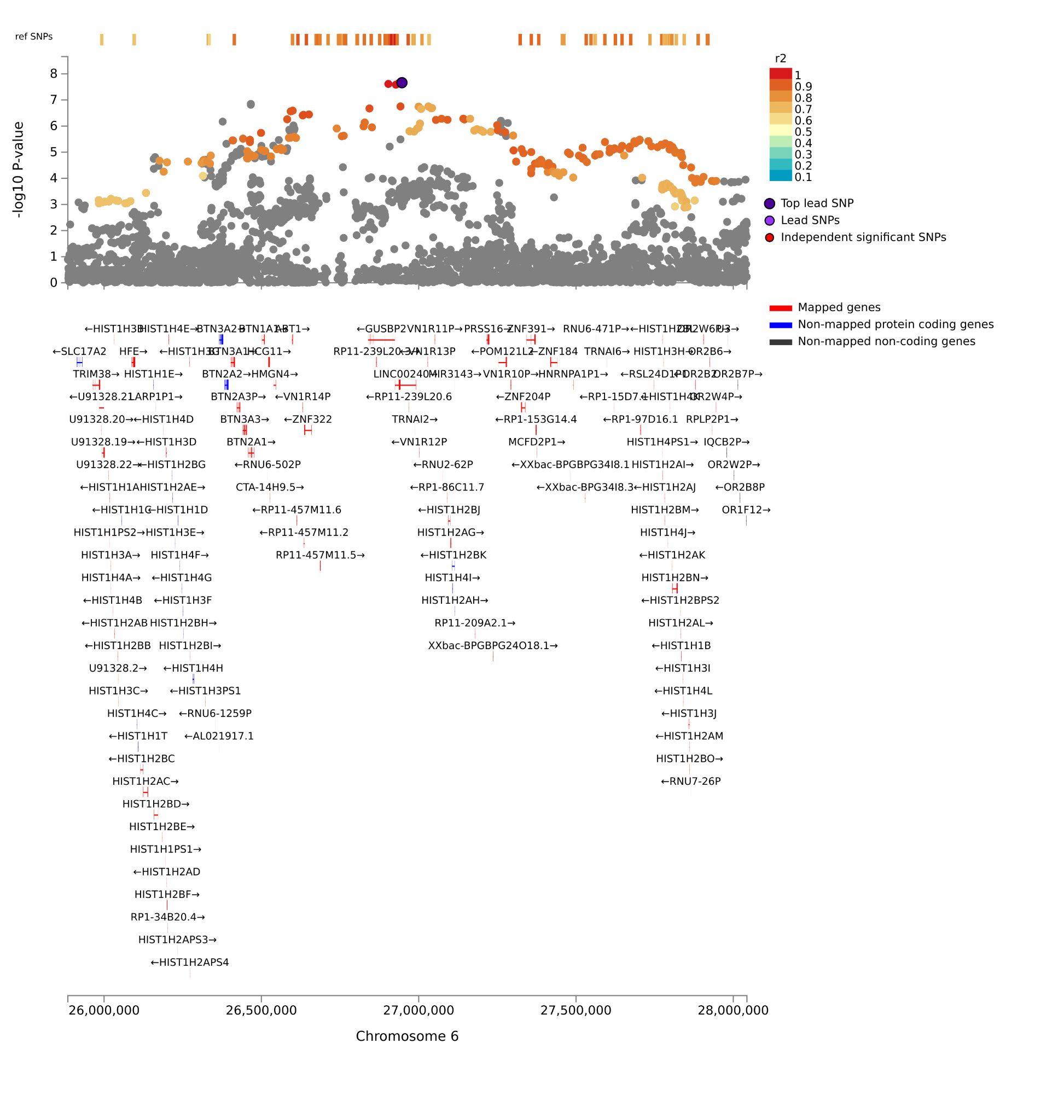

**Figure S25. EUR Regional Plot of Genome-Wide Significant Loci on Chromosome 6.** Plots are zoomed in to highlight the genomic region (x-axis) that contains the index SNP and SNPs in linkage disequilibrium with the index SNP. -log10 *p*-values are presented on the y-axis. Purple circles reflect the variant with the smallest *p*-value in the locus.

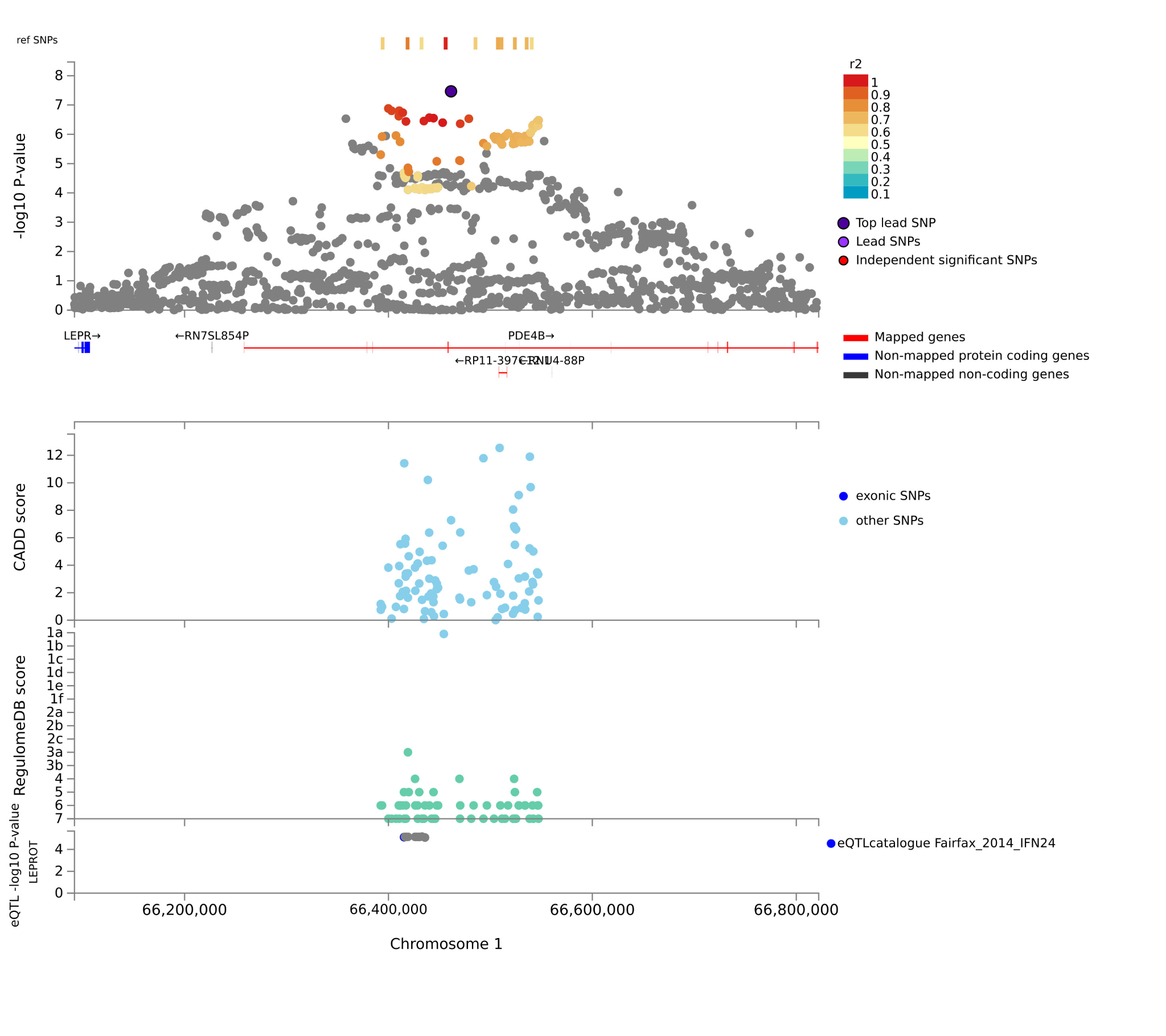

**Figure S26. EUR Regional Plot of Genome-Wide Significant Loci on Chromosome 1.** Plots are zoomed in to highlight the genomic region (x-axis) that contains the index SNP and SNPs in linkage disequilibrium with the index SNP. -log10 *p*-values are presented on the y-axis. Purple circles reflect the variant with the smallest *p*-value in the locus.

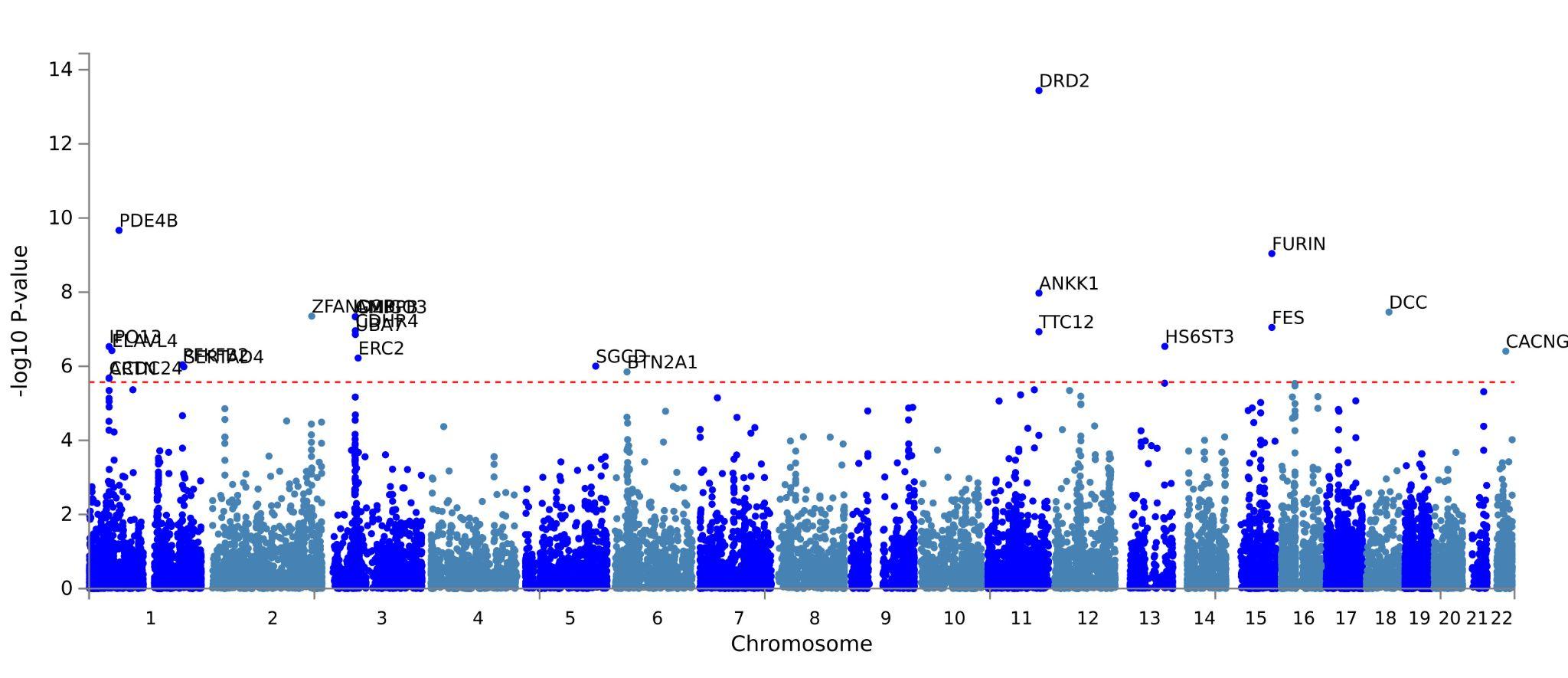

**Figure S27: Multi-Ancestry Gene-Based Manhattan Plot.** Gene analysis was performed by using MAGMA (v1.6)^1^ with its default setting in FUMA^2^. SNPs were assigned to the genes obtained from Ensembl build 85 (only protein-coding genes). Genome-wide significance (red dashed line) was set at 0.05 / (the number of tested genes = 18627) = 2.684x10^-6^. Genes whose *p*-value reached the genome-wide significance are labeled in the Manhattan plot.

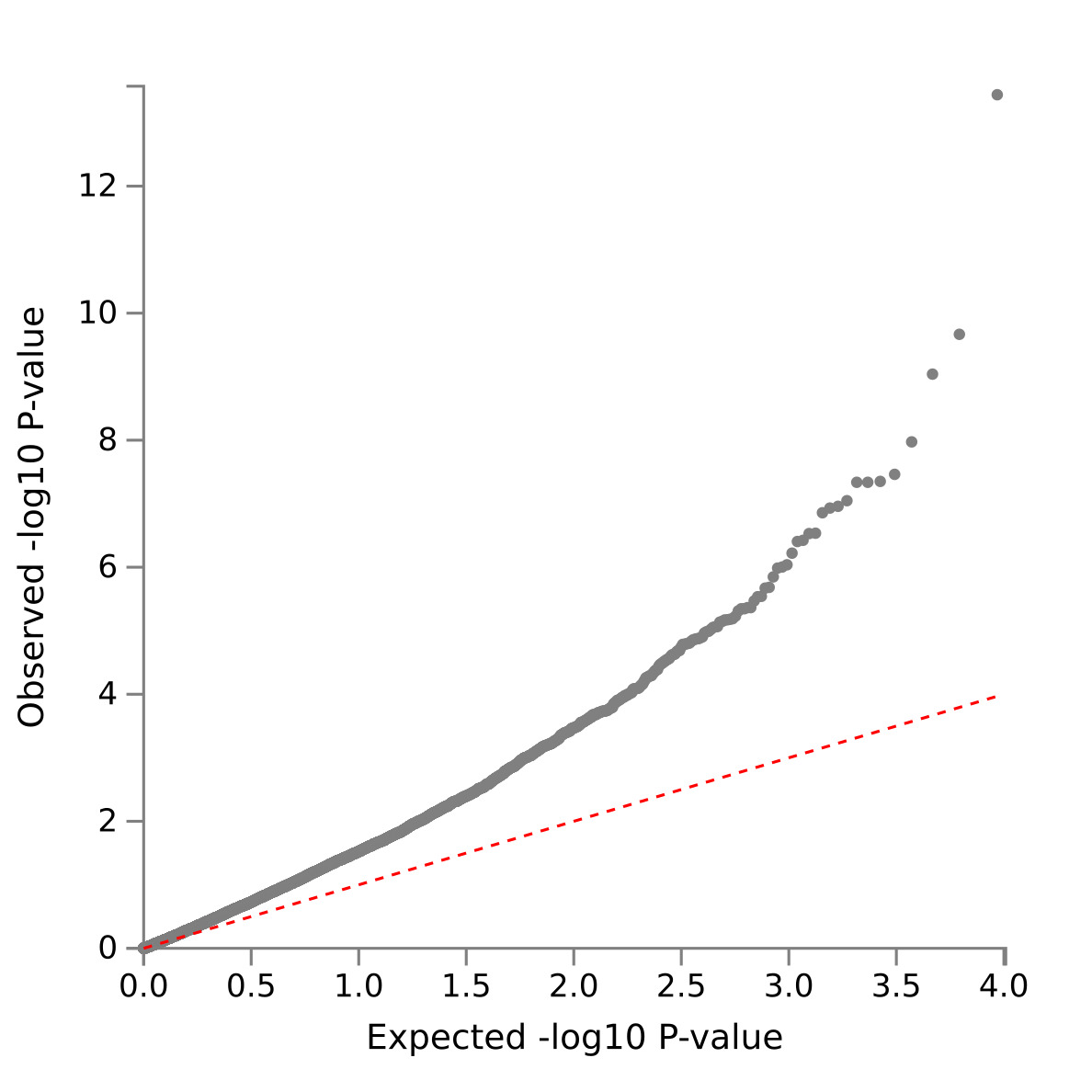

**Figure S28. Multi-Ancestry Gene-Based QQ-Plot of Suicide Attempt.** The y-axis reflects observed -log10 *p*-values. The x-axis is the number of significant *p*-values expected under the null hypothesis.

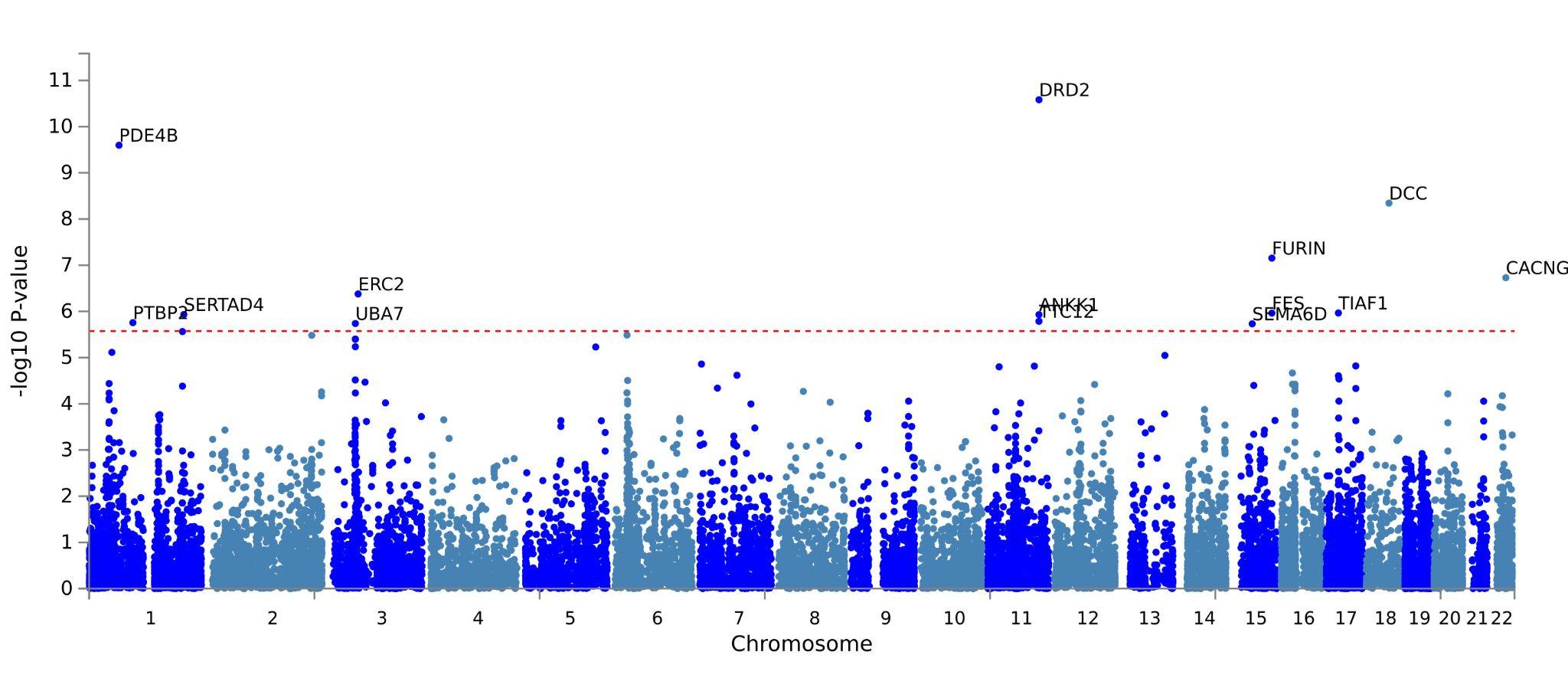

**Figure S29: EUR Gene-Based Manhattan Plot.** Gene analysis was performed by using MAGMA (v1.6)^1^ with its default setting in FUMA^2^. SNPs were assigned to the genes obtained from Ensembl build 85 (only protein-coding genes). Genome-wide significance (red dashed line) was set at 0.05 / (the number of tested genes = 18627) = 2.684x10^-6^. Genes whose *p*-value reached the genome-wide significance are labeled in the Manhattan plot.

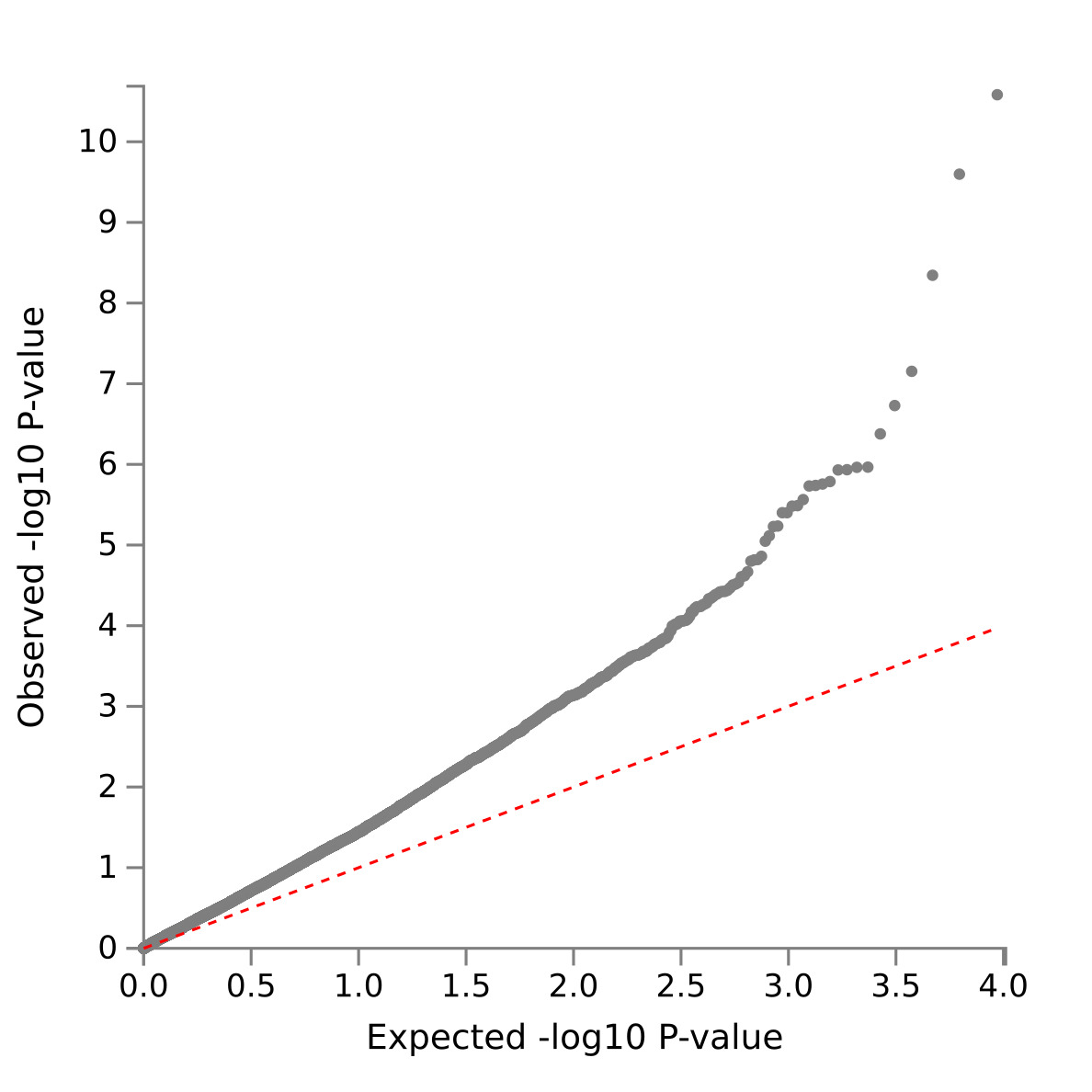

**Figure S30: EUR Gene-Based QQ-Plot of Suicide Attempt.** The y-axis reflects observed -log10 *p*-values. The x-axis is the number of significant *p*-values expected under null hypothesis.

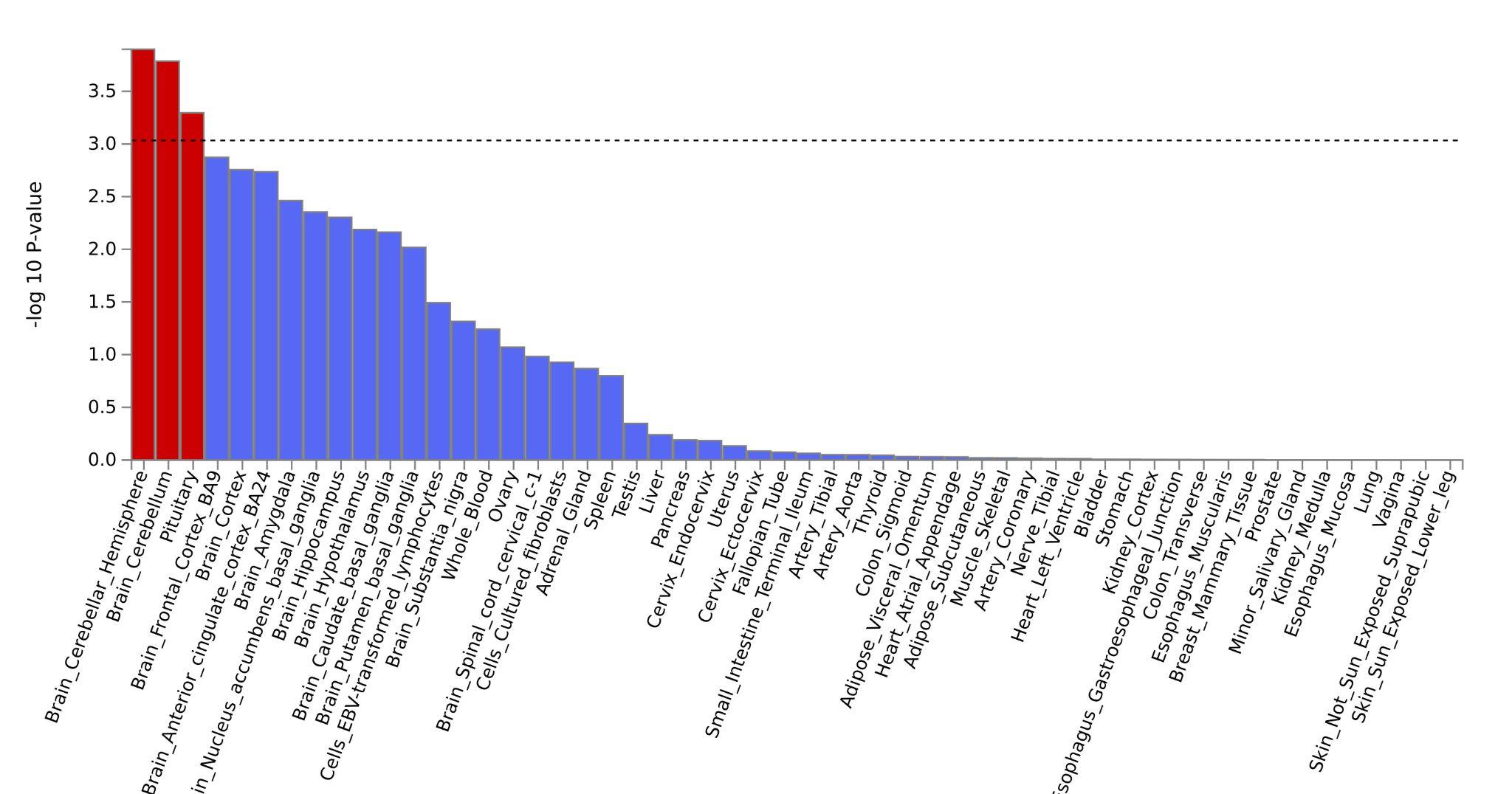

**Figure S31: Multi-Ancestry MAGMA Tissue Expression Analysis.** To test the (positive) relationship between highly expressed genes in a specific tissue and genetic associations, gene-property analysis is performed using average expression of genes per tissue type as a gene covariate. Gene expression values are log2 transformed average Reads Per Kilobase Million (RPKM) per tissue type, after winsorized at 50 based on GTEx RNA-seq data. Tissue expression analysis is performed for 30 general tissue types and 53 specific tissue types separately, the 53 specific types presented in this figure. MAGMA was performed using the result of the gene analysis (gene-based *p*-values) and tested for one side (greater) with conditioning on average expression across all tissue types. Gene expression analysis summary statistics across general and specific tissue types for the multi-ancestry SA GWAS are provided in Table S9.

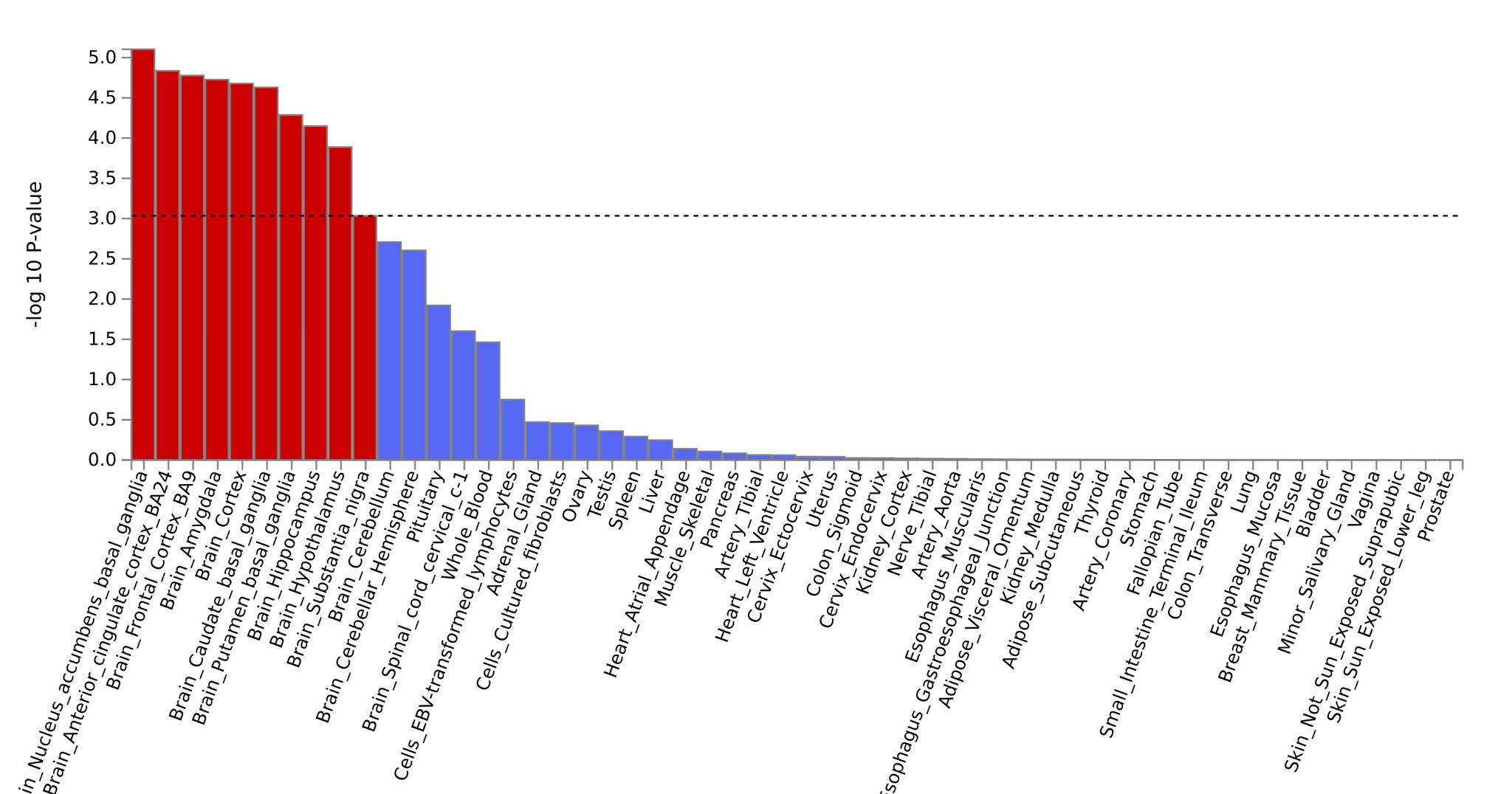

**Figure S32: EUR MAGMA Tissue Expression Analysis.** To test the (positive) relationship between highly expressed genes in a specific tissue and genetic associations, gene-property analysis is performed using average expression of genes per tissue type as a gene covariate. Gene expression values are log2 transformed average Reads Per Kilobase Million (RPKM) per tissue type, after winsorized at 50 based on GTEx RNA-seq data. Tissue expression analysis is performed for 30 general tissue types and 53 specific tissue types separately, the 53 specific types presented in this figure. MAGMA was performed using the result of the gene analysis (gene-based *p*-values) and tested for one side (greater) with conditioning on average expression across all tissue types. Gene expression analysis summary statistics across general and specific tissue types for the EUR SA GWAS are provided in Table S10.

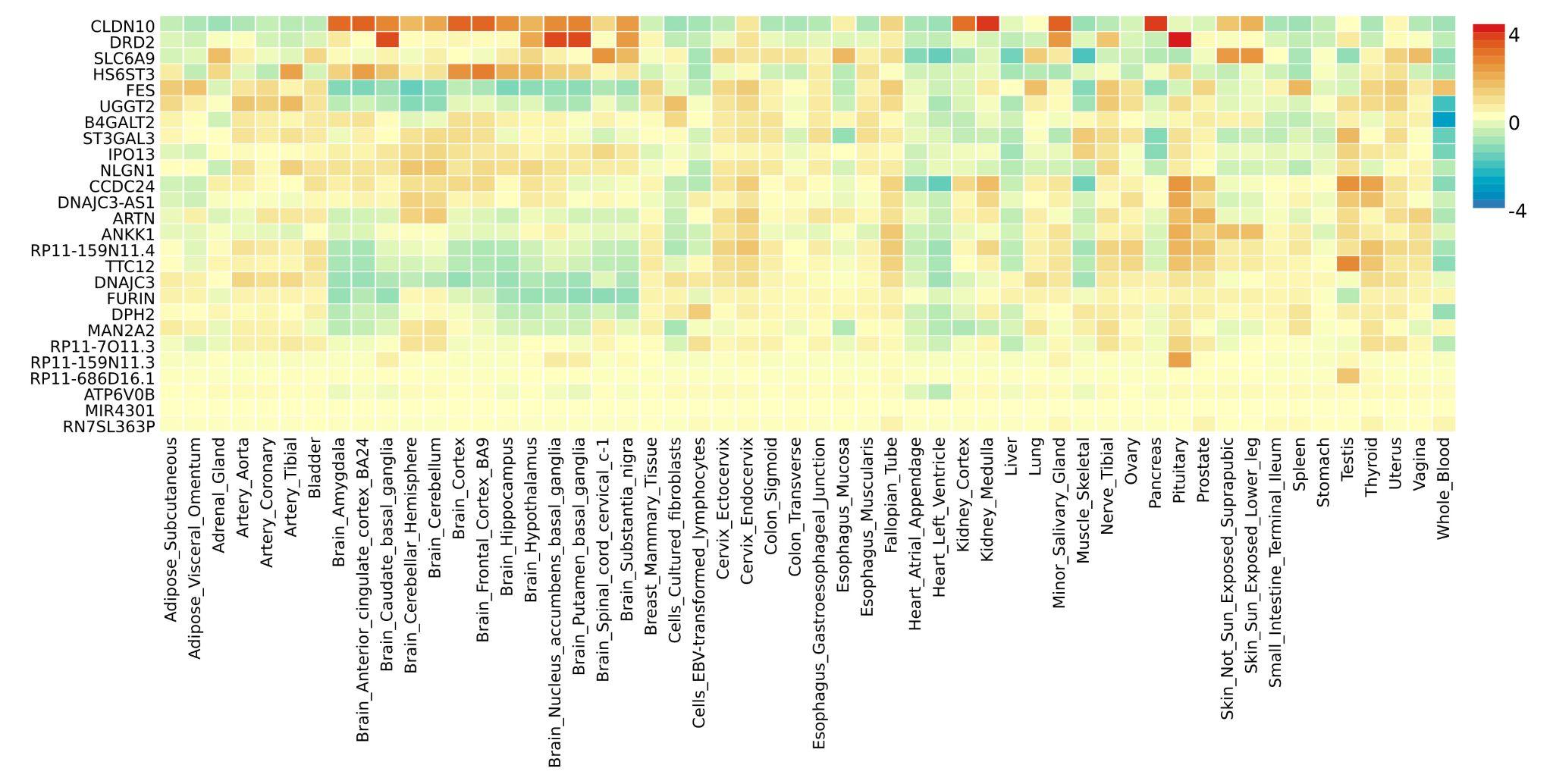

**Figure S33: Multi-Ancestry GTex Gene Expression Heatmap.** The expression value depends on the data set, RPKM (Read Per Kilobase per Million) for GTEx v6 and BrainSpan, TPM (Transcripts Per Million) for GTEx v7. Cells filled in red represent higher expression compared to cells filled in blue across genes and labels. This heatmap depicts the average of normalized expression (zero mean across samples) following winsorization at 50 and log2 transformation of the expression value with pseudocount 1. Genes and labels are both ordered by hierarchical clustering. Hierarchical clustering is performed using the Python scipy package^3^ (using "average" method).

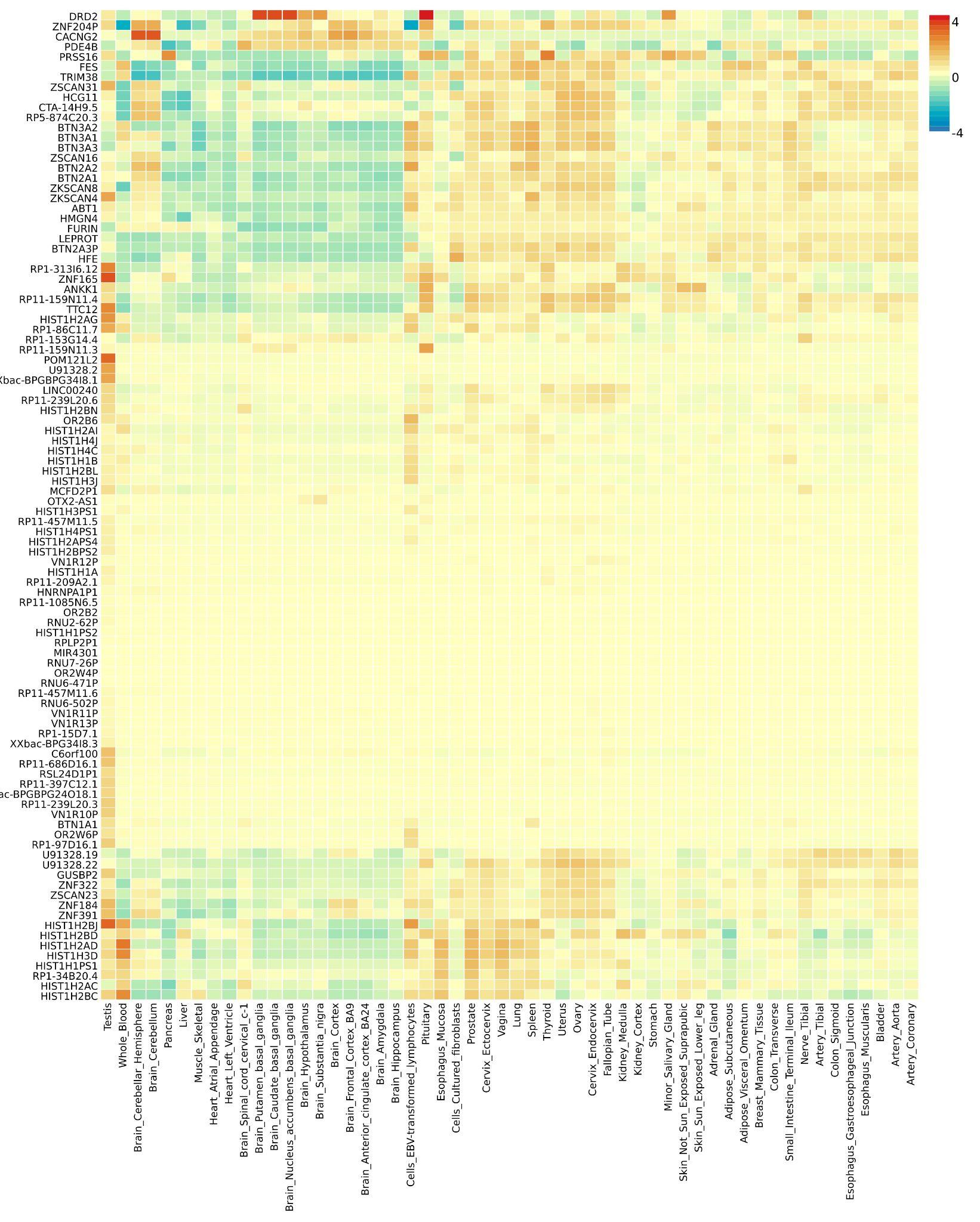

**Figure S34: EUR GTex Gene Expression Heatmap.** The expression value depends on the data set, RPKM (Read Per Kilobase per Million) for GTEx v6 and BrainSpan, TPM (Transcripts Per Million) for GTEx v7. Cells filled in red represent higher expression compared to cells filled in blue across genes and labels. This heatmap depicts the average of normalized expression (zero mean across samples) following winsorization at 50 and log2 transformation of the expression value with pseudocount 1. Genes and labels are both ordered by hierarchical clustering. Hierarchical clustering is performed using the Python scipy package^3^ (using "average" method).

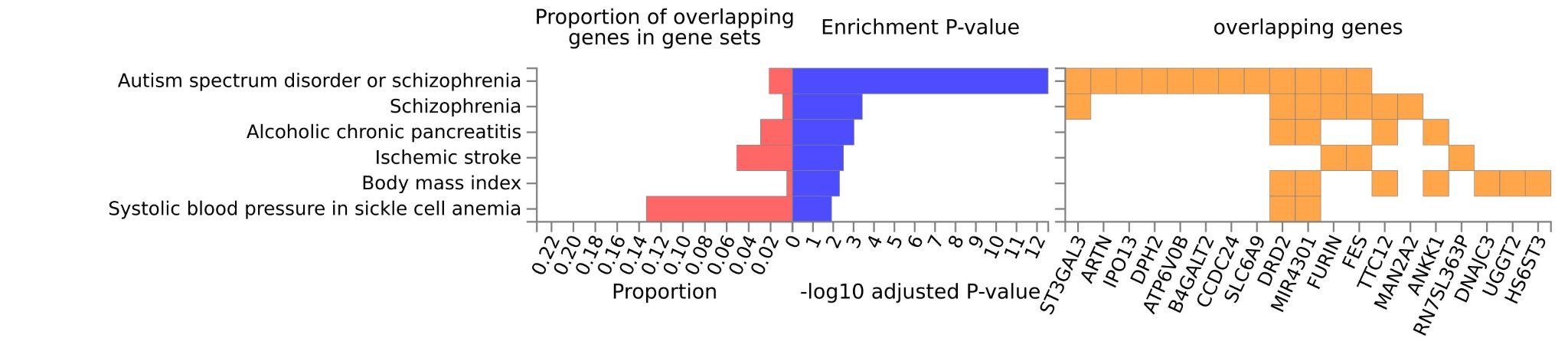

**Figure S35: Multi-Ancestry Enrichment of Genes in Gene Sets from GWAS Catalog.** Hypergeometric tests are performed to test if genes of interest are overrepresented in any of the pre-defined gene sets. Multiple test correction is performed per category, (i.e. canonical pathways, GO biological processes and so on, separately). Gene sets were obtained from MsigDB, WikiPathways and reported genes from the GWAS Catalog. GWAS Catalog results are shown here.

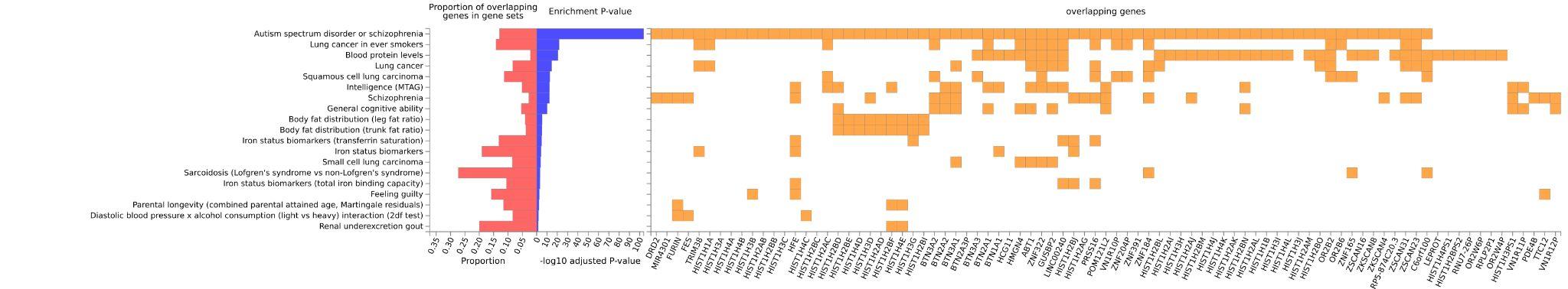

**Figure S36: EUR Enrichment of Genes in Gene Sets from GWAS Catalog.** Hypergeometric tests are performed to test if genes of interest are overrepresented in any of the pre-defined gene sets. Multiple test correction is performed per category, (i.e. canonical pathways, GO biological processes and so on, separately). Gene sets were obtained from MsigDB, WikiPathways and reported genes from the GWAS Catalog. GWAS Catalog results are shown here.

**Figure References**

1. de Leeuw, C. A., Mooij, J. M., Heskes, T. & Posthuma, D. MAGMA: generalized gene-set analysis of GWAS data. *PLoS Comput. Biol.* **11**, e1004219 (2015).
2. Watanabe, K., Taskesen, E., van Bochoven, A. & Posthuma, D. Functional mapping and annotation of genetic associations with FUMA. *Nat. Commun.* **8**, 1826 (2017).
3. Virtanen, P. *et al.* SciPy 1.0: fundamental algorithms for scientific computing in Python. *Nat. Methods* **17**, 261–272 (2020).
