## Supplementary material for "Genome-wide association study meta-analysis of suicide attempt identifies twelve genome-wide significant loci and implicates genetic risks for specific health factors": Supplementary_Materials_Docherty_ISGC_MVP_SA_GWAS_01012023.docx

**CONTENTS**

**ISGC COHORT ASCERTAINMENT, CASE AND CONTROL DEFINITIONS**	**1**

**ISGC COHORT GENOTYPING, QC, IMPUTATION AND ANALYSIS**	**13**

**ACKNOWLEDGEMENTS**	**18**

**CONSORTIUM AUTHORS**	**29**

### ISGC COHORT ASCERTAINMENT, CASE AND CONTROL DEFINITIONS

**Psychiatric Genomics Consortium Major Depressive Disorder**

Subjects were drawn from 14 major depressive disorder (MDD) case-control cohorts in the Psychiatric Genomics Consortium (PGC), where information on suicide attempt (SA) had been collected^1^. MDD was diagnosed using structured psychiatric interviews according to international consensus criteria (DSM-IV, ICD-9, or ICD-10). Items from these interviews provided information on self-harm, suicidal ideation, plans and SA for patients with MDD. Patients with MDD endorsing SA were included as cases in this study. The controls for the primary genome-wide association study (GWAS) of SA included patients with MDD who did not endorse SA as well as healthy controls. MDD cases who were missing information on SA were excluded from the study. The healthy controls from PGC MDD cohorts were largely screened for the absence of depression and other psychiatric disorders (12/14 cohorts), however information on SA was not available for these individuals. All subjects were of European ancestry and gave written informed consent to participate in the source studies. The source, inclusion and exclusion criteria for each individual PGC MDD cohort have been reported in detail previously^2^, as well as the specific items used to ascertain information on SA from psychiatric interviews^1^.

**Psychiatric Genomics Consortium Bipolar Disorder**

Subjects were drawn from 22 bipolar disorder (BIP) case-control cohorts in the PGC, where information on SA had been collected^1^. As described for the PGC MDD cohorts, structured psychiatric interviews were used to diagnose BIP and ascertain information on SA. Cases and controls were defined in the same way as for the PGC MDD sample. The healthy controls from most PGC BIP cohorts were screened for the absence of lifetime psychiatric disorders. The source, inclusion and exclusion criteria for each individual PGC BIP cohort have been reported in detail previously^3^, as well as the specific items used to ascertain information on SA from psychiatric interviews^1^ .

**Psychiatric Genomics Consortium Schizophrenia**

Subjects were drawn from 9 schizophrenia (SCZ) case-control cohorts in the PGC, where information on SA had been collected^1^. The same procedures were used to make psychiatric diagnoses, ascertain information on SA and define cases and controls, as described previously for PGC MDD and BIP studies. The source, inclusion and exclusion criteria for each individual PGC SCZ cohort have been reported in detail previously^4^, as well as the specific items used to ascertain information on SA from psychiatric interviews^1^.

**Psychiatric Genomics Consortium Eating Disorders**

Subjects originated from 4 anorexia nervosa (AN) case-control cohorts in PGC, where information on SA had been collected. The ascertainment, phenotype measurement, and inclusion and exclusion criteria have been described previously for these cohorts^5,6^. The cohorts were the Children’s Hospital of Philadelphia/Price Foundation Collaborative Group (CHOP/PFCG) case-control cohort, and the France, Spain, and USA/Canada case cohorts from the Genetic Consortium for Anorexia Nervosa/Wellcome Trust Case Control Consortium-3 (GCAN/WTCCC-3) with controls sourced as described in Duncan et al^6^. Control cohorts from a similar geographic location and genotyping platform were preferentially sought. PGC AN cases had DSM-III-R or DSM-IV diagnoses of AN or EDNOS-AN (i.e., without the requirement of amenorrhea) based on structured diagnostic interviews. Controls had not been screened for AN but prevalence of lifetime AN is rare (~1%), nor had they been screened for SA. The same procedures described for the PGC MDD cohorts were used to define cases and controls.

**CONVERGE**

MDD cases were recruited from 58 provincial mental health centers and psychiatric departments within general hospitals, from 23 provinces in China^7^. Controls were recruited from patients undergoing minor surgery at general hospitals or local community centers. All subjects were Han Chinese women with four Han Chinese grandparents. Cases were excluded if they had a history of bipolar disorder, psychosis, or mental retardation. Cases were between ages 30-60 and had at least two episodes of MDD based on DSM-IV criteria, and with the first episode occurring between ages 14-50. They could not have abused drugs or alcohol prior to their first depressive episode. All subjects were interviewed using a computerized assessment program. The MDD diagnosis was determined using the Composite International Diagnostic Interview (WHO lifetime version 2.1; Chinese version). Cases were asked whether they had contemplated suicide during their worst depressive episode, and if so, whether they made a plan. Those who endorsed making a plan were asked whether they had attempted suicide. Controls were asked whether they had thought a lot about death or harming themselves and excluded if they responded in the affirmative after all disorders). Controls from Mannheim were blood donors who filled out a questionnaire including questions on mental and somatic health. For the current study the following information was used: self-report of psychiatric disorders, self-report of diagnosis of psychiatric disorder by a healthcare professional, and a questionnaire version of the SCID items for depression criteria A1–A9. The subgroup of subjects affirming at least one of the two SCID depression screening items were asked for their lifetime history of suicide attempts. Control subjects with a history of suicide attempt were excluded.

**Army STARRS**

Subjects come from several components of the Army Study To Assess Risk and Resilience in Servicemembers (STARRS): New Soldier Study (NSS), Pre/Post Deployment Study (PPDS), and Soldier Health Outcomes Study A (SHOS-A). Detailed information about the design and conduct of STARRS is available in a separate report^8^. Soldiers from the respective studies are unique and independent as confirmed by analysis of genetic relatedness. Suicidal behaviors were assessed using a version of the Columbia Suicidal Severity Rating Scale (C-SSRS)^9^ assessing lifetime occurrence of suicidal ideation (“Did you ever in your life have thoughts of killing yourself” or “Did you ever wish you were dead or would go to sleep and never wake up?”) and, among respondents who reported lifetime suicidal ideation, suicide plans (“Did you ever have any intention to act [on these thoughts/on that wish]?” and, if so, “Did you ever think about how you might kill yourself [e.g., taking pills, shooting yourself] or work out a plan of how to kill yourself?”) and suicide attempts (“Did you ever make a suicide attempt [i.e., purposefully hurt yourself with at least some intention to die]?”). For the primary GWAS of SA (n=670 cases), controls (n=10637)included individuals with no lifetime history of SA (who may or may not have a lifetime history of suicidal ideation). The GWAS of SA within psychiatric diagnosis included 376 cases of SA with MDD and 3447 individuals with MDD and no history of SA as controls.

**German Borderline Genomics Consortium**

Subjects were drawn from a GWAS sample on Borderline Personality Disorder^10^. The selected subjects consist of cases recruited in Berlin and Mannheim, and controls recruited in Mainz and from a sample of blood donors recruited in Mannheim, Germany. The diagnosis of Borderline Personality Disorder was assigned according to DSM-IV criteria on the basis of structured clinical interviews (either IPDE or SCID-II). Life-time attempt of suicide and, in the case of a positive answer, the number of attempts were documented. Diagnostic interviews were conducted by trained and experienced raters. Controls from Mainz were screened for a list of psychiatric disorders (panic disorder, agoraphobia, social phobia, specific phobia, generalized anxiety disorder, PTSD, obsessive-compulsive disorder, major depression, dysthymia, mania, hypochondriacal disorder, somatoform disorder, pain, conversion disorders, anorexia nervosa, bulimia nervosa, harmful alcohol use, alcoholism, harmful drug use, drug addiction, schizophrenia, schizotypal disorders). Controls from Mannheim were blood donors who filled out a questionnaire including questions on mental and somatic health. For the current study the following information was used: self-report of psychiatric disorders, self-report of diagnosis of psychiatric disorder by a healthcare professional, and a questionnaire version of the SCID items for depression criteria A1–A9. The subgroup of subjects affirming at least one of the two SCID depression screening items were asked for their lifetime history of suicide attempts. Control subjects with a history of suicide attempt were excluded.

**Grady Trauma Project (GTP)**

The subjects for this study were part of a larger investigation of genetic and environmental factors that predict the response to stressful life events in a predominantly African American, urban population of low socioeconomic status. Participants were approached while in the waiting rooms of primary care, diabetes, or obstetrical-gynecological clinics of Grady Memorial Hospital in Atlanta, Georgia. Screen interviews, including participants’ demographic information (e.g., self-identified race, sex, and age), prior hospitalization for psychiatric diseases, and psychiatric symptoms including Posttraumatic Stress Disorder (PTSD), depression, schizophrenia, and bipolar disorder, were completed on site. Suicide attempt history was assessed based on self-report (yes/no) when obtaining demographic information. Further details regarding the GTP dataset can be found in Gillespie et al^11^. Written and verbal informed consent was obtained for all participants and all procedures in this study were approved by the institutional review boards of Emory University School of Medicine and Grady Memorial Hospital, Atlanta, Georgia. The primary GWAS of SA included 669 cases and 4473 controls and the GWAS of SA within psychiatric diagnosis included 355 cases and 1116 controls, all with PTSD.

**UK Biobank**

The UK Biobank is a prospective cohort study of 501,726 individuals, recruited at 23 centres across the United Kingdom^12^. Extensive phenotypic data are available for UK Biobank participants from health records and questionnaires, including an online follow-up questionnaire focussing on mental health (Mental Health Questionnaire, MHQ [Resource 22 on [http://biobank.ctsu.ox.ac.uk](http://biobank.ctsu.ox.ac.uk/)]). A total of 157,366 participants provided responses to an online mental health questionnaire (MHQ) as a follow up to initial phenotyping in the UK Biobank sample. Of these, 6,872 were asked this question from Data-Field 20483, Category: Self-harm behaviors, “Have you harmed yourself with the intention of ending your life?” Most participants were not asked this question as it required a positive response to a previous self-harm question. In total, 3,563 of 6,872 respondents indicated “yes”, 3,089 responded “no” and 220 preferred not to answer. In an effort to maximize power and because the phenotype is rare, we included all UK Biobank participants as controls in the primary GWAS of SA except for those responding yes to attempting suicide; this includes those that did not take the mental health assessment at all and those who preferred not to answer. After reducing our sample to a set of homogenous individuals with white British ancestry, we retained case-control data of 2,433 individuals having attempted suicide and 334,766 controls.

For the GWAS of SA within psychiatric diagnosis, cases were individuals with a mood disorder who reported a lifetime suicide attempt and controls were individuals with a mood disorder who reported no lifetime deliberate self-harm. Participants were classified as having a mood disorder if they either self-reported a professional diagnosis of depression or bipolar disorder as part of the MHQ [UK Biobank field 20544, responses 10 or 11] or if they met criteria for depression on MHQ questions derived from the Composite International Diagnostic Interview (CIDI). To meet these latter criteria, participants must have reported ever feeling depressed [UK Biobank field 20446] or anhedonic [UK Biobank field 20441] for two weeks in a row, for at least most of the day [UK Biobank field 20436] almost every day [UK Biobank field 20439] with more than a little interference with daily activities [UK Biobank field 20440]. In addition, they must have reported experiencing at least five of the following symptoms in this period of depression or anhedonia: depression [UK Biobank field 20446], anhedonia [UK Biobank field 20441], tiredness [UK Biobank field 20449], weight change [UK Biobank field 20536], sleep change [UK Biobank field 20532], loss of concentration [UK Biobank field 20435], worthlessness [UK Biobank field 20450] and thoughts of death [UK Biobank field 20437]. The MHQ additionally contained screening questions for bipolar disorder^13^. However, for the purpose of defining potential bipolar disorder, all individuals scoring positively on these screening questions were also required to meet the CIDI depression criteria defined above, and as such participants with potential bipolar disorder were a subset of those meeting criteria for depression. Individuals who self-reported a professional diagnosis of psychosis on the MHQ [UK Biobank field 20544, responses 2 or 3] were excluded. Cases of suicide attempt with mood disorders (n=2149) were defined as those who answered yes to the question “Have you ever harmed yourself with the intention to end your life?” [UK Biobank field f20483]. Controls with mood disorders were defined as those who reported no self-harm on the MHQ (n=35912).

**Taiwan Major Depressive Disorder**

MDD patients were drawn from a family study of mood disorders in Taiwan. Patients aged between 18 to 70 years, who met diagnostic criteria of MDD using the Diagnostic and Statistical Manual of Mental Disorders, fourth edition (DSM-IV) were consecutively referred by psychiatrists in clinical settings. Exclusion criteria include patients diagnosed with schizophrenia, schizoaffective or substance-induced mood disorders. SA was measured based on the Chinese version of the Composite International Diagnostic Interview (CIDI), the modified Schedule of Affective Disorder and Schizophrenia-Lifetime (SADS-L), or Hamilton Depression Rating Scale (HAM-D). SA cases were identified with the answer of “Yes” in item “Have you ever attempted suicide?” in CIDI, or “Yes” in item “Have you ever had suicide attempt and really wanted to die?” in SADS-L, or the score of suicide item equal or greater than 3 in HAM-D. Each participant was interviewed with either of the aforementioned instruments. The Taiwan MDD cohort was included in the GWAS of SA within psychiatric diagnosis only and 222 MDD cases with a history of SA were compared with 318 MDD cases without a history of SA as controls. More details regarding sample recruitment were described elsewhere^14^.

**Taiwan Bipolar Disorder**

The inclusion and exclusion criteria of Taiwan BIP cohort is the same as those of Taiwan MDD. Patients who met bipolar disorder subtype I or bipolar disorder subtype II using the DSM-IV were referred by psychiatrists in clinical settings. CIDI or SADS-L were used to collect SA information through interviews as the same in Taiwan MDD data. SA cases were defined as subjects who answered “Yes” for the SA item (n=235) and controls were individuals with BIP who answered “No” to the SA item (n=397). Details of sample recruitment and assessment please refer to Tsai et a ^15^.

**Taiwan Schizophrenia**

Schizophrenia patients were recruited from two study projects: Schizophrenia Trio Genomic Research in Taiwan (S-TOGET) and Taiwan Schizophrenia Linkage Study (TSLS). Participants enrolled from the S-TOGET project were parent-proband trio samples. There were in total 3008 families with probands diagnosed as schizophrenia or schizoaffective disorder based on DSM-IV in psychiatric hospitals or community care centers nation-wide in Taiwan. After excluding patients without suicide information, there were in total 1119 probands retained. Details about the ascertainment of the S-TOGET sample can be found elsewhere^16^.

Samples from TSLS were probands with clinical record of schizophrenia or depressive type of schizoaffective disorder from hospitals or psychiatric service stations. According to the inclusion criteria of TSLS, proband had to have at least one other sibling affected with similar diagnosis. In the present study, only 94 probands but not family members were included in analysis. More detailed information of TSLS is included in a previous report^17^.

SA information for SCZ patients in both S-TOGET and TSLS were measured using the Diagnostic Interview for Genetic Studies (DIGS), a semi-structured psychiatric interview. The item related to suicide in this instrument was ”Have you ever attempted suicide (YES/NO)?” SA cases were defined as those who answered “YES”.

**iPSYCH**

All individuals included in this study were a part of the Danish iPSYCH 2012 population-based case-control cohort^18^. SA cases were identified according to information available from the Danish Psychiatric Central Research Register and the National Registry of Patients both complete until December 31, 2016. SA cases were identified as individuals with ICD-10 diagnoses of SA (ICD-10: X60-X84, equivalent to intentional self-harm), with SA indicated as ‘reason for contact’ in the registers, and with a main diagnosis of poisoning (ICD-10: T39, T42, T43, and T58). The SA case group also included individuals with a diagnosis in the ICD-10: F chapter as main diagnosis and report of poisoning by drugs or other substances (ICD-10: T36–T50, T52–T60) or injuries to hand, wrist, and forearm (ICD-10: S51, S55, S59, S61, S65, S69). Individuals who died by suicide according to Cause of Death Register available until December 31, 2015 were also classified as SA cases. Only contacts starting at age 10 years old or older were considered to be reliably reported SA cases. Individuals not fulfilling any of the above SA case criteria were considered to be controls for the primary GWAS of SA. The study was approved by the regional Danish ethics committee and the Danish Data Protection Agency.

**Janssen**

The Janssen lifetime suicide attempt cohort consisted of subjects of European ancestry and was drawn from multiple clinical trial samples (NCT00044681, NCT00397033, NCT00412373, NCT00334126, NCT01193153, NCT00094926) conducted by Janssen Research & Development, LLC as well healthy control samples from NINDS Human Genetics Repository (neurologically normal European control panel NDPT020, NDPT079, NDPT084, NDPT090, NDPT093, NDPT094, NDPT095, NDPT096, NDPT098, and NDPT099) managed by Coriell Institute for Medical Research (Camden, NJ) and from BioIVT (Westbury, NY). A subset of clinical trial samples (NCT00334126, NCT00397033, NCT00412373, and NCT00044681) was described previously^2,4,19,20^^,^^10,21^ . The clinical diagnosis of MDD, schizophrenia, schizoaffective disorder, and bipolar disorder in Janssen clinical studies were based on expert clinician interviews conducted using DSM-IV-TR criteria. In two studies (NCT00397033 and NCT00412373), the diagnosis of schizoaffective disorder was confirmed using an interview based SCID (Structured Clinical Interview for DSM-IV-TR). The lifetime suicide attempt history was based on detailed clinical interview and medical records. The disease diagnosis for Coriell cohort was based on medical history including bipolar/manic depressive disorder, depression, schizophrenia, and suicide attempt. All patients who provided genetic samples gave written informed consent to the genetic testing. The primary GWAS included 255 cases and 1684 controls.

**Genetic Investigation of Suicide and SA (GISS)**

Sample recruitment, selection criteria, demographics, ancestry and psychiatric diagnoses have been described previously^22,23^^,^^24^^,^^25^. Briefly, lifetime SA was the main outcome ascertained in the offspring of nuclear family trios (all complete with both biological parents and one SA offspring per trio; n = 660). Trios were collected in Ukraine by first recruiting offspring from emergency care due to a severe SA, defined as a score of = 2 on the Medical Damage Rating Scale (MDS),^26^ which represented the primary ascertainment criteria for inclusion. Persons who have engaged in suicidal thoughts without actual behavior would not be included. Other exclusion criteria were subject adopted, mental retardation, organic mental disorder, or other chronic medical illness involving the central nervous system. The SA were verified independently by both parents, the suicide attempter and by examining medical records. The suicidal intent of the SA was assessed by using both objective (levels of precaution) and subjective (intent to die) aspects.^27^ Previous life-time SA was documented, as well as the history of suicides in family and relatives. Secondary outcomes included ICD-10 diagnoses according to the Composite International Diagnostic Interview (CIDI), personality traits according to the NEO personality inventory (NEO-PI-R), levels of anger, Beck’s depression inventory, the WHO well-being index and the Global assessment of functioning (GAF) scale. Exposures to lifetime stressful and traumatic life-events (SLEs) were also assessed. Overall, the SA offspring included 51.1% males (n=337)/48.9% females (n=323), with mean ages of 24.6 (S.D. ± 7.3)/23.8 (S.D. ± 7.1) years, and 94.4% (n=318)/93.2% (n=301) of the SA subjects had = 3 Ukrainian or Russian grandparents, respectively. Overall, n=498 SA subjects did not have any of the major psychiatric diagnoses, e.g. schizophrenia (ICD-10 code F20), schizoaffective disorder (F25) or moderate / severe depression diagnoses (F32-33). The collection of research subjects followed the code of ethics of the World Medical Association (Declaration of Helsinki), and written consent was obtained. The study was approved by the Research Ethics Committee at the Karolinska Institute (Dnr 97–188) and by the Ministry of Health in Ukraine.

**Australian Genetics of Depression Study and QSkin**

Sample recruitment has been described in detail elsewhere^28^. In brief, two separate approaches were used. First, a nationwide recruitment based on antidepressant prescription history was possible through the Australian Government Department of Human Services (DHS; now known as *Services Australia*) which keeps the pharmaceutical benefits scheme national database. After obtaining the relevant ethics approvals by both the DHS and QIMR Berghofer, the researchers engaged the DHS to send ~110,000 invitations, in two waves, to participants with a prescription history of antidepressants. The second strategy consisted of a media publicity campaign launched on April 4, 2017. Under both strategies, participants were directed to a website which provided information on the study and collected informed consent for participation, including donation of a saliva sample for genotyping. Consenting participants were then referred to a modular online questionnaire consisting of a *core module*, which assessed essential clinical information on mental health diagnoses, treatment history, effectiveness and side effects, and multiple satellite modules. As of the 3 September 2018, 20,689 (75% female, mean age 43 years) participants had completed the online core module and provided consent to donate a saliva sample. Most of them (19,803) reported being diagnosed with depression and 17,698 met the DSM-5 criteria for a major depressive episode. SA was assessed using the suicidal ideation attributes scale (SIDAS)^29^ and defined as an episode of self-harm with some intent to die. Healthy controls were ascertained from the QSkin Sun & Health Study (QSkin). QSkin consists of a randomly sampled cohort of individuals between 40 and 69 years from the state of Queensland^30^. A genetic study within QSkin has been initiated following a similar protocol for DNA collection by mail. During saliva donation participants were directed to fill in a short questionnaire on previous diagnoses of physician and psychiatric disorders^28^. Due to a lack of suicide attempt assessment in QSKIN, participants with a history of any psychiatric disorder were excluded. The final samples (unrelated individuals with genotype data passing quality control filters) comprised 2,792 SA cases and 20,193 controls for the primary GWAS of SA and 2,792 SA cases and 8,718 individuals with depression without a history of SA, for the GWAS of SA within psychiatric diagnosis.

**Yale-Penn (European and African American cohorts)**

Participants in this study were recruited from five sites in the eastern United States, for studies of the genetics of drug or alcohol dependence - the Yale-Penn study^31^^,^^32^. All participants were interviewed using the Semi-Structured Assessment for Drug Dependence and Alcoholism (SSADDA)^33^, which contains several items relevant to suicidal behavior. Specifically, if a participant responded “Yes” to the item “Have you ever tried to kill yourself?” they were considered as a case. If they responded “No” to both this question and also “Have you ever thought about killing yourself?” they were treated as a control. Participants provided written informed consent and the study was approved by the institutional review board at each participating site (Yale Human Research Protection Program, University of Pennsylvania Institutional Review Board, University of Connecticut Human Subjects Protection Program, Medical University of South Carolina Institutional Review Board for Human Research, and the McLean Hospital Institutional Review Board).

**Columbia University**

Sample selection and diagnoses have been described previously^34^. Briefly, 2,382 unrelated individuals of European ancestry from three sites (New York, USA; Montreal, Canada, Munich, Germany) were recruited between 1991 and 2011. and gave written informed consent to participate as required by the relevant Institutional Review Boards. In total, 1,765 live subjects and 617 postmortem subjects were genotyped using the Illumina Omni1-Quad Beadchip (1,014,770 SNPs). Subjects with SA were defined as individuals who died by suicide or attempted suicide and in 64 percent of cases were known to have had a DSM-IV defined MDD, diagnosed by a SCID I structured clinical interview. SA was defined as a self-injurious act that has at least partial intent to end one’s life. A group of subjects with MDD and without a history of a suicide attempt provided a psychiatric control group. Additionally, unrelated healthy volunteers of German descent were randomly selected from the general population of Munich, Germany, and contacted by mail. In New York and Montreal healthy volunteers were solicited through advertising. The Montreal sample was confined to French Canadians, whereas the New York sample included Europeans of any origin. Healthy volunteers were assessed by psychiatrists or clinical psychologists and evaluated using the SCID-NP version and were free of axis I diagnoses, cluster B personality disorder, substance use disorder and lifetime history of a suicide attempt. One thousand nine hundred and forty-two of the genotyped samples passed QC procedures. After filtering of ethnic outliers, 1,810 subjects remained: 925 males and 885 females, 577 cases with suicidal behavior (260 suicide attempters and 317 suicides), and 1,233 subjects without SB (1,096 live subjects without a history of attempt and 137 sudden death victims). A breakdown of subjects by diagnosis and site has been summarized in a table previously published^34^.

**Japan**

For the Japanese cohort, we used data from 746 suicide decedents (386 suicides who died between June 1996 and July 2012 in the 1st set and 360 suicides who died between August 2012 and February 2017 in the 2nd set)^35^. Autopsies on suicides were performed and the decision of assigning the status “suicide” was made through discussion with the Medical Examiner’s Office of the Hyogo Prefecture and the Division of Legal Medicine in the Kobe University Graduate School of Medicine. For non-suicide controls, we used genome-wide genotype data from 14,049 subjects (7,458 controls in the 1st set and 6,591 controls in the 2nd set) in the Biobank Japan project who had been genotyped as case subjects for non-psychiatric disorders and healthy volunteers.

**University of Utah**

The Utah GWAS samples included 4380 persons who died by suicide and 20,702 ancestry matched controls, genotyped on the PsychChip by the Psychiatric Genomics Consortium. Suicide cause-of-death determination results from a detailed investigation, done by the centralized Utah State Office of the Medical Examiner, of the scene of the death and circumstances of death, determination of medical conditions by full autopsy, review of medical and other public records concerning the case, interviews with survivors, in addition to standard toxicology workups. Suicide determination is traditionally made quite conservatively due to its impact on surviving relatives. DNA from suicide deaths was extracted from whole blood using the Qiagen Autopure LS automated DNA extractor (www.qiagen.com). Controls for the University of Utah sample were drawn from the following cohorts which had been genotyped on the PsychChip by the Psychiatric Genomics Consortium. The boldfaced first line for each sample is study PI, PubMed ID if published, study name, PGC internal tag or study identifier and number of controls.

**Braff D | PMID: 17035358 | Consortium on the Genetics of Schizophrenia (COGS-1) | cogs1 (n=416)**

Participants were recruited from seven sites in the United States, as part of the Consortium on the Genetics of Schizophrenia (COGS-1) family study: University of California at San Diego (UCSD) and Los Angeles (UCLA), University of Colorado (CUHSC), Mount Sinai School of Medicine (MSSM), University of Pennsylvania (PENN), Harvard Medical School (HMS) and University of Washington (UW). Participants provided written informed consent and the study was approved by the institutional review board at each participating site. Unrelated community comparison subjects without personal or family history of psychosis were recruited. To parallel psychiatric comorbidity in relatives of probands, nonpsychotic axis I psychopathology was accepted in approximately 30% of the community comparison subjects but clinical stability and/or remission was required. Subjects were excluded if they had ECT in the last 6 months, substance abuse or dependence, head injury with loss of consciousness >15 minutes, and for any neurological or severe systemic illness. All subjects underwent a standardized clinical assessment using the Diagnostic Interview for Genetic Studies (DIGS) Details of the ascertainment, diagnostic, and screening procedures are provided elsewhere^36^. Written informed consent was obtained for each subject per local IRB protocols.

**Sonuga-Barke E | Not published | South Hampshire ADHD Register - University of Southampton (SHaRE)| barke (n=65)**

SHaRE was a clinical database including child and adolescent patients from CAMHS clinics across the south coast in the UK. Controls were ascertained from local schools of a similar age and sex to patients. All undertook a detailed clinical and psychometric assessment. DNA was extracted from cheek cells and genotyped on the PsychChip by the Psychiatric Genetics Consortium.

**Baune, BT, Dannlowski, U | Not published | [PGC Psychchip] | bdtrs (n=722)**

The Bipolar Disorder treatment response Study (BP-TRS) comprises BD inpatient cases and screened controls of European background. Psychiatric diagnosis of Bipolar Disorders was ascertained using SCID or MINI 6.0 using DSM-IV criteria in a face-to-face interview by a trained psychologist / psychiatrist for both cases and controls. Healthy controls were included if no current or lifetime psychiatric diagnosis was identified.

**Bau C | Not published | [PGC Psychchip] |clait (n=272)**

The Brazilian ADHD Porto Alegre Cohort is part of the International Multi-centre persistent ADHD CollaboraTion (IMpACT). It comprises adult patients and controls ascertained in the Hospital de Clínicas de Porto Alegre. Individuals from the control group were recruited in the blood donation centre. The inclusion criteria were (A) being Brazilian of European descent and (B) aged 18 years or older. The exclusion criteria were: (A) positive screening in the 6-item Adult ADHD Self-Rated Scale Screener (ASRS), (B) evidence of a clinically significant neurological disease that might affect cognition (e.g., delirium, dementia, epilepsy, head trauma, and multiple sclerosis), and (C) current or past history of psychosis. The control group also underwent a broad sociodemographic assessment and a screening for comorbidities with the SCID epidemiologic screener. The study was carried out in accordance with the Declaration of Helsinki, and all participants signed an informed consent form previously approved by the institutional review board of the hospital (No. 00000921).

**Ophoff R, Posthuma D, Lochner C, Franke B | Not published | [PGC Psychchip] | dutch (n=1111)**

The following is an aggregation of Dutch population control samples. Ophoff R: Controls were collected at different sites in the Netherlands and were volunteers with no psychiatric history after screening with the (MINI^37^). Ethical approval was provided by UCLA and local ethics committees and all participants gave written informed consent. Lochner C: Controls include population based-controls ascertained from blood banks and controls recruited through university campuses and newspaper advertisements, who underwent a psychiatric interview and had no current or lifetime psychiatric disorder^38^^,^^39^. Franke B: The controls included are healthy individuals from the Dutch part of the International Multicenter ADHD Genetics (IMAGE) project^40^^,^^41^. Posthuma D: Data were provided for 960 unscreened Dutch population controls from the Netherlands Study of Cognition, Environment and Genes (NESCOG)^42^. The study was approved by the institutional review board of Vrije Universiteit Amsterdam and participants provided informed consent.

**Gawlik M | Not published | [PGC Psychchip] | gawli (n=572)**

Patients were recruited at the Department of Psychiatry, Psychosomatics and Psychotherapy, University of Würzburg, Germany. Diagnosis according to DSM-IV (Diagnostic and Statistical Manual of Mental Disorders-fourth edition) was made by the best estimate lifetime diagnosis method, based on all available information, including medical records, and the family history method. Healthy control subjects were recruited from the blood donor centre at the University of Würzburg.

**Reif, A | Not published | [PGC Psychchip] | germ1 (n=1072)**

Control subjects were healthy participants who were recruited from the community of the same region as cases for a genetic study of bipolar disorder. They were of European descent and fluent in German. Exclusion criteria were manifest or lifetime DSM-IV axis I disorder, severe medical conditions, intake of psychoactive medication as well as alcohol abuse or abuse of illicit drugs. Absence of DSM-IV axis I disorder was ascertained using the German versions of the Mini International Psychiatric Interview. IQ was above 85 as ascertained by the German version of the Culture Fair Intelligence Test 2^43^. Study protocols were reviewed and approved by the ethical committee of the Medical Faculty of the University of Frankfurt. All subjects provided written informed consent.

**Pato, C | Not published | [PGC Psychchip] | gpcw1 (n=1858)**

Genomic Psychiatry Consortium (GPC) cases and controls were collected via the University of Southern California healthcare system, as previously described^44^. Using a combination of focused, direct interviews and data extraction from medical records, diagnoses were established using the OPCRIT and were based on DSM-IV-TR criteria. Age and gender-matched controls were ascertained from the University of Southern California health system and assessed using a validated screening instrument and medical records.

**Spalletta G | Not published | [PGC Psychchip] | spal1 (n=40)**

The IRCCS Santa Lucia Foundation of Rome, Italy, sample of healthy people was recruited from the hospital personnel and using local advertisement and was screened for a current or lifetime history of psychiatric and personality disorders according to the DSM-IV-TR, using the SCID_non patient edition. Exclusion criteria are as follows: history of alcohol or drug abuse in the last 2 years before the assessment, lifetime drug dependence, traumatic head injury with loss of consciousness, past or present major medical illnesses or neurological disorders, any psychiatric disorders or mental retardation, dementia or cognitive deterioration according to DSM-IV-TR criteria and Mini-Mental State Examination (MMSE) normative data within the Italian population, any potential brain abnormalities and vascular lesions as apparent on conventional T1 and T2 weighted and FLAIR magnetic resonance imaging scans. All included subjects signed an informed consent approved by the local ethic committee.

**Serretti A | Not published | [PGC Psychchip] | serr1 (n=147)**

The sample has been described previously (Mapping genomic loci prioritises genes and implicates synaptic biology in schizophrenia. The Schizophrenia Working Group of the Psychiatric Genomics Consortium - manuscript submitted). Briefly, healthy controls were recruited and included in the context of a medical screening, no formal psychiatric interview was administered but the absence of major and invalidating psychiatric disorder was recorded. The study was approved by the San Raffaele Pisana and by ASL RME Ethics Committees, and all participants provided written informed consent.

**Nurnberger JI, Edenberg HJ, McInnis M, Wilcox HC, Glowinski AL, Fullerton JM | PMID: 29173741| [PGC Psychchip] | iupui (n=65)**

Young people with familial risk of bipolar disorder and healthy controls (aged 12 to 21 years) were ascertained from 4 independent sites in the United States: Johns Hopkins University, University of Michigan, Washington University in St. Louis, and Indiana University^45,46^. Recruitment procedures and clinical batteries were aligned with those also employed by the Australian Bipolar High Risk Study site (represented in the *neura* cohort). Control parents were recruited through general medicine clinics, motor vehicle records, and campus advertising. Exclusion criteria for control parents included BPI, BPII, recurrent major depression, schizoaffective disorder, or schizophrenia in either parent; we also excluded parents with a first-degree relative with a psychiatric hospitalization.^45,46^

**Rivera M, Cervilla J.A | Not published | [PGC Psychchip] | marg1 (n=1354)**

All control participants were part of the PISMA study, the first epidemiological study focussed on mental health disorders, and their associated factors, ever undertaken in a representative sample of the entire Andalusian population (Spain)^47^. This was a cross-sectional study targeting a large representative stratified sample of community-dwelling Andalusian adults between 18 and 75 years of age. All provinces in the Andalusian community were included. A comprehensive account of risk, neuropsychological, personality and psychiatric assessments were undergone in the PISMA sample (4507 participants) and have been reported elsewhere^47^. Interviews were undertaken by psychologists specially trained by the PI of the study (J.A.Cervilla). Interviewers demonstrated sufficient knowledge on both interviewing techniques on all protocol scales and inventories, most of which had been originally designed for administration by lay-interviewers. Teaching techniques used included lectures, role playing between interviewers and scoring of videoed interviews held by experts on volunteers. All instruments used had, nonetheless, previously been validated and demonstrated sufficient inter and intra-rater reliabilities along with most other psychometric properties. Specific inter-rater reliabilities between interviewers on such instruments after training sessions were high ^47^. The psychiatric interview to identify mental disorder (MDs) diagnoses was performed using the MINI, which generates diagnoses compatible with both Axis I DSM-4 and ICD-10 criteria for 16 common MDs, two additional diagnoses of major depression with melancholia and mood disorder with psychotic symptoms, one Axis II diagnosis (antisocial personality disorder), as well as a suicidal risk estimate. A biological sample was obtained from each participant using the Oragene DNA saliva collection kit (OG-500; DNA Genotek Inc.). DNA extraction was performed using Oragene saliva Kit protocol as per manufacturer's instructions. Samples were genotyped on the PsychChip array at the Stanley Centre. Participants in the PISMA study gave their informed consent. The study had ethics approval granted by “Comité de Ética en Investigación, Universidad de Granada” which permits inclusion of the data in meta-analyses. Genotype data can be accessed for secondary analysis after explicit PI approval. This study was funded by Consejería de Innovación, Proyecto de Excelencia CTS-2010-6682.

**Liberzon, I., King, A.P., Galea, S., Calabrese, J. | PMID 25162199 | Ohio Army National Guard (OHARNG) | mich1 (n=111)**

The Ohio Army National Guard (OHARNG) study^48^ was a prospective, longitudinal study of Ohio Army National Guard soldiers who were initially recruited and had a comprehensive intake psychiatric assessment (CATI telephone interview with standardized instruments) after their unit was activated and before their unit was deployed to Iraq or Afghanistan. Saliva samples for DNA (Oragene tube) were obtained at follow-up assessment Waves 2-4 by return mail to our lab, and DNA for GWAS analysis was isolated and stored. The control subjects included in this sample were healthy European-American male soldiers who did not meet criteria for PTSD, MDD, or any other psychiatric diagnosis at intake or any follow-up assessment Wave. Controls (N=125) were matched by age and lifetime “trauma load” to N=125 PTSD cases within the same cohort. A total of 37 potentially traumatic events were identified using the Clinician-Administered PTSD Scale (CAPS-IV)^49^ and the 1996 Detroit Area Survey of Trauma^49^ PTSD symptoms were assessed using a 17-item structured interview scale derived from the PTSD Checklist (PCL) for DSM-IV performed by trained lay telephone interviewers using epidemiological methods (forced choice symptom severity range, 1-5). Reliability of the telephone interview was validated against the criterion standard (in-person CAPS interview by mental health professional) in a clinical subsample (n = 500), demonstrating high specificity (0.92)^50^. Respondents were considered to have a diagnosis (cases) if lifetime DSM-IV PTSD criteria were met. Respondents were considered to have a current diagnosis if past month DSM-IV criteria were met. The PCL calculates PTSD symptom severity, which ranged from 17 to 85, by sum of scores of items endorsed. For this cohort (125 cases, 125 controls), the mean severity was 38.4 and the standard deviation 17.6.

**Fullerton JM, Mitchell PB, Schofield PR, Green MJ, Weickert CS, Weickert TW | Not published | [PGC Psychchip] | neura (n=161)**

The NeuRA collection comprised psychiatrically screened healthy control subjects from three cohort studies ascertained in Australia: the Bipolar High Risk “kids and sibs” study^46^^,^^51^, the Imaging Genetics in Psychosis Study (IGP)^52^ and the Cognitive and Affective Symptoms of Schizophrenia Intervention (CASSI) trial^53^. The Bipolar High Risk study is a collaborative study with 4 US sites (represented in the *iupui* cohort), and young Australian participants aged 12-30^45^. Healthy controls from each study were recruited from the community, had no personal lifetime history of a DSM-IV Axis-I diagnosis as determined by psychiatric interview, and no history of psychotic disorders among first-degree biological relatives.

**Koenen K | Not published | Nurses’ Health Study II| nhsii (n=739)**

In 2008 the [Trauma and PTSD Screening Questionnaire](https://drive.google.com/open?id=1dnSrXQZ1ACeURNVZiQ7AtnLkcBmPDPxE) was mailed to 60,804 Nurses’ Health Study II (NHSII) participants who had completed recent questionnaires. The response rate was 84% (N = 50,953). We identified 17,666 women for diagnostic interviews who reported exposure to at least one traumatic event on the modified Brief Trauma Questionnaire and agreed to be interviewed^54,55^. We then identified probable PTSD cases and probable controls using Breslau’s lifetime PTSD screen^56^, which classifies PTSD cases with 80% sensitivity, 97% specificity, 71% positive predictive value, and 98% negative predictive value. We randomly selected 2,112 probable PTSD cases and 2,001 probable controls for diagnostic interviews. The Partners Human Research Committee approved this study; the protocol has been published^57^.

PTSD was then assessed using the PTSD Checklist (PCL-C), a 17-item self-report measure of DSM-IV PTSD symptoms^58,59^. Participants rated each of the 17 symptoms on a scale indicating how much they had been bothered by a particular symptom as a result of the event, from “not at all” to “extremely.” The Checklist assesses re-experiencing symptoms (Criterion B), avoidance/numbing symptoms (Criterion C), and arousal symptoms (Criterion D). To be a PTSD case, respondents must have reported experiencing one or more of the 5 re-experiencing symptoms, 3 or more of the 7 avoidance/numbing symptoms, and 2 or more of the 5 arousal symptoms at least “moderately.” Additional questions assessed the other three DSM-IV criteria: intense fear, horror, or helplessness in response to the event (Criterion A2), symptom duration of at least one month (Criterion E), and clinically significant impairment in functioning due to symptoms (Criterion F). The PCL-C had excellent internal consistency (Cronbach’s α=0.87). Respondents were considered affected by lifetime PTSD if all six DSM-IV criteria were met in reference to the worst event.

**Krebs M-O | Not published | [PGC Psychchip] | paris n=420**

Controls from the Psydev Paris cohort were healthy unrelated French adults (both genders) recruited from among staff members at the GHU Paris or from physiotherapist schools as part of a study PsyDev (Promotor Inserm RBM03-021). They gave their written consent after receiving a full descrip­tion of the study and study procedures were approved by the French ethics committees CPP Paris Ile de France 4 and were in accordance with the Declaration of Helsinki. They were screened for medical and psychiatric history either using the Diagnosis Interview for Genetic Studies (DIGS version 3.0) conducted by trained psychiatrists and psycholo­gists and/or self-rated questionnaires followed by face-to- face interviews. Exclusion criteria included personal or in first degree relatives with psychiatric history, personal history of neurologic signs, unstable medical condition, pregnancy and substance dependence. All controls were of European ancestry ("Caucasian") and were born in France.

**Campion D, Laurent C, Levinson D | Not published | [PGC Psychchip] | rouen (n=190)**

Controls from the Rouen cohort were recruited from among staff members and blood donors at the Centre Hospitalier Universitaire Rouen (France) as part of a study on hyperprolactinemia in schizophrenia^60^. All controls were of European ancestry and were born in France. All controls denied (by self-report in response to direct questions) any history of psychiatric disorder in themselves or in first-degree relatives or current use of medications or drugs other than oral contraceptives in women. The protocol was approved by the appropriate regional ethics committee. All participants gave written informed consent.

**Gareeva, A; Khusnutdinova, E; Escott-Price, V | Not published | [PGC Psychchip] | russ1 n=344**

All controls have a negative family history for neuro-psychiatric disorders. For all individuals key phenotypic information has been collected, including information about sex, age, ethnicity, age at onset and family history of psychiatric disorders. All subjects have provided written and informed consent. This study has been approved by the local bioethical committee of the Institute of Biochemistry and Genetics of Ufa Federal Research Center of the Russian Academy of Sciences (IBG UFRC RAS). Peripheral blood was taken from all participants of the study. DNA was extracted from peripheral blood by the phenol and chloroform method.

**Perlis, R; Sklar, P; Smoller, J| Not published | [PGC Psychchip] | smol0 (n=1052), smol2 (n=493), smol3 (n=555)**

Perlis, R; Sklar, P; Smoller, J: EHR data were obtained from a health care system of more than 4.6 million patients^61^ spanning more than 20 years. Experienced clinicians reviewed charts to identify text features and coded data consistent or inconsistent with a diagnosis of bipolar disorder. Natural language processing was used to train a diagnostic algorithm with 95% specificity for classifying bipolar disorder. Filtered coded data were used to derive three additional classification rules for case subjects and one for control subjects. No EHR-classified control subject received a diagnosis of bipolar disorder on the basis of direct interview (positive predictive value (PPV)=1.0). For most subphenotypes, PPV exceeded 0.80. The EHR-based classifications were used to accrue bipolar disorder cases and controls for genetic analyses. Samples were genotyped on the Psychchip array.

**Ribases M | PMID 32279069 | [PGC Psychchip] | span1 (n=2054), span2 (n=430)**

The Spanish controls were part of the Mental-Cat clinical sample or the INSchool population-based cohort. A total of 1,774 controls from the Mental-Cat cohort (60.5% males) were evaluated and recruited prospectively from a restricted geographic area at the Hospital Universitari Vall d’Hebron of Barcelona (Spain) and consisted of unrelated healthy blood donors^62^. The INSchool sample consisting of 771 children (76.2% males) from schools in Catalonia were involved for screening using the Achenbach System of Empirically Based Assessment (ASEBA) with the Child Behavior Checklist CBCL/4-18 (completed by parents or surrogates), the Teacher Report Form TRF/5-18 (completed by teachers and other school staff) and the Youth Self-Report YSR/11-18 (completed by youths); the Strengths and Difficulties Questionnaire (SDQ) and the Conner’s ADHD Rating Scales (Parents and Teachers). Genomic DNA samples were obtained either from peripheral blood lymphocytes by the salting out procedure or from saliva using the Oragene DNA Self-Collection Kit (DNA Genotek, Kanata, Ontario Canada). DNA concentrations were determined using the Pico- Green dsDNA Quantitation Kit (Molecular Probes, Eugene, OR) and genotyped with the Illumina Infinium PsychArray-24 v1.1 at the Genomics Platform of the Broad Institute. The study was approved by the Clinical Research Ethics Committee (CREC) of Hospital Universitari Vall d'Hebron, all methods were performed in accordance with the relevant guidelines and regulations and written informed consent was obtained from participant parents before inclusion into the study. Detailed information has been published previously^62^.

**Landen M, Hillert J, Alfredsson L | Not published | [PGC Psychchip] | swed1 (n=2886)**

Population-based controls, randomly selected from the Swedish national population register, were collected as part of two case-control studies of multiple sclerosis: GEMS (Genes and Environment in Multiple Sclerosis) and EIMS (Epidemiological Investigation of Multiple Sclerosis)^63^.

**Di Florio A, McQuillin A, McIntosh A, Breen G | Not published | [PGC Psychchip] | ukwa1 (n=2527)**

McQuillin A: A subset of the UCL control subjects (n=814) were recruited from London branches of the National Blood Service, from local NHS family doctor clinics and from university student volunteers. All control subjects were interviewed with the SADS-L to exclude all psychiatric disorders. All volunteers read an information sheet approved by the Metropolitan Medical Research Ethics Committee who also approved the project for all NHS hospitals. Written informed consent was obtained from each volunteer. A subset (n=448) of the control subjects were random UK blood donors obtained from the ECACC DNA Panels (<https://www.phe-culturecollections.org.uk/products/dna/hrcdna/hrcdna.jsp>).

McIntosh AM: Cases with bipolar disorder were recruited from the clinical case loads of treating psychiatrists from Edinburgh and across the central belt of Scotland. Controls were identified from non-genetic family members and from the extended networks of the participants themselves. All participants were of European ancestry and diagnosis was confirmed using an established battery developed for ICCCBD.

Breen G: Controls were drawn from blood donors to the UK Motor Neuron Disease Association DNA Biobank.^64^

**Gatt JM, Williams LM, Bryant R, Fullerton JM, Schofield PR | PMID: 32785990| [PGC Psychchip] | unsw1 (n= 641)**

This sample is drawn from the TWIN-E study, an ongoing longitudinal prospective study of 1,660 individuals (aged 18–62 years) sourced from Twins Research Australia. The baseline study was originally conducted at the University of Sydney, under approval from the Human Research Ethics Committee (03-2009/11430). Participants were community dwelling, healthy, same-sex, adult twin-pairs with English as their primary language. Participants did not complete formal psychiatric assessments, but provided questionnaire, neurocognitive, electrophysiological, neuroimaging and saliva samples for genetic material^65^. The genotyped sample comprised 1,333 DNA samples comprising ~710 unrelated individuals (pi_hat<0.2) plus co-twins, as previously described^66^.

**Mathews CA| Not published | [PGC Psychchip] | matt1 (n=20)**

Control samples were ascertained as part of ongoing genetic and neurophysiological studies of hoarding, obsessive compulsive and tic disorders. Controls reported no current or lifetime history of mania or hypomania at the time of ascertainment. Sixty-two of the 104 controls were screened for psychiatric illness using the Structured Clinical Interview for DSM-IV TR diagnoses and diagnoses of bipolar disorder, lifetime or current, were ruled out through a best estimate consensus diagnosis. Other psychiatric diagnoses were not excluded. The remaining 42 participants were not formally screened, but reported no lifetime or current history of bipolar disorder, obsessive compulsive, hoarding, or tic disorders. Samples were genotyped on the Psychchip array. Ethical approvals were obtained from the University of Florida Human Subjects Review Board.

**Medland SE, Martin NG | Not published | [PGC Psychchip] | usadd-mart1 (n=395)**

Control samples were ascertained as part of a study on ADHD traits and inattention more broadly. Controls were screened for ADHD using the SWAN questionnaire^67^ and did not meet criteria for ADHD at the time of recruitment. Samples were genotyped on the Psychchip array. Ethical approvals were obtained from the QIMR Berghofer Medical Research Institute Human Research Ethics Committee.

**Waldman I | Not published | [PGC Psychchip] | wald1 (n=55), wald2 (n=110)**

Control samples were ascertained as part of an ongoing genetic study of ADHD and other Externalizing disorders (I.e., Oppositional Defiant Disorder and Conduct Disorder). Controls reported no current diagnoses of Externalizing or Internalizing disorders at the time of ascertainment. Controls were assessed for psychiatric conditions using the Emory Diagnostic Rating Scale (EDRS)^68^, a questionnaire that assessed parent ratings of symptoms of common DSM-IV Externalizing and Internalizing disorders (e.g., Major Depressive Disorder and various anxiety disorders). Samples were genotyped on the Psychchip array. Ethical approvals were obtained from the Emory University and University of Arizona Human Subjects Review Boards.

### ISGC COHORT GENOTYPING, QC, IMPUTATION AND ANALYSIS

**Psychiatric Genomics Consortium Major Depressive Disorder**

Cohorts were genotyped following their local protocols, after which standardized quality control and imputation and analyses were performed centrally using RICOPILI (Rapid Imputation for COnsortias PIpeLIne), for each cohort separately^69^. These procedures have been described in detail previously^2^. Briefly, the quality control parameters for retaining SNPs and subjects were: SNP missingness < 0.05 (before sample removal), subject missingness < 0.02, autosomal heterozygosity deviation (F_het_ < 0.2), SNP missingness < 0.02 (after sample removal), difference in SNP missingness between psychiatric cases and healthy controls < 0.02 and SNP Hardy-Weinberg equilibrium (*P* > 10^−10^ in psychiatric cases, *P* > 10^−6^ in healthy controls). Genotype imputation was performed using the pre-phasing/ imputation stepwise approach implemented in IMPUTE2/ SHAPEIT (chunk size of 3 Mb and default parameters) to the 1000 Genomes Project reference panel^70^,^71^,^72^. Relatedness between subjects was calculated using identity by descent and one of each pair of related individuals (pi_hat > 0.2) was excluded. Relatedness with subjects in the PGC BIP and PGC SCZ samples was also calculated and one of each pair of relatives (pi_hat > 0.2) was excluded across all three of the samples. Overlapping individuals between PGC MDD and the UK Biobank sample were determined using genotype-based checksums ([https://personal.broadinstitute.org/sripke/share_links/zpXkV8INxUg9bayDpLToG4g58TMtjN_PGC_SCZ_w3.0718d.76](https://urldefense.proofpoint.com/v2/url?u=https-3A__personal.broadinstitute.org_sripke_share-5Flinks_zpXkV8INxUg9bayDpLToG4g58TMtjN-5FPGC-5FSCZ-5Fw3.0718d.76&d=DwMFaQ&c=shNJtf5dKgNcPZ6Yh64b-A&r=TmdNCZIwdRIVqqv7d3xHrbkQQi1zMoIzfjFfR7w4-Kk&m=YxSUDT6YJS4kZ7GPhOGc8ZUUxuuyYP_16GcVfXsOEBQ&s=G-3h0PoE6Tj3aYUojDDjmvs6FhUSi5t8MxM66rT76pI&e=)), and excluded from the PGC MDD study. One of the PGC MDD cohorts (BACCs) was excluded from the primary GWAS of SA due to overlapping controls with one of the PGC BIP cohorts (BOMA-Germany).

GWAS were performed using PLINK 1.9 by comparing imputed marker dosages under an additive logistic regression model between cases and controls in each of the 14 cohorts separately^73^. Principal components (PCs) generated using EIGENSTRAT were used as covariates in all GWAS as required, to control for population stratification^74^. SNPs were filtered from the GWAS summary statistics from each cohort using sample minor allele frequency (MAF) >= 1% and sample MAF corresponding to a minor allele count of 10 in cases or controls (whichever had smaller N), in order to control test statistic inflation at low MAFs from small cohorts. Meta-analyses were then performed across cohorts using an inverse variance-weighted fixed effects model in METAL, to obtain results for the primary GWAS of SA and the GWAS of SA within psychiatric diagnosis^75^.

**Psychiatric Genomics Consortium Bipolar Disorder**

Genotyping, QC imputation and analyses were conducted in the same manner as described for the PGC MDD sample and have been described in full previously^1,3^.

**Psychiatric Genomics Consortium Schizophrenia**

Genotyping, QC imputation and analyses were conducted in the same manner as described for the PGC MDD sample and have been described in full previously^1,4^. The Danish PGC SCZ cohort was excluded from the primary GWAS of SA, to ensure no overlap with the Danish iPSYCH cohort.

**Psychiatric Genomics Consortium Eating Disorders**

Genotyping has been described previously for these cohorts^5,6^. Quality control, principal components analysis to identify and remove ancestry outliers and generate covariates, and imputation to the 1000 Genomes Phase 3 reference panel were performed within PGC’s GWAS pipeline RICOPILI ^5,69^ as described in full previously^5^. The first 5 PCs were included as covariates and GWASs were performed within RICOPILI using imputed variant dosages and an additive model. Identical individuals between PGC ED cohorts and PGC MDD, BIP and SCZ cohorts were detected using genotype-based checksums ([https://personal.broadinstitute.org/sripke/share_links/zpXkV8INxUg9bayDpLToG4g58TMtjN_PGC_SCZ_w3.0718d.76](https://urldefense.proofpoint.com/v2/url?u=https-3A__personal.broadinstitute.org_sripke_share-5Flinks_zpXkV8INxUg9bayDpLToG4g58TMtjN-5FPGC-5FSCZ-5Fw3.0718d.76&d=DwMFaQ&c=shNJtf5dKgNcPZ6Yh64b-A&r=TmdNCZIwdRIVqqv7d3xHrbkQQi1zMoIzfjFfR7w4-Kk&m=YxSUDT6YJS4kZ7GPhOGc8ZUUxuuyYP_16GcVfXsOEBQ&s=G-3h0PoE6Tj3aYUojDDjmvs6FhUSi5t8MxM66rT76pI&e=)). The USA/Canada GCAN/WTCCC-3 cohort was excluded from the primary GWAS of SA due to overlap of controls with one of the PGC BIP cohorts.

**CONVERGE**

DNA sequencing, variant calling, and imputation have been previously described^7^. Briefly, sequencing reads were aligned to GRCh37.p5 with Stampy (c.10.17)^76^ using default parameters after filtering out reads of poor quality. Variant discovery and genotyping at all SNPs in the 1000 Genomes Phase 1 East Asian (ASN) ^77^ was performed using the GATK’s UnifiedGenotyper (version 2.7-2-g6bda569). Imputation was performed using BEAGLE (version 3.3.2)^78^. GWAS were performed using PLINK 1.9 by comparing imputed marker dosages under an additive logistic regression model between cases and controls. Based on prior studies, the first two principal components were included as covariates; these were derived from an eigen-decomposition of the genetic relatedness matrix. Variants were excluded from analysis if they had an INFO score <0.3, minor allele frequency <0.001, or HWE p<1e-7.

**Army STARRS**

Samples were genotyped using the Illumina OmniExpress + Exome array with additional custom content (N SNP = 967,537) or the Illumina PsychChip (N SNP = 571,054; 477,757 SNPs overlap with OmniExpress + Exome array). Relatedness testing was carried out with PLINK v1.90 and pairs of subjects with π of >0.2 were identified, randomly retaining one member of each related pair. We used a two-step pre-phasing/imputation approach for genotype imputation, with reference to the 1000 Genomes Project multi-ethnic panel (August 2012 phase 1 integrated release; 2,186 phased haplotypes with 40,318,245 variants). We removed SNPs that were not present in the 1000 Genomes Project reference panel, had non-matching alleles to 1000 Genome Project reference, or had ambiguous, unresolvable alleles (AT/GC SNPs with minor allele frequency [MAF] > 0.1). For the Illumina OmniExpress array 664,457 SNPs and for the Illumina PsychChip 360,704 SNPs entered the imputation procedure. For quality control (QC) purposes we kept autosomal SNPs with missing rate < 0.05; kept samples with individual-wise missing rate < 0.02; and kept SNPs with missing rate < 0.02. After QC, we merged our study samples with HapMap3 samples. We kept SNPs with minor allele frequency (MAF) > 0.05 and LD pruned at R^2^ > 0.05. In order to avoid long range LD structure from interfering with the PCA analysis, we excluded SNPs in the MHC region (Chr 6:25-35Mb) and Chr 8 inversion (Chr 8:7-13Mb). We used PLINK v1.90 to conduct genome-wide association tests for each model on imputed SNP dosage with logistic regression adjusted for age, sex, and the top 10 within-population principal components (PCs).

**German Borderline Genomics Consortium**

Genotyping was performed using the Infinium PsychArray-24 Bead Chip as previously described^10^. Updated quality control and imputation were carried out using the RICOPILI GWAS pipeline^69^ for the present manuscript. Briefly, the exclusion criteria for SNPs and subjects in the first round of quality control were: genotyping call rate for given SNPs or individuals <98%, difference in SNP genotyping call rate between cases and controls >2%, deviation of autosomal heterozygosity from the mean (|Fhet|>0.2), or a deviation from Hardy-Weinberg equilibrium (p<1x10−10 in cases; p<1x10−6 in controls). Imputation was conducted using a publicly available reference panel consisting of 54,330 phased haplotypes with 36,678,882 variants from the haplotype reference consortium (EGAD00001002729) and the prephasing/imputation stepwise approach in EAGLE/MINIMAC3 (default parameters and a variable chunk size of 132 genomic chunks)^79,80^. Relatedness testing and population structure analysis were performed using a subset of 65,408 SNPs that fulﬁlled strict quality criteria after imputation (INFO >0.8, missingness <1%, minor allele frequency >0.05), and which had been subjected to LD pruning (r2>0.02) in the second round of quality control. In the case of cryptically related subjects with pi-hat >0.2, one member of each pair was removed at random following the preferential retention of cases over controls. Principal components (PCs) were estimated from the quality-controlled genotypic data, and phenotype association was tested using logistic regression. The effect of individual PCs on genome-wide test statistics was assessed using λ. The GWAS was performed using an additive logistic regression model including the PCs associated with Borderline Borderline Personality Disorder case-control status (1-4; 7) as covariates to test single-marker associations in PLINK 1.9.

**Grady Trauma Project (GTP)**

Genotyping for the Grady Trauma Project was performed using the Omni-Quad 1M Bead Chip. Quality control and imputation (1000 Genomes Phase 3-hg19) were performed by using the Psychiatric Genomics Consortium PTSD Workgroup guidelines^81^. Only individuals with African American ancestry based on SNPweights software17 were included in the models. Principal components for ancestry were calculated according to the PGC guidelines in each separate ancestry group^81^. For each model, GWAS was performed using an additive logistic regression adjusting for 5 ancestry principal components (PLINK 1.9).

**UK Biobank**

Genotypic data were available for 488,380 individuals and were imputed to the Haplotype Reference Consortium (HRC), UK10K and 1,000 Genomes Phase 3 reference panels using IMPUTE4 to identify ≈ 93M variants for 487,409 individuals^82^. Variants for analysis were limited to those with minor allele frequency >= 0.01, imputation INFO-score >= 0.4, and which were either genotyped or imputed to the HRC reference panel, leaving a total of 7794483 SNPs for analysis. Using the genotyped SNPs, individuals were removed if: recommended by the UK Biobank core analysis team for unusual levels of missingness or heterozygosity; SNP genotype call rate < 98%; related to another individual in the dataset (KING r < 0.044, equivalent to removing up to third-degree relatives inclusive); phenotypic and genotypic gender information was discordant (X-chromosome homozygosity (FX) < 0.9 for phenotypic males, FX > 0.5 for phenotypic females). Removal of relatives was performed using a greedy algorithm, which minimises exclusions (for example, by excluding the child in a mother-father-child trio). All analyses were limited to individuals of White Western European ancestry, as defined by 4-means clustering on the first two genetic principal components provided by the UK Biobank^82^. Principal component analysis was also performed on the European-only subset of the data using the software flashpca2^83^. The GWAS of SA within psychiatric diagnosis was performed using BGenie v.1.2^82^, covarying for 6 PCs, and factors capturing site of recruitment and genotyping batch. QC, imputation and analysis for the primary GWAS of SA followed similar procedures and has been described previously^84^.

**Taiwan MDD**

Genotyping for MDD cases was obtained using Affymetrix CHB Array with 642,832 markers, Affymetrix TWB Array with 642,545 markers, and Illumina Human Omni Express Exome Beadchips with 949,974 markers. Samples with a completion call rate below 95 % were repeatedly assayed on a new aliquot DNA. Imputation was conducted by Michigan Imputation Server (https://imputationserver.sph.umich.edu/index.html#!) using 1000G phase 3 v5 as a reference panel, Eagle v2.3 for phasing, and EAS population for QC. We imputed ~46 million variants for both MDD and based on 1000 genome data of the East Asian panel. Samples that did not meet the 95% threshold of call rate were removed. We also removed kinship-pairs and outliers in population stratification. Markers with call rate <95%, minor allele frequency <0.01, p-value of Hardy-Weinberg equilibrium <1E-6, or imputation INFO score <0.7 were excluded. GWAS were performed using PLINK 1.9 and adjusted for 5 ancestry principal components.

**Taiwan BIP**

Genotyping for BIP cases was obtained using Affymetrix CHB Array with 642,832 markers, Affymetrix 6.0 Human Omni Express with 730,525 markers, and Affymetrix TWB Array with 642,545 markers. Samples with a completion call rate below 95 % were repeatedly assayed on a new aliquot DNA. The imputation processes, QC criteria and GWAS analysis were all the same as those in Taiwan MDD.

**Taiwan SCZ**

Genotyping for SCZ cases was obtained using the Axiom Genome-wide CHB 1 Array Plate in TSLS participants ^85^. Samples with a completion call rate below 95 % were dropped from analysis. The imputation processes, QC criteria and GWAS analysis were all the same as those in Taiwan MDD and BIP.

**iPSYCH**

Genotyping, QC and imputation procedures for iPSYCH 2012 cohort were conducted in the same manner as described for the previous GWAS of SA^1,86,87^. Genotyping waves with less than 50 SA cases were removed from the analysis followed by removal of related individuals, duplicated samples, and restricting individuals to European population and Danish origin only. After the filtering of genotyping data 7,003 SA cases and 52,227 non-SA controls were identified. The GWAS analysis was adjusted for sex, the first 10 principal components of genetic ancestry and genotyping batch. Association analyses were performed and are reported only for variants for which P-value was calculated and for variants with MAF ≥ 1% or ≤ 99% in the control group.

**Janssen**

Clinical samples from NCT00334126, NCT00397033, and NCT00412373 were genotyped using Illumina Human1M-DuoV3, while samples from NCT00044681 were genotyped using HumanOmni5Exome-4v1. The rest of the samples were genotyped using PsychArray. Standard QC were applied. Genotype data were imputed using Impute2 against 1000 Genome reference panel (integrated_phase1_v3). The imputed genotype dosages were assessed for association in a logistic regression model, correcting for four principal components to account for population substructure.

**Genetic Investigation of Suicide and SA (GISS)**

SNP genotyping was done using the HumanOmni1-Quad_v1 chip (Illumina Inc.) at the SNP&SEQ Technology Platform facility (snpseq.medsci.uu.se), assaying ~1 million SNPs with each trio plated consecutively. For the raw data, 96.7% of SNPs had call rate >99%, >99.99% of calls were reproducible, >99.99% of family-wize calls had no mendelian errors, and duplicated individuals could be ruled out. SNPs were filtered to obtain call rates >= 95%, hardy weinberg equilibrium (HWE) exakt P = 10^-6^, minor allele frequency (MAF) = 0.01 and no mendelian errors, whereby 739,780 autosomal- and 17,501 X-chromosomal SNPs remained. Phased reference panels (1000 genomes, phase 1; filtered for 1.00<MAF<0.005), BEAGLE v.3.3.2 and utils were downloaded (faculty.washington.edu/browning) ^88^. SNPs were checked against the phased EUR individuals in the 1000 genomes reference-panel, for inconsistencies in SNP-strands, -positions, -names, MAFs, linkage disequilibrium (LD) and number of alleles, using the available check_strands python routines. 729,956 autosomal SNPs remained for imputation using ~9 million reference panel SNPs (1.00 > MAF > 0.005). The X-chromosome was not imputed. Phasing (nsamples=2) and imputation (nsamples=1) were executed separately, running one chromosome at a time in low-memory mode on a desktop PC. Only SNPs imputed with Beagle allelic R^2^ = 0.7 were retained. ~5.5% of SNPs had a rare frequency (MAF < 0.01). The net imputation SNP gain after accounting for LD with r^2^-threshold < 0.8 pruning and MAF > 0.01, was from 450,348 autosomal SNPs pre-imputation to 1,035,345 autosomal SNPs post-imputation, i.e. ~2.3 fold. Quantiles vs quantiles (QQ) plots showed that observed SNP P-values followed the uniform null (genomic inflation = 1.002), as previously described.^23^ For this analysis, the ~6.8 million post-imputation SNP data was converted into a case-control sample by use of --tucc command in plink v.1.07 (660 cases and 660 controls; each control is a non-SA pseudo-sib, matched to a case on all other features), followed by analysis with --assoc --ci 0.95 in plink v.1.9.

**Australian Genetics of Depression Study and QSkin**

Samples from the AGDS were genotyped on three different genotyping centers using the same array (GSAMD-24v1-0_20011747). Healthy controls (QSkin cohort) were also genotyped using the GSA array. Quality control procedures that follow were applied to both AGDS and QSKIN genotypes. A common set of high QC markers between the different genotyping batches was obtained prior to joint imputation. Marker exclusion criteria (prior to imputation) included: unknown or ambiguous map position and strand alignment in a BLAST search, missingness >5%, p(HWE test)< 10^-6), MAF<1%, GenTrain score <0.6. The Michigan imputation server was used to impute the genotypes using the HRCr1.1 as a reference panel. Individuals were excluded based on a high missingness (missing rate > 3%), inconsistent (and unresolvable) sex, or if deemed ancestry outliers from the European population (6 sd deviations from the first two genetic principal components). The GWAS was done employing a logistic regression using PLINK 1.9 and imputed dosage genotypes while correcting for the genotyping center and the first five ancestry principal components as covariates.

**Yale-Penn (European and African American cohorts)**

We included two different sets of identically ascertained subjects who were genotyped on different platforms. Yale-Penn 1, collected earlier, was genotyped using the HumanOmni-Quad v1.0 array (Illumina) containing 988,306 autosomal single nucleotide polymorphism (SNP) markers. Yale-Penn 2 was genotyped on the HumanCore Exome array (Illumina) containing 550,601 SNPs. Individuals and SNPs with call rates <98% were removed. Only imputed SNPs with an accuracy greater than 0.8 were retained, and all SNPs with a Hardy-Weinberg equilibrium P < 10−5 were removed. SNPs with MAF < 1% were removed. Subject population was defined based on two ancestry groups, European American (EA) and African American (AA), assigned using 1000 genome phase 3 for EUR and AFR as reference.

**Columbia University**

QC procedures were performed using PLINK. Markers were retained if they had a minor allele frequency (MAF) of 1% or more, a call rate 95%, and no significant departures from Hardy–Weinberg Equilibrium (HWE P>0.0001). Samples with ambiguous sex, genotyping call completeness <95%, and duplicated individuals were excluded. Multidimensional Scaling Analysis (MDS) in PLINK, and comparison to HapMap Phase 3 populations were used to exclude individuals of non-European ancestry. The majority of samples from all three sites were found to be superimposed on the CEU population, outliers more than 3 trimmed standard deviations away from the sample average (using 5% trimming) were deleted,and, after rerunning the MDS analysis, the first five components were retained. For the purposes of the present analysis, genotypes were imputed using the following Imputation reference panel**:** 1000 Genomes Phase 3 (Version 5), and genome build**:** 37. Logistic regression was run on the imputed data, using the “dosage” statement in PLINK, adjusted for the following covariates: sex, age and first 5 MDS components. MDS components were calculated on the unimputed data. In the logistic regression, cases were all subjects with suicidal behavior, regardless of whether they were cases of SA or suicide, while controls were live subjects or sudden death victims. without a history of attempt but with or without a psychiatric diagnosis.

**Japan**

The details of genotyping, QC and imputation are reported in our previous GWAS paper^35^. Briefly, samples were genotyped using Illumina HumanOmniExpress and HumanOmniExpressExome BeadChips for the 1st and 2nd set of samples ascertained, respectively. We performed QC using PLINK 1.9. Firstly, for each set, we excluded SNPs with a call rate < 0.98 and MAF < 0.01, and those with p < 1.0 × 10^−6^ for HWE in controls. Related individuals were excluded (PI_HAT ≥ 0.175). We performed PCA, and confirmed all of the above subjects were in the Japanese cluster. After estimating haplotypes using SHAPEIT2 (v2.r778), we performed genotype imputation by Minimac3 (1.0.13) using ALL samples in the 1000 Genomes Project phase 3v5 as a reference. In order to finalize the summary statistics of imputed data which consist of 746 suicide decedents and 14,049 controls, we combined the summary statistics of imputed variants of the 1st and 2nd control sets as implemented in Rvtests software, performing meta-analysis with METAL software using a fixed effects model with inverse-variance weighted approach, with adjustment for 10 PCs.

**University of Utah**

Suicide cases were genotyped using Illumina Infinium PsychArray platform measuring 593,260 single nucleotide polymorphisms (SNPs). Genotypes were subsequently imputed in all cases and controls jointly. Cases resulted from population-based ascertainment and cryptic relatedness was modeled via the derivation of genomic relatedness matrices. Genotyping quality control was performed using SNP clustering in [Illumina Genome Studio](https://www.illumina.com/techniques/microarrays/array-data-analysis-experimentaldesign/genomestudio.html). SNPs were retained if the GenTrain score was > 0.5 and the Cluster separation score was > 0.4. SNPs were converted to HG19 plus strand, and SNPs with >5% missing genotypes were removed. Samples with a call rate < 95% were removed. The ancestry PCA was performed using RaMWAS^89^. Approximately 20% of the population-based suicide cases had a significant degree of non-Northwestern European ancestry (chiefly of admixed ancestry) and were excluded from GWAS analyses. To exclude ancestrally heterogeneous samples, the top principal components (defined as those components which accounted for > 0.1% of the genotype variance, *n*_pc_ = 4) were used to establish PC centroid limits centered around 1000 Genomes CEU data, such that 99% of the CEU data fell within the limits. Only suicide and control samples also falling within these limits were considered ancestrally homogenous and thus were included in the GWAS. PCA was performed on control, suicide, and 1000 Genomes cohorts after LD pruning at a 0.2 threshold. European ancestry cases and controls were well-matched to 1000 Genomes CEPH. The Haplotype Reference Consortium is comprised in part by UK controls used in the GWAS, so we imputed genotypes based on the 1000 Genomes reference panel using minimac3^79^ and Eagle^79,90^ . SNPs with ambiguous strand orientation, >5% missing calls, or Hardy-Weinberg equilibrium p < 0.001 were excluded. SNPs with minor allele frequency below 0.01 or imputation R^2^ < 0.5 were also excluded. Genomic data were handled using PLINK 1.9^73^. Final GWAS analysis was performed on 7,519,308 variants passing quality control. GWAS were performed by comparing imputed marker dosages under an additive logistic regression model between cases and controls.

### ACKNOWLEDGEMENTS

**General Acknowledgements**

Only a brief list of acknowledgements was possible in the main manuscript. The full list of acknowledgements is provided here. We thank the participants who donated their time, life experiences and DNA to this research, and the clinical and scientific teams that worked with them.

This work was supported by the National Institute of Mental Health (R01MH123619, R01MH123489, R01MH099134, K01MH093731, R01MH116269, R01MH121455) and the Department of Veterans Affairs. Research reported in this publication was also supported by NIGMS of the National Institutes of Health under award number T32GM007347. This work was also supported by the Brain & Behavior Research Foundation (NARSAD Young Investigator Awards No. 29551 to N.M., No. 28686 to A.S., and No. 28132 to E.D.), the Simons Foundation, the Huntsman Mental Health Institute, the American Foundation for Suicide Prevention, and the Clark Tanner Research Foundation. This work was also supported by research funding from Janssen Research & Development, LLC to University of Utah. Several statistical analyses were carried out on the Mount Sinai high performance computing cluster ([http://hpc.mssm.edu](http://hpc.mssm.edu/)), which is supported by the Office of Research Infrastructure of the National Institutes of Health (Grant Nos. S10OD018522 and S10OD026880). This work was also conducted in part using the resources of the Advanced Computing Center for Research and Education at Vanderbilt University, Nashville, TN and the Computational Core for Translational Sciences at the University of Utah, Salt Lake City, UT.

**Cohort Acknowledgements**

**Psychiatric Genomics Consortium**

We are deeply indebted to the investigators who comprise the PGC. The PGC has received major funding from the US National Institute of Mental Health (PGC3: U01 MH109528; PGC2: U01 MH094421; PGC1: U01 MH085520).

Some data used in this study were obtained from dbGaP. Funding support for the Genome-Wide Association of Schizophrenia Study was provided by the National Institute of Mental Health (R01 MH67257, R01 MH59588, R01 MH59571, R01 MH59565, R01 MH59587, R01 MH60870, R01 MH59566, R01 MH59586, R01 MH61675, R01 MH60879, R01 MH81800, U01 MH46276, U01 MH46289 U01 MH46318, U01 MH79469, and U01 MH79470) and the genotyping of samples was provided through the Genetic Association Information Network (GAIN). The datasets used for the analyses described in this manuscript were obtained from the database of Genotypes and Phenotypes (dbGaP) found at [http://www.ncbi.nlm.nih.gov/gap](https://urldefense.proofpoint.com/v2/url?u=http-3A__www.ncbi.nlm.nih.gov_gap&d=DwQGaQ&c=shNJtf5dKgNcPZ6Yh64b-A&r=TmdNCZIwdRIVqqv7d3xHrbkQQi1zMoIzfjFfR7w4-Kk&m=QTImDg_2NzYPEatBs2VtWov-B1B7tTKik39eZIg44C4&s=ANpNOVa2wKSt6huWGFLj8fMPhjC5GNiFuDA0tDPmnAU&e=) through dbGaP accession number phs000021.v3.p2. Samples and associated phenotype data for the Genome-Wide Association of Schizophrenia Study were provided by the Molecular Genetics of Schizophrenia Collaboration (PI: Pablo V. Gejman, Evanston Northwestern Healthcare (ENH) and Northwestern University, Evanston, IL, USA).” Funding support for the Whole Genome Association Study of Bipolar Disorder was provided by the National Institute of Mental Health (NIMH) and the genotyping of samples was provided through the Genetic Association Information Network (GAIN). The datasets used for the analyses described in this manuscript were obtained from the database of Genotypes and Phenotypes (dbGaP) found at http://www.ncbi.nlm.nih.gov/gap through dbGaP accession number [phs000017.v3.p1](https://www.ncbi.nlm.nih.gov/projects/gap/cgi-bin/study.cgi?study_id=phs000017.v3.p1). Samples and associated phenotype data for the Collaborative Genomic Study of Bipolar Disorder were provided by the NIMH Genetics Initiative for Bipolar Disorder. Data and biomaterials were collected in four projects that participated in NIMH Bipolar Disorder Genetics Initiative. From 1991-98, the Principal Investigators and Co-Investigators were: Indiana University, Indianapolis, IN, U01 MH46282, John Nurnberger, M.D., Ph.D., Marvin Miller, M.D., and Elizabeth Bowman, M.D.; Washington University, St. Louis, MO, U01 MH46280, Theodore Reich, M.D., Allison Goate, Ph.D., and John Rice, Ph.D.; Johns Hopkins University, Baltimore, MD U01 MH46274, J. Raymond DePaulo, Jr., M.D., Sylvia Simpson, M.D., MPH, and Colin Stine, Ph.D.; NIMH Intramural Research Program, Clinical Neurogenetics Branch, Bethesda, MD, Elliot Gershon, M.D., Diane Kazuba, B.A., and Elizabeth Maxwell, M.S.W. Data and biomaterials were collected as part of ten projects that participated in the NIMH Bipolar Disorder Genetics Initiative. From 1999-03, the Principal Investigators and Co-Investigators were: Indiana University, Indianapolis, IN, R01 MH59545, John Nurnberger, M.D., Ph.D., Marvin J. Miller, M.D., Elizabeth S. Bowman, M.D., N. Leela Rau, M.D., P. Ryan Moe, M.D., Nalini Samavedy, M.D., Rif El-Mallakh, M.D. (at University of Louisville), Husseini Manji, M.D. (at Wayne State University), Debra A. Glitz, M.D. (at Wayne State University), Eric T. Meyer, M.S., Carrie Smiley, R.N., Tatiana Foroud, Ph.D., Leah Flury, M.S., Danielle M. Dick, Ph.D., Howard Edenberg, Ph.D.; Washington University, St. Louis, MO, R01 MH059534, John Rice, Ph.D, Theodore Reich, M.D., Allison Goate, Ph.D., Laura Bierut, M.D. ; Johns Hopkins University, Baltimore, MD, R01 MH59533, Melvin McInnis M.D. , J. Raymond DePaulo, Jr., M.D., Dean F. MacKinnon, M.D., Francis M. Mondimore, M.D., James B. Potash, M.D., Peter P. Zandi, Ph.D, Dimitrios Avramopoulos, and Jennifer Payne; University of Pennsylvania, PA, R01 MH59553, Wade Berrettini M.D.,Ph.D. ; University of California at Irvine, CA, R01 MH60068, William Byerley M.D., and Mark Vawter M.D. ; University of Iowa, IA, R01 MH059548, William Coryell M.D. , and Raymond Crowe M.D. ; University of Chicago, IL, R01 MH59535, Elliot Gershon, M.D., Judith Badner Ph.D. , Francis McMahon M.D. , Chunyu Liu Ph.D., Alan Sanders M.D., Maria Caserta, Steven Dinwiddie M.D., Tu Nguyen, Donna Harakal; University of California at San Diego, CA, R01 MH59567, John Kelsoe, M.D., Rebecca McKinney, B.A.; Rush University, IL, R01 MH059556, William Scheftner M.D. , Howard M. Kravitz, D.O., M.P.H., Diana Marta, B.S., Annette Vaughn-Brown, MSN, RN, and Laurie Bederow, MA; NIMH Intramural Research Program, Bethesda, MD, 1Z01MH002810-01, Francis J. McMahon, M.D., Layla Kassem, PsyD, Sevilla Detera-Wadleigh, Ph.D, Lisa Austin,Ph.D, Dennis L. Murphy, M.D.

**Psychiatric Genomics Consortium Major Depressive Disorder Cohorts**

GSK_MUNICH: We thank all participants in the GSK-Munich study. We thank numerous people at GSK and Max-Planck Institute, BKH Augsburg and Klinikum Ingolstadt in Germany who contributed to this project. JANSSEN: Funded by Janssen Research & Development, LLC. We are grateful to the study volunteers for participating in the research studies and to the clinicians and support staff for enabling patient recruitment and blood sample collection. We thank the staff in the former Neuroscience Biomarkers of Janssen Research & Development for laboratory and operational support (e.g., biobanking, processing, plating, and sample de-identification), and to the staff at Illumina for genotyping Janssen DNA samples. MARS: This work was funded by the Max Planck Society, by the Max Planck Excellence Foundation, and by a grant from the German Federal Ministry for Education and Research (BMBF) in the National Genome Research Network framework (NGFN2 and NGFN-Plus, FKZ 01GS0481), and by the BMBF Program FKZ 01ES0811. We acknowledge all study participants. We thank numerous people at Max-Planck Institute, and all study sites in Germany and Switzerland who contributed to this project. Controls were from the Dortmund Health Study which was supported by the German Migraine & Headache Society, and by unrestricted grants to the University of Münster from Almirall, Astra Zeneca, Berlin Chemie, Boehringer, Boots Health Care, Glaxo-Smith-Kline, Janssen Cilag, McNeil Pharma, MSD Sharp & Dohme, and Pfizer. Blood collection was funded by the Institute of Epidemiology and Social Medicine, University of Münster. Genotyping was supported by the German Ministry of Research and Education (BMBF grant 01ER0816). PsyColaus: The CoLaus|PsyCoLaus study was and is supported by research grants from GlaxoSmithKline, the Faculty of Biology and Medicine of Lausanne, and the Swiss National Science Foundation (grants 3200B0-105993, 3200B0-118308,33CSCO-122661, 33CS30-139468, 33CS30-148401, and 33CS30_177535). RADIANT: This report represents independent research funded by the National Institute for Health Research (NIHR) Biomedical Research Centre at South London and Maudsley NHS Foundation Trust, and King’s College London. The views expressed are those of the authors and not necessarily those of the NHS, the NIHR, or the Department of Health. SHIP-LEGEND/TREND: SHIP is part of the Community Medicine Research net of the University of Greifswald which is funded by the Federal Ministry of Education and Research (grants 01ZZ9603, 01ZZ0103, and 01ZZ0403), the Ministry of Cultural Affairs, and the Social Ministry of the Federal State of Mecklenburg-West Pomerania. Genotyping in SHIP was funded by Siemens Healthineers and the Federal State of Mecklenburg-West Pomerania. Genotyping in SHIP-TREND-0 was supported by the Federal Ministry of Education and Research (grant 03ZIK012). STAR*D: The authors appreciate the efforts of the STAR*D investigator team for acquiring, compiling, and sharing the STAR*D clinical data set. QIMR: We thank the participants and their families for their willing participation in our studies. MR received support from the Australian National Health and Medical Research Council (NHMRC) Centre for Research Excellence on Suicide Prevention (CRESP) [GNT1042580]. SEM was supported by fellowships from the NHMRC APP1172917 and APP1103623. This work was supported by NHMRC grants APP1086683 and APP1138514.

AMM is supported by the Wellcome Trust (104036/Z/14/Z, 216767/Z/19/Z), UKRI MRC (MC_PC_17209, MR/S035818/1) and the European Union’s Horizon 2020 research and innovation programme under grant agreement No 847776.

The following table lists the funding that supported the primary studies analyzed.

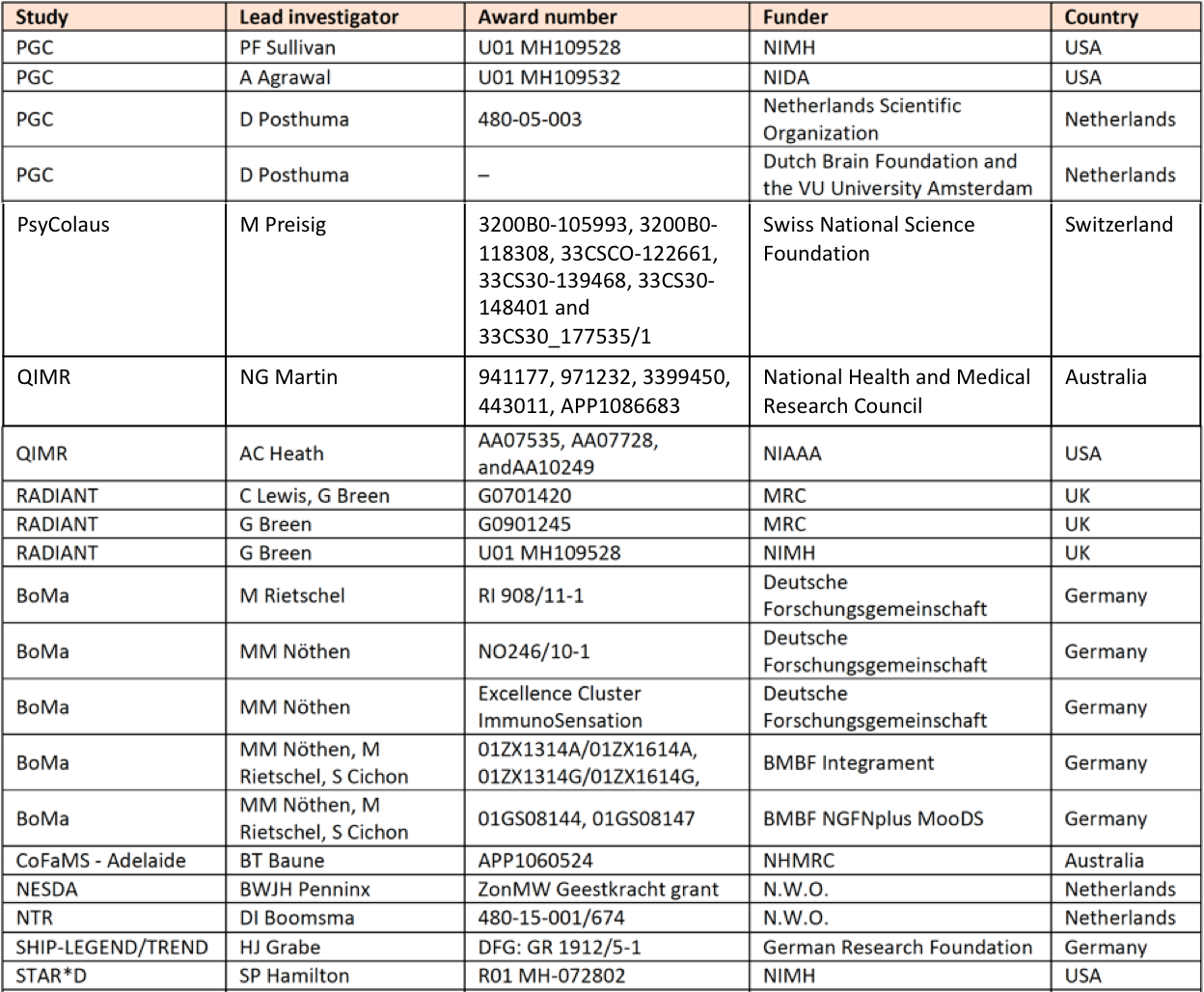

**Psychiatric Genomics Consortium Bipolar Disorder Cohorts**

BACCS: This work was supported in part by the NIHR Maudsley Biomedical Research Centre (‘BRC’) hosted at King’s College London and South London and Maudsley NHS Foundation Trust, and funded by the National Institute for Health Research under its Biomedical Research Centres funding initiative. The views expressed are those of the authors and not necessarily those of the BRC, the NHS, the NIHR or the Department of Health or King’s College London. We gratefully acknowledge capital equipment funding from the Maudsley Charity (Grant Reference 980) and Guy’s and St Thomas’s Charity (Grant Reference STR130505). Work on the Toronto (Centre for Addiction & Mental Health) cohort was supported in part by an operating grant from the Canadian Institutes of Health Research, MOP-172013.

BOMA-Germany I, BOMA-Germany II, BOMA-Germany III, PsyCourse: This work was supported by the German Ministry for Education and Research (BMBF) through the Integrated Network IntegraMent (Integrated Understanding of Causes and Mechanisms in Mental Disorders), under the auspices of the e:Med program (grant 01ZX1314A/01ZX1614A to MMN and SC, grant 01ZX1314G/01ZX1614G to MR, grant 01ZX1314K to TGS). This work was supported by the German Ministry for Education and Research (BMBF) grants NGFNplus MooDS (Systematic Investigation of the Molecular Causes of Major Mood Disorders and Schizophrenia; grant 01GS08144 to MMN and SC, grant 01GS08147 to MR). This work was also supported by the Deutsche Forschungsgemeinschaft (DFG), grants NO246/10-1 and NO 246/10-2 to MMN (FOR 2107), grant RI 908/11-2 to MR (FOR 2107), grant WI 3439/3-2 to SHW, grants SCHU 1603/4-1, SCHU 1603/5-1 (KFO 241) and SCHU 1603/7-1 (PsyCourse) to TGS. This work was also supported by ERA-NET NEURON “EMBED”, BMBF (Federal Ministry of Education and Research) grant 01EW1904 to MR. This work was supported by the Swiss National Science Foundation (SNSF, grant 156791 to SC). MMN is supported through the Excellence Cluster ImmunoSensation. TGS is supported by an unrestricted grant from the Dr. Lisa-Oehler Foundation. MH was supported by the Deutsche Forschungsgemeinschaft.

France: This research was supported by Foundation FondaMental, Créteil, France and by the Investissements d’Avenir Programs managed by the ANR under references ANR-11-IDEX-0004-02 and ANR-10-COHO-10-01.

Halifax: Halifax data were obtained with support from the Canadian Institutes of Health Research (grant # 166098) and from Genome Atlantic

Michigan (NIMH/Pritzker Neuropsychiatric Disorders Research Consortium): We thank the participants who donated their time and DNA to make this study possible. We thank members of the NIMH Human Genetics Initiative and the University of Michigan Prechter Bipolar DNA Repository for generously providing phenotype data and DNA samples. Many of the authors are members of the Pritzker Neuropsychiatric Disorders Research Consortium which is supported by the Pritzker Neuropsychiatric Disorders Research Fund L.L.C. A shared intellectual property agreement exists between this philanthropic fund and the University of Michigan, Stanford University, the Weill Medical College of Cornell University, HudsonAlpha Institute of Biotechnology, the Universities of California at Davis, and at Irvine, to encourage the development of appropriate findings for research and clinical applications.

TOP: The TOP Study was supported by the Research Council of Norway (#213837, #217776, #223273), South-East Norway Health Authority (#2015-078, #2017-112, #2019-108) and K.G. Jebsen Stiftelsen and a research grant from Mrs. Throne-Holst.

The following table lists the funding that supported the primary studies analyzed.

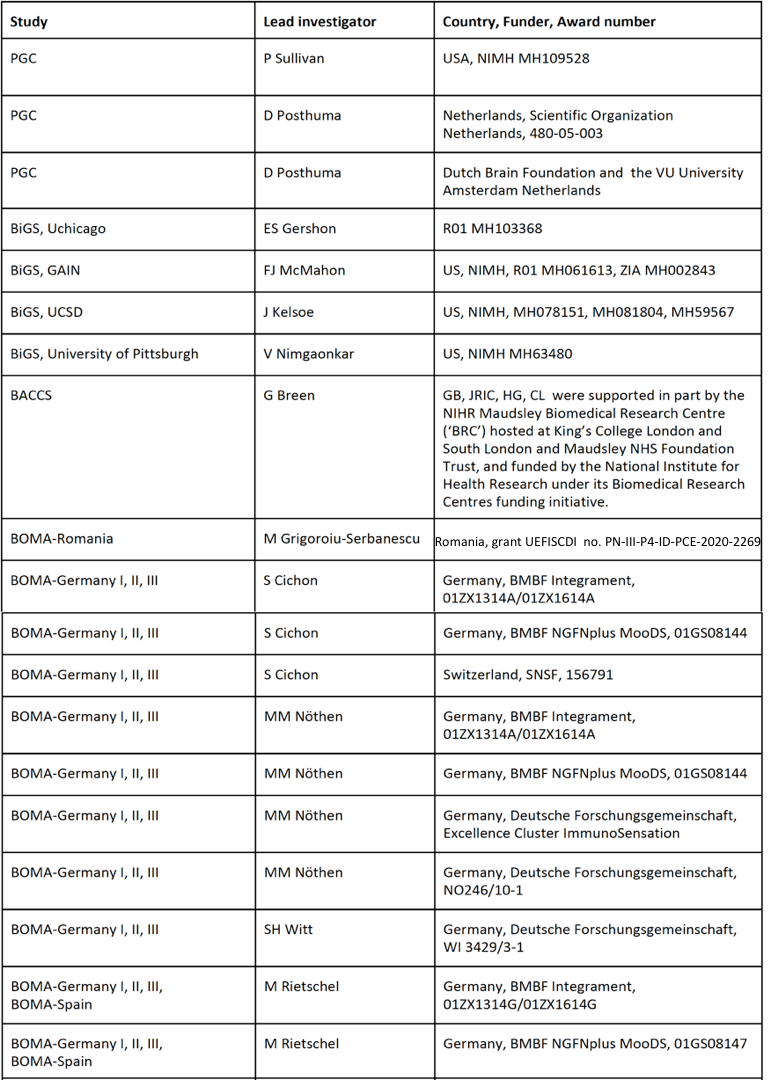

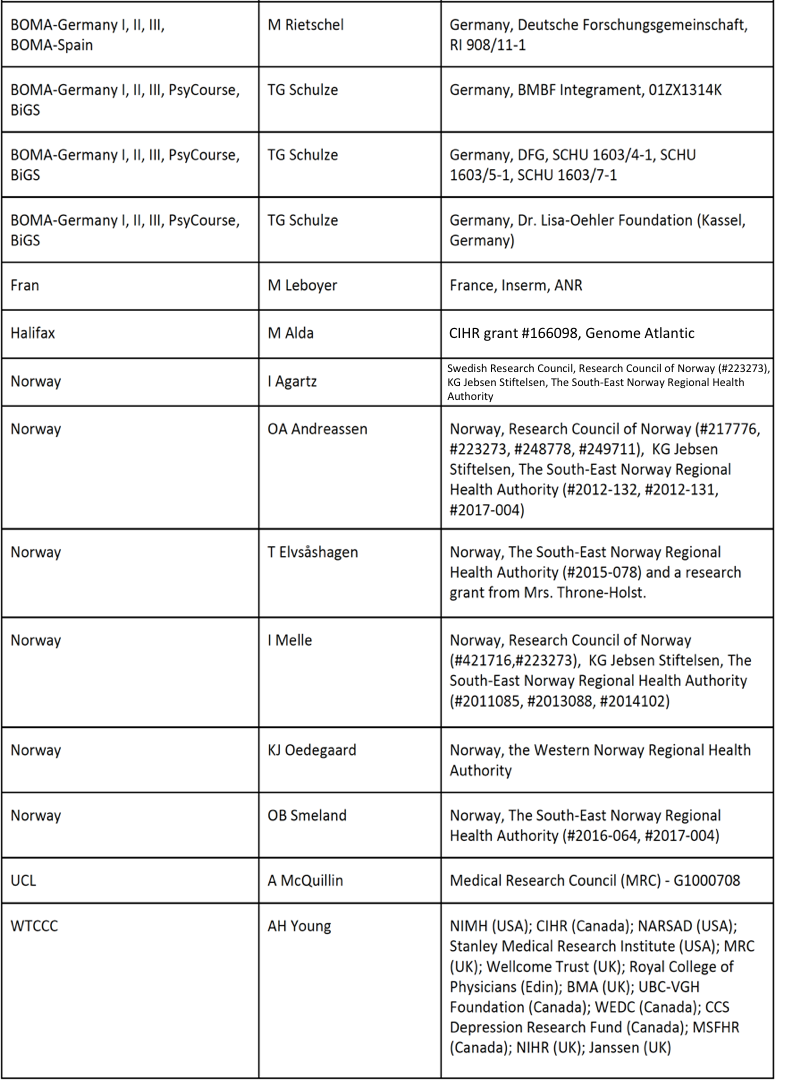

**Psychiatric Genomics Consortium Schizophrenia Cohorts**

Portugal: CNP and MTP are or have been supported by grants from the NIMH (MH085548, MH085542, MH071681, MH061884, MH58693, and MH52618) and the NCRR (RR026075). CNP, MTP, and AHF are or have been supported by grants from the Department of Veterans Affairs Merit Review Program

Bulgarian Trio sample: Work in Cardiff was supported by MRC Centre (G0800509) and MRC Programme (G0801418) Grants. The recruitment of families in Bulgaria was funded by the Janssen Research Foundation, Beerse, Belgium. We are grateful to the study volunteers for participating in the Janssen research studies and to the clinicians and support staff for enabling patient recruitment and blood sample collection. Informed consent was obtained from all participants or their parents or guardians.

Dutch sample: High-Density Genome-Wide Association Study Of Schizophrenia In Large Dutch Sample (R01 MH078075 NIH/National Institute Of Mental Health PI: Roel A. Ophoff).

Denmark: The Danish Aarhus study was supported by grants from The Lundbeck Foundation, The Danish Strategic Research Council, Aarhus University, and The Stanley Research Foundation.

TOP: The TOP Study was supported by the Research Council of Norway (#213837, #217776, #223273), South-East Norway Health Authority (#2015-078, #2017-112, #2019-108) and K.G. Jebsen Stiftelsen.

Bonn/Mannheim: The Bonn/Mannheim sample was genotyped within a study that was supported by the German Federal Ministry of Education and Research (BMBF) through the Integrated Genome Research Network (IG) MooDS (Systematic Investigation of the Molecular Causes of Major Mood Disorders and Schizophrenia; grant 01GS08144 to M.M.N. and S.C., grant 01GS08147 to M.R.), under the auspices of the National Genome Research Network plus (NGFNplus), and through the Integrated Network IntegraMent (Integrated Understanding of Causes and Mechanisms in Mental Disorders), under the auspices of the e:Med Programme.(GSK control sample; Müller-Myhsok). This work has been funded by the Bavarian Ministry of Commerce and by the Federal Ministry of Education and Research in the framework of the National Genome Research Network, Förderkennzeichen 01GS0481 and the Bavarian Ministry of Commerce. M.M.N. is a member of the DFG-funded Excellence-Cluster ImmunoSensation. M.M.N. also received support from the Alfried Krupp von Bohlen und Halbach-Stiftung.

Molecular Genetics of Schizophrenia: The collection was established as part of the Wellcome Trust Case- Control Consortium. We thank the study participants, and the research staff at the study sites. This study was supported by NIMH grant R01MH062276 (to DF Levinson, C Laurent, M Owen and D Wildenauer), grant R01MH068922 (to PV Gejman), grant R01MH068921 (to AE Pulver) and grant R01MH068881 (to B Riley). The authors are grateful to the many family members who participated in the studies that recruited these samples, to the many clinicians who assisted in their recruitment. In addition to the support acknowledged for the Multicenter Genetics Studies of Schizophrenia and Molecular Genetics of Schizophrenia studies, Dr. DF Levinson received additional support from the Walter E. Nichols, M.D., Professorship in the School of Medicine, the Eleanor Nichols Endowment, the Walter F. & Rachael L. Nichols Endowment and the William and Mary McIvor Endowment, Stanford University. This study was supported by NIH R01 grants (MH67257 to N.G.B., MH59588 to B.J.M., MH59571 to P.V.G., MH59565 to R.F., MH59587 to F.A., MH60870 to W.F.B., MH59566 to D.W.B., MH59586 to J.M.S., MH61675 to D.F.L., MH60879 to C.R.C., and MH81800 to P.V.G.), NIH U01 grants (MH46276 to C.R.C., MH46289 to C. Kaufmann, MH46318 to M.T. Tsuang, MH79469 to P.V.G., and MH79470 to D.F.L.), the Genetic Association Information Network (GAIN), and by The Paul Michael Donovan Charitable Foundation. Genotyping was carried out by the Center for Genotyping and Analysis at the Broad Institute of Harvard and MIT (S. Gabriel and D. B. Mirel), which is supported by grant U54 RR020278 from the National Center for Research Resources.

**Psychiatric Genomics Consortium Eating Disorder cohorts:** This work was funded by a grant from the WTCCC3 WT088827/Z/09 entitled “A genomewide association study of anorexia nervosa.”

Genetics of Anorexia Nervosa (GAN), National Institute of Mental Health: The data and collection of biomaterials for the GAN study have been supported by National Institutes of Health grants ([MH066122](https://www.ncbi.nlm.nih.gov/nuccore/MH066122), [MH066117](https://www.ncbi.nlm.nih.gov/nuccore/MH066117), [MH066145](https://www.ncbi.nlm.nih.gov/nuccore/MH066145), [MH066296](https://www.ncbi.nlm.nih.gov/nuccore/MH066296), [MH066147](https://www.ncbi.nlm.nih.gov/nuccore/MH066147), MH0662, [MH066193](https://www.ncbi.nlm.nih.gov/nuccore/MH066193), [MH066287](https://www.ncbi.nlm.nih.gov/nuccore/MH066287), [MH066288](https://www.ncbi.nlm.nih.gov/nuccore/MH066288), [MH066146](https://www.ncbi.nlm.nih.gov/nuccore/MH066146)).

Canada: The Ontario Mental Health Foundation (OMHF). The collection of the Toronto DNA samples was supported by a grant from the OMHF, awarded to Allan S. Kaplan and Robert D. Levitan (Polymorphism in Serotonin System Genes: Putative Role in Increased Eating Behaviour in Seasonal Affective Disorder and Bulimia Nervosa).

Spain: Department of Psychiatry University Hospital of Bellvitge-IDIBELL, Barcelona. Financial support was received from Fondo de Investigación Sanitaria-FIS (PI20/132) and AGAUR. CIBER Fisiopatología de la Obesidad y Nutrición (CIBERobn) is an initiative of ISCIII. We thank CERCA Program/Generalitat de Catalunya for institutional support. Additional support received from EU Grant Eat2beNice (H2020-SFS-2016-2; Ref 728018) and PRIME (PRIME 847879) and COST (Hyperchildnet, CA19115).

France: Institut National de la Santé et de la Recherche Médicale (INSERM), France. This French cohort was recruited with grants from EC Framework V ‘Factors in Healthy Eating’ (a consortium coordinated by Janet Treasure and David Collier, King's College London), and from INRA/INSERM (4M406D), and the participation of Audrey Versini's work was supported by grants from ‘Région Ile-de-France’. Cases were ascertained from Sainte-Anne Hospital (Paris) and Robert Debre Hospital (Paris).

PF/CHOP: The Price Foundation provided the funding for sample collection and all genome-wide genotyping was funded by an Institute Development Award to the Center for Applied Genomics from the Children’s Hospital of Philadelphia (CHOP).

**Grady Trauma Project:** This work was primarily supported by the National Institute of Mental Health (MH071537, PI-Ressler). Support also included Emory and Grady Memorial Hospital General Clinical Research Center, NIH National Centers for Research Resources (M01RR00039), and the Burroughs Wellcome Fund. We would like to thank Rebecca Hinrichs, Angelo Brown, and all of the Grady Trauma Project staff, multiple PIs, research assistants, and participants for their time and participation.

**Army STARRS:** Army STARRS was sponsored by the Department of the Army and funded under cooperative agreement number U01MH087981 (2009-2015; MPIs Ursano RJ and Stein MB) with the U.S. Department of Health and Human Services, National Institutes of Health, National Institute of Mental Health (NIH/NIMH). Subsequently, STARRS-LS was sponsored and funded by the Department of Defense (USUHS grant number HU0001-15-2-0004).

**Japan:** Sample collection, genotyping, and GWAS of DNA from suicide decedents and non-suicide controls in Japan and the current collaboration was supported, in part, by JSPS KAKENHI Grant Number 17H04249, 20KK0194, SENSHIN Medical Research Foundation, the Biobank Japan, and the Rotary Club of Osaka-Midosuji District 2660 Rotary International in Japan.

**Genetic Investigation of Suicide and SA (GISS).** Genetic Investigation of Suicide and SA (GISS) was funded by the Knut and Alice Wallenberg Foundation and National Centre for Suicide Research and Prevention of Mental Ill-Health (NASP), Karolinska Institutet, Stockholm, Sweden, with Danuta Wasserman as the Principal Investigator, Marcus Sokolowski chief geneticist, Vsevolod Rozanov responsible for material collection and Jerzy Wasserman chief of quality assurance of data collection and analyses

**COLUMBIA Suicide GWAS Study:** The study was funded by an NIMH grant to three sites including Columbia University with J. John Mann as the Principal Investigator for the coordinating site: R01 MH082041, entitled Suicidal Behavior in Mood Disorders: Genes and Intermediate Phenotypes [multi-site with Canada (PI: Gustavo Turecki) & Germany (PI: Dan Rujescu)] 2008-2013. Hanga Galfalvy was the lead statistician.

**CONVERGE**: This work was funded by the Wellcome Trust (WT090532/Z/09/Z, WT083573/Z/07/Z, WT089269/Z/09/Z) and the National Institutes of Health (MH100549 to K.S.K and AA027522 to A.C.E.). The China, Oxford, and VCU Experimental Research on Genetic Epidemiology (CONVERGE) consortium gratefully acknowledges the support of all partners in hospitals across China.

**AGDS and QSKIN:** Data collection for AGDS was possible thanks to a funding from the Australian National Health and Medical Research Council (NHMRC) to NGM (GNT1086683). We thank our colleagues Richard Parker, Simone Cross, Scott Gordon and Kerrie McAloney for their valuable work coordinating all the administrative, operational and technical aspects of the AGDS. AIC is supported by a UQ Research Training Scholarship from The University of Queensland (UQ). MER thanks the support of NHMRC and Australian Research Council (ARC), through a NHMRC-ARC Dementia Research Development Fellowship (GNT1102821). The QSkin Study is supported by NHMRC grants APP1073898, APP1063061 and APP1058522. DCW is supported by a Research Fellowship from the NHRMC (APP1155413).

**German Borderline Genomics Consortium:** This work was supported by the German Research Foundation to Christian Schmahl (KFO256 and GRK2350) and Stephanie Witt (GRK2350) and by the European Union to Marcella Rietschel (01EW1810).

**iPSYCH, Denmark:** The iPSYCH team was supported by grants from the Lundbeck Foundation (R102-A9118, R155-2014-1724 and R248-2017-2003), the EU H2020 Program (Grant No. 667302, “CoCA” to ADB), NIMH (1U01MH109514-01 to ADB) and the universities and university hospitals of Aarhus and Copenhagen. The Danish National Biobank resource was supported by the Novo Nordisk Foundation. High-performance computer capacity for handling and statistical analysis of iPSYCH data on the GenomeDK HPC facility was provided by the Center for Genomics and Personalized Medicine and the Centre for Integrative Sequencing, iSEQ, Aarhus University, Denmark (grant to ADB).

**Taiwan MDD, BPD, and SCZ Sample:** We thank all participants, interviewers, and psychiatrists who joined in this project. Sample collection, DNA extraction, and genotyping in this project was supported by Ministry of Science and Technology (105-2628-B-002-028-MY3 and 108-2314-B-002-136-MY3) and National Health Research Institutes (NHRI-EX108-10627NI) to Pohsiu Kuo.

**UK Biobank:** This study represents independent research funded partly by the National Institute for Health Research (NIHR) Maudsley Biomedical Research Centre at South London and Maudsley NHS Foundation Trust and King’s College London. High performance computing facilities were funded with capital equipment grants from the GSTT Charity (TR130505) and Maudsley Charity (980). This research used data derived from the UK Biobank Resource under application 18177 (Prof Lewis). The views expressed are those of the authors and not necessarily those of the NHS, the NIHR or the Department of Health and Social Care.

**Yale-Penn:** We wish to thank all of the research participants in this study. Adult subject recruitment and assessment were overseen at the Yale School of Medicine and the APT Foundation by James Poling, Ph.D., Aryeh Herman, Psy.D., and Dr Gelernter; at the University of Connecticut Health Center by Henry R. Kranzler, M.D.; at McLean Hospital by Roger Weiss, M.D.; at the Medical University of South Carolina by Kathleen Brady, M.D., Ph.D. and Raymond Anton, M.D.; and at the University of Pennsylvania initially by David Oslin, M.D. and then by Dr Kranzler. Some genotyping services were provided by the Center for Inherited Disease Research (CIDR) and the Yale University Center for Genome Analysis. CIDR is fully funded through a federal contract from the National Institutes of Health to The Johns Hopkins University (contract number N01-HG-65403). Ann Marie Lacobelle, M.S. and Christa Robinson, A.S. provided excellent technical assistance; the SSADDA interviewers, led by Yaira Nunez, devoted substantial time and effort to phenotype the study sample; and Richard Sherva, Ph.D., Ryan Koesterer, M.A., and John Farrell, Ph.D. at Boston University offered valuable assistance with data cleaning and management. This study was supported by grants from the National Institutes of Health (NIH) (RC2 DA028909, R01 DA12690, R01 DA12849, R01 DA18432, R01 AA11330, R01 AA017535) and the US Department of Veterans Affairs Medical Research Program. Daniel Levey was supported in part by a NARSAD Young Investigator Award.

**Janssen:** I am grateful to the study volunteers for participating in the research studies and to the clinicians and support staff for enabling patient recruitment and blood sample collection. We thank the staff in the former Pharmacogenomics/Neuroscience Biomarkers of Janssen Research & Development for laboratory and operational (including but not limited to sample banking, processing, plating, and clinical data/sample de-identification) support, and the staff at Illumina for genotyping Janssen DNA samples.

**University of Utah:** Collection, genotyping, and analysis of DNA from suicide deaths in the Utah Suicide Genetic Risk Study is supported by NIMH grants R01MH099134, R01MH122412, R01MH123489 (H Coon), R01MH123619 (A Docherty), The Utah Division of Substance Abuse and Mental Health (H Coon), a research contract with Janssen Research & Development, LLC (H Coon, Q Li), the Clark Tanner Foundation (A Shabalin/H Coon), and GCRC M01-RR025764 from the National Center for Research Resources. We gratefully acknowledge the staff of the Utah State Office of the Medical Examiner whose many hours of work have made this study possible.

**Psychiatric Genomics Consortium PsychChip controls**

**cogs1:** We would like to acknowledge all of the Consortium on the Genetics of Schizophrenia (COGS-I) Investigators: David L. Braff, M.D., Kristin S. Cadenhead, M.D., Monica E. Calkins, Ph.D., Dorcas J. Dobie, M.D., Robert Freedman, M.D., Michael F. Green, Ph.D., Tiffany A. Greenwood, Ph.D., Raquel E. Gur, M.D., Ph.D.,Ruben C. Gur, Ph.D., Gregory A. Light, Ph.D. , Keith H. Nuechterlein, Ph.D., Ann Olincy, M.D., Allen D. Radant, M.D. , Larry J. Seidman, Ph.D., Larry J. Siever, M.D., Jeremy M. Silverman, Ph.D., William S. Stone, Ph.D., Catherine A. Sugar, Ph.D. , Neal R. Swerdlow, M.D., Ph.D. , Debby W. Tsuang, M.D., Ming T. Tsuang, M.D., Ph.D., D.Sc., Bruce I. Turetsky, M.D. We also wish to thank all of the participants and support staff that made this study possible. This study was supported by grants from the National Institute of Mental Health (NIMH): R01-MH065571, R01-MH065588, R01-MH065562, R01-MH065707, R01-MH065554, R01- MH065578, and R01-MH065558.

**clait:** The Brazilian ADHD Porto Alegre Cohort received financial support from Conselho Nacional de Desenvolvimento Científico e Tecnológico (Grants 466722/2014-1, 424041/2016-2, 426905/2016-2, 431472/2018-1, 140853/2019-7). Also, this study was financed in part by Coordenação de Aperfeiçoamento de Pessoal de Nível Superior - Brasil (CAPES, - Finance Code 001), FIPE-HCPA 160600 and 01-321 and FAPERGS PPSUS- 19/2551-0001668-9 and 19/2551-0001731-6.

**germ1:** Collection of this sample was supported by the BMBF (BipoLife) and the European Commission (grant agreement no. 667302, “CoCA”), to A. Reif.

**iupui:** Funding for this project comes from Collaborative R01s MH68009, MH073151, and MH068006 and genotyping was supported by the Australian National Health and Medical Research Council through Project Grant 1066177 (to JMF & JIN). The authors gratefully acknowledge the Heinz C. Prechter Bipolar Research Fund at the University of Michigan.

**swed1:** Population-based healthy controls were collected as part of the SWEBIC case-control study of bipolar disorder (funded by the Stanley Center for Psychiatric Research and the Swedish Research Council 2018-02653) and two case-control studies of multiple sclerosis: GEMS (Genes and Environment in Multiple Sclerosis) and EIMS (Epidemiological Investigation of Multiple Sclerosis).

**neura:** Funding for the Bipolar High Risk project comes from the Australian National Health and Medical Research Council (NHMRC) through Program Grant 1037196 and Project Grants 1066177 and 1063960, and the Lansdowne Foundation. The IGP cohort collection was supported by NHMRC Project Grants 630471 and 108160. MJG was supported by NHMRC Fellowships 1061875 & 1121474. The CASSI study was supported by NHMRC Project Grant 568807, and the Schizophrenia Research Institute, utilizing infrastructure funding from NSW Ministry of Health and the Macquarie Group Foundation. CSW was also supported by NHMRC Fellowship 1021970. Some samples were sourced from the Australian Schizophrenia Research Bank, which is supported by the NHMRC of Australia, the Pratt Foundation, Ramsay Health Care and the Viertel Charitable Foundation. DNA was extracted by Genetic Repositories Australia, an Enabling Facility that was supported by NHMRC Enabling Grant 401184).

**nhsii:** Nurses Health Study II. NHSII PTSD Sub-Study was funded by National Institute on Mental Health awards RO1 MH093612, MH078928 to Karestan C Koenen. The NHSII cohort is funded in part by U01CA176726 and R01CA67262. We are grateful to all the participants in this study for their contributions.

**paris:** For the PsyDev Cohort, funding was provided by INSERM, ANR EPINEP (ANR 08-MNP-007), and Eranet Neuron AUSZ (ANR 2011-A00812-39), PHRC ICAAR (AOM07-118). The authors gratefully acknowledge all participants as well as Narjès Bendjemaa, Caroline Gaillard, and Guillaume Ciesco for their contribution on recruitment, and JP Bleton, M Boutroy (AFMK-APHP); J Signeyrole and F Marizy (Fondation EFOM) who welcomed us for the recruitment of volunteers. The genotyping was funded by U.S. National Institute of Health (U01MH096296)

**span1, span2:** CSM was a recipient of a Sara Borrell contract (CD15/00199) and a mobility grant (MV16/00039) from the Instituto de Salud Carlos III, Ministerio de Economía, Industria y Competitividad, Spain. MR was a recipient of a Miguel de Servet contract (CP09/00119 and CPII15/00023) and LV is a recipient of a pre-doctoral fellowship (FI18/00285) from the Instituto de Salud Carlos III, Ministerio de Economía, Industria y Competitividad, Spain. MSA is a recipient of a Juan de la Cierva Incorporación contract (IJC2018-035346-I) from the Ministry of Science, Innovation and Universities, Spain. This investigation was supported by Instituto de Salud Carlos III (PI16/01505,PI17/00289, PI18/01788, PI19/00721, P19/01224 and PI20/00041), and cofinanced by the European Regional Development Fund (ERDF), “la Marató de TV3” (092330/31), the Agència de Gestió d’Ajuts Universitaris i de Recerca-AGAUR, Generalitat de Catalunya (2014SGR1357 and 2017SGR1461) and the Pla estratègic de recerca i innovació en salut (PERIS), Generalitat de Catalunya (MENTAL-Cat; SLT006/17/287). This project has also received funding from the European Union’s Horizon 2020 Research and Innovation Programme under the grant agreements No 667302 (CoCA), 728018 (Eat2beNICE) and 848228 (DISCOvERIE).

**unsw1:** Recruitment of twins was supported by an Australian Research Council Linkage Grant LP0883621 (LMW, JMG, PRS), facilitated through access to Twins Research Australia, a national resource supported by NHMRC Centre of Research Excellence Grant (1079102). Subsequent studies were supported by National Health and Medical Research Council (NHMRC) Project Grants 1122816 (JMG), 1066177 (JMF), and Program Grant 1037196 (PRS).

**spal1: Rome, Italy, IRCCS Santa Lucia Foundation:** This study collection and the PI (Gianfranco Spalletta) were funded by the Italian Ministry of Health RC-10-11-12-13-14-15-16-17-18-19-20/A.

**Million Veteran Program** - The views expressed in this article are those of the authors, and do not necessarily reflect the position or policy of the Department of Veterans Affairs (VA). The Million Veteran Program (MVP) is funded by grant #MVP000 from the VA Office of Research and Development (ORD). This work was also funded by grant #1I01CX001729 from the Clinical Services Research & Development (CSRD) Service of VA ORD to Drs. Beckham and Kimbrel and by the MVP CHAMPION program, which is a collaboration between the VA and the Department of Energy (DoE).

### CONSORTIUM AUTHORS

**International Suicide Genetics Consortium**

Niamh Mullins 1, 2, Jooeun Kang 3, Adrian I Campos 4, 5, Jonathan R I Coleman 6, 7, Alexis C Edwards 8, Hanga Galfalvy 9, 10, Daniel F Levey 11, 12, Adriana Lori 13, Andrey Shabalin 14, Anna Starnawska 15, 16, 17, 18, Mei-Hsin Su 19, Hunna J Watson 20, 21, 22, Mark Adams 23, Swapnil Awasthi 24, Michael Gandal 25, Jonathan D Hafferty 23, Akitoyo Hishimoto 26, Minsoo Kim 25, Satoshi Okazaki 27, Ikuo Otsuka 10, 27, Stephan Ripke 24, 28, 29, Erin B Ware 30, 31, Andrew W Bergen 32, 33, Wade H Berrettini 34, Martin Bohus 35, Harry Brandt 36, Xiao Chang 37, Wei J Chen 19, 38, 39, Hsi-Chung Chen 39, Steven Crawford 36, Scott Crow 40, Emily DiBlasi 14, Philibert Duriez 41, 42, Fernando Fernández-Aranda 43, Manfred M Fichter 44, 45, Steven Gallinger 46, Stephen J Glatt 47, Philip Gorwood 41, 42, Yiran Guo 37, Hakon Hakonarson 37, 48, Katherine A Halmi 49, Hai-Gwo Hwu 50, Sonia Jain 51, Stéphane Jamain 52, Susana Jiménez-Murcia 43, Craig Johnson 53, Allan S Kaplan 54, 55, 56, Walter H Kaye 57, Pamela K Keel 58, James L Kennedy 54, 55, 56, Kelly L Klump 59, Dong Li 37, Shih-Cheng Liao 39, Klaus Lieb 60, Lisa Lilenfeld 61, Chih-Min Liu 39, Pierre J Magistretti 62, 63, Christian R Marshall 64, James E Mitchell 65, Eric T Monson 14, Richard M Myers 66, Dalila Pinto 1, 2, Abigail Powers 13, Nicolas Ramoz 42, Stefan Roepke 67, Vsevolod Rozanov 68, 69, Stephen W Scherer 70, Christian Schmahl 35, Marcus Sokolowski 71, Michael Strober 72, 73, Laura M Thornton 22, Janet Treasure 74, 75, Ming T Tsuang 76, Stephanie H Witt 77, D Blake Woodside 55, 56, 78, 79, Zeynep Yilmaz 22, 80, 81, Lea Zillich 77, Rolf Adolfsson 82, Ingrid Agartz 83, 84, 85, Tracy M Air 86, Martin Alda 87, 88, Lars Alfredsson 89, 90, Ole A Andreassen 91, 92, Adebayo Anjorin 93, Vivek Appadurai 94, 95, María Soler Artigas 96, 97, 98, 99, Sandra Van der Auwera 100, M Helena Azevedo 101, Nicholas Bass 102, Claiton HD Bau 103, 104, Bernhard T Baune 105, 106, Frank Bellivier 107, 108, 109, 110, Klaus Berger 111, Joanna M Biernacka 112, Tim B Bigdeli 113, 114, Elisabeth B Binder 13, 115, Michael Boehnke 116, Marco P Boks 117, Rosa Bosch 96, 97, 118, David L Braff 119, Richard Bryant 120, Monika Budde 121, Enda M Byrne 122, 123, Wiepke Cahn 124, Miguel Casas 96, 97, 99, 118, Enrique Castelao 125, Jorge A Cervilla 126, Boris Chaumette 127, 128, 129, Sven Cichon 130, 131, 132, 133, Aiden Corvin 134, Nicholas Craddock 135, David Craig 136, Franziska Degenhardt 133, Srdjan Djurovic 137, 138, Howard J Edenberg 139, 140, Ayman H Fanous 113, 114, Jerome C Foo 141, Andreas J Forstner 130, 133, 142, Mark Frye 143, Janice M Fullerton 144, 145, Justine M Gatt 120, 144, Pablo V Gejman 146, 147, Ina Giegling 148, 149, Hans J Grabe 100, Melissa J Green 144, 150, Eugenio H Grevet 151, 152, Maria Grigoroiu-Serbanescu 153, Blanca Gutierrez 154, Jose Guzman-Parra 155, Steven P Hamilton 156, Marian L Hamshere 135, Annette Hartmann 148, Joanna Hauser 157, Stefanie Heilmann-Heimbach 133, Per Hoffmann 131, 132, 133, Marcus Ising 158, Ian Jones 135, Lisa A Jones 159, Lina Jonsson 160, René S Kahn 2, 161, John R Kelsoe 119, 162, Kenneth S Kendler 114, Stefan Kloiber 54, 158, 163, Karestan C Koenen 164, 165, 166, Manolis Kogevinas 167, Bettina Konte 148, Marie-Odile Krebs 127, 128, 129, Mikael Landén 160, 168, Jacob Lawrence 169, Marion Leboyer 107, 170, 171, Phil H Lee 28, 29, 172, Douglas F Levinson 173, Calwing Liao 174, 175, Jolanta Lissowska 176, Susanne Lucae 158, Fermin Mayoral 155, Susan L McElroy 177, Patrick McGrath 178, Peter McGuffin 7, Andrew McQuillin 102, Sarah Medland 179, Divya Mehta 180, 181, Ingrid Melle 91, 182, Yuri Milaneschi 183, Philip B Mitchell 150, Esther Molina 184, Gunnar Morken 185, 186, Preben Bo Mortensen 16, 80, 95, 187, Bertram Müller-Myhsok 115, 188, 189, Caroline Nievergelt 119, Vishwajit Nimgaonkar 190, Markus M Nöthen 133, Michael C O'Donovan 135, Roel A Ophoff 191, 192, Michael J Owen 135, Carlos Pato 193, 194, Michele T Pato 194, Brenda WJH Penninx 183, 183, Jonathan Pimm 102, Giorgio Pistis 125, James B Potash 195, Robert A Power 7, 196, 197, Martin Preisig 125, Digby Quested 198, Josep Antoni Ramos-Quiroga 96, 97, 99, 118, Andreas Reif 199, Marta Ribasés 96, 97, 98, 99, Vanesa Richarte 96, 97, 118, Marcella Rietschel 200, Margarita Rivera 7, 201, Andrea Roberts 202, Gloria Roberts 150, Guy A Rouleau 175, 203, Diego L Rovaris 204, Dan Rujescu 148, Cristina Sánchez-Mora 96, 97, 98, 99, Alan R Sanders 146, 147, Peter R Schofield 144, 145, Thomas G Schulze 121, 141, 205, 206, 207, Laura J Scott 116, Alessandro Serretti 208, Jianxin Shi 209, Stanley I Shyn 210, Lea Sirignano 141, Pamela Sklar 1, 2, 211, Olav B Smeland 91, 92, Jordan W Smoller 28, 166, 212, Edmund J S Sonuga-Barke 213, Gianfranco Spalletta 214, 215, John S Strauss 54, 163, Beata Świątkowska 216, Maciej Trzaskowski 122, Gustavo Turecki 217, Laura Vilar-Ribó 96, 99, John B Vincent 218, Henry Völzke 219, James TR Walters 135, Cynthia Shannon Weickert 144, 150, Thomas W Weickert 144, 150, Myrna M Weissman 220, 221, Leanne M Williams 222, Naomi R Wray 122, 181, Clement C Zai 28, 163, 165, 223, 224, 225, Allison E. Ashley-Koch 226, Jean C. Beckham 227, 228, Elizabeth R. Hauser 226, 229, Michael A. Hauser 226, Nathan A. Kimbrel 227, 228, Jennifer H. Lindquist 230, Benjamin McMahon 231, David W. Oslin 232, 233, Xuejun Qin 226, Esben Agerbo 80, 187, 234, Anders D Børglum 15, 16, 17, 18, Gerome Breen 6, 7, Annette Erlangsen 18, 235, 236, 237, Tõnu Esko 238, 239, Joel Gelernter 11, 12, David M Hougaard 234, 240, Ronald C Kessler 241, Henry R Kranzler 242, 243, Qingqin S Li 244, Nicholas G Martin 245, Andrew M McIntosh 23, Sarah E Medland 245, Ole Mors 234, 246, Merete Nordentoft 234, 247, Catherine M Olsen 248, David Porteous 249, Robert J Ursano 250, Danuta Wasserman 71, Thomas Werge 94, 234, 251, 252, David C Whiteman 248, Cynthia M Bulik 22, 168, 253, Hilary Coon 14, 254, Ditte Demontis 15, 16, 17, 18, Anna R Docherty 8, 14, Po-Hsiu Kuo 19, 39, Cathryn M Lewis 7, 255, J John Mann 256, Miguel E Rentería 4, 5, Daniel J Smith 257, Eli A Stahl 1, 2, 238, Murray B Stein 258, Fabian Streit 77, Virginia Willour 259, Douglas M Ruderfer 3, 260, 261

**Affiliations**

1, Department of Genetics and Genomic Sciences, Icahn School of Medicine at Mount Sinai, New York, NY, US

2, Department of Psychiatry, Icahn School of Medicine at Mount Sinai, New York, NY, US

3, Division of Genetic Medicine, Department of Medicine, Vanderbilt Genetics Institute, Vanderbilt University Medical Center, Nashville, TN, US

4, Department of Genetics and Computational Biology, QIMR Berghofer Medical Research Institute, Brisbane, QLD, Australia

5, School of Biomedical Sciences, Faculty of Medicine, The University of Queensland, Brisbane, QLD, Australia

6, National Institute for Health Research (NIHR) Maudsley Biomedical Research Centre at South London and Maudsley NHS Foundation Trust, King's College London, London, UK

7, Social Genetic and Developmental Psychiatry Centre, King's College London, London, UK

8, Department of Psychiatry, Virginia Commonwealth University, Richmond, VA, US

9, Department of Biostatistics, Columbia University, New York, NY, US

10, Department of Psychiatry, Columbia University, New York, NY, US

11, Department of Psychiatry, Veterans Affairs Connecticut Healthcare Center, West Haven, CT, US

12, Division of Human Genetics, Department of Psychiatry, Yale University School of Medicine, New Haven, CT, US

13, Department of Psychiatry and Behavioral Sciences, Emory University School of Medicine, Atlanta, GA, US

14, Department of Psychiatry, University of Utah School of Medicine, Salt Lake City, UT, US

15, Centre for Genomics and Personalized Medicine, CGPM, Aarhus University, Aarhus, Denmark

16, Centre for Integrative Sequencing, iSEQ, Aarhus University, Aarhus, Denmark

17, Department of Biomedicine, Aarhus University, Aarhus, Denmark

18, The Lundbeck Foundation Initiative for Integrative Psychiatric Research, iPSYCH, Aarhus University, Aarhus, Denmark

19, Institute of Epidemiology and Preventive Medicine, College of Public Health, National Taiwan University, Taipei, Taiwan

20, School of Psychology, Curtin University, Perth, Western Australia, Australia

21, Division of Paediatrics, The University of Western Australia, Perth, Western Australia, Australia

22, Department of Psychiatry, University of North Carolina at Chapel Hill, Chapel Hill, NC, US

23, Division of Psychiatry, University of Edinburgh, Edinburgh, UK

24, Department of Psychiatry and Psychotherapy, Charité - Universitätsmedizin Berlin, Berlin, Germany

25, Department of Psychiatry and Biobehavioral Science, Semel Institute, David Geffen School of Medicine, University of California, Los Angeles, Los Angeles, CA, US

26, Department of Psychiatry, Yokohama City University Graduate School of Medicine, Yokohama, Japan

27, Department of Psychiatry, Kobe University Graduate School of Medicine, Kobe, Japan

28, Stanley Center for Psychiatric Research, Broad Institute, Cambridge, MA, US

29, Analytical and Translational Genetics Unit, Massachusetts General Hospital, Boston, MA, US

30, Population Studies Center, Institute for Social Research, University of Michigan, Ann Arbor, MI, US

31, Survery Research Center, Institute for Social Research, University of Michigan, Ann Arbor, MI, US

32, BioRealm, LLC, Walnut, CA, US

33, Oregon Research Institute, Eugene, OR, US

34, Department of Psychiatry, Center for Neurobiology and Behavior, Perelman School of Medicine at the University of Pennsylvania, Philadelphia, PA, US

35, Department of Psychosomatic Medicine and Psychotherapy, Central Institute of Mental Health, Medical Faculty Mannheim, University of Heidelberg, Mannheim, Germany

36, The Center for Eating Disorders at Sheppard Pratt, Baltimore, MD, US

37, Center for Applied Genomics, Children's Hospital of Philadelphia, Philadelphia, PA, US

38, Center for Neuropsychiatric Research, National Health Research Institutes, Miaoli County, Taiwan

39, Department of Psychiatry, National Taiwan University Hospital, Taipei, Taiwan

40, Department of Psychiatry, University of Minnesota, Minneapolis, MN, US

41, Hôpital Sainte Anne, GHU Paris Psychiatrie et Neurosciences, Paris, France

42, Institute of Psychiatry and Neuroscience of Paris (IPNP), INSERM U1266, Université de Paris, Paris, France

43, Department of Psychiatry, University Hospital Bellvitge-IDIBELL and CIBEROBN, Barcelona, Spain

44, Department of Psychiatry and Psychotherapy, Ludwig-Maximilians-University (LMU), Munich, Germany

45, Schön Klinik Roseneck affiliated with the Medical Faculty of the University of Munich (LMU), Munich, Germany

46, Department of Surgery, Faculty of Medicine, University of Toronto, Toronto, Canada

47, Department of Psychiatry and Behavioral Sciences, SUNY Upstate Medical University, Syracuse, NY, US

48, The Perelman School of Medicine, University of Pennsylvania, Philadelphia, PA, US

49, Department of Psychiatry, Weill Cornell Medical College, New York, NY, US

50, Department of Psychiatry, National Taiwan University Hospital and College of Medicine, Taipei, Taiwan

51, Biostatistics Research Center, Herbert Wertheim School of Public Health and Human Longevity Science, University of California San Diego, La Jolla, CA, US

52, Inserm U955, Institut Mondor de recherches Biomédicales, Laboratoire, Neuro-Psychiatrie Translationnelle, and Fédération Hospitalo-Universitaire de Précision Médecine en Addictologie et Psychiatrie (FHU ADAPT), University Paris-Est-Créteil, Créteil, France

53, Eating Recovery Center, Denver, CO, US

54, Centre for Addiction and Mental Health, Toronto, ON, Canada

55, Department of Psychiatry, University of Toronto, Toronto, Canada

56, Institute of Medical Science, University of Toronto, Toronto, Canada

57, Department of Psychiatry, University of California San Diego, San Diego, CA, US

58, Department of Psychology, Florida State University, Tallahassee, FL, US

59, Department of Psychology, Michigan State University, Lansing, MI, US

60, Department of Psychiatry and Psychotherapy, University Medical Center, Mainz, Germany

61, Department of Clinical Psychology, The Chicago School of Professional Psychology, Washington DC, Washington, DC, US

62, BESE Division, King Abdullah University of Science and Technology, Thuwal, Saudi Arabia

63, Department of Psychiatry, University of Lausanne-University Hospital of Lausanne (UNIL-CHUV), Lausanne, Switzerland

64, Department of Paediatric Laboratory Medicine, The Hospital for Sick Children, Toronto, Canada

65, Department of Psychiatry and Behavioral Science, University of North Dakota School of Medicine and Health Sciences, Fargo, ND, US

66, HudsonAlpha Institute for Biotechnology, Huntsville, AL, US

67, Department of Psychiatry, Charité - Universitätsmedizin Berlin, Corporate Member of Freie Universität Berlin, Humboldt-Universität zu Berlin, Berlin Institute of Health, Campus Benjamin Franklin, Berlin, Germany

68, Department of Psychology, Saint-Petersburg State University, Saint-Petersburg, Russian Federation

69, Department of Borderline Disorders and Psychotherapy, V.M. Bekhterev National Medical Research Center for Psychiatry and Neurology, Saint-Petersburg, Russian Federation

70, Department of Genetics and Genomic Biology, The Hospital for Sick Children, Toronto, Canada

71, National Centre for Suicide Research and Prevention of Mental Ill-Health (NASP), LIME, Karolinska Institutet, Stockholm, Sweden

72, David Geffen School of Medicine, University of California Los Angeles, Los Angeles, LA, US

73, Department of Psychiatry and Biobehavioral Science, Semel Institute for Neuroscience and Human Behavior, University of California Los Angeles, Los Angeles, LA, US

74, Institute of Psychiatry, Psychology and Neuroscience, Department of Psychological Medicine, King’s College London, London, UK

75, National Institute for Health Research Biomedical Research Centre, King’s College London and South London and Maudsley National Health Service Foundation Trust, London, UK

76, Center for Behavioral Genomics, Department of Psychiatry, University of California, San Diego, San Diego, CA, US

77, Department of Genetic Epidemiology in Psychiatry, Central Institute of Mental Health, Medical Faculty Mannheim, University of Heidelberg, Mannheim, Germany

78, Centre for Mental Health, University Health Network, Toronto, Canada

79, Program for Eating Disorders, University Health Network, Toronto, Canada

80, National Centre for Register-Based Research, Aarhus University, Aarhus, Denmark

81, Department of Genetics, University of North Carolina at Chapel Hill, Chapel Hill, NC, US

82, Department of Clinical Sciences, Psychiatry, Umeå University Medical Faculty, Umeå, Sweden

83, Department of Psychiatric Research, Diakonhjemmet Hospital, Oslo, Norway

84, Department of Clinical Neuroscience, Centre for Psychiatry Research, Karolinska Institutet, Stockholm, Sweden

85, NORMENT, Institute of Clinical Medicine, University of Oslo, Oslo, Norway

86, Discipline of Psychiatry, University of Adelaide, Adelaide, SA, Australia

87, Department of Psychiatry, Dalhousie University, Halifax, NS, Canada

88, National Institute of Mental Health, Klecany, CZ

89, Department of Clinical Neuroscience, Karolinska Institutet, Stockholm, Sweden

90, Inst of Environmental Medicine, Karolinska Institutet, Stockholm, Sweden

91, Division of Mental Health and Addiction, Oslo University Hospital, Oslo, Norway

92, NORMENT, University of Oslo, Oslo, Norway

93, Psychiatry, Berkshire Healthcare NHS Foundation Trust, Bracknell, UK

94, Institute of Biological Psychiatry, Copenhagen Mental Health Services, Copenhagen University Hospital, Copenhagen, Denmark

95, The Lundbeck Foundation Initiative for Integrative Psychiatric Research, iPSYCH, Copenhagen, Denmark

96, Department of Psychiatry, Hospital Universitari Vall d’Hebron, Barcelona, Spain

97, Biomedical Network Research Centre on Mental Health (CIBERSAM), Instituto de Salud Carlos III, Madrid, Spain

98, Department of Genetics, Microbiology & Statistics, University of Barcelona, Barcelona, Spain

99, Psychiatric Genetics Unit, Group of Psychiatry, Mental Health and Addiction, Vall d’Hebron Research Institute (VHIR), Universitat Autònoma de Barcelona, Barcelona, Spain

100, Department of Psychiatry and Psychotherapy, University Medicine Greifswald, Greifswald, Mecklenburg-Vorpommern, Germany

101, Department of Psychiatry, University of Coimbra, Coimbra, Portugal

102, Division of Psychiatry, University College London, London, UK

103, Laboratory of Developmental Psychiatry, Hospital de Clínicas de Porto Alegre, Porto Alegre, RS, Brazil

104, Department of Genetics, Universidade Federal do Rio Grande do Sul, Porto Alegre, RS, Brazil

105, Department of Psychiatry, Melbourne Medical School, University of Melbourne, Melbourne, Australia

106, Department of Psychiatry, University of Münster, Münster, Germany

107, Department of Psychiatry and Addiction Medicine, Assistance Publique - Hôpitaux de Paris, Paris, France

108, Paris Bipolar and TRD Expert Centres, FondaMental Foundation, Paris, France

109, UMR-S1144 Team 1 : Biomarkers of relapse and therapeutic response in addiction and mood disorders, INSERM, Paris, France

110, Psychiatry, Université Paris Diderot, Paris, France

111, Institute of Epidemiology and Social Medicine, University of Münster, Münster, Nordrhein-Westfalen, Germany

112, Health Sciences Research, Mayo Clinic, Rochester, MN, US

113, Department of Psychiatry and Behavioral Sciences, State University of New York Downstate Medical Center, New York, NY, US

114, Department of Psychiatry, Virginia Commonwealth University, Richmond, VA, US

115, Department of Translational Research in Psychiatry, Max Planck Institute of Psychiatry, Munich, Germany

116, Center for Statistical Genetics and Department of Biostatistics, University of Michigan, Ann Arbor, MI, US

117, Psychiatry, UMC Utrecht Hersencentrum, Utrecht, Netherlands

118, Department of Psychiatry and Legal Medicine, Universitat Autònoma de Barcelona, Barcelona, Spain

119, Department of Psychiatry, University of California San Diego, La Jolla, CA, US

120, School of Psychology, University of New South Wales, Sydney, NSW, Australia

121, Institute of Psychiatric Phenomics and Genomics (IPPG), University Hospital, LMU Munich, Munich, Germany

122, Institute for Molecular Bioscience, The University of Queensland, Brisbane, QLD, Australia

123, Centre for Children’s Health Research, The University of Queensland, Brisbane, QLD, Australia

124, Department of Psychiatry, UMC Utrecht Hersencentrum Rudolf Magnus, Utrecht, Netherlands

125, Department of Psychiatry, Lausanne University Hospital and University of Lausanne, Lausanne, Vaud, Switzerland

126, Mental Health Unit, Department of Psychiatry, Faculty of Medicine, Granada University Hospital Complex, University of Granada, Granada, Spain

127, Institut de Psychiatrie, CNRS GDR 3557, Paris, France

128, Department of Evaluation, Prevention and Therapeutic innovation, GHU Paris Psychiatrie et Neurosciences, Paris, France

129, Team Pathophysiology of psychiatric diseases, Université de Paris, Institute of Psychiatry and Neuroscience of Paris (IPNP), INSERM U1266, Paris, France

130, Institute of Neuroscience and Medicine (INM-1), Research Centre Jülich, Jülich, Germany

131, Institute of Medical Genetics and Pathology, University Hospital Basel, Basel, Switzerland

132, Department of Biomedicine, University of Basel, Basel, Switzerland

133, Institute of Human Genetics, University of Bonn, School of Medicine & University Hospital Bonn, Bonn, Germany

134, Neuropsychiatric Genetics Research Group, Dept of Psychiatry and Trinity Translational Medicine Institute, Trinity College Dublin, Dublin, Ireland

135, Medical Research Council Centre for Neuropsychiatric Genetics and Genomics, Division of Psychological Medicine and Clinical Neurosciences, Cardiff University, Cardiff, UK

136, Department of Translational Genomics, University of Southern California, Pasadena, CA, US

137, Department of Medical Genetics, Oslo University Hospital, Oslo, Norway

138, NORMENT, KG Jebsen Centre for Psychosis Research, Department of Clinical Science, University of Bergen, Bergen, Norway

139, Department of Medical & Molecular Genetics, Indiana University, Indianapolis, IN, US

140, Biochemistry and Molecular Biology, Indiana University School of Medicine, Indianapolis, IN, US

141, Department of Genetic Epidemiology in Psychiatry, Central Institute of Mental Health, Medical Faculty Mannheim, Heidelberg University, Mannheim, Germany

142, Centre for Human Genetics, University of Marburg, Marburg, Germany

143, Department of Psychiatry & Psychology, Mayo Clinic, Rochester, MN, US

144, Neuroscience Research Australia, Sydney, NSW, Australia

145, School of Medical Sciences, University of New South Wales, Sydney, NSW, Australia

146, Department of Psychiatry and Behavioral Sciences, NorthShore University HealthSystem, Evanston, IL, US

147, Department of Psychiatry and Behavioral Neuroscience, University of Chicago, Chicago, IL, US

148, Dept. of Psychiatry, Psychotherapy and Psychosomatics, Martin-Luther-University Halle-Wittenberg, Halle (Saale), Germany

149, Department of Psychiatry, University of Munich, Munich, Germany

150, School of Psychiatry, University of New South Wales, Sydney, NSW, Australia

151, ADHD Outpatient Program, Adult Division, Hospital de Clínicas de Porto Alegre, Porto Alegre, RS, Brazil

152, Department of Psychiatry, Universidade Federal do Rio Grande do Sul, Porto Alegre, RS, Brazil

153, Biometric Psychiatric Genetics Research Unit, Alexandru Obregia Clinical Psychiatric Hospital, Bucharest, Romania

154, Department of Psychiatry, Faculty of Medicine and Biomedical Research Centre (CIBM), University of Granada, Granada, Spain

155, Mental Health Department, University Regional Hospital. Biomedicine Institute (IBIMA), Málaga, Spain

156, Psychiatry, Kaiser Permanente Northern California, San Francisco, CA, US

157, Department of Psychiatry, Laboratory of Psychiatric Genetics, Poznan University of Medical Sciences, Poznan, Poland

158, Max Planck Institute of Psychiatry, Munich, Germany

159, Department of Psychological Medicine, University of Worcester, Worcester, UK

160, Department of Psychiatry and Neuroscience, University of Gothenburg, Gothenburg, Sweden

161, Psychiatry, UMC Utrecht Hersencentrum Rudolf Magnus, Utrecht, Netherlands

162, Institute for Genomic Medicine, University of California San Diego, La Jolla, CA, US

163, Department of Psychiatry, University of Toronto, Toronto, ON, Canada

164, Stanley Center for Psychiatric Research, Broad Institute, Cambridge, MA, US

165, Department of Epidemiology, Harvard TH Chan School of Public Health, Boston, MA, US

166, Department of Psychiatry, Massachusetts General Hospital, Boston, MA, US

167, Center for Research in Environmental Epidemiology (CREAL), Barcelona, Spain

168, Department of Medical Epidemiology and Biostatistics, Karolinska Institutet, Stockholm, Sweden

169, Psychiatry, North East London NHS Foundation Trust, Ilford, UK

170, INSERM, Paris, France

171, Faculté de Médecine, Université Paris Est, Créteil, France

172, Psychiatric and Neurodevelopmental Genetics Unit, Massachusetts General Hospital, Boston, MA, US

173, Psychiatry & Behavioral Sciences, Stanford University, Stanford, CA, US

174, Department of Human Genetics, McGill University, Montreal, QC, Canada

175, Montreal Neurological Institute and Hospital, Montreal, QC, Canada

176, Cancer Epidemiology and Prevention, M. Sklodowska-Curie Cancer Center and Institute of Oncology, Warsaw, Poland

177, Research Institute, Lindner Center of HOPE, Mason, OH, US

178, Psychiatry, Columbia University College of Physicians and Surgeons, New York, NY, US

179, Genetics and Computational Biology, QIMR Berghofer Medical Research Institute, Brisbane, QLD, Australia

180, School of Psychology and Counseling, Queensland University of Technology, Brisbane, QLD, Australia

181, Queensland Brain Institute, The University of Queensland, Brisbane, QLD, Australia

182, Division of Mental Health and Addiction, University of Oslo, Institute of Clinical Medicine, Oslo, Norway

183, Department of Psychiatry, Amsterdam UMC, Vrije Universiteit and GGZ inGeest, Amsterdam, Netherlands

184, Department of Nursing, Faculty of Health Sciences and Biomedical Research Centre (CIBM), University of Granada, Granada, Spain

185, Mental Health, Faculty of Medicine and Health Sciences, Norwegian University of Science and Technology - NTNU, Trondheim, Norway

186, Psychiatry, St Olavs University Hospital, Trondheim, Norway

187, Centre for Integrated Register-based Research, Aarhus University, Aarhus, Denmark

188, Munich Cluster for Systems Neurology (SyNergy), Munich, Germany

189, University of Liverpool, Liverpool, UK

190, Psychiatry and Human Genetics, University of Pittsburgh, Pittsburgh, PA, US

191, Psychiatry, Erasmus University Medical Center, Rotterdam, Netherlands

192, Jane and Terry Semel Institute for Neuroscience and Human Behavior, Los Angeles, CA, US

193, College of Medicine Institute for Genomic Health, SUNY Downstate Medical Center College of Medicine, Brooklyn, NY, US

194, Institute for Genomic Health, SUNY Downstate Medical Center College of Medicine, Brooklyn, NY, US

195, Psychiatry, University of Iowa, Iowa City, IA, US

196, Genetics, BioMarin Pharmaceuticals, London, UK

197, St Edmund Hall, University of Oxford, Oxford, UK

198, Department of Psychiatry, University of Oxford, Oxford, UK

199, Department of Psychiatry, Psychosomatic Medicine and Psychotherapy, University Hospital Frankfurt, Frankfurt, Germany

200, Department of Genetic Epidemiology in Psychiatry, Central Institute of Mental Health, Medical Faculty Mannheim, Heidelberg University, Mannheim, Baden-Württemberg, Germany

201, Department of Biochemistry and Molecular Biology II and Institute of Neurosciences, Biomedical Research Centre (CIBM), University of Granada, Granada, Spain

202, Department of Environmental Health, Harvard TH Chan School of Public Health, Boston, MA, US

203, Department of Neurology and Neurosurgery, McGill University, Faculty of Medicine, Montreal, QC, Canada

204, Department of Physiology and Biophysics, Instituto de Ciencias Biomedicas Universidade de Sao Paulo, São Paulo, SP, Brazil

205, Department of Psychiatry and Behavioral Sciences, Johns Hopkins University School of Medicine, Baltimore, MD, US

206, Human Genetics Branch, Intramural Research Program, National Institute of Mental Health, Bethesda, MD, US

207, Department of Psychiatry and Psychotherapy, University Medical Center Göttingen, Göttingen, Germany

208, Department of Biomedical and NeuroMotor Sciences, University of Bologna, Bologna, Italy

209, Division of Cancer Epidemiology and Genetics, National Cancer Institute, Bethesda, MD, US

210, Behavioral Health Services, Kaiser Permanente Washington, Seattle, WA, US

211, Department of Neuroscience, Icahn School of Medicine at Mount Sinai, New York, NY, US

212, Psychiatric and Neurodevelopmental Genetics Unit (PNGU), Massachusetts General Hospital, Boston, MA, US

213, Institute of Psychology, Psychiatry & Neuroscience, King's College London, London, UK

214, Menninger Department of Psychiatry and Behavioral Sciences, Baylor College of Medicine, Houston, Houston, TX, US

215, Laboratory of Neuropsychiatry, IRCCS Santa Lucia Foundation, Rome, Rome, Italy

216, Department of Environmental Epidemiology, Nofer Institute of Occupational Medicine, Lodz, Poland

217, Department of Psychiatry, McGill University, Montreal, QC, Canada

218, Molecular Brain Science, Centre for Addiction and Mental Health, Toronto, ON, Canada

219, Institute for Community Medicine, University Medicine Greifswald, Greifswald, Mecklenburg-Vorpommern, Germany

220, Columbia University College of Physicians and Surgeons, New York, NY, US

221, Division of Translational Epidemiology, New York State Psychiatric Institute, New York, NY, US

222, Department of Psychiatry and Behavioral Sciences, Stanford University, Stanford, CA, US

223, Institute of Medical Science, University of Toronto, Toronto, ON, Canada

224, Molecular Brain Science, Campbell Family Mental Health Research Institute, Centre for Addiction and Mental Health, Toronto, ON, Canada

225, Laboratory Medicine and Pathobiology, University of Toronto, Toronto, ON, Canada

226, Duke Molecular Physiology Institute, Duke University Medical Center, Durham, NC, USA

227, VISN 6 Mid-Atlantic Mental Illness Research, Education, and Clinical Center, Durham Veterans Affairs Health Care System, Durham, NC, USA

228, Department of Psychiatry and Behavioral Sciences, Duke University School of Medicine, Durham, NC, USA

229, Cooperative Studies Program Epidemiology Center, Durham Veterans Affairs Health Care System, Durham, NC, USA

230, VA Health Services Research and Development Center of Innovation to Accelerate Discovery and Practice Transformation, Durham Veterans Affairs Health Care System, Durham, NC, USA

231, Theoretical Division, Los Alamos National Laboratory, Los Alamos National Laboratory, Los Alamos, NM, USA

232, VISN 4 Mental Illness Research, Education, and Clinical Center, Corporal Michael J. Crescenz VA Medical Center, Philadelphia, PA, USA

233, Department of Psychiatry, Perelman School of Medicine, University of Pennsylvania, Philadelphia, PA, USA

234, The Lundbeck Foundation Initiative for Integrative Psychiatric Research, iPSYCH, Aarhus, Denmark

235, Center of Mental Health Research, Australian National University, Canberra, Australia

236, Department of Mental Health, Johns Hopkins Bloomberg School of Public Health, Baltimore, MD, US

237, Danish Research Institute for Suicide Prevention, Mental Health Centre Copenhagen, Copenhagen, Denmark

238, Program in Medical and Population Genetics, Broad Institute, Cambridge, MA, US

239, Estonian Genome Center, Institute of Genomics, University of Tartu, Tartu, Estonia

240, Center for Neonatal Screening, Department for Congenital Disorders, Statens Serum Institut, Copenhagen, Denmark

241, Department of Health Care Policy, Harvard Medical School, Boston, MA, US

242, Department of Psychiatry, University of Pennsylvania Perelman School of Medicine, Philadelphia, PA, US

243, VISN 4 MIRECC, Crescenz VAMC, Philadelphia, PA, US

244, Neuroscience, Janssen Research & Development, LLC, Titusville, NJ, US

245, Department of Genetics and Computational Biology, QIMR Berghofer Medical Research Institute, Herston, QLD, Australia

246, Psychosis Research Unit, Aarhus University Hospital, Risskov, Aarhus, Denmark

247, Mental Health Center Copenhagen, Copenhagen University Hospital, Copenhagen, Denmark

248, Department of Population Health, QIMR Berghofer Medical Research Institute, Herston, QLD, Australia

249, Institute for Genetics and Molecular Medicine, University of Edinburgh, Edinburgh, UK

250, Department of Psychiatry, Uniformed University of the Health Sciences, Bethesda, MD, US

251, Department of Clinical Medicine, University of Copenhagen, Copenhagen, Denmark

252, Lundbeck Foundation GeoGenetics Centre, GLOBE Institute, University of Copenhagen, Copenhagen, Denmark

253, Department of Nutrition, University of North Carolina at Chapel Hill, Chapel Hill, NC, US

254, Biomedical Informatics, University of Utah School of Medicine, Salt Lake City, UT, US

255, Department of Medical & Molecular Genetics, King's College London, London, UK

256, Departments of Psychiatry and Radiology, Columbia University, New York, NY, US

257, Institute of Health and Wellbeing, University of Glasgow, Glasgow, UK

258, Department of Psychiatry and School of Public Health, University of California San Diego, La Jolla, CA, US

259, Department of Psychiatry, University of Iowa, Iowa City, IA, US

260, Department of Biomedical Informatics, Vanderbilt University Medical Center, Nashville, TN, US

261, Department of Psychiatry and Behavioral Sciences, Vanderbilt University Medical Center, Nashville, TN, US

The **MVP Suicide Exemplar Workgroup** contributors for this manuscript include Silvia Crivelli, PhD, Lawrence Berkeley National Laboratory; Michelle F. Dennis, BA, Durham Veterans Affairs (VA) Health Care System & Duke University School of Medicine; Phillip D. Harvey, PhD, University of Miami Miller School of Medicine, Miami, FL, United States of America; Bruce W. Carter VA Medical Center; Elizabeth R. Hauser, PhD, Durham VA Health Care System & Duke University School of Medicine; Michael A. Hauser, PhD, Duke University School of Medicine; Jennifer E. Huffman, PhD, Massachusetts Veterans Epidemiology Research and Information Center (MAVERIC), VA Boston Healthcare System; Daniel Jacobson, PhD, Oak Ridge National Laboratory; Jennifer H. Lindquist, MS, Durham VA Health Care System; Ravi Madduri, PhD, Argonne National Laboratory; Maren K. Olsen, PhD, Duke University School of Medicine; John Pestian, PhD, Oak Ridge National Laboratory.

**The VA Million Veteran Program (MVP)** contributors for this manuscript include J. Michael Gaziano, MD, MPH, co-chair, VA Boston Healthcare System; Sumitra Muralidhar, PhD, co-chair, US Department of Veterans Affairs; Rachel Ramoni, DMD, ScD, US Department of Veterans Affairs; Jean Beckham, PhD, Durham VA Medical Center; Kyong-Mi Chang, MD, Philadelphia VA Medical Center; Christopher J. O’Donnell, MD, MPH, VA Boston Healthcare System; Philip S. Tsao, PhD, VA Palo Alto Health Care System; James Breeling, MD, Ex-Officio, US Department of Veterans Affairs; Grant Huang, PhD, Ex-Officio, US Department of Veterans Affairs; and J.P. Casas Romero, MD, PhD, Ex-Officio, VA Boston Healthcare System. MVP Program Office: Sumitra Muralidhar, PhD, and Jennifer Moser, PhD, both of US Department of Veterans Affairs. MVP Recruitment/Enrollment: Recruitment/Enrollment Director/Deputy Director, Boston—Stacey B. Whitbourne, PhD; Jessica V. Brewer, MPH, VA Boston Healthcare System; MVP Coordinating Centers: Clinical Epidemiology Research Center (CERC), West Haven—Mihaela Aslan, PhD, West Haven VA Medical Center; Cooperative Studies Program Clinical Research Pharmacy Coordinating Center, Albuquerque—Todd Connor, PharmD; Dean P Argyres, BS, MS, New Mexico VA Health Care System; Genomics Coordinating Center, Palo Alto—Philip S. Tsao, PhD, VA Palo Alto Health Care System; MVP Boston Coordinating Center, Boston—J. Michael Gaziano, MD, MPH, VA Boston Healthcare System; MVP Information Center, Canandaigua—Brady Stephens, MS, Canandaigua VA Medical Center; VA Central Biorepository, Boston—Mary T Brophy MD, MPH; Donald E Humphries, PhD; Luis E Selva, PhD, VA Boston Healthcare System; MVP Informatics, Boston—Nhan Do, MD; Shahpoor Shayan, VA Boston Healthcare System; MVP Data Operations/Analytics, Boston—Kelly Cho, PhD, VA Boston Healthcare System. MVP Science: Science Operations—Christopher J. O’Donnell, MD, MPH, VA Boston Healthcare System; Genomics Core— Christopher J. O’Donnell, MD, MPH; Saiju Pyarajan PhD, VA Boston Healthcare System; Philip S. Tsao, PhD, VA Palo Alto Health Care System; Phenomics Core—Kelly Cho, MPH, PhD, VA Boston Healthcare System; Data and Computational Sciences—Saiju Pyarajan, PhD, VA Boston Healthcare System; Statistical Genetics—Elizabeth Hauser, PhD, Durham VA Medical Center; Yan Sun, PhD, Atlanta VA Medical Center; Hongyu Zhao, PhD, West Haven VA Medical Center. Current MVP Local Site Investigators: Atlanta VA Medical Center, Peter Wilson, MD; Bay Pines VA Healthcare System, Rachel McArdle, PhD; Birmingham VA Medical Center, Louis Dellitalia, MD; Central Western Massachusetts Healthcare System, Kristin Mattocks, PhD, MPH; Cincinnati VA Medical Center, John Harley, MD, PhD; Clement J. Zablocki VA Medical Center, Jeffrey Whittle, MD, MPH; VA Northeast Ohio Healthcare System, Frank Jacono, MD; Durham VA Medical Center, Jean Beckham, PhD; Edith Nourse Rogers Memorial Veterans Hospital; Edward Hines, Jr VA Medical Center, Salvador Gutierrez, MD; Veterans Health Care System of the Ozarks, Gretchen Gibson, DDS, MPH; Fargo VA Health Care System, Kimberly Hammer, PhD; VA Health Care Upstate New York, Laurence Kaminsky, PhD; New Mexico VA Health Care System, Gerardo Villareal, MD; VA Boston Healthcare System, Scott Kinlay, MBBS, PhD; VA Western New York Healthcare System, Junzhe Xu, MD; Ralph H Johnson VA Medical Center, Mark Hamner, MD; Columbia VA Health Care System, Roy Mathew, MD; VA North Texas Health Care System, Sujata Bhushan, MD; Hampton VA Medical Center, Pran Iruvanti, DO, PhD; Richmond VA Medical Center, Michael Godschalk, MD; Iowa City VA Health Care System, Zuhair Ballas, MD; Eastern Oklahoma VA Health Care System, Douglas Ivins, MD; James A. Haley Veterans’ Hospital, Stephen Mastorides, MD; James H. Quillen VA Medical Center, Jonathan Moorman, MD, PhD; John D Dingell VA Medical Center, Saib Gappy, MD; Louisville VA Medical Center, Jon Klein, MD, PhD; Manchester VA Medical Center, Nora Ratcliffe, MD; Miami VA Health Care System, Hermes Florez, MD, PhD; Michael E. DeBakey VA Medical Center, Olaoluwa Okusaga, MD; Minneapolis VA Health Care System, Maureen Murdoch, MD, MPH; N FL/S GA Veterans Health System, Peruvemba Sriram, MD; Northport VA Medical Center, Shing Shing Yeh, PhD, MD; Overton Brooks VA Medical Center, Neeraj Tandon, MD; Philadelphia VA Medical Center, Darshana Jhala, MD; Phoenix VA Health Care System, Samuel Aguayo, MD; Portland VA Medical Center, David Cohen, MD; Providence VA Medical Center, Satish Sharma, MD; Richard Roudebush VA Medical Center, Suthat Liangpunsakul, MD, MPH; Salem VA Medical Center, Kris Ann Oursler, MD; San Francisco VA Health Care System, Mary Whooley, MD; South Texas Veterans Health Care System, Sunil Ahuja, MD; Southeast Louisiana Veterans Health Care System, Joseph Constans, PhD; Southern Arizona VA Health Care System, Paul Meyer, MD, PhD; Sioux Falls VA Health Care System, Jennifer Greco, MD; St Louis VA Health Care System, Michael Rauchman, MD; Syracuse VA Medical Center, Richard Servatius, PhD; VA Eastern Kansas Health Care System, Melinda Gaddy, PhD; VA Greater Los Angeles Health Care System, Agnes Wallbom, MD, MS, 11301 Wilshire Blvd, Los Angeles, CA 90073; VA Long Beach Healthcare System, Timothy Morgan, MD; VA Maine Healthcare System, Todd Stapley, DO; VA New York Harbor Healthcare System, Scott Sherman, MD, MPH; VA Pacific Islands Health Care System, George Ross, MD; VA Palo Alto Health Care System, Philip Tsao, PhD; VA Pittsburgh Health Care System, Patrick Strollo, Jr, MD; VA Puget Sound Health Care System, Edward Boyko, MD; VA Salt Lake City Health Care System, Laurence Meyer, MD, PhD; VA San Diego Healthcare System, Samir Gupta, MD, MSCS; VA Sierra Nevada Health Care System, Mostaqul Huq, PharmD, PhD; VA Southern Nevada Healthcare System, Joseph Fayad, MD; VA Tennessee Valley Healthcare System, Adriana Hung, MD, MPH; Washington DC VA Medical Center, Jack Lichy, MD, PhD; W.G., Bill Hefner VA Medical Center, Robin Hurley, MD; White River Junction VA Medical Center, Brooks Robey, MD; William S. Middleton Memorial Veterans Hospital, Robert Striker, MD, PhD.

**Other Participating Consortia:**

German Borderline Genomics Consortium

Stephanie H Witt^1,2^, Fabian Streit^1^, Martin Jungkunz^3,4^, Josef Frank^1^, Swapnil Awasthi^5^, Lea Zillich^1^, Alisha Hall^1^, Cornelia E Schwarze^6^, Ina Giegling^7^, Norbert Dahmen^8^, Björn H Schott^5,9,10^, Nikolaus Kleindienst^3^, Markus M Nöthen^11^, Stephan Ripke^5,12,13^, Arian Mobascher^14^, Dan Rujescu^15^, Klaus Lieb^8^, Stefan Roepke^16^, Christian Schmahl^3^, Martin Bohus^3,17^, Marcella Rietschel^1^

##### Affiliation

^1^ Central Institute of Mental Health, Department of Genetic Epidemiology in Psychiatry, Medical Faculty Mannheim, Heidelberg University, Mannheim, Germany

^2^ Central Institute of Mental Health, Center for Innovative Psychiatry and Psychotherapy, Medical Faculty Mannheim, Heidelberg University, Mannheim, Germany

^3^ Central Institute of Mental Health, Department of Psychosomatic Medicine and Psychotherapy, Medical Faculty Mannheim, Heidelberg University, Mannheim, Germany

^4^ Section for Translational Medical Ethics, National Center for Tumor Diseases, German Cancer Research Center (DKFZ), Heidelberg, Germany

^5^ Charité Universitätsmedizin Berlin, Department of Psychiatry and Psychotherapy, Campus Mitte, Berlin, Germany

^6^ Department of Developmental and Biological Psychology, Heidelberg University, Heidelberg, Germany

^7^ Medical University of Vienna, Comprehensive Centers for Clinical Neurosciences and Mental Health, Vienna, Austria

^8^ University Medical Center, Department of Psychiatry and Psychotherapy, Mainz, Germany

^9^ Leibniz Institute for Neurobiology, Magdeburg, Germany

^10^ Department of Psychiatry and Psychotherapy, University Medical Center Göttingen, Göttingen, Germany

^11^ University Hospital Bonn, Institute of Human Genetics, Bonn, Germany

^12^ Broad Institute of MIT and Harvard, Stanley Center for Psychiatric Research and Medical and Population Genetics Program, Cambridge, MA, USA

^13^ Massachusetts General Hospital and Department of Medicine, Harvard Medical School, Analytic and Translational Genetics Unit, Boston, MA, USA

^14^ St. Elisabeth Krankenhaus Lahnstein, Department of Psychiatry and Psychotherapy, Lahnstein, Germany

^15^ Medical University of Vienna, Department of Psychiatry and Psychotherapy, Vienna, Austria

^16^ Department of Psychiatry and Psychotherapy, Charité - Universitätsmedizin Berlin, Corporate Member of Freie Universität Berlin, Humboldt-Universität zu Berlin, Berlin Institute of Health, Campus Benjamin Franklin, Charité - Medical Faculty Berlin, Berlin

^17^ Department of Clinical Psychology and Psychotherapy, Ruhr University Bochum, Germany

**The Major Depressive Disorder Working Group of the Psychiatric Genomics Consortium**

| Naomi R Wray* 1, 2  Stephan Ripke* 3, 4, 5  Manuel Mattheisen* 6, 7, 8  Maciej Trzaskowski 1  Enda M Byrne 1  Abdel Abdellaoui 9  Mark J Adams 10  Esben Agerbo 11, 12, 13  Tracy M Air 14  Till F M Andlauer 15, 16  Silviu-Alin Bacanu 17  Marie Bækvad-Hansen 13, 18  Aartjan T F Beekman 19  Tim B Bigdeli 17, 20  Elisabeth B Binder 15, 21  Julien Bryois 22  Henriette N Buttenschøn 13, 23, 24  Jonas Bybjerg-Grauholm 13, 18  Na Cai 25, 26  Enrique Castelao 27  Jane Hvarregaard Christensen 8, 13, 24  Toni-Kim Clarke 10  Jonathan R I Coleman 28  Lucía Colodro-Conde 29  Baptiste Couvy-Duchesne 2, 30  Nick Craddock 31  Gregory E Crawford 32, 33  Gail Davies 34  Franziska Degenhardt 35  Eske M Derks 29  Nese Direk 36, 37  Anna R. Docherty, 20  Conor V Dolan 9  Erin C Dunn 38, 39, 40  Thalia C Eley 28  Valentina Escott-Price 41  Farnush Farhadi Hassan Kiadeh 42  Hilary K Finucane 43, 44  Jerome C Foo 45  Andreas J Forstner 35, 46, 47, 48  Josef Frank 45  Héléna A Gaspar 28  Michael Gill 49  Fernando S Goes 50  Scott D Gordon 29  Shantel Marie Weinsheimer 13, 54  Jürgen Wellmann 101  Gonneke Willemsen 9  Stephanie H Witt 45  Yang Wu 1  Hualin S Xi 112  Jian Yang 2, 113  Futao Zhang 1  Volker Arolt 114  Bernhard T Baune 114, 115, 116  Klaus Berger 101  Dorret I Boomsma 9  Sven Cichon 35, 47, 117, 118  Udo Dannlowski 114  EJC de Geus 9, 119  J Raymond DePaulo 50  Enrico Domenici 120  Katharina Domschke 121, 122  Tõnu Esko 5, 78  Hans J Grabe 109  Steven P Hamilton 123 | Jakob Grove 8, 13, 24, 51  Lynsey S Hall 10, 52  Christine Søholm Hansen 13, 18  Thomas F Hansen 53, 54, 55  Stefan Herms 35, 47  Ian B Hickie 56  Per Hoffmann 35, 47  Georg Homuth 57  Carsten Horn 58  Jouke-Jan Hottenga 9  David M Hougaard 13, 18  David M Howard 10, 28  Marcus Ising 59  Rick Jansen 19  Ian Jones 60  Lisa A Jones 61  Eric Jorgenson 62  James A Knowles 63  Isaac S Kohane 64, 65, 66  Julia Kraft 4  Warren W. Kretzschmar 67  Zoltán Kutalik 68, 69  Yihan Li 67  Penelope A Lind 29  Donald J MacIntyre 70, 71  Dean F MacKinnon 50  Robert M Maier 2  Wolfgang Maier 72  Jonathan Marchini 73  Hamdi Mbarek 9  Patrick McGrath 74  Peter McGuffin 28  Sarah E Medland 29  Divya Mehta 2, 75  Christel M Middeldorp 9, 76, 77  Evelin Mihailov 78  Yuri Milaneschi 19  Lili Milani 78  Francis M Mondimore 50  Grant W Montgomery 1  Sara Mostafavi 79, 80  Niamh Mullins 28  Matthias Nauck 81, 82  Bernard Ng 80  Michel G Nivard 9  Dale R Nyholt 83  Paul F O'Reilly 28  Hogni Oskarsson 84  Caroline Hayward 124  Andrew C Heath 89  Kenneth S Kendler 17  Stefan Kloiber 59, 125, 126  Glyn Lewis 127  Qingqin S Li 128  Susanne Lucae 59  Pamela AF Madden 89  Patrik K Magnusson 22  Nicholas G Martin 29  Andrew M McIntosh 10, 34  Andres Metspalu 78, 129  Ole Mors 13, 130  Preben Bo Mortensen 11, 12, 13, 24  Bertram Müller-Myhsok 15, 131, 132  Merete Nordentoft 13, 133  Markus M Nöthen 35  Michael C O'Donovan 60  Sara A Paciga 134  Nancy L Pedersen 22 | Michael J Owen 60  Jodie N Painter 29  Carsten Bøcker Pedersen 11, 12, 13  Marianne Giørtz Pedersen 11, 12, 13  Roseann E Peterson 17, 85  Wouter J Peyrot 19  Giorgio Pistis 27  Danielle Posthuma 86, 87  Jorge A Quiroz 88  Per Qvist 8, 13, 24  John P Rice 89  Brien P. Riley 17  Margarita Rivera 28, 90  Saira Saeed Mirza 36  Robert Schoevers 91  Eva C Schulte 92, 93  Ling Shen 62  Jianxin Shi 94  Stanley I Shyn 95  Engilbert Sigurdsson 96  Grant C B Sinnamon 97  Johannes H Smit 19  Daniel J Smith 98  Hreinn Stefansson 99  Stacy Steinberg 99  Fabian Streit 45  Jana Strohmaier 45  Katherine E Tansey 100  Henning Teismann 101  Alexander Teumer 102  Wesley Thompson 13, 54, 103, 104  Pippa A Thomson 105  Thorgeir E Thorgeirsson 99  Matthew Traylor 106  Jens Treutlein 45  Vassily Trubetskoy 4  André G Uitterlinden 107  Daniel Umbricht 108  Sandra Van der Auwera 109  Albert M van Hemert 110  Alexander Viktorin 22  Peter M Visscher 1, 2  Yunpeng Wang 13, 54, 104  Bradley T. Webb 111  Brenda WJH Penninx 19  Roy H Perlis 38, 135  David J Porteous 105  James B Potash 136  Martin Preisig 27  Marcella Rietschel 45  Catherine Schaefer 62  Thomas G Schulze 45, 93, 137, 138, 139  Jordan W Smoller 38, 39, 40  Kari Stefansson 99, 140  Henning Tiemeier 36, 141, 142  Rudolf Uher 143  Henry Völzke 102  Myrna M Weissman 74, 144  Thomas Werge 13, 54, 145  Cathryn M Lewis* 28, 146  Douglas F Levinson* 147  Gerome Breen* 28, 148  Anders D Børglum* 8, 13, 24  Patrick F Sullivan* 22, 149, 150 |
| --- | --- | --- |

Affiliations

1, Institute for Molecular Bioscience, The University of Queensland, Brisbane, QLD, AU

2, Queensland Brain Institute, The University of Queensland, Brisbane, QLD, AU

3, Analytic and Translational Genetics Unit, Massachusetts General Hospital, Boston, MA, US

4, Department of Psychiatry and Psychotherapy, Universitätsmedizin Berlin Campus Charité Mitte, Berlin, DE

5, Medical and Population Genetics, Broad Institute, Cambridge, MA, US

6, Department of Psychiatry, Psychosomatics and Psychotherapy, University of Wurzburg, Wurzburg, DE

7, Centre for Psychiatry Research, Department of Clinical Neuroscience, Karolinska Institutet, Stockholm, SE

8, Department of Biomedicine, Aarhus University, Aarhus, DK

9, Dept of Biological Psychology & EMGO+ Institute for Health and Care Research, Vrije Universiteit Amsterdam, Amsterdam, NL

10, Division of Psychiatry, University of Edinburgh, Edinburgh, GB

11, Centre for Integrated Register-based Research, Aarhus University, Aarhus, DK

12, National Centre for Register-Based Research, Aarhus University, Aarhus, DK

13, iPSYCH, The Lundbeck Foundation Initiative for Integrative Psychiatric Research,, DK

14, Discipline of Psychiatry, University of Adelaide, Adelaide, SA, AU

15, Department of Translational Research in Psychiatry, Max Planck Institute of Psychiatry, Munich, DE

16, Department of Neurology, Klinikum rechts der Isar, Technical University of Munich, Munich, DE

17, Department of Psychiatry, Virginia Commonwealth University, Richmond, VA, US

18, Center for Neonatal Screening, Department for Congenital Disorders, Statens Serum Institut, Copenhagen, DK

19, Department of Psychiatry, Vrije Universiteit Medical Center and GGZ inGeest, Amsterdam, NL

20, Virginia Institute for Psychiatric and Behavior Genetics, Richmond, VA, US

21, Department of Psychiatry and Behavioral Sciences, Emory University School of Medicine, Atlanta, GA, US

22, Department of Medical Epidemiology and Biostatistics, Karolinska Institutet, Stockholm, SE

23, Department of Clinical Medicine, Translational Neuropsychiatry Unit, Aarhus University, Aarhus, DK

24, iSEQ, Centre for Integrative Sequencing, Aarhus University, Aarhus, DK

25, Human Genetics, Wellcome Trust Sanger Institute, Cambridge, GB

26, Statistical genomics and systems genetics, European Bioinformatics Institute (EMBL-EBI), Cambridge, GB

27, Department of Psychiatry, Lausanne University Hospital and University of Lausanne, Lausanne, CH

28, Social, Genetic and Developmental Psychiatry Centre, King's College London, London, GB

29, Genetics and Computational Biology, QIMR Berghofer Medical Research Institute, Brisbane, QLD, AU

30, Centre for Advanced Imaging, The University of Queensland, Brisbane, QLD, AU

31, Psychological Medicine, Cardiff University, Cardiff, GB

32, Center for Genomic and Computational Biology, Duke University, Durham, NC, US

33, Department of Pediatrics, Division of Medical Genetics, Duke University, Durham, NC, US

34, Centre for Cognitive Ageing and Cognitive Epidemiology, University of Edinburgh, Edinburgh, GB

35, Institute of Human Genetics, University of Bonn, School of Medicine & University Hospital Bonn, Bonn, DE

36, Epidemiology, Erasmus MC, Rotterdam, Zuid-Holland, NL

37, Psychiatry, Dokuz Eylul University School Of Medicine, Izmir, TR

38, Department of Psychiatry, Massachusetts General Hospital, Boston, MA, US

39, Psychiatric and Neurodevelopmental Genetics Unit (PNGU), Massachusetts General Hospital, Boston, MA, US

40, Stanley Center for Psychiatric Research, Broad Institute, Cambridge, MA, US

41, Neuroscience and Mental Health, Cardiff University, Cardiff, GB

42, Bioinformatics, University of British Columbia, Vancouver, BC, CA

43, Department of Epidemiology, Harvard T.H. Chan School of Public Health, Boston, MA, US

44, Department of Mathematics, Massachusetts Institute of Technology, Cambridge, MA, US

45, Department of Genetic Epidemiology in Psychiatry, Central Institute of Mental Health, Medical Faculty Mannheim, Heidelberg University, Mannheim, Baden-Württemberg, DE

46, Department of Psychiatry (UPK), University of Basel, Basel, CH

47, Department of Biomedicine, University of Basel, Basel, CH

48, Centre for Human Genetics, University of Marburg, Marburg, DE

49, Department of Psychiatry, Trinity College Dublin, Dublin, IE

50, Psychiatry & Behavioral Sciences, Johns Hopkins University, Baltimore, MD, US

51, Bioinformatics Research Centre, Aarhus University, Aarhus, DK

52, Institute of Genetic Medicine, Newcastle University, Newcastle upon Tyne, GB

53, Danish Headache Centre, Department of Neurology, Rigshospitalet, Glostrup, DK

54, Institute of Biological Psychiatry, Mental Health Center Sct. Hans, Mental Health Services Capital Region of Denmark, Copenhagen, DK

55, iPSYCH, The Lundbeck Foundation Initiative for Psychiatric Research, Copenhagen, DK

56, Brain and Mind Centre, University of Sydney, Sydney, NSW, AU

57, Interfaculty Institute for Genetics and Functional Genomics, Department of Functional Genomics, University Medicine and Ernst Moritz Arndt University Greifswald, Greifswald, Mecklenburg-Vorpommern, DE

58, Roche Pharmaceutical Research and Early Development, Pharmaceutical Sciences, Roche Innovation Center Basel, F. Hoffmann-La Roche Ltd, Basel, CH

59, Max Planck Institute of Psychiatry, Munich, DE

60, MRC Centre for Neuropsychiatric Genetics and Genomics, Cardiff University, Cardiff, GB

61, Department of Psychological Medicine, University of Worcester, Worcester, GB

62, Division of Research, Kaiser Permanente Northern California, Oakland, CA, US

63, Psychiatry & The Behavioral Sciences, University of Southern California, Los Angeles, CA, US

64, Department of Biomedical Informatics, Harvard Medical School, Boston, MA, US

65, Department of Medicine, Brigham and Women's Hospital, Boston, MA, US

66, Informatics Program, Boston Children's Hospital, Boston, MA, US

67, Wellcome Trust Centre for Human Genetics, University of Oxford, Oxford, GB

68, Institute of Social and Preventive Medicine (IUMSP), Lausanne University Hospital and University of Lausanne, Lausanne, VD, CH

69, Swiss Institute of Bioinformatics, Lausanne, VD, CH

70, Division of Psychiatry, Centre for Clinical Brain Sciences, University of Edinburgh, Edinburgh, GB

71, Mental Health, NHS 24, Glasgow, GB

72, Department of Psychiatry and Psychotherapy, University of Bonn, Bonn, DE

73, Statistics, University of Oxford, Oxford, GB

74, Psychiatry, Columbia University College of Physicians and Surgeons, New York, NY, US

75, School of Psychology and Counseling, Queensland University of Technology, Brisbane, QLD, AU

76, Child and Youth Mental Health Service, Children's Health Queensland Hospital and Health Service, South Brisbane, QLD, AU

77, Child Health Research Centre, University of Queensland, Brisbane, QLD, AU

78, Estonian Genome Center, University of Tartu, Tartu, EE

79, Medical Genetics, University of British Columbia, Vancouver, BC, CA

80, Statistics, University of British Columbia, Vancouver, BC, CA

81, DZHK (German Centre for Cardiovascular Research), Partner Site Greifswald, University Medicine, University Medicine Greifswald, Greifswald, Mecklenburg-Vorpommern, DE

82, Institute of Clinical Chemistry and Laboratory Medicine, University Medicine Greifswald, Greifswald, Mecklenburg-Vorpommern, DE

83, Institute of Health and Biomedical Innovation, Queensland University of Technology, Brisbane, QLD, AU

84, Humus, Reykjavik, IS

85, Virginia Institute for Psychiatric & Behavioral Genetics, Virginia Commonwealth University, Richmond, VA, US

86, Clinical Genetics, Vrije Universiteit Medical Center, Amsterdam, NL

87, Complex Trait Genetics, Vrije Universiteit Amsterdam, Amsterdam, NL

88, Solid Biosciences, Boston, MA, US

89, Department of Psychiatry, Washington University in Saint Louis School of Medicine, Saint Louis, MO, US

90, Department of Biochemistry and Molecular Biology II, Institute of Neurosciences, Biomedical Research Center (CIBM), University of Granada, Granada, ES

91, Department of Psychiatry, University of Groningen, University Medical Center Groningen, Groningen, NL

92, Department of Psychiatry and Psychotherapy, University Hospital, Ludwig Maximilian University Munich, Munich, DE

93, Institute of Psychiatric Phenomics and Genomics (IPPG), University Hospital, Ludwig Maximilian University Munich, Munich, DE

94, Division of Cancer Epidemiology and Genetics, National Cancer Institute, Bethesda, MD, US

95, Behavioral Health Services, Kaiser Permanente Washington, Seattle, WA, US

96, Faculty of Medicine, Department of Psychiatry, University of Iceland, Reykjavik, IS

97, School of Medicine and Dentistry, James Cook University, Townsville, QLD, AU

98, Institute of Health and Wellbeing, University of Glasgow, Glasgow, GB

99, deCODE Genetics / Amgen, Reykjavik, IS

100, College of Biomedical and Life Sciences, Cardiff University, Cardiff, GB

101, Institute of Epidemiology and Social Medicine, University of Münster, Münster, Nordrhein-Westfalen, DE

102, Institute for Community Medicine, University Medicine Greifswald, Greifswald, Mecklenburg-Vorpommern, DE

103, Department of Psychiatry, University of California, San Diego, San Diego, CA, US

104, KG Jebsen Centre for Psychosis Research, Norway Division of Mental Health and Addiction, Oslo University Hospital, Oslo, NO

105, Medical Genetics Section, CGEM, IGMM, University of Edinburgh, Edinburgh, GB

106, Clinical Neurosciences, University of Cambridge, Cambridge, GB

107, Internal Medicine, Erasmus MC, Rotterdam, Zuid-Holland, NL

108, Roche Pharmaceutical Research and Early Development, Neuroscience, Ophthalmology and Rare Diseases Discovery & Translational Medicine Area, Roche Innovation Center Basel, F. Hoffmann-La Roche Ltd, Basel, CH

109, Department of Psychiatry and Psychotherapy, University Medicine Greifswald, Greifswald, Mecklenburg-Vorpommern, DE

110, Department of Psychiatry, Leiden University Medical Center, Leiden, NL

111, Virginia Institute for Psychiatric & Behavioral Genetics, Virginia Commonwealth University, Richmond, VA, US

112, Computational Sciences Center of Emphasis, Pfizer Global Research and Development, Cambridge, MA, US

113, Institute for Molecular Bioscience; Queensland Brain Institute, The University of Queensland, Brisbane, QLD, AU

114, Department of Psychiatry, University of Münster, Münster, Nordrhein-Westfalen, DE

115, Department of Psychiatry, Melbourne Medical School, University of Melbourne, Melbourne, AU

116, Florey Institute for Neuroscience and Mental Health, University of Melbourne, Melbourne, AU

117, Institute of Medical Genetics and Pathology, University Hospital Basel, University of Basel, Basel, CH

118, Institute of Neuroscience and Medicine (INM-1), Research Center Juelich, Juelich, DE

119, Amsterdam Public Health Institute, Vrije Universiteit Medical Center, Amsterdam, NL

120, Centre for Integrative Biology, Università degli Studi di Trento, Trento, Trentino-Alto Adige, IT

121, Department of Psychiatry and Psychotherapy, Medical Center - University of Freiburg, Faculty of Medicine, University of Freiburg, Freiburg, DE

122, Center for NeuroModulation, Faculty of Medicine, University of Freiburg, Freiburg, DE

123, Psychiatry, Kaiser Permanente Northern California, San Francisco, CA, US

124, Medical Research Council Human Genetics Unit, Institute of Genetics and Molecular Medicine, University of Edinburgh, Edinburgh, GB

125, Department of Psychiatry, University of Toronto, Toronto, ON, CA

126, Centre for Addiction and Mental Health, Toronto, ON, CA

127, Division of Psychiatry, University College London, London, GB

128, Neuroscience Therapeutic Area, Janssen Research and Development, LLC, Titusville, NJ, US

129, Institute of Molecular and Cell Biology, University of Tartu, Tartu, EE

130, Psychosis Research Unit, Aarhus University Hospital, Risskov, Aarhus, DK

131, Munich Cluster for Systems Neurology (SyNergy), Munich, DE

132, University of Liverpool, Liverpool, GB

133, Mental Health Center Copenhagen, Copenhagen Universtity Hospital, Copenhagen, DK

134, Human Genetics and Computational Biomedicine, Pfizer Global Research and Development, Groton, CT, US

135, Psychiatry, Harvard Medical School, Boston, MA, US

136, Psychiatry, University of Iowa, Iowa City, IA, US

137, Department of Psychiatry and Behavioral Sciences, Johns Hopkins University, Baltimore, MD, US

138, Department of Psychiatry and Psychotherapy, University Medical Center Göttingen, Goettingen, Niedersachsen, DE

139, Human Genetics Branch, NIMH Division of Intramural Research Programs, Bethesda, MD, US

140, Faculty of Medicine, University of Iceland, Reykjavik, IS

141, Child and Adolescent Psychiatry, Erasmus MC, Rotterdam, Zuid-Holland, NL

142, Psychiatry, Erasmus MC, Rotterdam, Zuid-Holland, NL

143, Psychiatry, Dalhousie University, Halifax, NS, CA

144, Division of Translational Epidemiology, New York State Psychiatric Institute, New York, NY, US

145, Department of Clinical Medicine, University of Copenhagen, Copenhagen, DK

146, Department of Medical & Molecular Genetics, King's College London, London, GB

147, Psychiatry & Behavioral Sciences, Stanford University, Stanford, CA, US

148, NIHR Maudsley Biomedical Research Centre, King's College London, London, GB

149, Genetics, University of North Carolina at Chapel Hill, Chapel Hill, NC, US

150, Psychiatry, University of North Carolina at Chapel Hill, Chapel Hill, NC, US

**Bipolar Disorder Working Group of the Psychiatric Genomics Consortium**

Niamh Mullins^1,2^, Andreas J. Forstner^3,4,5^, Kevin S. O’Connell^6,7^, Brandon Coombes^8^, Jonathan R. I. Coleman^9,10^, Zhen Qiao^11^, Thomas D. Als^12,13,14^, Tim B. Bigdeli^15,16^, Sigrid Børte^17,18,19^, Julien Bryois^20^, Alexander W. Charney^2^, Ole Kristian Drange^21,22^, Michael J. Gandal^23^, Saskia P. Hagenaars^9,10^, Masashi Ikeda^24^, Nolan Kamitaki^25,26^, Minsoo Kim^23^, Kristi Krebs^27^, Georgia Panagiotaropoulou^28^, Brian M. Schilder^1,29,30,31^, Laura G. Sloofman^1^, Stacy Steinberg^32^, Vassily Trubetskoy^28^, Bendik S. Winsvold^19,33^, Hong-Hee Won^34^, Liliya Abramova^35^, Kristina Adorjan^36,37^, Esben Agerbo^14,38,39^, Mariam Al Eissa^40^, Diego Albani^41^, Ney Alliey-Rodriguez^42,43^, Adebayo Anjorin^44^, Verneri Antilla^45^, Anastasia Antoniou^46^, Swapnil Awasthi^28^, Ji Hyun Baek^47^, Marie Bækvad-Hansen^14,48^, Nicholas Bass^40^, Michael Bauer^49^, Eva C. Beins^3^, Sarah E. Bergen^20^, Armin Birner^50^, Carsten Bøcker Pedersen^14,38,39^, Erlend Bøen^51^, Marco P. Boks^52^, Rosa Bosch^53,54,55,56^, Murielle Brum^57^, Ben M. Brumpton^19^, Nathalie Brunkhorst-Kanaan^57^, Monika Budde^36^, Jonas Bybjerg-Grauholm^14,48^, William Byerley^58^, Murray Cairns^59^, Miquel Casas^53,54,55,56^, Pablo Cervantes^60^, Toni-Kim Clarke^61^, Cristiana Cruceanu^60,62^, Alfredo Cuellar-Barboza^63,64^, Julie Cunningham^65^, David Curtis^66,67^, Piotr M. Czerski^68^, Anders M. Dale^69^, Nina Dalkner^50^, Friederike S. David^3^, Franziska Degenhardt^3,70^, Srdjan Djurovic^71,72^, Amanda L. Dobbyn^1,2^, Athanassios Douzenis^46^, Torbjørn Elvsåshagen^18,73,74^, Valentina Escott-Price^75^, I. Nicol Ferrier^76^, Alessia Fiorentino^40^, Tatiana M. Foroud^77^, Liz Forty^75^, Josef Frank^78^, Oleksandr Frei^6,18^, Nelson B. Freimer^23,79^, Louise Frisén^80^, Katrin Gade^36,81^, Julie Garnham^82^, Joel Gelernter^83,84,85^, Marianne Giørtz Pedersen^14,38,39^, Ian R. Gizer^86^, Scott D. Gordon^87^, Katherine Gordon-Smith^88^, Tiffany A. Greenwood^89^, Jakob Grove^12,13,14,90^, José Guzman-Parra^91^, Kyooseob Ha^92^, Magnus Haraldsson^93^, Martin Hautzinger^94^, Urs Heilbronner^36^, Dennis Hellgren^20^, Stefan Herms^3,95,96^, Per Hoffmann^3,95,96^, Peter A. Holmans^75^, Laura Huckins^1,2^, Stéphane Jamain^97,98^, Jessica S. Johnson^1,2^, Janos L. Kalman^36,37,99^, Yoichiro Kamatani^100,101^, James L. Kennedy^102,103,104,105^, Sarah Kittel-Schneider^57,106^, James A. Knowles^107,108^, Manolis Kogevinas^109^, Maria Koromina^110^, Thorsten M. Kranz^57^, Henry R. Kranzler^111,112^, Michiaki Kubo^113^, Ralph Kupka^114,115,116^, Steven A. Kushner^117^, Catharina Lavebratt^118,119^, Jacob Lawrence^120^, Markus Leber^121^, Heon-Jeong Lee^122^, Phil H. Lee^123^, Shawn E. Levy^124^, Catrin Lewis^75^, Calwing Liao^125,126^, Susanne Lucae^62^, Martin Lundberg^118,119^, Donald J. MacIntyre^127^, Sigurdur H. Magnusson^32^, Wolfgang Maier^128^, Adam Maihofer^89^, Dolores Malaspina^1,2^, Eirini Maratou^129^, Lina Martinsson^80^, Manuel Mattheisen^12,13,14,106,130^, Nathaniel W. McGregor^131^, Peter McGuffin^9^, James D. McKay^132^, Helena Medeiros^108^, Sarah E. Medland^87^, Vincent Millischer^118,119^, Grant W. Montgomery^11^, Jennifer L. Moran^25,133^, Derek W. Morris^134^, Thomas W. Mühleisen^4,95^, Niamh O’Brien^40^, Claire O’Donovan^82^, Loes M. Olde Loohuis^23,79^, Lilijana Oruc^135^, Sergi Papiol^36,37^, Antonio F. Pardiñas^75^, Amy Perry^88^, Andrea Pfennig^49^, Evgenia Porichi^46^, James B. Potash^136^, Digby Quested^137,138^, Towfique Raj^1,29,30,31^, Mark H. Rapaport^139^, J. Raymond DePaulo^136^, Eline J. Regeer^140^, John P. Rice^141^, Fabio Rivas^91^, Margarita Rivera^142,143^, Julian Roth^106^, Panos Roussos^1,2,29^, Douglas M. Ruderfer^144^, Cristina Sánchez-Mora^53,54,56,145^, Eva C. Schulte^36,37^, Fanny Senner^36,37^, Sally Sharp^40^, Paul D. Shilling^89^, Engilbert Sigurdsson^93,146^, Lea Sirignano^78^, Claire Slaney^82^, Olav B. Smeland^6,7^, Daniel J. Smith^147^, Janet L. Sobell^148^, Christine Søholm Hansen^14,48^, Maria Soler Artigas^53,54,56,145^, Anne T. Spijker^149^, Dan J. Stein^150^, John S. Strauss^102^, Beata Świątkowska^151^, Chikashi Terao^101^, Thorgeir E. Thorgeirsson^32^, Claudio Toma^152,153,154^, Paul Tooney^59^, Evangelia-Eirini Tsermpini^110^, Marquis P. Vawter^155^, Helmut Vedder^156^, James T. R. Walters^75^, Stephanie H. Witt^78^, Simon Xi^157^, Wei Xu^158^, Jessica Mei Kay Yang^75^, Allan H. Young^159,160^, Hannah Young^1^, Peter P. Zandi^136^, Hang Zhou^83,84^, Lea Zillich^78^, HUNT All-In Psychiatry^161^, Rolf Adolfsson^162^, Ingrid Agartz^51,130,163^, Martin Alda^82,164^, Lars Alfredsson^165^, Gulja Babadjanova^166^, Lena Backlund^118,119^, Bernhard T. Baune^167,168,169^, Frank Bellivier^170,171^, Susanne Bengesser^50^, Wade H. Berrettini^172^, Douglas H. R. Blackwood^61^, Michael Boehnke^173^, Anders D. Børglum^14,174,175^, Gerome Breen^9,10^, Vaughan J. Carr^176^, Stanley Catts^177^, Aiden Corvin^178^, Nicholas Craddock^75^, Udo Dannlowski^167^, Dimitris Dikeos^179^, Tõnu Esko^26,27,180,181^, Bruno Etain^170,171^, Panagiotis Ferentinos^9,46^, Mark Frye^64^, Janice M. Fullerton^152,153^, Micha Gawlik^106^, Elliot S. Gershon^42,182^, Fernando S. Goes^136^, Melissa J. Green^152,176^, Maria Grigoroiu-Serbanescu^183^, Joanna Hauser^68^, Frans Henskens^59^, Jan Hillert^80^, Kyung Sue Hong^47^, David M. Hougaard^14,48^, Christina M. Hultman^20^, Kristian Hveem^19,184^, Nakao Iwata^24^, Assen V. Jablensky^185^, Ian Jones^75^, Lisa A. Jones^88^, René S. Kahn^2,52^, John R. Kelsoe^89^, George Kirov^75^, Mikael Landén^20,186^, Marion Leboyer^97,98,187^, Cathryn M. Lewis^9,10,188^, Qingqin S. Li^189^, Jolanta Lissowska^190^, Christine Lochner^191^, Carmel Loughland^59^, Nicholas G. Martin^87,192^, Carol A. Mathews^193^, Fermin Mayoral^91^, Susan L. McElroy^194^, Andrew M. McIntosh^127,195^, Francis J. McMahon^196^, Ingrid Melle^6,197^, Patricia Michie^59^, Lili Milani^27^, Philip B. Mitchell^176^, Gunnar Morken^21,198^, Ole Mors^14,199^, Preben Bo Mortensen^12,14,38,39^, Bryan Mowry^177^, Bertram Müller-Myhsok^62,200,201^, Richard M. Myers^124^, Benjamin M. Neale^25,45,180^, Caroline M. Nievergelt^89,202^, Merete Nordentoft^14,203^, Markus M. Nöthen^3^, Michael C. O’Donovan^75^, Ketil J. Oedegaard^204,205^, Tomas Olsson^206^, Michael J. Owen^75^, Sara A. Paciga^207^, Chris Pantelis^208^, Carlos Pato^108^, Michele T. Pato^108^, George P. Patrinos^110,209,210^, Roy H. Perlis^211,212^, Danielle Posthuma^213,214^, Josep Antoni Ramos-Quiroga^53,54,55,56^, Andreas Reif^57^, Eva Z. Reininghaus^50^, Marta Ribasés^53,54,56,145^, Marcella Rietschel^78^, Stephan Ripke^25,28,45^, Guy A. Rouleau^126,215^, Takeo Saito^24^, Ulrich Schall^59^, Martin Schalling^118,119^, Peter R. Schofield^152,153^, Thomas G. Schulze^36,78,81,136,216^, Laura J. Scott^173^, Rodney J. Scott^59^, Alessandro Serretti^217^, Cynthia Shannon Weickert^152,176,218^, Jordan W. Smoller^25,133,219^, Hreinn Stefansson^32^, Kari Stefansson^32,220^, Eystein Stordal^221,222^, Fabian Streit^78^, Patrick F. Sullivan^20,223,224^, Gustavo Turecki^225^, Arne E. Vaaler^226^, Eduard Vieta^227^, John B. Vincent^102^, Irwin D. Waldman^228^, Thomas W. Weickert^152,176,218^, Thomas Werge^14,229,230,231^, Naomi R. Wray^11,232^, John-Anker Zwart^18,19,33^, Joanna M. Biernacka^8,64^, John I. Nurnberger^233^, Sven Cichon^3,4,95,96^, Howard J. Edenberg^77,234^, Eli A. Stahl^1,2,180^, Andrew McQuillin^40^, Arianna Di Florio^75,224^, Roel A. Ophoff^23,79,117,235^, Ole A. Andreassen^6,7^

**Affiliations**

^1^Department of Genetics and Genomic Sciences, Icahn School of Medicine at Mount Sinai, New York, NY, USA. ^2^Department of Psychiatry, Icahn School of Medicine at Mount Sinai, New York, NY, USA. ^3^Institute of Human Genetics, University of Bonn, School of Medicine and University Hospital Bonn, Bonn, Germany. ^4^Institute of Neuroscience and Medicine (INM-1), Research Centre Jülich, Jülich, Germany. ^5^Centre for Human Genetics, University of Marburg, Marburg, Germany. ^6^Division of Mental Health and Addiction, Oslo University Hospital, Oslo, Norway. ^7^NORMENT, University of Oslo, Oslo, Norway. ^8^Department of Health Sciences Research, Mayo Clinic, Rochester, MN, USA. ^9^Social, Genetic and Developmental Psychiatry Centre, King’s College London, London, UK. ^10^NIHR Maudsley BRC, King’s College London, London, UK. ^11^Institute for Molecular Bioscience, The University of Queensland, Brisbane, QLD, Australia. ^12^iSEQ, Center for Integrative Sequencing, Aarhus University, Aarhus, Denmark. ^13^Department of Biomedicine - Human Genetics, Aarhus University, Aarhus, Denmark. ^14^iPSYCH, The Lundbeck Foundation Initiative for Integrative Psychiatric Research, Denmark. ^15^Department of Psychiatry and Behavioral Sciences, SUNY Downstate Health Sciences University, Brooklyn, NY, USA. ^16^VA NY Harbor Healthcare System, Brooklyn, NY, USA. ^17^Research and Communication Unit for Musculoskeletal Health, Division of Clinical Neuroscience, Oslo University Hospital, Ullevål, Oslo, Norway. ^18^Institute of Clinical Medicine, University of Oslo, Oslo, Norway. ^19^K. G. Jebsen Center for Genetic Epidemiology, Department of Public Health and Nursing, Faculty of Medicine and Health Sciences, Norwegian University of Science and Technology, Trondheim, Norway. ^20^Department of Medical Epidemiology and Biostatistics, Karolinska Institutet, Stockholm, Sweden. ^21^Department of Mental Health, Faculty of Medicine and Health Sciences, Norwegian University of Science and Technology (NTNU), Trondheim, Norway. ^22^Department of Østmarka, Division of Mental Health Care, St. Olavs Hospital, Trondheim University Hospital, Trondheim, Norway. ^23^Department of Psychiatry and Biobehavioral Science, Semel Institute, David Geffen School of Medicine, University of California, Los Angeles, Los Angeles, CA, USA. ^24^Department of Psychiatry, Fujita Health University School of Medicine, Toyoake, Japan. ^25^Stanley Center for Psychiatric Research, Broad Institute, Cambridge, MA, USA. ^26^Department of Genetics, Harvard Medical School, Boston, MA, USA. ^27^Estonian Genome Center, Institute of Genomics, University of Tartu, Tartu, Estonia. ^28^Department of Psychiatry and Psychotherapy, Charité - Universitätsmedizin, Berlin, Germany. ^29^Department of Neuroscience, Icahn School of Medicine at Mount Sinai, New York, NY, USA. ^30^Ronald M. Loeb Center for Alzheimer’s Disease, Icahn School of Medicine at Mount Sinai, New York, NY, USA. ^31^Estelle and Daniel Maggin Department of Neurology, Icahn School of Medicine at Mount Sinai, New York, NY, USA. ^32^deCODE Genetics / Amgen, Reykjavik, Iceland. ^33^Department of Research, Innovation and Education, Division of Clinical Neuroscience, Oslo University Hospital, Oslo, Norway. ^34^Samsung Advanced Institute for Health Sciences and Technology (SAIHST), Sungkyunkwan University, Samsung Medical Center, Seoul, South Korea. ^35^Russian Academy of Medical Sciences, Mental Health Research Center, Moscow, Russian Federation. ^36^Institute of Psychiatric Phenomics and Genomics (IPPG), University Hospital, LMU Munich, Munich, Germany. ^37^Department of Psychiatry and Psychotherapy, University Hospital, LMU Munich, Munich, Germany. ^38^National Centre for Register-Based Research, Aarhus University, Aarhus, Denmark. ^39^Centre for Integrated Register-based Research, Aarhus University, Aarhus, Denmark. ^40^Division of Psychiatry, University College London, London, UK. ^41^Department of Neuroscience, Istituto Di Ricerche Farmacologiche Mario Negri IRCCS, Milano, Italy. ^42^Department of Psychiatry and Behavioral Neuroscience, University of Chicago, Chicago, IL, USA. ^43^Northwestern University, Chicago, IL, USA. ^44^Psychiatry, Berkshire Healthcare NHS Foundation Trust, Bracknell, UK. ^45^Analytic and Translational Genetics Unit, Massachusetts General Hospital, Boston, MA, USA. ^46^National and Kapodistrian University of Athens, 2nd Department of Psychiatry, Attikon General Hospital, Athens, Greece. ^47^Department of Psychiatry, Sungkyunkwan University School of Medicine, Samsung Medical Center, Seoul, South Korea. ^48^Center for Neonatal Screening, Department for Congenital Disorders, Statens Serum Institut, Copenhagen, Denmark. ^49^Department of Psychiatry and Psychotherapy, University Hospital Carl Gustav Carus, Technische Universität Dresden, Dresden, Germany. ^50^Medical University of Graz, Department of Psychiatry and Psychotherapeutic Medicine, Graz, Austria. ^51^Department of Psychiatric Research, Diakonhjemmet Hospital, Oslo, Norway. ^52^Psychiatry, Brain Center UMC Utrecht, Utrecht, The Netherlands. ^53^Instituto de Salud Carlos III, Biomedical Network Research Centre on Mental Health (CIBERSAM), Madrid, Spain. ^54^Department of Psychiatry, Hospital Universitari Vall d´Hebron, Barcelona, Spain. ^55^Department of Psychiatry and Forensic Medicine, Universitat Autònoma de Barcelona, Barcelona, Spain. ^56^Psychiatric Genetics Unit, Group of Psychiatry Mental Health and Addictions, Vall d´Hebron Research Institut (VHIR), Universitat Autònoma de Barcelona, Barcelona, Spain. ^57^Department of Psychiatry, Psychosomatic Medicine and Psychotherapy, University Hospital Frankfurt, Frankfurt am Main, Germany. ^58^Psychiatry, University of California San Francisco, San Francisco, CA, USA. ^59^University of Newcastle, Newcastle, NSW, Australia. ^60^Department of Psychiatry, Mood Disorders Program, McGill University Health Center, Montreal, QC, Canada. ^61^Division of Psychiatry, University of Edinburgh, Edinburgh, UK. ^62^Department of Translational Research in Psychiatry, Max Planck Institute of Psychiatry, Munich, Germany. ^63^Department of Psychiatry, Universidad Autonoma de Nuevo Leon, Monterrey, Mexico. ^64^Department of Psychiatry and Psychology, Mayo Clinic, Rochester, MN, USA. ^65^Department of Laboratory Medicine and Pathology, Mayo Clinic, Rochester, MN, USA. ^66^Centre for Psychiatry, Queen Mary University of London, London, UK. ^67^UCL Genetics Institute, University College London, London, UK. ^68^Department of Psychiatry, Laboratory of Psychiatric Genetics, Poznan University of Medical Sciences, Poznan, Poland. ^69^Center for Multimodal Imaging and Genetics, Departments of Neurosciences, Radiology, and Psychiatry, University of California, San Diego, CA, USA. ^70^Department of Child and Adolescent Psychiatry, Psychosomatics and Psychotherapy, University Hospital Essen, University of Duisburg-Essen, Duisburg, Germany. ^71^Department of Medical Genetics, Oslo University Hospital Ullevål, Oslo, Norway. ^72^NORMENT, Department of Clinical Science, University of Bergen, Bergen, Norway. ^73^Department of Neurology, Oslo University Hospital, Oslo, Norway. ^74^NORMENT, KG Jebsen Centre for Psychosis Research, Oslo University Hospital, Oslo, Norway. ^75^Medical Research Council Centre for Neuropsychiatric Genetics and Genomics, Division of Psychological Medicine and Clinical Neurosciences, Cardiff University, Cardiff, UK. ^76^Academic Psychiatry, Newcastle University, Newcastle upon Tyne, UK. ^77^Department of Medical and Molecular Genetics, Indiana University, Indianapolis, IN, USA. ^78^Department of Genetic Epidemiology in Psychiatry, Central Institute of Mental Health, Medical Faculty Mannheim, Heidelberg University, Mannheim, Germany. ^79^Center for Neurobehavioral Genetics, Semel Institute for Neuroscience and Human Behavior, Los Angeles, CA, USA. ^80^Department of Clinical Neuroscience, Karolinska Institutet, Stockholm, Sweden. ^81^Department of Psychiatry and Psychotherapy, University Medical Center Göttingen, Göttingen, Germany. ^82^Department of Psychiatry, Dalhousie University, Halifax, NS, Canada. ^83^Department of Psychiatry, Yale School of Medicine, New Haven, CT, USA. ^84^Veterans Affairs Connecticut Healthcare System, West Haven, CT, USA. ^85^Departments of Genetics and Neuroscience, Yale University School of Medicine, New Haven, CT, USA. ^86^Department of Psychological Sciences, University of Missouri, Columbia, MO, USA. ^87^Genetics and Computational Biology, QIMR Berghofer Medical Research Institute, Brisbane, QLD, Australia. ^88^Psychological Medicine, University of Worcester, Worcester, UK. ^89^Department of Psychiatry, University of California San Diego, La Jolla, CA, USA. ^90^Bioinformatics Research Centre, Aarhus University, Aarhus, Denmark. ^91^Mental Health Department, University Regional Hospital, Biomedicine Institute (IBIMA), Málaga, Spain. ^92^Department of Psychiatry, Seoul National University College of Medicine, Seoul, South Korea. ^93^Landspitali University Hospital, Reykjavik, Iceland. ^94^Department of Psychology, Eberhard Karls Universität Tübingen, Tubingen, Germany. ^95^Department of Biomedicine, University of Basel, Basel, Switzerland. ^96^Institute of Medical Genetics and Pathology, University Hospital Basel, Basel, Switzerland. ^97^Neuropsychiatrie Translationnelle, Inserm U955, Créteil, France. ^98^Faculté de Santé, Université Paris Est, Créteil, France. ^99^International Max Planck Research School for Translational Psychiatry (IMPRS-TP), Munich, Germany. ^100^Laboratory of Complex Trait Genomics, Department of Computational Biology and Medical Sciences, Graduate School of Frontier Sciences, The University of Tokyo, Tokyo, Japan. ^101^Laboratory for Statistical and Translational Genetics, RIKEN Center for Integrative Medical Sciences, Yokohama, Japan. ^102^Campbell Family Mental Health Research Institute, Centre for Addiction and Mental Health, Toronto, ON, Canada. ^103^Neurogenetics Section, Centre for Addiction and Mental Health, Toronto, ON, Canada. ^104^Department of Psychiatry, University of Toronto, Toronto, ON, Canada. ^105^Institute of Medical Sciences, University of Toronto, Toronto, ON, Canada. ^106^Department of Psychiatry, Psychosomatics and Psychotherapy, Center of Mental Health, University Hospital Würzburg, Würzburg, Germany. ^107^Cell Biology, SUNY Downstate Medical Center College of Medicine, Brooklyn, NY, USA. ^108^Institute for Genomic Health, SUNY Downstate Medical Center College of Medicine, Brooklyn, NY, USA. ^109^ISGlobal, Barcelona, Spain. ^110^University of Patras, School of Health Sciences, Department of Pharmacy, Laboratory of Pharmacogenomics and Individualized Therapy, Patras, Greece. ^111^Mental Illness Research, Education and Clinical Center, Crescenz VAMC, Philadelphia, PA, USA. ^112^Center for Studies of Addiction, University of Pennsylvania Perelman School of Medicine, Philadelphia, PA, USA. ^113^RIKEN Center for Integrative Medical Sciences, Yokohama, Japan. ^114^Psychiatry, Altrecht, Utrecht, The Netherlands. ^115^Psychiatry, GGZ inGeest, Amsterdam, The Netherlands. ^116^Psychiatry, VU medisch centrum, Amsterdam, The Netherlands. ^117^Department of Psychiatry, Erasmus MC, University Medical Center Rotterdam, Rotterdam, The Netherlands. ^118^Department of Molecular Medicine and Surgery, Karolinska Institutet, Stockholm, Sweden. ^119^Center for Molecular Medicine, Karolinska University Hospital, Stockholm, Sweden. ^120^Psychiatry, North East London NHS Foundation Trust, Ilford, UK. ^121^Clinic for Psychiatry and Psychotherapy, University Hospital Cologne, Cologne, Germany. ^122^Department of Psychiatry, Korea University College of Medicine, Seoul, South Korea. ^123^Psychiatric and Neurodevelopmental Genetics Unit, Center for Genomic Medicine, Massachusetts General Hospital and Harvard Medical School, Boston, MA, USA. ^124^HudsonAlpha Institute for Biotechnology, Huntsville, AL, USA. ^125^Department of Human Genetics, McGill University, Montréal, QC, Canada. ^126^Montreal Neurological Institute and Hospital, McGill University, Montréal, QC, Canada. ^127^Division of Psychiatry, Centre for Clinical Brain Sciences, The University of Edinburgh, Edinburgh, UK. ^128^Department of Psychiatry and Psychotherapy, University of Bonn, Bonn, Germany. ^129^National and Kapodistrian University of Athens, Medical School, Clinical Biochemistry Laboratory, Attikon General Hospital, Athens, Greece. ^130^Department of Clinical Neuroscience, Centre for Psychiatry Research, Karolinska Institutet, Stockholm, Sweden. ^131^Systems Genetics Working Group, Department of Genetics, Stellenbosch University, Stellenbosch, South Africa. ^132^Genetic Cancer Susceptibility Group, International Agency for Research on Cancer, Lyon, France. ^133^Department of Psychiatry, Massachusetts General Hospital, Boston, MA, USA. ^134^Centre for Neuroimaging and Cognitive Genomics (NICOG), National University of Ireland Galway, Galway, Ireland. ^135^Medical faculty, University Sarajevo School of Science and Technology, Sarajevo, Bosnia and Herzegovina. ^136^Department of Psychiatry and Behavioral Sciences, Johns Hopkins University School of Medicine, Baltimore, MD, USA. ^137^Oxford Health NHS Foundation Trust, Warneford Hospital, Oxford, UK. ^138^Department of Psychiatry, University of Oxford, Warneford Hospital, Oxford, UK. ^139^Department of Psychiatry and Behavioral Sciences, Emory University School of Medicine, Atlanta, GA, USA. ^140^Outpatient Clinic for Bipolar Disorder, Altrecht, Utrecht, The Netherlands. ^141^Department of Psychiatry, Washington University in Saint Louis, Saint Louis, MO, USA. ^142^Department of Biochemistry and Molecular Biology II, Faculty of Pharmacy, University of Granada, Spain. ^143^Institute of Neurosciences, Biomedical Research Center (CIBM), University of Granada, Spain. ^144^Medicine, Psychiatry, Biomedical Informatics, Vanderbilt University Medical Center, Nashville, TN, USA. ^145^Department of Genetics, Microbiology and Statistics, Faculty of Biology, Universitat de Barcelona, Barcelona, Catalonia, Spain. ^146^Faculty of Medicine, Department of Psychiatry, School of Health Sciences, University of Iceland, Reykjavik, Iceland. ^147^Institute of Health and Wellbeing, University of Glasgow, Glasgow, UK. ^148^Psychiatry and the Behavioral Sciences, University of Southern California, Los Angeles, CA, USA. ^149^Mood Disorders, PsyQ, Rotterdam, The Netherlands. ^150^SAMRC Unit on Risk and Resilience in Mental Disorders, Dept of Psychiatry and Neuroscience Institute, University of Cape Town, Cape Town, South Africa. ^151^Department of Environmental Epidemiology, Nofer Institute of Occupational Medicine, Lodz, Poland. ^152^Neuroscience Research Australia, Sydney, NSW, Australia. ^153^School of Medical Sciences, University of New South Wales, Sydney, NSW, Australia. ^154^Centro de Biología Molecular Severo Ochoa, Universidad Autónoma de Madrid and CSIC, Madrid, Spain. ^155^Department of Psychiatry and Human Behavior, School of Medicine, University of California, Irvine, CA, USA. ^156^Psychiatry, Psychiatrisches Zentrum Nordbaden, Wiesloch, Germany. ^157^Computational Sciences Center of Emphasis, Pfizer Global Research and Development, Cambridge, MA, USA. ^158^Dalla Lana School of Public Health, University of Toronto, Toronto, ON, Canada. ^159^Department of Psychological Medicine, Institute of Psychiatry, Psychology and Neuroscience, King’s College London, London, UK. ^160^South London and Maudsley NHS Foundation Trust, Bethlem Royal Hospital, Monks Orchard Road, Beckenham, Kent, UK. ^161^A list of members and affiliations appears in the Supplementary Note. ^162^Department of Clinical Sciences, Psychiatry, Umeå University Medical Faculty, Umeå, Sweden. ^163^NORMENT, KG Jebsen Centre for Psychosis Research, Division of Mental Health and Addiction, Institute of Clinical Medicine and Diakonhjemmet Hospital, University of Oslo, Oslo, Norway. ^164^National Institute of Mental Health, Klecany, Czech Republic. ^165^Institute of Environmental Medicine, Karolinska Institutet, Stockholm, Sweden. ^166^Institute of Pulmonology, Russian State Medical University, Moscow, Russian Federation. ^167^Department of Psychiatry, University of Münster, Münster, Germany. ^168^Department of Psychiatry, Melbourne Medical School, The University of Melbourne, Melbourne, VIC, Australia. ^169^The Florey Institute of Neuroscience and Mental Health, The University of Melbourne, Parkville, VIC, Australia. ^170^Université de Paris, INSERM, Optimisation Thérapeutique en Neuropsychopharmacologie, UMRS-1144, Paris, France. ^171^APHP Nord, DMU Neurosciences, GHU Saint Louis-Lariboisière-Fernand Widal, Département de Psychiatrie et de Médecine Addictologique, Paris, France. ^172^Psychiatry, University of Pennsylvania, Philadelphia, PA, USA. ^173^Center for Statistical Genetics and Department of Biostatistics, University of Michigan, Ann Arbor, MI, USA. ^174^Department of Biomedicine and the iSEQ Center, Aarhus University, Aarhus, Denmark. ^175^Center for Genomics and Personalized Medicine, CGPM, Aarhus, Denmark. ^176^School of Psychiatry, University of New South Wales, Sydney, NSW, Australia. ^177^University of Queensland, Brisbane, QLD, Australia. ^178^Neuropsychiatric Genetics Research Group, Dept of Psychiatry and Trinity Translational Medicine Institute, Trinity College Dublin, Dublin, Ireland. ^179^National and Kapodistrian University of Athens, 1st Department of Psychiatry, Eginition Hospital, Athens, Greece. ^180^Medical and Population Genetics, Broad Institute, Cambridge, MA, USA. ^181^Division of Endocrinology, Children’s Hospital Boston, Boston, MA, USA. ^182^Department of Human Genetics, University of Chicago, Chicago, IL, USA. ^183^Biometric Psychiatric Genetics Research Unit, Alexandru Obregia Clinical Psychiatric Hospital, Bucharest, Romania. ^184^HUNT Research Center, Department of Public Health and Nursing, Faculty of Medicine and Health Sciences, Norwegian University of Science and Technology, Trondheim, Norway. ^185^University of Western Australia, Nedlands, WA, Australia. ^186^Institute of Neuroscience and Physiology, University of Gothenburg, Gothenburg, Sweden. ^187^Department of Psychiatry and Addiction Medicine, Assistance Publique - Hôpitaux de Paris, Paris, France. ^188^Department of Medical and Molecular Genetics, King’s College London, London, UK. ^189^Neuroscience Therapeutic Area, Janssen Research and Development, LLC, Titusville, NJ, USA. ^190^Cancer Epidemiology and Prevention, M. Sklodowska-Curie National Research Institute of Oncology, Warsaw, Poland. ^191^SA MRC Unit on Risk and Resilience in Mental Disorders, Dept of Psychiatry, Stellenbosch University, Stellenbosch, South Africa. ^192^School of Psychology, The University of Queensland, Brisbane, QLD, Australia. ^193^Department of Psychiatry and Genetics Institute, University of Florida, Gainesville, FL, USA. ^194^Research Institute, Lindner Center of HOPE, Mason, OH, USA. ^195^Centre for Cognitive Ageing and Cognitive Epidemiology, University of Edinburgh, Edinburgh, UK. ^196^Human Genetics Branch, Intramural Research Program, National Institute of Mental Health, Bethesda, MD, USA. ^197^Division of Mental Health and Addiction, University of Oslo, Institute of Clinical Medicine, Oslo, Norway. ^198^Psychiatry, St Olavs University Hospital, Trondheim, Norway. ^199^Psychosis Research Unit, Aarhus University Hospital - Psychiatry, Risskov, Denmark. ^200^Munich Cluster for Systems Neurology (SyNergy), Munich, Germany. ^201^University of Liverpool, Liverpool, UK. ^202^Research/Psychiatry, Veterans Affairs San Diego Healthcare System, San Diego, CA, USA. ^203^Mental Health Services in the Capital Region of Denmark, Mental Health Center Copenhagen, University of Copenhagen, Copenhagen, Denmark. ^204^Division of Psychiatry, Haukeland Universitetssjukehus, Bergen, Norway. ^205^Faculty of Medicine and Dentistry, University of Bergen, Bergen, Norway. ^206^Department of Clinical Neuroscience and Center for Molecular Medicine, Karolinska Institutet at Karolinska University Hospital, Solna, Sweden. ^207^Human Genetics and Computational Biomedicine, Pfizer Global Research and Development, Groton, CT, USA. ^208^University of Melbourne, VIC, Australia. ^209^United Arab Emirates University, College of Medicine and Health Sciences, Department of Pathology, Al-Ain, United Arab Emirates. ^210^United Arab Emirates University, Zayed Center of Health Sciences, Al-Ain, United Arab Emirates. ^211^Psychiatry, Harvard Medical School, Boston, MA, USA. ^212^Division of Clinical Research, Massachusetts General Hospital, Boston, MA, USA. ^213^Department of Complex Trait Genetics, Center for Neurogenomics and Cognitive Research, Amsterdam Neuroscience, Vrije Universiteit Amsterdam, Amsterdam, The Netherlands. ^214^Department of Clinical Genetics, Amsterdam Neuroscience, Vrije Universiteit Medical Center, Amsterdam, The Netherlands. ^215^Department of Neurology and Neurosurgery, McGill University, Faculty of Medicine, Montreal, QC, Canada. ^216^Department of Psychiatry and Behavioral Sciences, SUNY Upstate Medical University, Syracuse, NY, USA. ^217^Department of Biomedical and NeuroMotor Sciences, University of Bologna, Bologna, Italy. ^218^Department of Neuroscience, SUNY Upstate Medical University, Syracuse, NY, USA. ^219^Psychiatric and Neurodevelopmental Genetics Unit (PNGU), Massachusetts General Hospital, Boston, MA, USA. ^220^Faculty of Medicine, University of Iceland, Reykjavik, Iceland. ^221^Department of Psychiatry, Hospital Namsos, Namsos, Norway. ^222^Department of Neuroscience, Norges Teknisk Naturvitenskapelige Universitet Fakultet for naturvitenskap og teknologi, Trondheim, Norway. ^223^Department of Genetics, University of North Carolina at Chapel Hill, Chapel Hill, NC, USA. ^224^Department of Psychiatry, University of North Carolina at Chapel Hill, Chapel Hill, NC, USA. ^225^Department of Psychiatry, McGill University, Montreal, QC, Canada. ^226^Dept of Psychiatry, Sankt Olavs Hospital Universitetssykehuset i Trondheim, Trondheim, Norway. ^227^Clinical Institute of Neuroscience, Hospital Clinic, University of Barcelona, IDIBAPS, CIBERSAM, Barcelona, Spain. ^228^Department of Psychology, Emory University, Atlanta, GA, USA. ^229^Institute of Biological Psychiatry, Mental Health Services, Copenhagen University Hospital, Copenhagen, Denmark. ^230^Department of Clinical Medicine, University of Copenhagen, Copenhagen, Denmark. ^231^Center for GeoGenetics, GLOBE Institute, University of Copenhagen, Copenhagen, Denmark. ^232^Queensland Brain Institute, The University of Queensland, Brisbane, QLD, Australia. ^233^Psychiatry, Indiana University School of Medicine, Indianapolis, IN, USA. ^234^Biochemistry and Molecular Biology, Indiana University School of Medicine, Indianapolis, IN, USA. ^235^Department of Human Genetics, David Geffen School of Medicine, University of California Los Angeles, Los Angeles, CA, USA.

**Eating Disorders Working Group of the Psychiatric Genomics Consortium**

| Roger AH Adan ^1, 2, 3^  Lars Alfredsson ^4^  Tetsuya Ando ^5^  Ole A Andreassen ^6^  Harald Aschauer ^7^  Jessica H Baker ^8^  Vladimir Bencko ^9^  Andrew W Bergen ^10, 11^  Wade H Berrettini ^12^  Andreas Birgegård ^13, 14, 15^  Joseph M Boden ^16^  Ilka Boehm ^17^  Claudette Boni ^18^  Vesna Boraska Perica ^19, 20^  Harry Brandt ^21^  Gerome Breen ^22, 23^  Julien Bryois ^15^  Katharina Buehren ^24^  Cynthia M Bulik ^8, 15, 25^  Roland Burghardt ^26^  Laura Carlberg ^27^  Matteo Cassina ^28^  Sven Cichon ^29, 30, 31^  Maurizio Clementi ^28^  Jonathan RI Coleman ^22, 23^  Roger D Cone ^32^  Philippe Courtet ^33^  Steven Crawford ^21^  Scott Crow ^34^  James J Crowley ^13, 35^  Unna N Danner ^2^  Oliver SP Davis ^36, 37^  Martina de Zwaan ^38^  George Dedoussis ^39^  Daniela Degortes ^40^  Janiece E DeSocio ^41^  Danielle M Dick ^42, 43, 44^  Dimitris Dikeos ^45^  Christian Dina ^46^  Monika Dmitrzak-Weglarz ^47^  Elisa Docampo Martinez ^48, 49, 50^  Laramie E Duncan ^51^  Karin Egberts^52^  Christian R Marshall ^126^  Nicholas G Martin ^72^  Manuel Mattheisen ^13, 14, 75, 127^  Morten Mattingsdal ^6^  Sara McDevitt ^128, 129^  Peter McGuffin ^22^  Sarah E Medland ^72^  Andres Metspalu ^53, 130^  Ingrid Meulenbelt ^131^  Nadia Micali ^132, 133^  James Mitchell ^134^  Karen Mitchell ^135, 136^  Palmiero Monteleone ^137^  Alessio Maria Monteleone ^124^  Grant W Montgomery ^72, 86, 138^  Preben Bo Mortensen ^76, 114, 115^  Melissa A Munn-Chernoff ^8^  Benedetta Nacmias ^139^  Marie Navratilova ^63^  Ioanna Ntalla ^39^  Catherine M Olsen ^140^  Roel A Ophoff ^141, 142^  Julie K O'Toole ^143^  Leonid Padyukov ^110^  Aarno Palotie ^54, 102, 144^  Jacques Pantel ^18^  Hana Papezova ^97^  Richard Parker ^72^  John F Pearson ^145^  Nancy L Pedersen ^15^ | Stefan Ehrlich ^17^  Geòrgia Escaramís ^48, 49, 50^  Tõnu Esko ^53, 54^  Thomas Espeseth ^55^  Xavier Estivill ^48, 49, 50, 56^  Anne Farmer ^22^  Angela Favaro ^40^  Fernando Fernández-Aranda ^57, 58^  Manfred M Fichter ^59, 60^  Krista Fischer ^53^  James AB Floyd ^61^  Manuel Föcker ^62^  Lenka Foretova ^63^  Andreas J Forstner ^30, 64, 65, 66^  Monica Forzan ^28^  Christopher S Franklin ^19^  Steven Gallinger ^67^  Giovanni Gambaro ^68^  Héléna A Gaspar ^22, 23^  Ina Giegling ^69^  Johanna Giuranna ^70^  Paola Giusti-Rodríquez ^35^  Fragiskos Gonidakis ^71^  Scott Gordon ^72^  Philip Gorwood ^73, 74^  Monica Gratacos Mayora ^48, 49, 50^  Jakob Grove ^75, 76, 77, 78^  Sébastien Guillaume ^33^  Yiran Guo ^79^  Hakon Hakonarson ^79, 80^  Katherine A Halmi ^81^  Ken B Hanscombe ^82^  Konstantinos Hatzikotoulas ^19, 83^  Joanna Hauser ^84^  Johannes Hebebrand ^70^  Sietske G Helder ^22, 85^  Anjali K Henders ^86^  Stefan Herms ^29, 30^  Beate Herpertz-Dahlmann ^24^  Wolfgang Herzog ^87^  Anke Hinney ^70^  L. John Horwood ^16^  Christopher Hübel ^15, 22^  Liselotte V Petersen ^76, 114, 115^  Dalila Pinto ^88^  Kirstin L Purves ^22^  Anu Raevuori ^101^  Nicolas Ramoz ^18^  Ted Reichborn-Kjennerud ^112, 146^  Valdo Ricca ^147^  Samuli Ripatti ^148^  Stephan Ripke ^149, 150, 151^  Franziska Ritschel ^17, 152^  Marion Roberts ^22^  Alessandro Rotondo ^153^  Dan Rujescu ^69^  Filip Rybakowski ^154^  Paolo Santonastaso ^155^  André Scherag ^156^  Stephen W Scherer ^157, 158^  Ulrike Schmidt ^22^  Nicholas J Schork ^159^  Alexandra Schosser ^160^  Jochen Seitz ^24^  Lenka Slachtova ^161^  P. Eline Slagboom ^131^  Margarita CT Slof-Op 't Landt ^162, 163^  Agnieszka Slopien ^164^  Nicole Soranzo ^19, 165, 166, 167^  Sandro Sorbi ^139, 168^  Lorraine Southam ^19^  Vidar W Steen ^169, 170^  Michael Strober ^171, 172^ | Laura M Huckins ^88^  James I Hudson ^89^  Hartmut Imgart ^90^  Hidetoshi Inoko ^91^  Vladimir Janout ^92^  Susana Jiménez-Murcia ^57, 58^  Craig Johnson ^93^  Jennifer Jordan ^94, 95^  Antonio Julià ^96^  Gursharan Kalsi ^22^  Deborah Kaminská ^97^  Allan S Kaplan ^98, 99, 100^  Jaakko Kaprio ^101, 102^  Leila Karhunen ^103^  Andreas Karwautz ^104^  Martien JH Kas ^1, 105^  Walter H Kaye ^106^  James L Kennedy ^98, 99, 100^  Martin A Kennedy ^107^  Anna Keski-Rahkonen ^101^  Kirsty Kiezebrink ^108^  Youl-Ri Kim ^109^  Katherine M Kirk ^72^  Lars Klareskog ^110^  Kelly L Klump ^111^  Gun Peggy S Knudsen ^112^  Maria C La Via ^8^  Mikael Landén ^15, 113^  Janne T Larsen ^76, 114, 115^  Stephanie Le Hellard ^116, 117, 118^  Virpi M Leppä ^15^  Robert D Levitan ^99^  Dong Li ^79^  Paul Lichtenstein ^15^  Lisa Lilenfeld ^119^  Bochao Danae Lin ^1^  Jolanta Lissowska ^120^  Astri Lundervold ^121^  Jurjen Luykx ^1^  Pierre J Magistretti ^122,123^  Mario Maj ^124^  Katrin Mannik ^53, 125^  Sara Marsal ^96^  Garret D Stuber ^8, 173^  Patrick F Sullivan ^8, 15, 35^  Beata Świątkowska ^174^  Jin P Szatkiewicz ^35^  Ioanna Tachmazidou ^19^  Elena Tenconi ^40^  Laura M Thornton ^8^  Alfonso Tortorella ^175, 176^  Federica Tozzi ^177^  Janet Treasure ^22^  Artemis Tsitsika ^178^  Marta Tyszkiewicz-Nwafor ^164^  Konstantinos Tziouvas ^179^  Annemarie A van Elburg ^2, 180^  Eric F van Furth ^162, 163^  Tracey D Wade ^181^  Gudrun Wagner ^104^  Esther Walton ^17^  Hunna J Watson ^8, 182, 183^  Thomas Werge ^184^  David C Whiteman ^140^  H.-Erich Wichmann ^185^  Elisabeth Widen ^102^  D. Blake Woodside ^99, 100, 186, 187^  Shuyang Yao ^15^  Zeynep Yilmaz ^8, 35^  Eleftheria Zeggini ^19, 83^  Stephanie Zerwas ^8^  Stephan Zipfel ^188^ |
| --- | --- | --- |

Affiliations

1. Brain Center Rudolf Magnus, Department of Translational Neuroscience, University Medical Center Utrecht, Utrecht, The Netherlands

2. Center for Eating Disorders Rintveld, Altrecht Mental Health Institute, Zeist, The Netherlands

3. Sahlgrenska Academy, University of Gothenburg, Gothenburg, Sweden

4. Institute of Environmental Medicine, Karolinska Institutet, Stockholm, Sweden

5. Department of Behavioral Medicine, National Institute of Mental Health, National Center of Neurology and Psychiatry, Kodaira, Tokyo, Japan

6. NORMENT Centre, Division of Mental Health and Addiction, University of Oslo, Oslo University Hospital, Oslo, Norway

7. Biopsychosocial Corporation, Vienna, Austria

8. Department of Psychiatry, University of North Carolina at Chapel Hill, Chapel Hill, North Carolina, US

9. First Faculty of Medicine, Institute of Hygiene and Epidemiology, Charles University, Prague, Czech Republic

10. BioRealm, LLC, Walnut, California, USA

11. Oregon Research Institute, Eugene, Oregon, USA

12. Department of Psychiatry, Center for Neurobiology and Behavior, University of Pennsylvania Perelman School of Medicine, Philadelphia, Pennsylvania, USA

13. Department of Clinical Neuroscience, Karolinska Institutet, Stockholm, Sweden

14. Center for Psychiatry Research, Stockholm Health Care Services, Stockholm City Council, Stockholm, Sweden

15. Department of Medical Epidemiology and Biostatistics, Karolinska Institutet, Stockholm, Sweden

16. Christchurch Health and Development Study, University of Otago, Christchurch, New Zealand

17. Division of Psychological and Social Medicine and Developmental Neurosciences, Faculty of Medicine, Technische Universität Dresden, Dresden, Germany

18. INSERM U894, Centre of Psychiatry and Neuroscience, Paris, France

19. Wellcome Sanger Institute, Wellcome Genome Campus, Hinxton, Cambridge, UK

20. Department of Medical Biology, School of Medicine, University of Split, Split, Croatia

21. The Center for Eating Disorders at Sheppard Pratt, Baltimore, Maryland, US

22. Institute of Psychiatry, Psychology and Neuroscience, Social, Genetic and Developmental Psychiatry (SGDP) Centre, King’s College London, London, UK

23. National Institute for Health Research Biomedical Research Centre, King’s College London and South London and Maudsley National Health Service Trust, London, UK

24. Department of Child and Adolescent Psychiatry, Psychosomatics and Psychotherapy, RWTH Aachen University, Aachen, Germany

25. Department of Nutrition, University of North Carolina at Chapel Hill, Chapel Hill, North Carolina, US

26. Klinikum Frankfurt/Oder, Frankfurt, Germany

27. Medical University of Vienna, Vienna, Austria

28. Clinical Genetics Unit, Department of Woman and Child Health, University of Padova, Padova, Italy

29. Institute of Medical Genetics and Pathology, University Hospital Basel, Basel, Switzerland

30. Department of Biomedicine, University of Basel, Basel, Switzerland

31. Institute of Neuroscience and Medicine (INM-1), Research Center Juelich, Juelich, Germany

32. Life Sciences Institute and Department of Molecular and Integrative Physiology, University of Michigan, Ann Arbor, Michigan, US

33. Department of Emergency Psychiatry and Post-Acute Care, CHRU Montpellier, University of Montpellier, Montpellier, France

34. Department of Psychiatry, University of Minnesota, Minneapolis, Minnesota, US

35. Department of Genetics, University of North Carolina at Chapel Hill, Chapel Hill, North Carolina, US

36. MRC Integrative Epidemiology Unit, University of Bristol, Bristol, UK

37. School of Social and Community Medicine, University of Bristol, Bristol, UK

38. Department of Psychosomatic Medicine and Psychotherapy, Hannover Medical School, Hannover, Germany

39. Department of Nutrition and Dietetics, Harokopio University, Athens, Greece

40. Department of Neurosciences, University of Padova, Padova, Italy

41. College of Nursing, Seattle University, Seattle, Washington, US

42. Department of Psychology, Virginia Commonwealth University, Richmond, Virginia, US

43. College Behavioral and Emotional Health Institute, Virginia Commonwealth University, Richmond, Virginia, US

44. Department of Human & Molecular Genetics, Virginia Commonwealth University, Richmond, Virginia, US

45. Department of Psychiatry, Athens University Medical School, Athens University, Athens, Greece

46. L'institut du thorax, INSERM, CNRS, UNIV Nantes, Nantes, France

47. Department of Psychiatric Genetics, Poznan University of Medical Sciences, Poznan, Poland

48. Barcelona Institute of Science and Technology, Barcelona, Spain

49. Universitat Pompeu Fabra, Barcelona, Spain

50. Centro de Investigación Biomédica en Red en Epidemiología y Salud Pública (CIBERESP), Barcelona, Spain

51. Department of Psychiatry and Behavioral Sciences, Stanford University, Stanford, California, US

52. Department of Child and Adolescent Psychiatry, Psychosomatics and Psychotherapy, University Hospital of Würzburg, Centre for Mental Health, Würzburg, Germany

53. Estonian Genome Center, University of Tartu, Tartu, Estonia

54. Program in Medical and Population Genetics, Broad Institute of the Massachusetts Institute of Technology and Harvard University, Cambridge, Massachusetts, US

55. Department of Psychology, University of Oslo, Oslo University Hospital, Oslo, Norway

56. Genomics and Disease, Bioinformatics and Genomics Programme, Centre for Genomic Regulation, Barcelona, Spain

57. Department of Psychiatry, University Hospital of Bellvitge –IDIBELL and CIBERobn, Barcelona, Spain

58. Department of Clinical Sciences, School of Medicine, University of Barcelona, Barcelona, Spain

59. Department of Psychiatry and Psychotherapy, Ludwig‐Maximilians‐University (LMU), Munich, Germany

60. Schön Klinik Roseneck affiliated with the Medical Faculty of the University of Munich, Munich,Germany

61. Genomics PLC, Oxford, UK

62. Department of Child and Adolescent Psychiatry, University of Münster, Münster, Germany

63. Department of Cancer, Epidemiology and Genetics, Masaryk Memorial Cancer Institute, Brno, Czech Republic

64. Centre for Human Genetics, University of Marburg, Marburg, Germany

65. Institute of Human Genetics, University of Bonn, School of Medicine & University Hospital Bonn, Bonn, Germany

66. Department of Psychiatry (UPK), University of Basel, Basel, Switzerland

67. Department of Surgery, Faculty of Medicine, University of Toronto, Toronto, Ontario, Canada

68. Division of Nephrology and Dialysis, Institute of Internal Medicine and Medical Specialties, Columbus-Gemelli University Hospital, Rome, Italy

69. Department of Psychiatry, Psychotherapy and Psychosomatics, Martin Luther University of Halle-Wittenberg, Halle (Saale), Germany

70. Department of Child and Adolescent Psychiatry, University Hospital Essen, University of Duisburg-Essen, Essen, Germany

71. 1st Psychiatric Department, National and Kapodistrian University of Athens, Medical School, Eginition Hospital, Athens, Greece

72. QIMR Berghofer Medical Research Institute, Brisbane, Queensland, Australia

73. INSERM U1266, Institute of Psychiatry and Neuroscience of Paris, Paris, France

74. CMME (GHU Paris Psychiatrie et Neurosciences), Paris Descartes University, Paris, France

75. Department of Biomedicine, Aarhus University, Aarhus, Denmark

76. The Lundbeck Foundation Initiative for Integrative Psychiatric Research (iPSYCH), Aarhus, Denmark

77. Centre for Integrative Sequencing, iSEQ, Aarhus University, Aarhus, Denmark

78. Bioinformatics Research Centre, Aarhus University, Aarhus, Denmark

79. Center for Applied Genomics, Children's Hospital of Philadelphia, Philadelphia, Pennsylvania, US

80. Department of Pediatrics, University of Pennsylvania Perelman School of Medicine, Philadelphia, Pennsylvania, US

81. Department of Psychiatry, Weill Cornell Medical College, New York, New York, US

82. Department of Medical and Molecular Genetics, King's College London, Guy’s Hospital, London, UK

83. Institute of Translational Genomics, Helmholtz Zentrum München - German Research Centre for Environmental Health, Neuherberg, Germany

84. Department of Adult Psychiatry, Poznan University of Medical Sciences, Poznan, Poland

85. Zorg op Orde, Delft, The Netherlands

86. Institute for Molecular Bioscience, University of Queensland, Brisbane, Queensland, Australia

87. Department of General Internal Medicine and Psychosomatics, Heidelberg University Hospital, Heidelberg University, Heidelberg, Germany

88. Department of Psychiatry, and Genetics and Genomics Sciences Division of Psychiatric Genomics, Icahn School of Medicine at Mount Sinai, New York, New York, US

89. Biological Psychiatry Laboratory, McLean Hospital/Harvard Medical School, Boston, Massachusetts, US

90. Eating Disorders Unit, Parklandklinik, Bad Wildungen, Germany

91. Department of Molecular Life Science Division of Basic Medical Science and Molecular Medicine, School of Medicine, Tokai University, Isehara, Japan

92. Faculty of Health Sciences, Palacky University, Olomouc, Czech Republic

93. Eating Recovery Center, Denver, Colorado, US

94. Department of Psychological Medicine, University of Otago, Christchurch, New Zealand

95. Canterbury District Health Board, Christchurch, New Zealand

96. Rheumatology Research Group, Vall d’Hebron Research Institute, Barcelona, Spain

97. Department of Psychiatry, First Faculty of Medicine, Charles University, Prague, Czech Republic

98. Centre for Addiction and Mental Health, Toronto, Ontario, Canada

99. Institute of Medical Science, University of Toronto, Toronto, Ontario, Canada

100. Department of Psychiatry, University of Toronto, Toronto, Ontario, Canada

101. Department of Public Health, University of Helsinki, Helsinki, Finland

102. Institute for Molecular Medicine FIMM, HiLIFE, University of Helsinki, Helsinki, Finland

103. Institute of Public Health and Clinical Nutrition, Department of Clinical Nutrition, University of Eastern Finland, Kuopio, Finland

104. Eating Disorders Unit, Department of Child and Adolescent Psychiatry, Medical University of Vienna, Vienna, Austria

105. Groningen Institute for Evolutionary Life Sciences, University of Groningen, Groningen, The Netherlands

106. Department of Psychiatry, University of California San Diego, San Diego, California, US

107. Department of Pathology and Biomedical Science, University of Otago, Christchurch, New Zealand

108. Institute of Applied Health Sciences, School of Medicine, Medical Sciences and Nutrition, University of Aberdeen, Aberdeen, UK

109. Department of Psychiatry, Seoul Paik Hospital, Inje University, Seoul, Korea

110. Rheumatology Unit, Department of Medicine, Center for Molecular Medicine, Karolinska Institutet and Karolinska University Hospital, Stockholm, Sweden

111. Department of Psychology, Michigan State University, East Lansing, Michigan, US

112. Department of Mental Disorders, Norwegian Institute of Public Health, Oslo, Norway

113. Department of Psychiatry and Neurochemistry Institute of Neuroscience and Physiology, The Sahlgrenska Academy at the University of Gothenburg, Gothenburg, Sweden

114. National Centre for Register-Based Research, Aarhus BSS, Aarhus University, Aarhus, Denmark

115. Centre for Integrated Register-based Research (CIRRAU), Aarhus University, Aarhus, Denmark

116. Department of Clinical Science, K.G. Jebsen Centre for Psychosis Research, Norwegian Centre for Mental Disorders Research (NORMENT), University of Bergen, Bergen, Norway

117. Dr. Einar Martens Research Group for Biological Psychiatry, Center for Medical Genetics and Molecular Medicine, Haukeland University Hospital, Bergen, Norway

118. Department of Clinical Medicine, Laboratory Building, Haukeland University Hospital, Bergen, Norway

119. The Chicago School of Professional Psychology, Washington DC, US

120. Department of Cancer Epidemiology and Prevention, M Skłodowska-Curie Cancer Center - Oncology Center, Warsaw, Poland

121. University of Bergen, K. G. Jebsen Center for Neuropsychiatric Disorders, Bergen, Norway

122. BESE Division, King Abdullah University of Science and Technology, Thuwal, Saudi Arabia

123. Department of Psychiatry, University of Lausanne-University Hospital of Lausanne (UNIL-CHUV), Lausanne, Switzerland

124. Department of Psychiatry, University of Campania "Luigi Vanvitelli", Naples, Italy

125. Center for Integrative Genomics, University of Lausanne, Lausanne, Switzerland

126. Department of Paediatric Laboratory Medicine Division of Genome Diagnostics, The Hospital for Sick Children, Toronto, Ontario, Canada

127. Department of Psychiatry, Psychosomatics and Psychotherapy, University of Würzburg, Würzburg, Germany

128. Department of Psychiatry, University College Cork, Cork, Ireland

129. Eist Linn Adolescent Unit, Bessborough, Health Service Executive South, Cork, Ireland

130. Institute of Molecular and Cell Biology, University of Tartu, Tartu, Estonia

131. Molecular Epidemiology Section (Department of Biomedical Datasciences), Leiden University Medical Centre, Leiden, The Netherlands

132. Department of Psychiatry, Faculty of Medicine, University of Geneva, Geneva, Switzerland

133. Division of Child and Adolescent Psychiatry, Geneva University Hospital, Geneva, Switzerland

134. Department of Psychiatry and Behavioral Science, University of North Dakota School of Medicine and Health Sciences, Fargo, North Dakota, US

135. National Center for PTSD, VA Boston Healthcare System, Boston, Massachusetts, US

136. Department of Psychiatry, Boston University School of Medicine, Boston, Massachusetts, US

137. Department of Medicine, Surgery and Dentistry "Scuola Medica Salernitana", University of Salerno, Salerno, Italy

138. Queensland Brain Institute, University of Queensland, Brisbane, Queensland, Australia

139. Department of Neuroscience, Psychology, Drug Research and Child Health (NEUROFARBA), University of Florence, Florence, Italy

140. Population Health Department, QIMR Berghofer Medical Research Institute, Brisbane, Queensland, Australia

141. Center for Neurobehavioral Genetics Semel Institute for Neuroscience and Human Behavior, University of California Los Angeles, Los Angeles, California, USA

142. Department of Psychiatry, Erasmus MC, University Medical Center Rotterdam, Rotterdam, The Netherlands

143. Kartini Clinic, Portland, Oregon, US

144. Center for Human Genome Research, Massachusetts General Hospital, Boston, Massachusetts, USA

145. Biostatistics and Computational Biology Unit, University of Otago, Christchurch, New Zealand

146. Institute of Clinical Medicine, University of Oslo, Oslo, Norway

147. Department of Health Science, University of Florence, Florence, Italy

148. Department of Biometry, University of Helsinki, Helsinki, Finland

149. Analytic and Translational Genetics Unit, Department of Medicine, Massachusetts General Hospital and Harvard Medical School, Boston, Massachusetts, US

150. Stanley Center for Psychiatric Research, Broad Institute of the Massachusetts Institute of Technology and Harvard University, Cambridge, Massachusetts, US

151. Department of Psychiatry and Psychotherapy, Charité - Universitätsmedizin, Berlin, Germany

152. Eating Disorders Research and Treatment Center, Department of Child and Adolescent Psychiatry, Faculty of Medicine, Technische Universität Dresden, Dresden, Germany

153. Department of Psychiatry, Neurobiology, Pharmacology, and Biotechnologies, University of Pisa, Pisa, Italy

154. Department of Psychiatry, Poznan University of Medical Sciences, Poznan, Poland

155. Department of Neurosciences, Padua Neuroscience Center, University of Padova, Padova, Italy

156. Institute of Medical Statistics, Computer and Data Sciences, Jena University Hospital, Jena, Germany

157. Department of Genetics and Genomic Biology, The Hospital for Sick Children, Toronto, Ontario, Canada

158. McLaughlin Centre, University of Toronto, Toronto, Ontario, Canada

159. J. Craig Venter Institute (JCVI), La Jolla, California, US

160. Department of Psychiatry and Psychotherapy, Medical University of Vienna, Vienna, Austria

161. Department of Pediatrics and Center of Applied Genomics, First Faculty of Medicine, Charles University, Prague, Czech Republic

162. Center for Eating Disorders Ursula, Rivierduinen, Leiden, The Netherlands

163. Department of Psychiatry, Leiden University Medical Centre, Leiden, The Netherlands

164. Department of Child and Adolescent Psychiatry, Poznan University of Medical Sciences, Poznan, Poland

165. Donor Health and Genomics, National Institute for Health Research Blood and Transplant Unit, Cambridge, UK

166. Division of Cardiovascular Medicine, British Heart Foundation Centre of Excellence, Cambridge, UK

167. Department of Haematology, University of Cambridge, Cambridge, UK

168. IRCCS Fondazione Don Carlo Gnocchi, Florence, Italy

169. Center for Medical Genetics and Molecular Medicine, Haukeland University Hospital, Bergen, Norway

170. Department of Clinical Science, University of Bergen, Bergen, Norway

171. Department of Psychiatry and Biobehavioral Science, Semel Institute for Neuroscience and Human Behavior, University of California Los Angeles, Los Angeles, California, US

172. David Geffen School of Medicine, University of California Los Angeles, Los Angeles, California, US

173. Department of Cell Biology and Physiology, University of North Carolina at Chapel Hill, Chapel Hill, North Carolina, US

174. Department of Environmental Epidemiology, Nofer Institute of Occupational Medicine, Lodz, Poland

175. Department of Psychiatry, University of Naples SUN, Naples, Italy

176. Department of Psychiatry, University of Perugia, Perugia, Italy

177. Brain Sciences Department, Stremble Ventures, Limassol, Cyprus

178. Adolescent Health Unit, Second Department of Pediatrics, "P. & A. Kyriakou" Children's Hospital, University of Athens, Athens, Greece

179. Pediatric Intensive Care Unit, "P. & A. Kyriakou" Children's Hospital, University of Athens, Athens, Greece

180. Faculty of Social and Behavioral Sciences, Utrecht University, Utrecht, The Netherlands

181. School of Psychology, Flinders University, Adelaide, South Australia, Australia

182. School of Psychology, Curtin University, Perth, Western Australia, Australia

183. School of Paediatrics and Child Health, University of Western Australia, Perth, Western Australia, Australia

184. Department of Clinical Medicine, University of Copenhagen, Copenhagen, Denmark

185. Helmholtz Centre Munich - German Research Center for Environmental Health, Munich, Germany

186. Centre for Mental Health, University Health Network, Toronto, Ontario, Canada

187. Program for Eating Disorders, University Health Network, Toronto, Ontario, Canada

188. Department of Internal Medicine VI, Psychosomatic Medicine and Psychotherapy, University Medical Hospital Tuebingen, Tuebingen, Germany

**References**

1.	Mullins, N. *et al.* GWAS of Suicide Attempt in Psychiatric Disorders and Association With Major Depression Polygenic Risk Scores. *Am. J. Psychiatry* **176**, 651–660 (2019).

2.	Wray, N. R. *et al.* Genome-wide association analyses identify 44 risk variants and refine the genetic architecture of major depression. *Nat. Genet.* **50**, 668–681 (2018).

3.	Stahl, E. A. *et al.* Genome-wide association study identifies 30 loci associated with bipolar disorder. *Nat. Genet.* **51**, 793–803 (2019).

4.	Schizophrenia Working Group of the Psychiatric Genomics Consortium. Biological insights from 108 schizophrenia-associated genetic loci. *Nature* **511**, 421–427 (2014).

5.	Watson, H. J. *et al.* Genome-wide association study identifies eight risk loci and implicates metabo-psychiatric origins for anorexia nervosa. *Nat. Genet.* **51**, 1207–1214 (2019).

6.	Duncan, L. *et al.* Significant Locus and Metabolic Genetic Correlations Revealed in Genome-Wide Association Study of Anorexia Nervosa. *Am. J. Psychiatry* **174**, 850–858 (2017).

7.	CONVERGE Consortium,. Sparse whole-genome sequencing identifies two loci for major depressive disorder. *Nature* **523**, 588–591 (2015).

8.	Ursano, R. J. *et al.* The Army study to assess risk and resilience in servicemembers (Army STARRS). *Psychiatry* **77**, 107–119 (2014).

9.	Posner, K. *et al.* The Columbia-Suicide Severity Rating Scale: initial validity and internal consistency findings from three multisite studies with adolescents and adults. *Am. J. Psychiatry* **168**, 1266–1277 (2011).

10.	Witt, S. H. *et al.* Genome-wide association study of borderline personality disorder reveals genetic overlap with bipolar disorder, major depression and schizophrenia. *Transl. Psychiatry* **7**, e1155 (2017).

11.	Gillespie, C. F. *et al.* Trauma exposure and stress-related disorders in inner city primary care patients. *Gen. Hosp. Psychiatry* **31**, 505–514 (2009).

12.	Sudlow, C. *et al.* UK biobank: an open access resource for identifying the causes of a wide range of complex diseases of middle and old age. *PLoS Med.* **12**, e1001779 (2015).

13.	Smith, D. J. *et al.* Prevalence and characteristics of probable major depression and bipolar disorder within UK biobank: cross-sectional study of 172,751 participants. *PLoS One* **8**, e75362 (2013).

14.	Wu, P.-J. *et al.* The profile and familiality of personality traits in mood disorder families. *J. Affect. Disord.* **138**, 367–374 (2012).

15.	Tsai, H.-C. *et al.* Empirically derived subgroups of bipolar I patients with different comorbidity patterns of anxiety and substance use disorders in Han Chinese population. *Journal of Affective Disorders* vol. 136 81–89 (2012).

16.	Rees, E. *et al.* Analysis of exome sequence in 604 trios for recessive genotypes in schizophrenia. *Transl. Psychiatry* **5**, e607 (2015).

17.	Hwu, H.-G. *et al.* Taiwan schizophrenia linkage study: The field study. *American Journal of Medical Genetics Part B: Neuropsychiatric Genetics* vol. 134B 30–36 (2005).

18.	Pedersen, C. B. *et al.* The iPSYCH2012 case–cohort sample: new directions for unravelling genetic and environmental architectures of severe mental disorders. *Molecular Psychiatry* vol. 23 6–14 (2018).

19.	Li, Q. *et al.* Genome-wide association study of paliperidone efficacy. *Pharmacogenet. Genomics* **27**, 7–18 (2017).

20.	Sleiman, P. *et al.* GWAS meta analysis identifies TSNARE1 as a novel Schizophrenia / Bipolar susceptibility locus. *Scientific Reports* vol. 3 (2013).

21.	Bigdeli, T. B. *et al.* Genetic effects influencing risk for major depressive disorder in China and Europe. *Transl. Psychiatry* **7**, e1074 (2017).

22.	Ben-Efraim, Y. J., Wasserman, D., Wasserman, J. & Sokolowski, M. Family-based study of AVPR1B association and interaction with stressful life events on depression and anxiety in suicide attempts. *Neuropsychopharmacology* **38**, 1504–1511 (2013).

23.	Sokolowski, M., Wasserman, J. & Wasserman, D. Polygenic associations of neurodevelopmental genes in suicide attempt. *Mol. Psychiatry* **21**, 1381–1390 (2016).

24.	Ben-Efraim, Y. J., Wasserman, D., Wasserman, J. & Sokolowski, M. Family-based study of HTR2A in suicide attempts: observed gene, gene × environment and parent-of-origin associations. *Mol. Psychiatry* **18**, 758–766 (2013).

25.	Sokolowski, M., Ben-Efraim, Y. J., Wasserman, J. & Wasserman, D. Glutamatergic GRIN2B and polyaminergic ODC1 genes in suicide attempts: associations and gene-environment interactions with childhood/adolescent physical assault. *Mol. Psychiatry* **18**, 985–992 (2013).

26.	Beck, A. T., Beck, R. & Kovacs, M. Classification of suicidal behaviors: I. Quantifying intent and medical lethality. *Am. J. Psychiatry* **132**, 285–287 (1975).

27.	Beck, A. T., Weissman, A., Lester, D. & Trexler, L. Classification of suicidal behaviors. II. Dimensions of suicidal intent. *Arch. Gen. Psychiatry* **33**, 835–837 (1976).

28.	Byrne, E. M. *et al.* Cohort profile: the Australian genetics of depression study. *BMJ Open* **10**, e032580 (2020).

29.	van Spijker, B. A. J. *et al.* The suicidal ideation attributes scale (SIDAS): Community-based validation study of a new scale for the measurement of suicidal ideation. *Suicide Life Threat. Behav.* **44**, 408–419 (2014).

30.	Olsen, C. M. *et al.* Cohort profile: The QSkin Sun and Health Study. *International Journal of Epidemiology* vol. 41 929–929i (2012).

31.	Gelernter, J. *et al.* Genome-wide association study of alcohol dependence:significant findings in African- and European-Americans including novel risk loci. *Mol. Psychiatry* **19**, 41–49 (2013).

32.	Levey, D. F. *et al.* Genetic associations with suicide attempt severity and genetic overlap with major depression. *Transl. Psychiatry* **9**, 1–12 (2019).

33.	Diagnostic reliability of the Semi-structured Assessment for Drug Dependence and Alcoholism (SSADDA). *Drug Alcohol Depend.* **80**, 303–312 (2005).

34.	Galfalvy, H. *et al.* A genome-wide association study of suicidal behavior. *American Journal of Medical Genetics Part B: Neuropsychiatric Genetics* vol. 168 557–563 (2015).

35.	Otsuka, I. *et al.* Genome-wide association studies identify polygenic effects for completed suicide in the Japanese population. *Neuropsychopharmacology* **44**, 2119–2124 (2019).

36.	Calkins, M. E. *et al.* The Consortium on the Genetics of Endophenotypes in Schizophrenia: Model Recruitment, Assessment, and Endophenotyping Methods for a Multisite Collaboration. *Schizophrenia Bulletin* vol. 33 33–48 (2006).

37.	Van Vliet, I. M. & De Beurs, E. The MINI-International Neuropsychiatric Interview. A brief structured diagnostic psychiatric interview for DSM-IV en ICD-10 psychiatric disorders. *Tijdschr. Psychiatr.* **49**, 393–397 (2007).

38.	Hemmings, S. M. J. *et al.* BDNF Val66Met modifies the risk of childhood trauma on obsessive-compulsive disorder. *J. Psychiatr. Res.* **47**, 1857–1863 (2013).

39.	Syal, S. *et al.* Grey matter abnormalities in social anxiety disorder: a pilot study. *Metab. Brain Dis.* **27**, 299–309 (2012).

40.	von Rhein, D. *et al.* The NeuroIMAGE study: a prospective phenotypic, cognitive, genetic and MRI study in children with attention-deficit/hyperactivity disorder. Design and descriptives. *Eur. Child Adolesc. Psychiatry* **24**, 265–281 (2015).

41.	Neale, B. M. *et al.* Genome-wide association scan of attention deficit hyperactivity disorder. *Am. J. Med. Genet. B Neuropsychiatr. Genet.* **147B**, 1337–1344 (2008).

42.	Polderman, T. J. C. *et al.* Attentional switching forms a genetic link between attention problems and autistic traits in adults. *Psychol. Med.* **43**, 1985–1996 (2013).

43.	[Weiss, R. H. Grundintelligenztest skala 2—revision CFT 20-R [culture fair intelligence test scale 2—revision]. *Hogrefe, Göttingen* (2006).](http://paperpile.com/b/AtB1ay/4p3g3)

44.	Pato, M. T. *et al.* The genomic psychiatry cohort: partners in discovery. *Am. J. Med. Genet. B Neuropsychiatr. Genet.* **162B**, 306–312 (2013).

45.	Wilcox, H. C. *et al.* Traumatic Stress Interacts With Bipolar Disorder Genetic Risk to Increase Risk for Suicide Attempts. *J. Am. Acad. Child Adolesc. Psychiatry* **56**, 1073–1080 (2017).

46.	Nurnberger, J. I., Jr *et al.* A high-risk study of bipolar disorder. Childhood clinical phenotypes as precursors of major mood disorders. *Arch. Gen. Psychiatry* **68**, 1012–1020 (2011).

47.	Cervilla, J. A. *et al.* Protocol and methodology of Study epidemiological mental health in Andalusia: PISMA-ep. *Rev. Psiquiatr. Salud Ment.* **9**, 185–194 (2016).

48.	Liberzon, I. *et al.* Interaction of the ADRB2 gene polymorphism with childhood trauma in predicting adult symptoms of posttraumatic stress disorder. *JAMA Psychiatry* **71**, 1174–1182 (2014).

49.	Breslau, N. *et al.* Trauma and posttraumatic stress disorder in the community: the 1996 Detroit Area Survey of Trauma. *Arch. Gen. Psychiatry* **55**, 626–632 (1998).

50.	Prescott, M. R. *et al.* Validation of lay-administered mental health assessments in a large Army National Guard cohort. *International Journal of Methods in Psychiatric Research* vol. 23 109–119 (2014).

51.	Perich, T. *et al.* What clinical features precede the onset of bipolar disorder? *J. Psychiatr. Res.* **62**, 71–77 (2015).

52.	Watkeys, O. J. *et al.* Derivation of poly-methylomic profile scores for schizophrenia. *Prog. Neuropsychopharmacol. Biol. Psychiatry* **101**, 109925 (2020).

53.	Weickert, T. W. *et al.* Adjunctive raloxifene treatment improves attention and memory in men and women with schizophrenia. *Mol. Psychiatry* **20**, 685–694 (2015).

54.	Morgan, C. A., 3rd *et al.* Symptoms of dissociation in humans experiencing acute, uncontrollable stress: a prospective investigation. *Am. J. Psychiatry* **158**, 1239–1247 (2001).

55.	Schnurr, P., Vielhauer, M. & Weathers, F. Brief Trauma Interview. *Unpublished Interview* (1995).

56.	Breslau, N., Peterson, E. L., Kessler, R. C. & Schultz, L. R. Short Screening Scale for DSM-IV Posttraumatic Stress Disorder. *American Journal of Psychiatry* vol. 156 908–911 (1999).

57.	Koenen, K. C. *et al.* Protocol for investigating genetic determinants of posttraumatic stress disorder in women from the Nurses’ Health Study II. *BMC Psychiatry* **9**, 29 (2009).

58.	Weathers FW, F. J. Psychometric review of PTSD Checklist (PCL-C, PCL-S, PCL-M, PCL-PR). in *Measurement of Stress, Trauma, and Adaptation* (ed. Hudnall Stamm, B.) (Sidran Press, 1996).

59.	American Psychiatric Association. *Diagnostic and Statistical Manual of Mental Disorders, 4th Edition, (DSM-IV)*. (American Psychiatric Association, 1994).

60.	Jacquet, H. *et al.* Hyperprolinemia is a risk factor for schizoaffective disorder. *Mol. Psychiatry* **10**, 479–485 (2005).

61.	Castro, V. M. *et al.* Validation of electronic health record phenotyping of bipolar disorder cases and controls. *Am. J. Psychiatry* **172**, 363–372 (2015).

62.	Rovira, P. *et al.* Shared genetic background between children and adults with attention deficit/hyperactivity disorder. *Neuropsychopharmacology* (2020) doi:[10.1038/s41386-020-0664-5](http://dx.doi.org/10.1038/s41386-020-0664-5).

63.	Hedström, A. K., Hillert, J., Olsson, T. & Alfredsson, L. Alcohol as a modifiable lifestyle factor affecting multiple sclerosis risk. *JAMA Neurol.* **71**, 300–305 (2014).

64.	Smith, L. *et al.* Establishing the UK DNA Bank for motor neuron disease (MND). *BMC Genet.* **16**, 84 (2015).

65.	Gatt, J. M. *et al.* The TWIN-E project in emotional wellbeing: study protocol and preliminary heritability results across four MRI and DTI measures. *Twin Res. Hum. Genet.* **15**, 419–441 (2012).

66.	Jamshidi, J. *et al.* Diverse phenotypic measurements of wellbeing: Heritability, temporal stability and the variance explained by polygenic scores. *Genes Brain Behav.* e12694 (2020).

67.	Swanson, J. M. et al. Overidentification of extreme behavior in the evaluation and diagnosis of ADHD/HKD. <http://www.adhd.net/SWAN_Paper.pdf> (2001).

68.	Waldman, I. D. *et al.* Association and linkage of the dopamine transporter gene and attention-deficit hyperactivity disorder in children: heterogeneity owing to diagnostic subtype and severity. *Am. J. Hum. Genet.* **63**, 1767–1776 (1998).

69.	Lam, M. *et al.* RICOPILI: Rapid Imputation for COnsortias PIpeLIne. *Bioinformatics* (2019) doi:[10.1093/bioinformatics/btz633](http://dx.doi.org/10.1093/bioinformatics/btz633).

70.	Howie, B., Marchini, J. & Stephens, M. Genotype Imputation with Thousands of Genomes. *G3: Genes|Genomes|Genetics* vol. 1 457–470 (2011).

71.	Delaneau, O., Marchini, J. & Zagury, J.-F. A linear complexity phasing method for thousands of genomes. *Nat. Methods* **9**, 179–181 (2011).

72.	1000 Genomes Project Consortium *et al.* A map of human genome variation from population-scale sequencing. *Nature* **467**, 1061–1073 (2010).

73.	Chang, C. C. *et al.* Second-generation PLINK: rising to the challenge of larger and richer datasets. *Gigascience* **4**, 7 (2015).

74.	Price, A. L. *et al.* Principal components analysis corrects for stratification in genome-wide association studies. *Nat. Genet.* **38**, 904–909 (2006).

75.	Willer, C. J., Li, Y. & Abecasis, G. R. METAL: fast and efficient meta-analysis of genomewide association scans. *Bioinformatics* **26**, 2190–2191 (2010).

76.	Lunter, G. & Goodson, M. Stampy: a statistical algorithm for sensitive and fast mapping of Illumina sequence reads. *Genome Res.* **21**, 936–939 (2011).

77.	The 1000 Genomes Project Consortium, *et al.* An integrated map of genetic variation from 1,092 human genomes. *Nature* **491**, 56–65 (2012).

78.	Browning, S. R. & Browning, B. L. Rapid and accurate haplotype phasing and missing-data inference for whole-genome association studies by use of localized haplotype clustering. *Am. J. Hum. Genet.* **81**, 1084–1097 (2007).

79.	Das, S. *et al.* Next-generation genotype imputation service and methods. *Nat. Genet.* **48**, 1284–1287 (2016).

80.	Loh, P.-R., Palamara, P. F. & Price, A. L. Fast and accurate long-range phasing in a UK Biobank cohort. *Nat. Genet.* **48**, 811–816 (2016).

81.	Nievergelt, C. M. *et al.* International meta-analysis of PTSD genome-wide association studies identifies sex- and ancestry-specific genetic risk loci. *Nat. Commun.* **10**, 4558 (2019).

82.	Bycroft, C. *et al.* The UK Biobank resource with deep phenotyping and genomic data. *Nature* **562**, 203–209 (2018).

83.	Abraham, G., Qiu, Y. & Inouye, M. FlashPCA2: principal component analysis of Biobank-scale genotype datasets. *Bioinformatics* **33**, 2776–2778 (2017).

84.	Ruderfer, D. M. *et al.* Significant shared heritability underlies suicide attempt and clinically predicted probability of attempting suicide. *Mol. Psychiatry* **25**, 2422–2430 (2020).

85.	Woolston, A. L. *et al.* Genetic loci associated with an earlier age at onset in multiplex schizophrenia. *Sci. Rep.* **7**, 6486 (2017).

86.	Børglum, A. D. *et al.* Genome-wide study of association and interaction with maternal cytomegalovirus infection suggests new schizophrenia loci. *Mol. Psychiatry* **19**, 325–333 (2014).

87.	Hollegaard, M. V. *et al.* Robustness of genome-wide scanning using archived dried blood spot samples as a DNA source. *BMC Genet.* **12**, 58 (2011).

88.	Browning, B. L. & Browning, S. R. A unified approach to genotype imputation and haplotype-phase inference for large data sets of trios and unrelated individuals. *Am. J. Hum. Genet.* **84**, 210–223 (2009).

89.	Shabalin, A. A. *et al.* RaMWAS: fast methylome-wide association study pipeline for enrichment platforms. *Bioinformatics* **34**, 2283–2285 (2018).

90.	Loh, P.-R. *et al.* Reference-based phasing using the Haplotype Reference Consortium panel. *Nat. Genet.* **48**, 1443–1448 (2016).
